# Global genomic surveillance of monkeypox virus

**DOI:** 10.1101/2024.08.15.24312031

**Authors:** James R. Otieno, Christopher Ruis, Bernard A. Onoja, Krutika Kuppalli, Ana Hoxha, Andreas Nitsche, Annika Brinkmann, Janine Michel, Placide Mbala-Kisengeni, Daniel Mukadi-Bamuleka, Muntasir Mohammed Osman, Hanadi Elawad Hussein, Muhammad Ali Raja, Richard Fotsing, Belinda L. Herring, Mory Keita, Jairo Mendez Rico, Lionel Gresh, Amal Barakat, Victoria Katawera, Karen Nahapetyan, Dhamari Naidoo, R. Andres Floto, Jane Cunningham, Maria D. Van Kerkhove, Rosamund Lewis, Lorenzo Subissi

**Affiliations:** World Health Organization, Geneva, Switzerland; Victor Phillip Dahdaleh Heart & Lung Research Institute, University of Cambridge, Cambridge, UK; Cambridge Centre for AI in Medicine, University of Cambridge, Cambridge, UK; Department of Veterinary Medicine, University of Cambridge, Cambridge, UK; Robert Koch Institute, Berlin, Germany; National Institute for Biomedical Research, Kinshasa, Democratic Republic of the Congo; University of Kinshasa, Kinshasa, Democratic Republic of the Congo; National Institute for Biomedical Research, Goma, Democratic Republic of the Congo; Health Emergencies and Epidemics Control, Federal Ministry of Health, Khartoum, Sudan; World Health Organization Country Office, Khartoum, Sudan; World Health Organization Country Office, Kinshasa, Democratic Republic of the Congo; World Health Organization Regional Office for Africa, Brazzaville, Republic of the Congo; World Health Organization Regional Office for the Americas, Washington, United States of America; World Health Organization Regional Office for the Eastern Mediterranean, Cairo, Egypt; World Health Organization Regional Office for the Western Pacific, Manila, the Philippines; World Health Organization Regional Office for Europe, Copenhagen, Denmark; World Health Organization Regional Office for South East Asia, New Delhi, India; Cambridge Centre for Lung Infection, Papworth Hospital, Cambridge, UK

## Abstract

Monkeypox virus (MPXV) is endemic in Western and Central Africa and, in May 2022, a clade IIb lineage (B.1) caused a global outbreak outside Africa, resulting in its detection in 117 countries/territories. To understand the global phylogenetics of MPXV, we carried out the first analysis of all available MPXV sequences, including 10,670 sequences from 65 countries collected between 1958 and 2024. Our analysis reveals high mobility of clade I viruses within Central Africa, sustained human-to-human transmission of clade IIb lineage A viruses within the Eastern Mediterranean region, and distinct mutational signatures that can distinguish sustained human-to-human from animal-to-animal transmission. Moreover, distinct clade I sequences from Sudan suggest local MPXV circulation in areas of Eastern Africa over the past four decades. Our study underscores the importance of genomic surveillance in tracking spatiotemporal dynamics of MXPV clades and the need to strengthen such surveillance, including in some parts of Eastern Africa.

## Introduction

Mpox, formerly known as monkeypox, is a disease that is caused by the *monkeypox virus* (MPXV). MPXV is a member of the *orthopoxvirus* genus, which also includes the *variola virus*, the causative agent of smallpox.^1^ In humans, mpox can be associated with a range of clinical symptoms, but classically presents with a short febrile prodromal phase, which lasts 1-5 days, followed by the appearance of skin and/or mucosal rash, which might include single or multiple lesions.^2–4^ The incubation period of mpox has historically ranged from 5 to 21 days.^5^ MPXV is divided genetically into two clades — clade I (formerly known as Congo Basin clade) and clade II (formerly known as West African clade), which is further classified into subclades IIa and IIb.^6^ Clade I and subclade IIa circulate endemically within as yet unknown animal reservoirs, potentially including rodents and non-human primates, and human cases are mostly the result of spill-over from these reservoirs.^1,7,8^ Historical surveillance has not been sufficient to identify the frequency of spillover. In 2022, mpox epidemiology shifted with emergence of a new human-adapted lineage – clade IIb - that spread worldwide, and based on the vast number of sequences from this outbreak, it was inferred that clade IIb has circulated continually within humans since at least 2016.^9^ Mpox human-to-human transmission primarily occurs through direct contact with infected lesions or bodily fluids, which includes sexual contact, but transmission can also occur through contact with fomites.^10^

In May 2022, a novel lineage of clade IIb termed B.1 emerged and spread globally, establishing efficient local transmission within many countries with no previous history of mpox transmission. As of 30 April 2024, the multi-country outbreak has been associated with 97,208 cases and 186 fatalities from 117 countries/areas/territories, representing a case fatality ratio (CFR) of 0.19%.^11^ The outbreak is primarily driven by sexual transmission among males who self-identify as men who have sex with men, with 7% of cases requiring hospitalization. Other groups at higher risk of hospitalization include female cases, those younger than 5 years of age or greater than 65 years of age, and the immunosuppressed (either due to being HIV positive or from other immunocompromising conditions).^12^ In response to the global outbreak, a number of countries have started to establish mpox surveillance programs.

As well as the global outbreak, detection of mpox is increasing within endemic regions. For example, in 2023, a total of 14 626 mpox suspected cases and suspected 654 deaths (CFR 4.5%) were reported in the DRC, representing the highest figures in the recorded in the country and the highest among countries in the WHO African Region.^13^ In 2024, a total of 7 851 mpox suspected cases were reported as of 26 May, including 384 suspected deaths (CFR 4.9%).^13^ This is substantially higher than previous years. This recent increase, along with the newly documented sexual transmission recorded in March and then July-September 2023 in Kwango and South Kivu provinces, respectively, confirm the growing importance of human-to-human transmission, including through sexual contact, in the DRC.^14–16^ Furthermore, this highlights the importance of MPXV as an emerging human pathogen.

The global mpox surveillance that was quickly established in 2022 provided a platform for the generation of genome sequencing data. This data has been useful to characterize MPXV evolution, understand the origins of emerging lineages and monitor local and global spread. Previous studies have shown that clade IIb exhibits a higher substitution rate than other *orthopoxvirus* variants. ^9,17–19^ This appears to be due to elevated TC>TT mutations (which represents C mutating to T with an upstream T nucleotide and also includes the reverse GA>AA mutations) driven by human APOBEC-3 enzymes causing cytosine deamination in the viral genome.^9,17–19^ This mutational signature has enabled inference that clade IIb is a human-adapted lineage.

Global genomic surveillance can also enable monitoring of the integrity and stability of the MPXV genomic termini, which can rearrange driving gene duplication or gene loss,^20^ which are also drivers of poxvirus evolution and adaption to the host, and are therefore important to monitor.^21,22^

Mpox surveillance programs and their associated genomic strategies globally remain critical to understand the disease and characterize the virus evolutionary trajectory and genetic diversity, as well as potential insights into the associated phenotype. Ultimately, this data supports the deployment of suitable countermeasures (diagnostics, therapeutics, and vaccines), leveraging efforts garnered from smallpox interventions prior to eradication and preparedness in the years since, as well as advance research and development for countermeasures. Here, we report a global analysis of the publicly available MPXV genomic sequence data, which provide key insights into the spatiotemporal spread, host species range and evolution of this ongoing threat.

## Results

We collected all available MXPV sequences and filtered these to retain 10 546 high quality sequences from 64 countries (see **Methods**). Of those, 6 585 sequences were extracted from GenBank (62%) and 3 914 sequences from GISAID (37%). Due to their distinct epidemiology and sequence diversity, we divided clade IIb into the A sublineages and the B.1 lineage for the analyses below, and refer to these groupings as clade IIb A and lineage B.1, respectively. As expected, the majority (97.7%) of the available MPXV sequences cluster within lineage B.1 (**Figure S1a**), representing intensive genomic sequencing efforts during the global outbreak. Clades I, IIa and IIb A have been sequenced far less often (**Figure S1a**). Correspondingly, the majority (98.6%) of MPXV sequences were collected from 2022-2024, with limited historical surveillance resulting in many years between 1958 and 2015 with no sequences in global databases. (**Figure S1b**). Nonetheless, we observe differences in the temporal, spatial and host species distributions of the major MPXV clades and discuss these below.

### MPXV clades exhibit distinct temporal sampling distributions

Subclade IIa and clade I were the first to be detected, in 1958 and 1970, respectively (**Figures 1**, **S1c**).^23,24^ Sporadic detection of both of these clades has continued through to recent years (**Figures 1**, **S1c**), showing continued circulation within the animal reservoir. Clade I continued to be detected in DRC and Sudan during the lineage B.1 outbreak between 2022 and 2024 (**Figure 1**), with DRC cases in 2024 being associated with a novel divergent lineage showing signatures of human-to-human transmission within South Kivu.^15,25^ Clade IIa has not been observed since 2018 (**Figure 1**). Except for a sample from 1971, all clade IIb genomes were sampled from 2017-2023. The 1971 sample likely reflects the fact that also clade IIb originated in an animal reservoir. Clade IIb A was first detected in Nigeria in 2017 and has continued circulating through human-to-human transmission to at least 2023,^8^ while the descendent lineage B.1 was first detected in 2022 (**Figures 1**, **S1c**).

**Figure 1.**
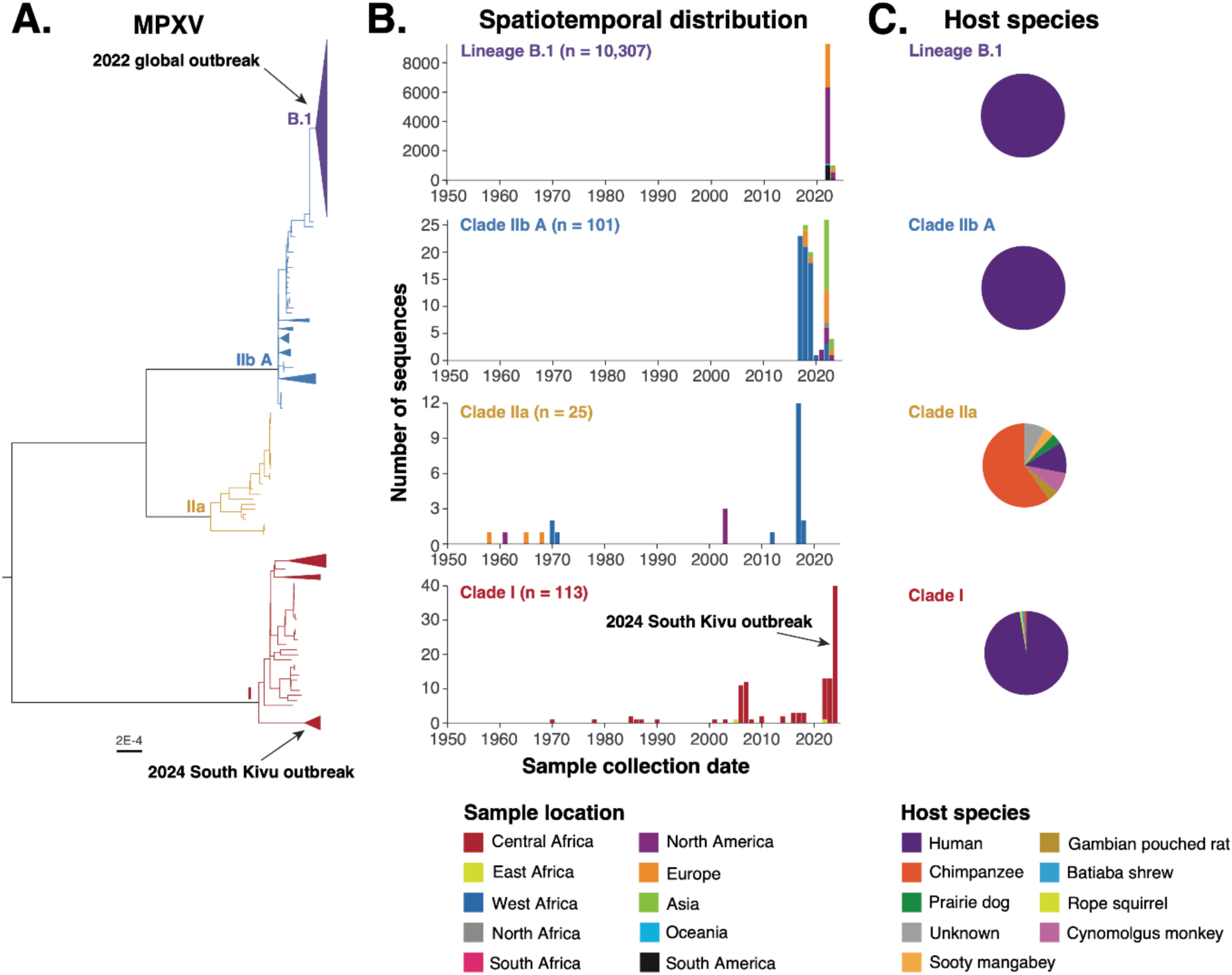
Spatiotemporal and host species distributions of MPXV sequences. (**A**) Maximum likelihood phylogenetic tree highlighting the major clades of MPXV. Branches are coloured by clade. Lineage B.1 clusters within clade IIb and caused the 2022 global MPXV outbreak; this lineage is therefore separated from the remainder of clade IIb. Scale bar shows the expected number of nucleotide substitutions per site. A subset of clades are collapsed for clarity. (**B**) The temporal and regional distribution of MPXV sequences is shown for each clade. The n numbers show the total number of sequences from the clade. (**C**) Distributions of the number of sequences from each host species.

### MPXV sampling shows distinct host species patterns

Although it likely originated from an animal reservoir, clade IIb circulates via human-to-human transmission^26^ and correspondingly both clade IIb A and B.1 lineages have, to date, been sampled exclusively in humans (**Figure 1**). While clades I and IIa both circulate within poorly understood animal reservoirs, the hosts from which they have been sampled are markedly different. The majority of clade I sequences (96%) have been sampled from humans, with single samples from an outbreak in captive chimpanzees,^27^ and from wild shrew (*Crocidura littoralis*) and rope squirrel (*Funisciurus Anerythrus*).^28^ Conversely, only 12% (3/25) of clade IIa samples with recorded host species were collected in humans (**Figure 1**), two in 1970 and one in 2003. Clade IIa has been isolated most often in chimpanzees (60%, **Figure 1**), although these samples are mostly from a single study within Taï National Park in Cote d’Ivoire,^7^ and chimpanzees are likely a spillover host rather than a reservoir host. Clade IIa has additionally been isolated from a wild sooty mangabey, imported cynomolgus monkeys in USA and Denmark, and a prairie dog during the 2003 USA outbreak (**Figure 1**).^29^

### Geographical distribution of MPXV clades

Clade I has mostly been isolated from the Congo Basin area, where it has been sampled in the DRC, the Republic of the Congo, Central African Republic, Cameroon and Gabon (**Figures 2**, **S2**). Sampling locations are broadly spread around these countries. Sequences from individual countries and provinces often do not cluster within the phylogenetic tree (**Figure 2**), demonstrating multiple introductions of clade I into local geographical regions.

**Figure 2.**
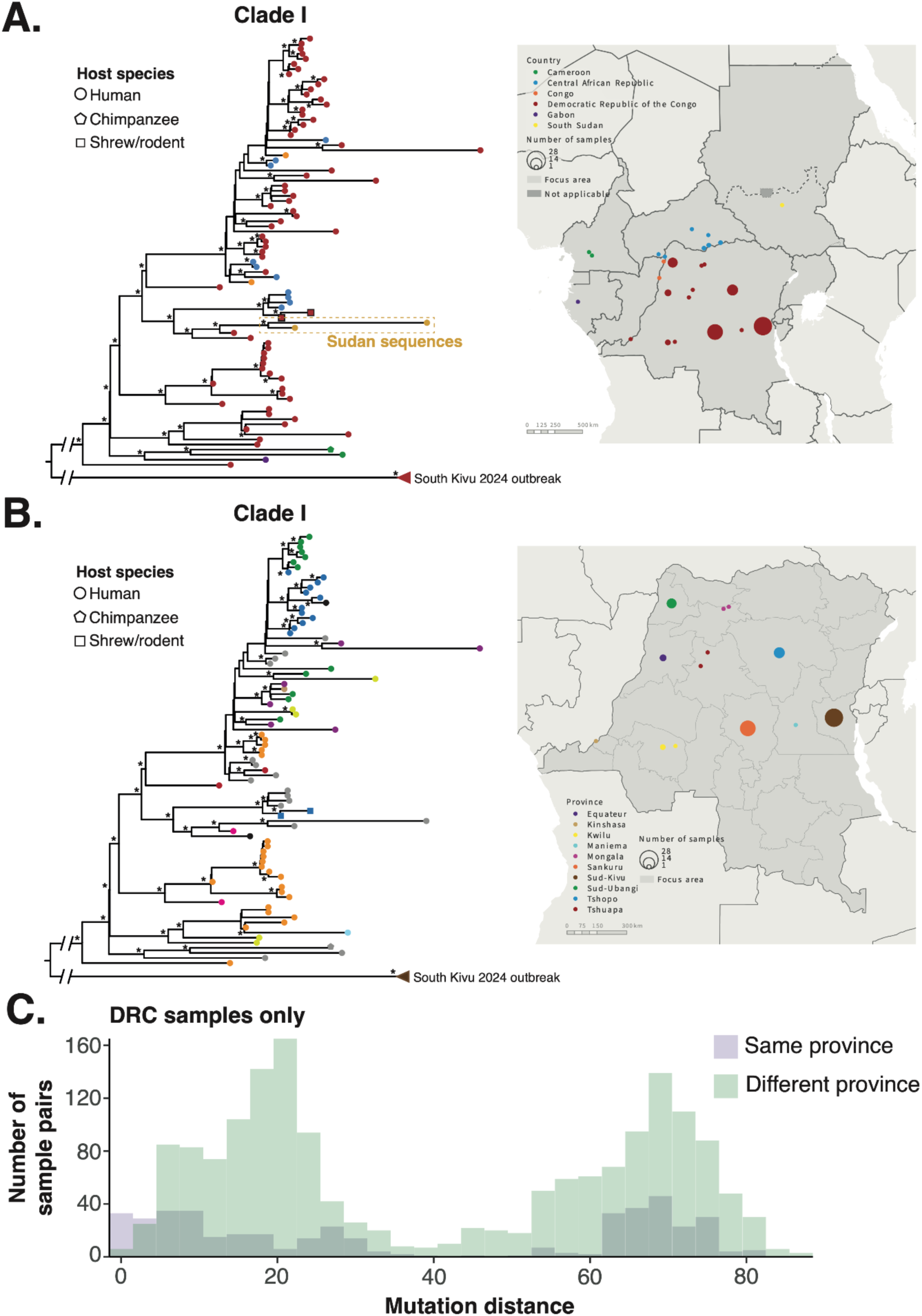
Regular international and inter-province transmission of clade I. (**A, B**) Maximum likelihood phylogenetic tree of 113 high-quality clade I sequences. (**A**) Tips are coloured by country to match the map and shapes show the host species from which the sequence was isolated. The Sudan sequences cluster is highlighted. The 2024 South Kivu outbreak clade has been collapsed for clarity. Asterisks show phylogenetic nodes with bootstrap support of 70 or higher. The map shows sampling locations with points proportional to the number of sequences from the location. (**B**) Tips are coloured by province within DRC to match the map. Tips collected outside DRC are coloured grey and tips sampled within DRC but without a recorded province are coloured black. (**C**) Mutation distance between all pairs of clade I samples stratified by whether the pairs are from the same *(purple*) or different (*green*) provinces.

Having first confirmed the presence of a temporal signal (see **Methods**), we sought to identify the timescale of these virus movements by reconstructing a temporal phylogenetic tree (**Figure 3**). We found that the most recent common ancestor of clade I (excluding the 2024 South Kivu outbreak which has a different substitution rate, see **Methods**)^9^ occurred in approximately 1917 (95% highest probability density (HPD) 1880-1949). We observe frequent international and inter-province transmission over the past several decades (**Figures 2**, **3**). For example, a clade sampled in Sud-Ubangi, Equateur and Kinshasa in 2023-2024 coalesces to a common ancestor in 2007 (95% HPD 2003-2011), supporting recent virus movement within the animal reservoir. Furthermore, we observe highly similar distributions of genetic relatedness between clade I samples from the same and different provinces within DRC (**Figure 2C**), further highlighting the regular movement of viruses between geographical locations.

**Figure 3.**
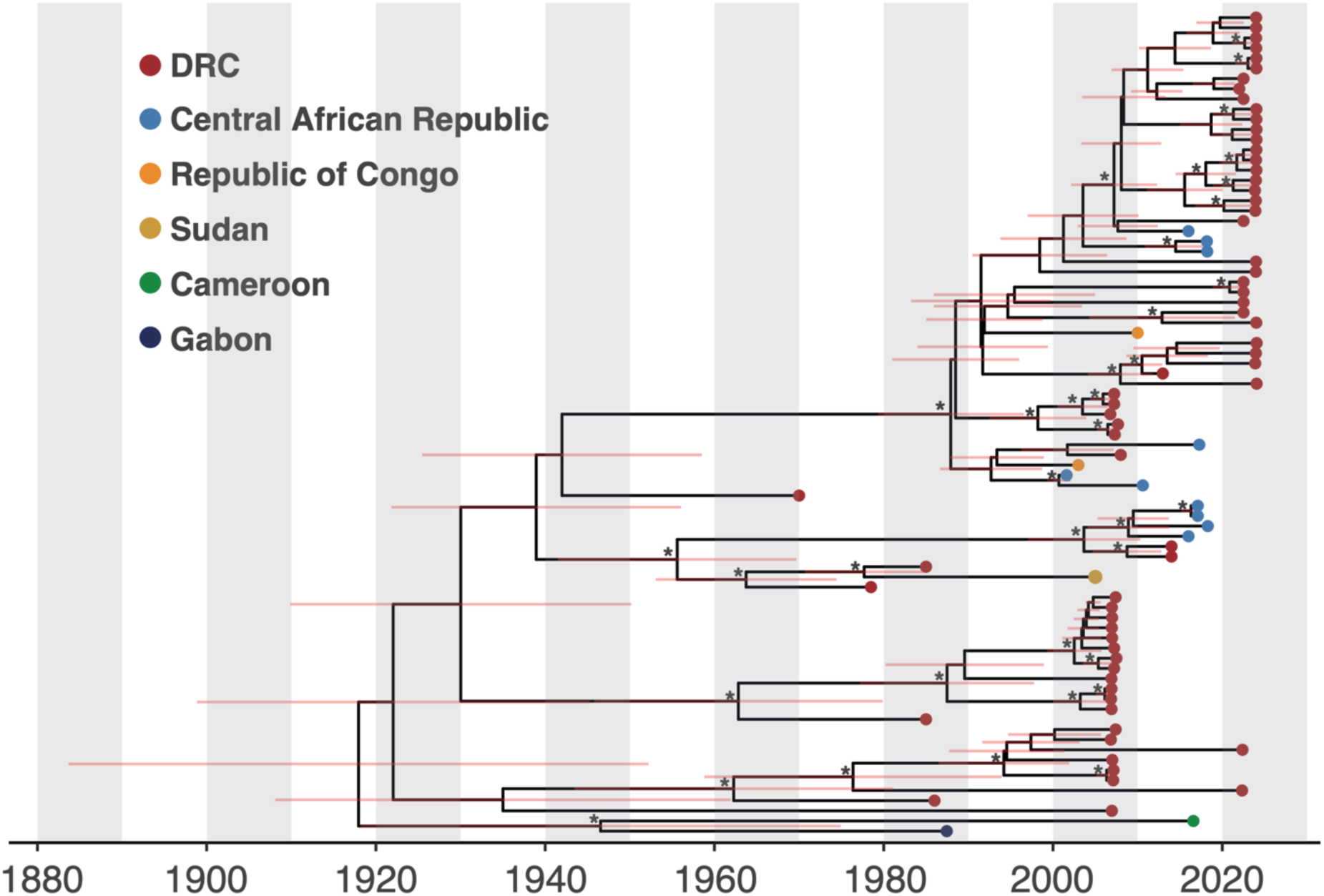
Temporal evolutionary history of clade I. The temporal maximum clade credibility phylogenetic tree is shown. Tips are coloured by country of isolation. Red bars show the 95% HPD on the date of the corresponding node. Asterisks show nodes with posterior support of 70 or higher.

Clade I has been isolated outside of the Congo Basin in Sudan in 2005 (in a region that is now part of South Sudan) and 2022 (**Figure 2A**). The two Sudan sequences cluster in the phylogenetic tree and share a ~10.5Kb duplication (**Figure S3**). We estimate that the Sudan sequences diverged from their closest sampled relative (a DRC sample from 1985) in 1978 (95% HPD 1969 – 1984); this lineage has therefore been sampled only in Sudan over roughly 46 years. It is likely that this lineage has circulated in the animal reservoir during this period as it exhibits 8.5% TC>TT mutations, highly similar to that expected in animals (8%) but far lower than expected from evolution in humans (85%).^9^ This is confirmed by the geographical spread (6 states in western, southern and eastern Sudan) of the 18 mpox cases that were laboratory confirmed in Sudan 2022 (**Figure S4**).

Clades IIa and IIb A both circulate in West Africa and have been exported to other regions (**Figures 1**, **4**). Clade IIa has not been observed outside West Africa since an outbreak in USA in 2003 (**Figure 1**). Samples of clade IIa from West Africa remain sparse with single sequences from Liberia and Sierra Leone from human cases in 1970, and two closely related clusters of sequences from chimpanzees in Cote d’Ivoire collected from 2017-2018 (**Figure 4A**).^7^ We therefore currently lack the resolution to examine spatial transmission patterns in more detail for clade IIa.

**Figure 4.**
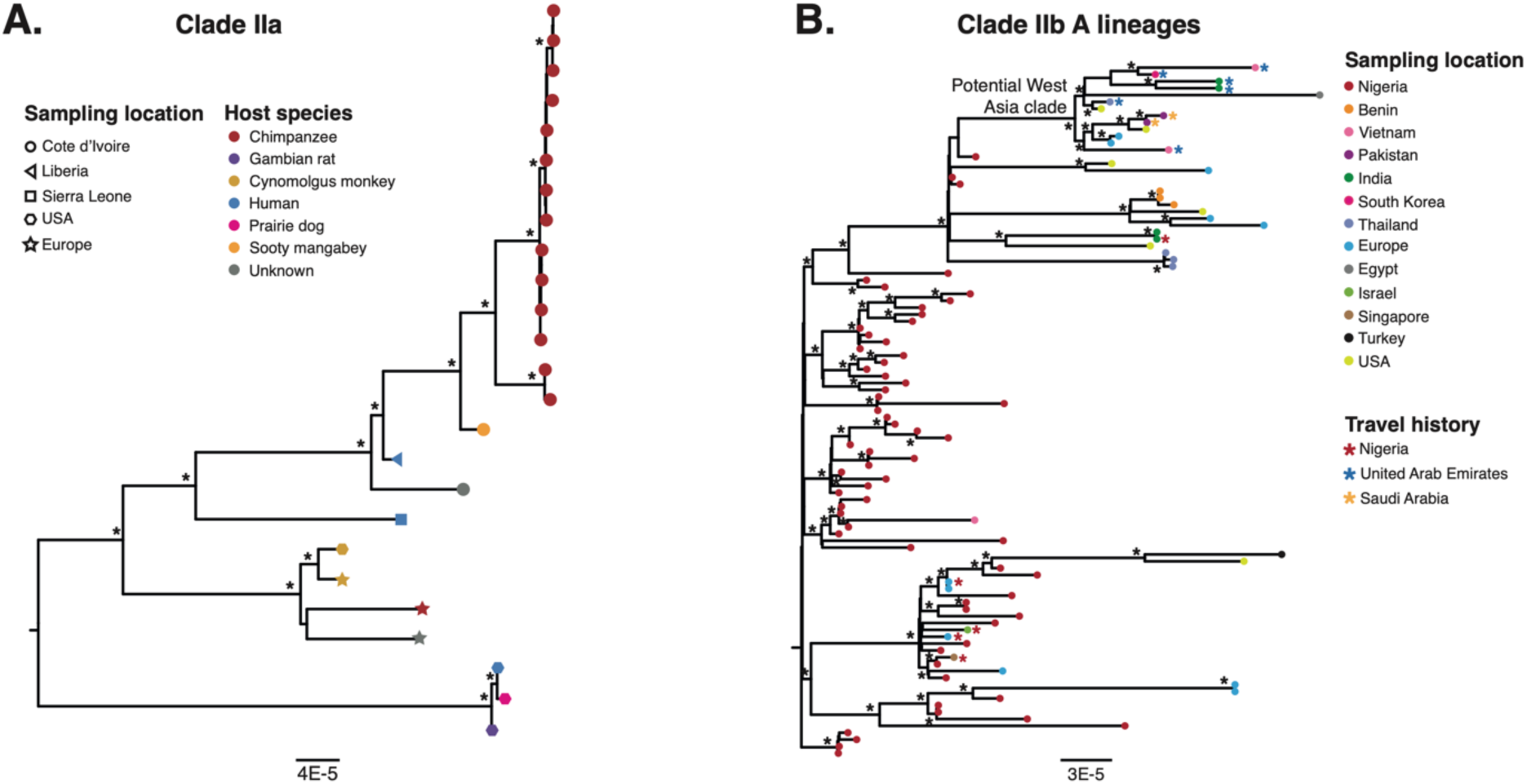
Spatial distributions of clades IIa and IIb A. (**A**) Maximum likelihood phylogenetic tree of 25 high quality clade IIa isolates. Tips are coloured by host species and shapes show sampling locations. Asterisks show nodes with bootstrap support of 70 or higher and the scale bar shows the expected number of nucleotide substitutions per site. (**B**) Maximum likelihood phylogenetic tree of 101 clade IIb A sequences. Tips are coloured by country of collection. Red, blue and orange asterisks show samples with travel history. The potential Eastern Mediterranean clade containing samples with travel history to United Arab Emirates and Saudi Arabia is highlighted. Black asterisks so nodes with bootstrap support of 70 or above and the scale bar shows the expected number of nucleotide substitutions per site.

While clade IIb A initially spread in Nigeria, it was then exported to other countries in Europe, Asia, North America and, more recently, North Africa (**Figures 1**, **4B**). Genetic sequences from multiple individuals infected with clade IIb A from United Kingdom, Israel, Singapore and India have travel history to Nigeria (**Figure 4B**), supporting infection in endemic regions of Nigeria and subsequent export. Additionally, eight sequences from India, South Korea, Vietnam and Thailand were isolated from travellers returning from United Arab Emirates or Saudi Arabia; these sequences cluster within a single phylogenetic lineage that also includes sequences from USA, UK, Slovenia and Egypt for which no information regarding recent travel is recorded (**Figure 4B**). This is consistent with sustained circulation of a lineage of clade IIb A in the Eastern Mediterranean region; no sequences are currently available from Eastern Mediterranean to examine this further.

### MPXV mutational spectra are influenced by transmission route

Previous studies have demonstrated that mutational spectra of MPXV lineages that transmit from human-to-human exhibit a high proportion of TC>TT mutations, implicating APOBEC-3 as a major driver of mutagenesis in human MPXV infections.^9^ We, for the first time, calculated complete single base substitution (SBS) mutational spectra for the dominant clades of MPXV and corrected these for genomic composition (**Figure 5**, **Methods**). Similar to previous studies,^9,19^ we find the spectrum of clade IIb (both A and B.1 lineages) to be dominated by C>T mutations where the C is preceded by a T, with preference for A or G following the substitution site (**Figure 5**).

**Figure 5.**
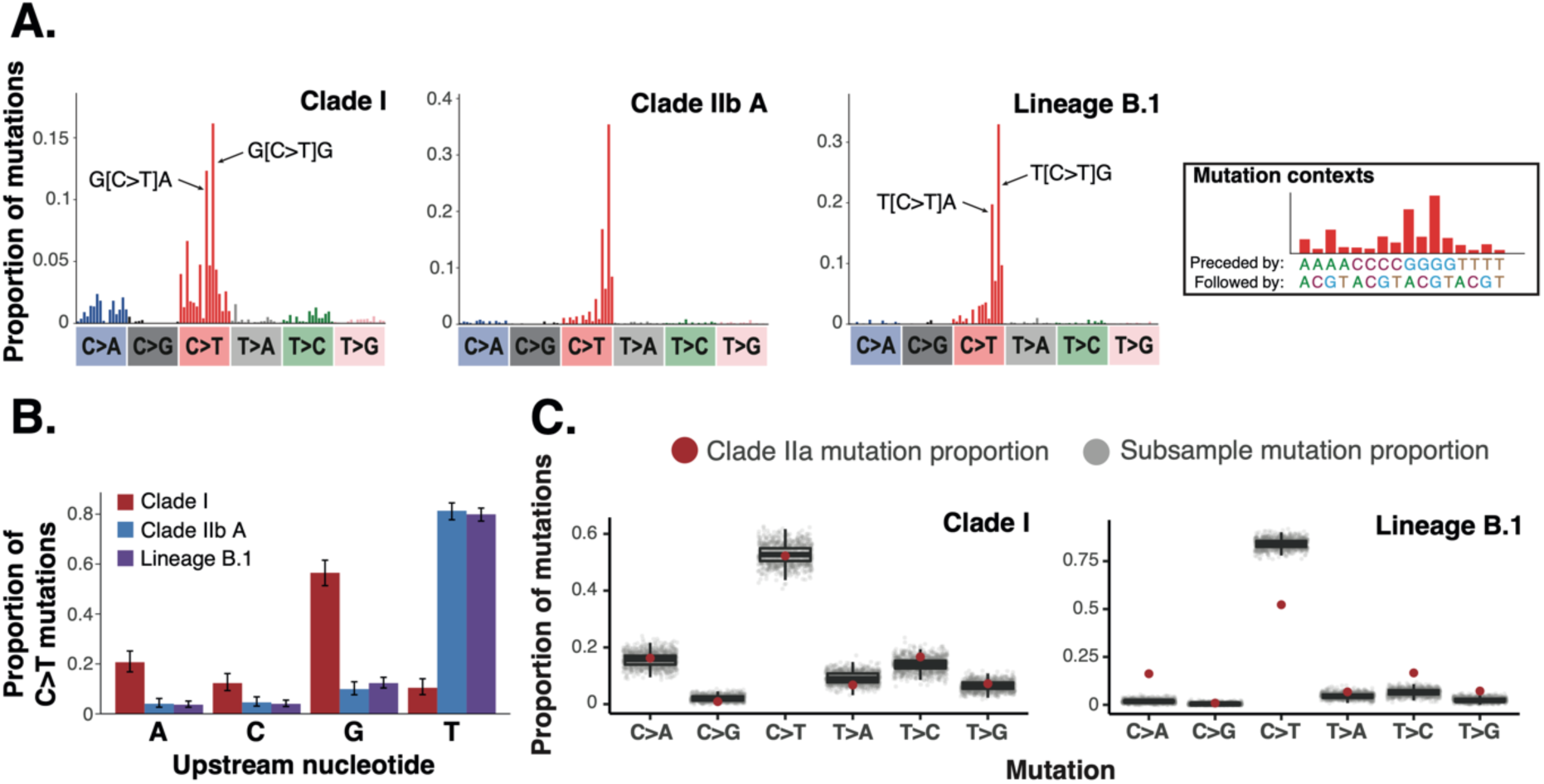
Mutational spectra differ between major MPXV clades. (**A**) Single base substitution (SBS) mutational spectra for clade I, clade IIb A and lineage B.1 (clade IIa not included because of insufficient mutations, see **Methods**). SBS spectra show the proportion of mutations of each mutation type within each surrounding nucleotide contexts; contexts for an example mutation type are shown in the right-hand panel. The two most prevalent contextual mutations are highlighted for clade I and lineage B.1. Mutational spectra are corrected for genome composition (see **Methods**). Symmetrical mutations (for example C>T and G>A) are combined as MPXV is a DNA pathogen.^30^ (**B**) The proportion of C>T mutations with each nucleotide upstream is shown for each clade. (**C**) To examine the potential for the clade IIa mutational spectrum to have been generated by the mutational spectra of clade I and lineage B.1, we compared the proportion of each mutation type in the clade IIa spectrum with that in 1000 subsamplings of the other clade spectrum to the number of mutations in the clade IIa spectrum (see **Methods**). Each grey point represents the mutation type proportion in one subsample of the respective mutational spectrum while the red point shows the mutation type proportion in clade IIa. The clade IIa mutation type proportions are within that expected from clade I but often outside that expected from lineage B.1. Boxplot centre lines show median value; upper and lower bounds show the 25th and 75th quantile, respectively; upper and lower whiskers show the largest and smallest values within 1.5 times the interquartile range above the 75th percentile and below the 25th percentile, respectively.

The clade I mutational spectrum shows that mutational processes within the animal reservoir are also dominated by C>T mutations (66% of mutational burden accounting for genome composition), but with different contextual preferences to evolution within humans (**Figure 5**). In clade I, C>T mutations occur most commonly where the C is preceded by G (57% of the C>T mutational burden, **Figure 5B**) and are most frequent in G[C>T]A and G[C>T]G contexts (**Figure 5A**). The C>T mutations are unlikely to be the result of spontaneous deamination of cytosine as this has a strong preference for CG>TG contexts in human DNA.^31^ The C>T mutations may therefore be the result of polymerase errors during genome replication, action of alternative APOBEC enzymes within the animal reservoir and/or additional mutagens. The clade IIb spectra exhibit similar enrichment of G over A and C as the nucleotide preceding C>T mutations (**Figure 5B**), which may suggest the GC>GT mutations are driven by a non-host species factor, but this will require additional future work to untangle.

C>A (which includes G>T mutations) is the second most common mutation in clade I (17% of the total mutational burden, **Figure 5**); reactive oxygen species are a potential driver of these mutations.^32^

The clade IIa mutation spectrum contains 222 mutations, which may be too low to examine mutational patterns in detail.^30^ We therefore used the clade I and lineage B.1 mutational spectra as references and investigated whether either could explain the clade IIa mutational spectrum. We found that the clade IIa mutational patterns could have been generated by the clade I spectrum but not by the lineage B.1 spectrum (**Figure 5C**). This suggests that clade IIa exhibits similar mutational processes as clade I, consistent with a lack of human APOBEC-3 activity and therefore both clades consistently circulating outside of humans.

## Discussion

Implementing measures to control MPXV cases will require in depth understanding of how the virus is transmitting within humans and animals across different spatial scales. We here carried out an in-depth analysis of spatiotemporal and host species patterns across all major MPXV clades. Our analysis revealed large differences in the spatiotemporal and host species patterns of the major clades. We identified regular international and inter-province transmission of clade I, including its likely circulation in animal reservoirs and/or endemicity in parts of eastern Africa, inferred transmission of clade IIb A in Eastern Mediterranean and showed that MPXV mutational patterns are associated with transmission route.

Our analysis provides the first evidence of regular recent transmission of clade I amongst countries and provinces within Central Africa (**Figures 2**, **3**). We also found co-circulation of multiple clade I lineages within individual countries and provinces (**Figures 2**, **3**). This demonstrates that high MXPV diversity is maintained within the clade I animal reservoir(s), suggesting high MPXV prevalence and thereby highlighting the potential for frequent spillover when humans interact with reservoir species. Our results also show that the animal reservoir is mobile. Understanding the speed and dynamics of virus movements may help to pinpoint possible reservoir species, as well as source locations, but this will require increased sequencing of epidemiologically representative samples in future.

While clades I and IIa both circulate within animals and spillover into humans, we observe highly different distributions of host species amongst sampled genetic sequences, with clade I mostly being sampled from humans and clade IIa from animals (**Figure 1C**). It is unclear whether this difference in host sampling is driven by distinct abilities of the clades to infect humans, differential disease severity in humans and/or animals altering likelihood of case detection, different contacts between humans/sampled animals and animal reservoirs (which may differ for the two clades) and/or different surveillance and sampling strategies in humans and animals within the affected countries. Identifying the driver(s) of differential host sampling will require stronger surveillance in humans and animals, and linkage with sample metadata to determine likely routes of infection.

While clade I has mostly been detected within the Congo Basin, our data highlight the potential for widespread endemic and/or enzootic circulation of this clade within Sudan (former Sudan, now divided into Sudan and South Sudan). We identified a clade I lineage that has been sampled only from Sudan over roughly 45 years (**Figures 2**, **3**). This lineage has likely circulated within the animal reservoir (due to a lack of APOBEC3-like mutations) and may have circulated continually in Sudan following introduction at any point during this time period or could have been introduced multiple times from an unsampled region either shortly before being sampled in Sudan or with some level of local transmission. Distinguishing between these possibilities will require additional sequencing data from Sudan. However, our results combined with recent epidemiological data highlight the potential for large numbers of MPXV cases in Sudan: following the declaration of the Public Health Emergency of International Concern in July 2022, the Federal Ministry of Health of Sudan, with support from WHO, started mpox surveillance and as a consequence, suspected mpox cases were reported from 12 states and 32 localities in Sudan, of which 5 states host refugees and internally displaced persons, and with >40% of suspected cases being children under five.^33^ Of the suspected cases, only a portion was tested and a total of 18 cases were laboratory confirmed from 6 states and 9 localities, and including one death (CFR 5.8%).^33^

Our data combined with the recent emergence of a novel human transmissible clade I lineage in South Kivu,^25^ where mpox cases have not previously been detected (except few cases in 2011/2012), highlights the potential for an increased risk of international spread of clade I MPXV. A recent study highlighted that the affected population in South Kivu had a median age of 22 years, 52% were females and 30% were sex workers, which represent a significant shift in the historical mpox epidemiology in the DRC, which involves children <15 years as the main affected age group.^13,25^ As a comparison, in the years preceding the eradication of smallpox (1956-1971), the maximum number of smallpox cases reported by the DRC (at that time Zaire) Ministry of Health to the WHO was 5,523 cases including 710 deaths in 1963,^34^ which is a third of the suspected mpox cases reported by the DRC in 2023.^13^ Both Sudan and DRC have reported cases in provinces that directly border with other African countries such as Burundi, Uganda, Rwanda, Angola, South Sudan, and Chad, and in some instances, such as for South Kivu province (DRC), at-risk populations are known to include regular cross-border commuters. Further studies to better understand transmission patterns in these settings (including if enzooticity has been established) and the geographical distribution of mpox in Eastern Africa are urgently needed.

The co-circulation of multiple clade I lineages in individual provinces combined with regular geographical movements and the potential for an East Africa clade I lineage suggests there is high prevalence and diversity of clade I within the animal reservoir(s). It is currently unclear whether this genotypic diversity is associated with phenotypic diversity. However, this high prevalence is likely to make control of clade I within the animal reservoir highly challenging. Prevention of human cases is therefore likely to require interventions at the human-animal interface and rapid detection and cessation of human-to-human transmission chains. Our results therefore underpin the importance of further studies to understand how humans become infected with clade I viruses and studies carrying out functional characterisation of diverse clade I viruses.

Local human-to-human transmission chains were established in many countries across all six WHO regions during the 2022 MPXV global outbreak.^11^ Local transmission of the ancestral clade IIb A lineages has also been identified in some cases outside of West Africa.^35^ We were here able to infer local transmission of clade IIb A MPXV in Eastern Mediterranean through travel data associated with sequences from other countries, despite sequences from that region being unavailable for analysis. This highlights the importance of associating detailed metadata with genetic sequences where possible.

Mutational signatures have provided major insights into MPXV and can be used to identify lineages that are transmitting from human-to-human and to infer outbreak origin dates.^9,19^ We here calculated the most in depth mutational spectra of MPXV to date and compared these across spectra, showing that C>T mutations are most common within both human and animal hosts, but differences in contextual preferences exist between species (**Figure 5**). Identifying the drivers of spectrum differences will require future work, but our analyses suggest that the ratio between GC>GT mutations and TC>TT mutations is a reliable marker to distinguish human-to-human transmission from transmission in animals, therefore potentially being able identify sustained human outbreaks from transmission from the reservoir.

The global outbreak has provided critical insights into the epidemiology of mpox in humans. However, it is unknown whether characteristics of lineage B.1 were acquired following adaptation in humans, or whether they may be generalisable across MPXV clades. This can only be revealed with stronger surveillance, sharing of sequences and continued genotypic and functional characterization of the differences between clades and lineages.^36^ Such characterization would shed light into the drivers of epidemiological differences between clades, but this work is technically challenging and currently few laboratories worldwide have such capacity. However, a small number of studies have found that the apparent difference in morbidity and mortality between clades I and II is likely driven by multiple proteins present in clade I but absent in clade II.^37,38^ One particular area of interest is gene duplication, such as that found in clade I sequences from Sudan. Such duplications have been described for clade IIb sequences from the 2022 outbreak, when they were assumed to be involved in immune evasion and host range.^21,39,40^ Gene duplication and loss in the MPXV terminal regions are also considered drivers of poxvirus evolution and adaption to the host.^21,22^ Analyses comparing strains with and without deletions will be essential to uncover their functional consequences and understand/forecast the epidemiology of lineages exhibiting such changes. In addition, deletions may lead to diagnostic failure, especially for nucleic acid amplification tests that target less conserved genes, such as some of the clade-specific PCRs, which are designed to distinguish clades. To date, there have been two reported examples of such diagnostic failure episodes, one for a variant of MPXV clade IIb detected in the US that did not spread widely, ^41^ and one for the clade I lineage currently circulating in South Kivu.^15^ This clearly highlights the critical importance of a strategic genomic surveillance system where mpox circulates.

As highlighted in the standing recommendations for mpox issued by the Director General of the WHO, it is critical that countries have national mpox strategic plans integrated into broader health systems, and that capacities that have been built in resource-limited settings and among marginalized groups should be sustained.^42^ Without surveillance, no genomic sequence data can be generated, and no virological characterization of circulating clades and lineages can be done. In this regard, more laboratories should engage in virological characterization of MPXV clades and lineages. Furthermore, countries are strongly encouraged to continue documenting and making sequences publicly available, prioritising specimens for both targeted sequencing (for example of imported cases, the first few cases of local emergence and cases with divergent demographic or clinical profiles) and representative sequencing. This will enable tracking of virus circulation and evolution over time. If we want to prevent the next mpox global outbreak, it is now time to strengthen mpox surveillance, including in Africa, focusing on populations at highest risk and ensuring integration with existing systems to ensure comprehensive and seamless delivery of care.

## Competing interests

No competing interests declared

## Data availability statement

All sequence accessions and inferred phylogenetic trees are available to be publicly shared without any restrictions.

## Disclaimer

The findings and conclusions in this report are those of the author(s) and do not necessarily represent the views of the funding agencies

## Online Methods

### Dataset assembly and filtering

We aimed to collate a dataset containing all available high quality MPXV genetic sequences. To do this, we initially downloaded all MPXV nucleotide sequences from GenBank (identified as sequences containing at least one of the search terms “monkeypox”, “mpox”, “MPXV” and “MPV”, *n*=7,252) and the Global Initiative for Sharing of All Influenza Data (GISAID) EpiPox database (*n*=8,843) as of 17 February 2024. We then combined these datasets and filtered to remove duplicate and lower quality sequences. To do this, we initially removed sequences containing fewer than 30,000 nucleotides (nt), with this cutoff chosen to retain historical MPXV sequences that were ~32,000 nt in length. We next discarded sequences with >20% indeterminate bases (Ns) and assigned clades and (for clade IIb) Pango lineages using Nextclade.^43^ Sequences that could not be assigned a clade were assumed to be low quality and were excluded from further analysis. Based on the clade and lineage assignments from Nextclade, we divided the sequence dataset into four groups: clade I, clade IIa, clade IIb A (containing sequences from the A sublineages within clade IIb but not those within lineage B.1 and its descendent lineages) and lineage B.1 (containing sequences from lineage B.1 and its descendent lineages).

We identified sequences that were duplicated between GenBank and GISAID initially by identifying sequences with the same sample name, country, and collection date. After removing one of each of these sequence pairs, we carried out an additional phylogenetic screen for duplicate sequences in the clade I, clade IIa and clade IIb A datasets. Sequences within each of the four datasets were aligned using squirrel v0.1 (https://github.com/aineniamh/squirrel),^9^ specifying the clade to which the sequences belong. We then reconstructed a phylogenetic tree for each clade using IQ-TREE v2.1.3, ^44^employing a JC model of nucleotide substitution. We identified closely related pairs of sequences in the resulting phylogenetic trees and checked their sequence names, countries and collection dates to determine whether they might be duplicates. Where the sequence names and collection dates were similar (i.e. the sequence names contained shared elements and the collection dates were the same to the most accurate level possible), we retained one of the sequences.

We carried out a further quality control check by analysing the root-to-tip distances of sequences compared to their collection date.^45^ For clade I, clade IIa and clade IIb A, we aligned sequences with squirrel v0.1 and reconstructed maximum likelihood phylogenetic trees using IQ-TREE v2.1.3 as above but including an outlier sequence (outlier accession numbers KJ642617.1 for clade I, KJ642616.1 for clade IIa and clade IIb A) that was used to root the tree. We then identified sequences that were clear outliers in a root-to-tip plot of the rooted tree in TempEst v1.5.3,^45^ and removed these from further analyses. Due to the large size of the lineage B.1 dataset (n = 10369 sequences prior to filtering), instead of reconstructing a phylogenetic tree, we carried out a root-to-tip-like analysis by comparing collection date with mutation distance to sample ON676708.1, which clusters immediately upstream of lineage B.1. We aligned the lineage B.1 dataset and ON676708.1 using squirrel v0.1 as above and calculated the number of mutations between each sequence and ON676708.1. This showed a strong correlation (**Figure S5**) so we generated a linear model between sample collection date and this mutation distance and removed samples whose residual within the linear model is more than five times the median absolute deviation away from the median residual (**Figure S5**). These filtering steps resulted in final datasets of 113 clade I sequences, 25 clade IIa sequences, 101 clade IIb A sequences and 10,307 lineage B.1 sequences. The majority of these sequences contain close to the complete genome (**Figure S6, Tables S1, S2**). In addition, we have added 47 sequences from a GitHub directory of a recent paper that describes highly divergent viruses.^16^

We identified the spatiotemporal and host species distributions of sequences within each dataset using location, collection data and host species metadata associated with the sequence accession on either GenBank or GISAID. Where this metadata was missing, we attempted to identify it within original publications.

### Reconstruction of phylogenetic trees

To visualise the phylogenetic relationships between the clades, we reconstructed a tree containing all sequences from clades I, IIa and IIb A and the sublineage references for each of the sublineages within B.1 (accession numbers obtained from https://github.com/mpxv-lineages/lineage-designation/blob/master/auto-generated/lineages.md). We aligned these sequences using squirrel v0.1 with clade set to clade II and reconstructed a maximum likelihood phylogenetic tree as above.

To examine phylogenetic relationships within each clade, we aligned sequences within the respective dataset using squirrel v0.1 with clade set to clade I for the clade I dataset and set to clade II for the remaining datasets. Final maximum likelihood phylogenetic trees were reconstructed for each dataset using IQ-TREE v2.1.3 as above. Topological robustness was assessed using 1000 bootstrap replicates. Travel histories for Clade IIb A were identified in GISAID metadata and from examination of original publications.

Phylogenetic trees were visualised using FigTree v1.4.4 and ggtree v3.0.2.^46^

### Reconstruction of the temporal history of Clade I

We aimed to reconstruct the temporal history of clade I. The recently identified outbreak in South Kivu contains evidence of APOBEC3 mutagenesis,^16^ which increases the substitution rate^9^ and may make the application of single clock model unreliable. We therefore did not include the sequences from this outbreak in the temporal reconstruction.

Methods to infer temporal history are only valid if there is a temporal signal within the dataset.^45^ We assessed temporal signal using root-to-tip randomisation where we compared the R2 correlation between sample collection date and root-to-tip distance with that in 1000 randomisations of collection dates. This supported the presence of a temporal signal (*P* < 0.001). We therefore reconstructed the temporal history of clade I using BEAST v2.6.6 employing the JC69 model of nucleotide substitution. We used a relaxed log-normal clock model with a log-normal prior on the substitution rate with mean 1.9E-6 (chosen to match the estimated slope in TempEst) and standard deviation 0.5. Population history was modelled using a coalescent constant population prior. Four independent runs were carried out for 150,000,000 total MCMC steps. Convergence was assessed using Tracer v1.7 and all ESS values were above 550. 10% burnin was removed from each run before the runs were combined and the maximum clade credibility tree identified and annotated using TreeAnnotator.

### Identification of genomic rearrangements

Genome rearrangements in the Sudan clade I sequences were identified as described in Brinkmann et al.^20^

### Calculation of mutational spectra

We calculated a single base substitution mutational spectrum for each of the major MPXV clades using the sequence alignments and maximum likelihood phylogenetic trees generated above and containing all high-quality sequences. Phylogenetic trees were outgroup rooted using the outgroups described above to enable the direction of each mutation (i.e. the ancestral and mutated nucleotides) to be robustly identified. The outgroup was removed prior to spectrum calculation. We reconstructed mutational spectra using MutTui v2.0.2 (https://github.com/chrisruis/MutTui).^30^ We rescaled the resulting mutational spectra using MutTui v2.0.2 to account for the number of A, C, G and T nucleotides and the distribution of nucleotide triplets across the genome.^30^

It has previously been suggested that a dataset requires at least 300-600 mutations for the mutational spectrum to be accurately estimated.^30^ The clade I, clade IIb A and lineage B.1 datasets each contain more than 600 mutations. However, the clade IIa spectrum contains 222 mutations so we did not attempt to examine the detailed contextual patterns in these mutations. To estimate whether the clade IIa mutational spectrum could have been generated by the spectrum of clade I or clade IIb, we compared the mutation type proportions in the clade IIa spectrum with those in 1000 random downsamplings of the clade I and lineage B.1 mutational spectra to 222 mutations.

### Data availability

Sequence alignments (excluding GISAID sequences), BEAST log files and mutational spectra are available at https://github.com/chrisruis/Mpox_global. GenBank sample accessions and metadata are listed in **Table S1**. GISAID sample accessions and acknowledgements are listed in **Table S2**.

## Supplementary figures

**Figure S1.**
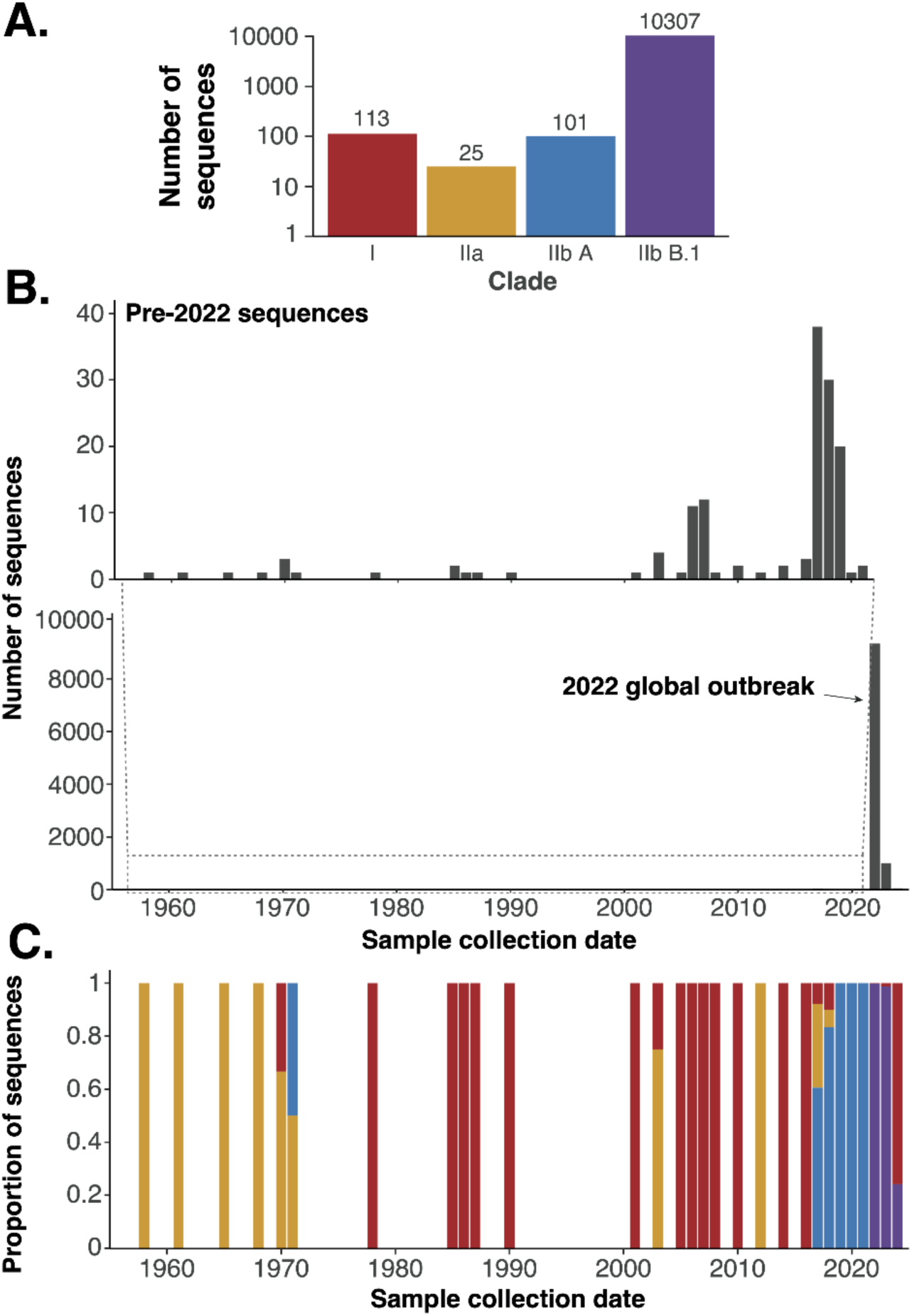
Distribution of MXPV genetic sequences. (**A**) The number of high-quality MXPV genetic sequences (see **Methods**) is shown for each major clade. (**B**) The number of MXPV sequences from each year is shown. Due to the large number of sequences in 2022 compared to other years, we extract the years before 2022 in the upper panel. (**C**) The proportion of sequences belonging to each major MPXV clade in each year is shown, colours match those in panel **A**.

**Figure S2:**
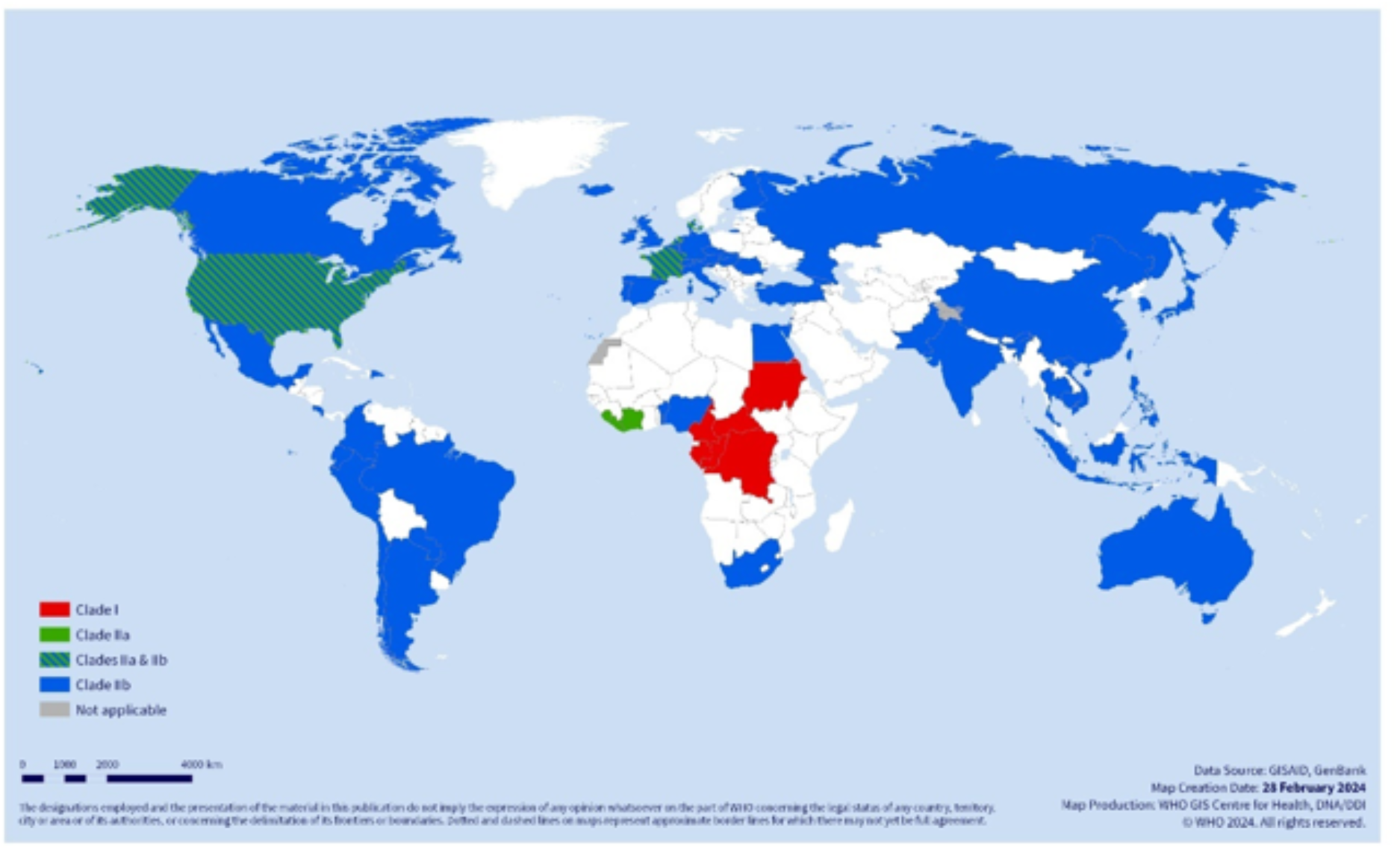
Geographic distribution of MPXV clades across WHO regions. The map was generated from 10,670 sequences obtained from GenBank and GISAID, 1958-February 2024. Clade I is red, clade IIa is green, and clade IIb is blue.

**Figure S3:**
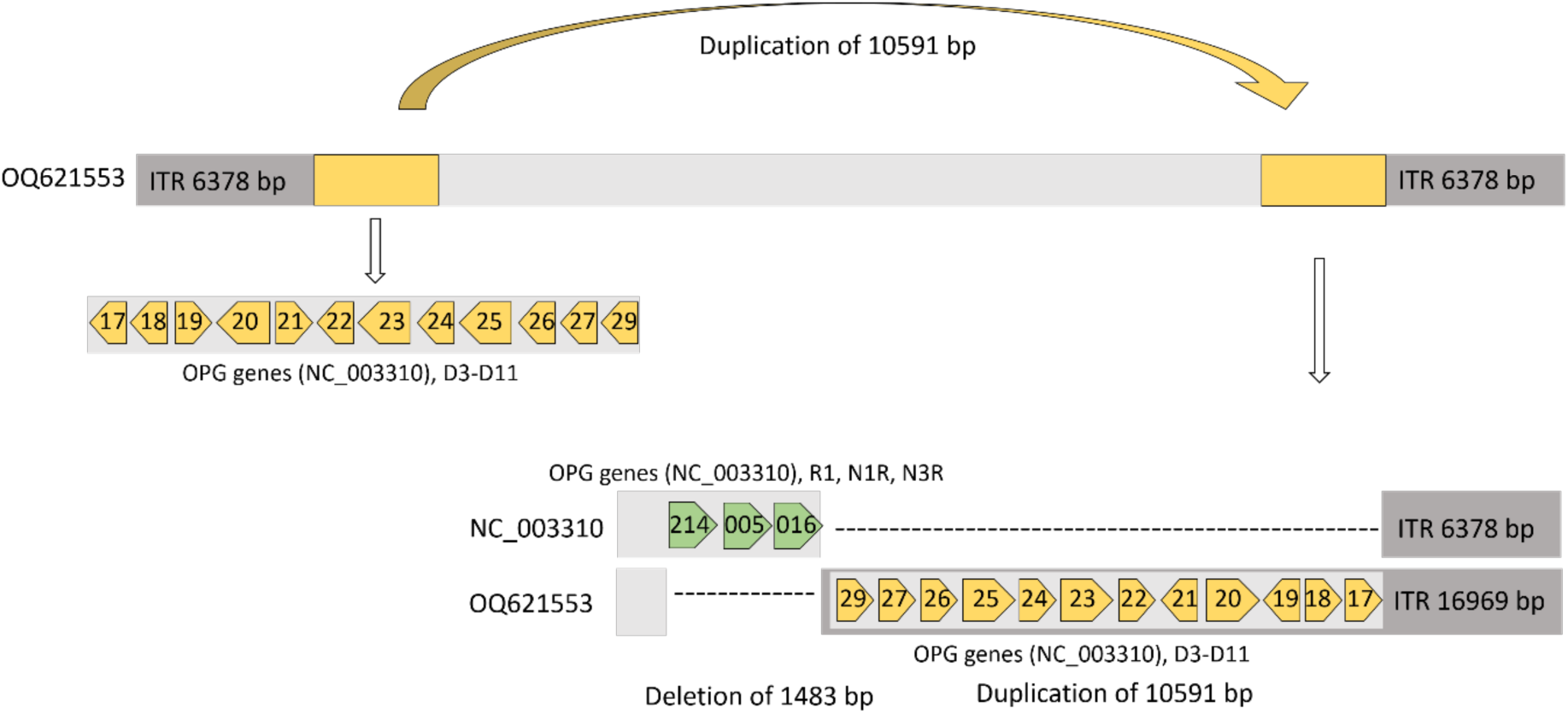
Duplication of the genome from Sudan 2005 and 2022 (KC257459, OQ621553). A 10591 bp region directly downstream the left ITR is duplicated to the right site of the genome, resulting in ITRs of 16969 bp and a genome length of 204,808 bp). Three genes (R1, N1R and N3R) are deleted at the site of duplication.

**Figure S4.**
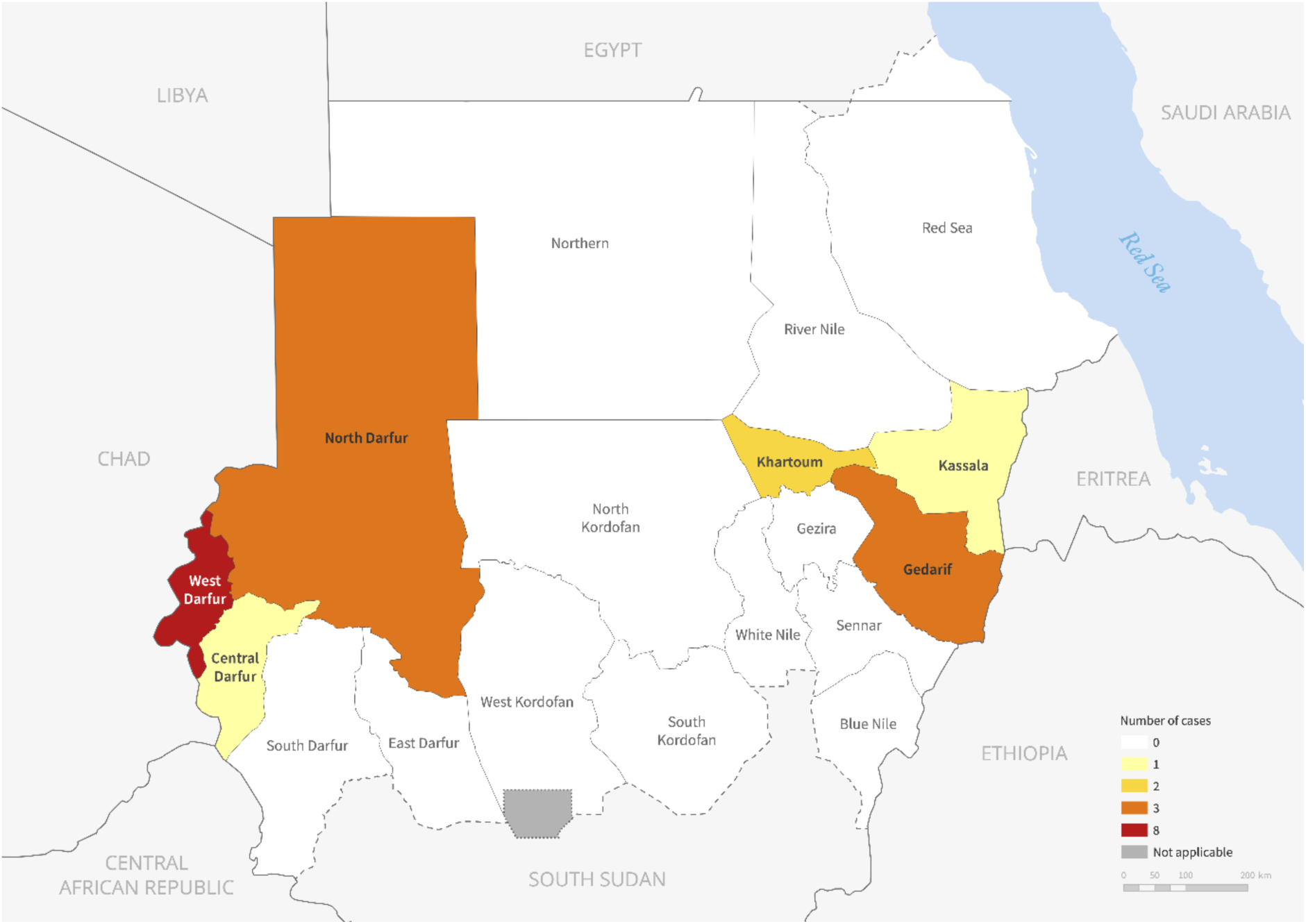
Geographical distribution of laboratory confirmed cases of mpox in Sudan in 2022. Confirmed cases were reported in Sudan states neighbouring Chad, Central African Republic, Ethiopia and Eritrea, while cases in 2005 were reported from a region that now belong to South Sudan.

**Figure S5.**
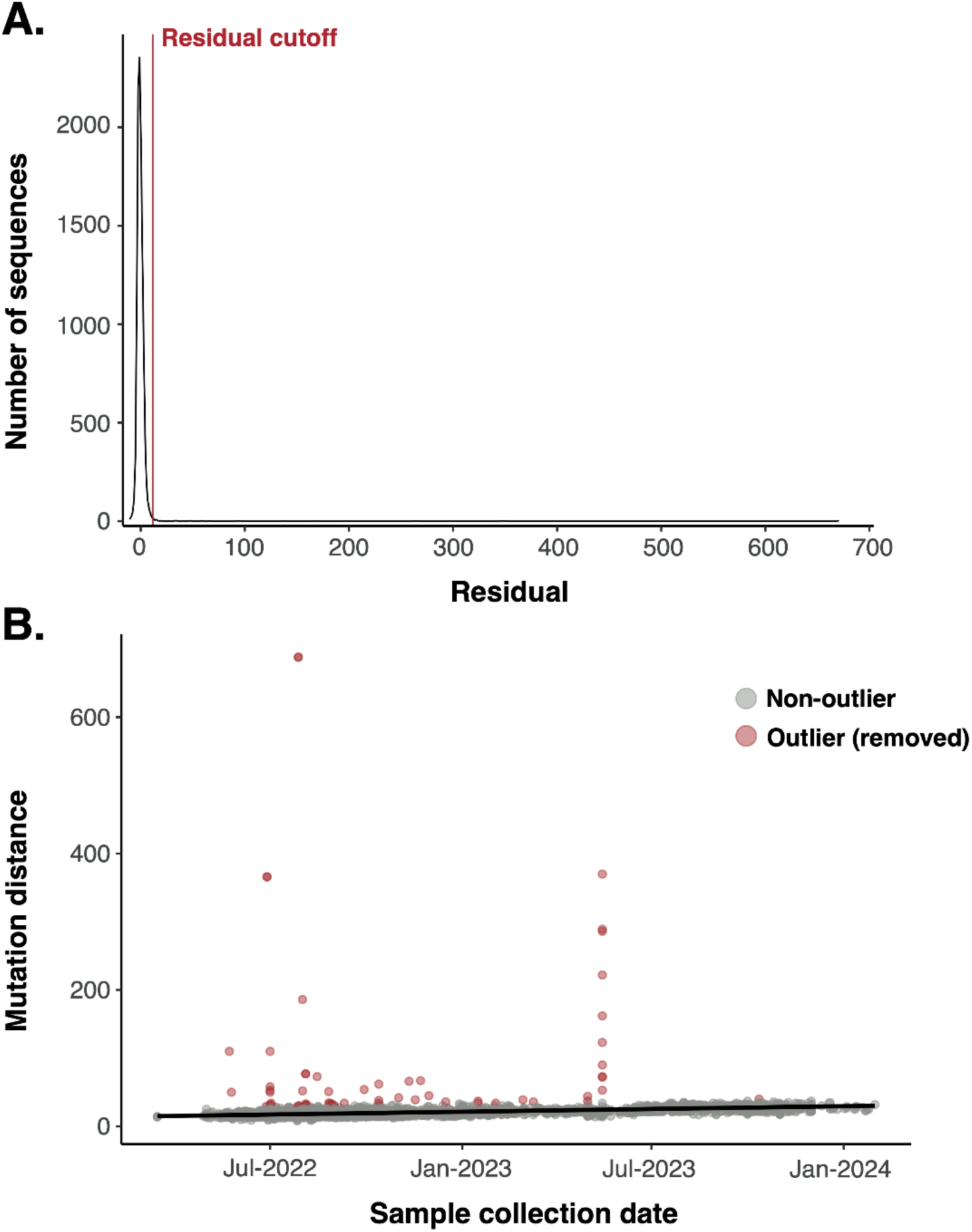
Identification of lineage B.1 outliers. (**A**) The distribution of residuals around the best fit line shown in **B**. The red vertical line shows five median absolute deviations above the median residual which was used as a cutoff to identify outliers. (**B**) The collection date of each lineage B.1 sequence is plotted against the number of mutations between the sequence and sample ON676708.1 which clusters immediately upstream of lineage B.1. Sequences shown in red contain greater diversity than expected given their sampling data so were excluded from further analyses.

**Figure S6.**
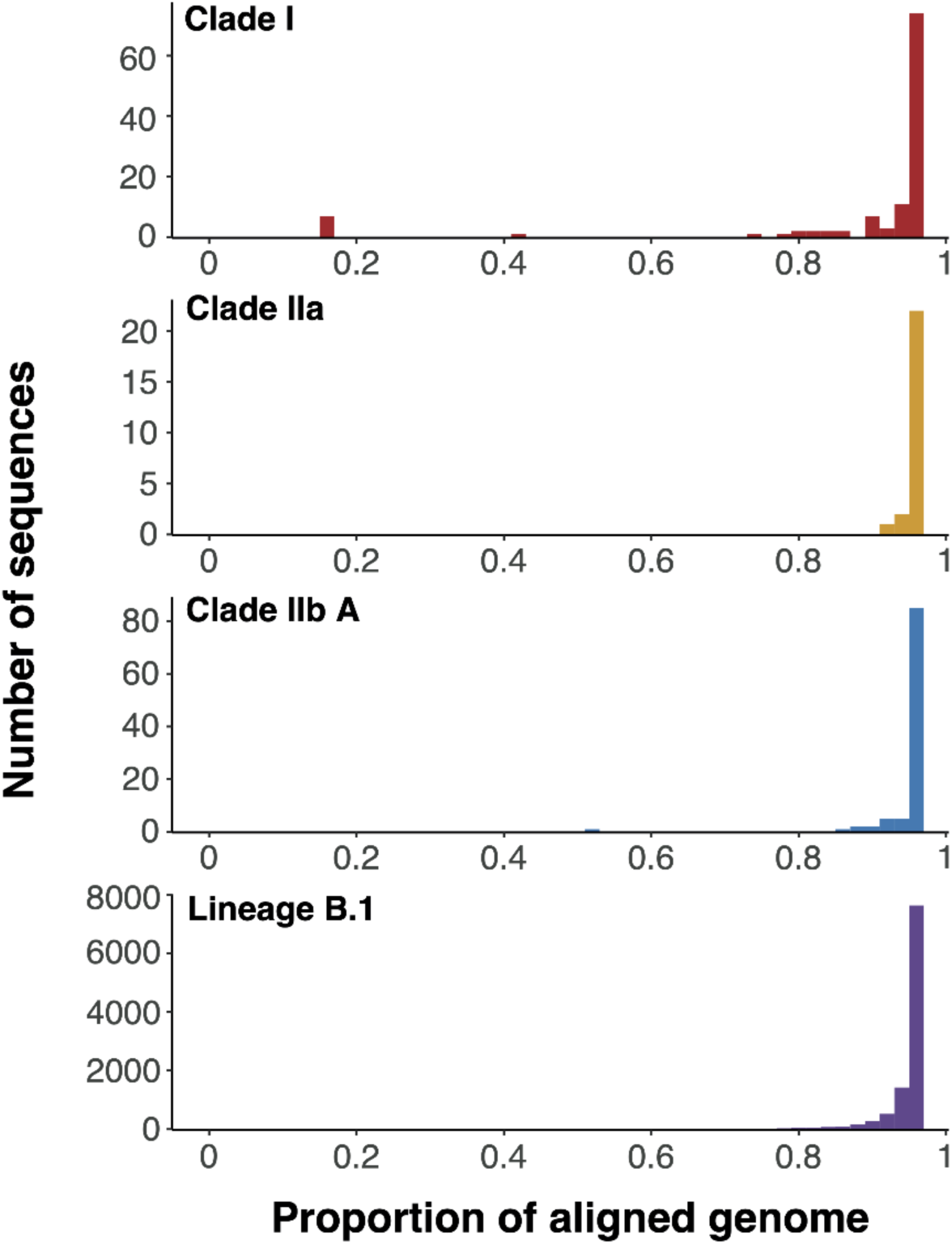
Most included MPXV sequences contain close to the complete genome. The proportion of the total genome covered is plotted for each sequence retained after filtering, split by major clade. Note that due to masking of ITR regions during alignment,^9^ no sequence will contain the complete genome.

**Table S1. GenBank sample accessions and metadata**

**Table S2. GISAID sample accessions and acknowledgements**

## Notes

### Competing Interest Statement

The authors have declared no competing interest.

### Funding Statement

This study did not receive any funding

