## Supplementary Table S2 for "Global genomic surveillance of monkeypox virus"

We gratefully acknowledge the following Authors from the Originating laboratories responsible for obtaining the specimens, as well as the Submitting laboratories where the genome data were generated and shared via GISAID, on which this research is based.

All Submitters of data may be contacted directly via www.gisaid.org

Authors are sorted alphabetically.

| Accession ID | Originating Laboratory | Submitting Laboratory | Authors |
|---|---|---|---|
| EPI_ISL_13056893 | Instituto Nacional de Saude Doutor Ricardo Jorge (INSA) | Instituto Nacional de Saude Doutor Ricardo Jorge (INSA) | Joana Isidro, Vítor Borges, Miguel Pinto, Daniel Sobral, João Dourado Santos, Alexandra Nunes, Verónica Mixão, Rita Ferreira, Daniela Santos, Silvia Duarte, Luís Vieira, Maria José Borrego, Sofia Núncio, Isabel Lopes de Carvalho, Ana Pelerito, Rita Cordeiro, João Paulo Gomes |
| EPI_ISL_13069002 | Erasmus Medical Center Department of Virology | Erasmus Medical Center Department of Virology | Bas Oude Munnink, Marjan Boter, Babette Weller, Richard Molenkamp, Janette Rahamat-Langendoen, Reina Sikkema, Marion Koopmans |
| EPI_ISL_13106454 | Hospital General Universitario Gregorio Marañón | Hospital General Universitario Gregorio Marañón | Sergio Buenestado Serrano, Rosalía Palomino Cabrera, Daniel Peñas Utrilla, Jorge Rodríguez-Grande, Pedro Sola Campoy, Laura Pérez-Lago, Cristina Rodriguez-Grande, Marta Herranz Martin, Julia Suárez, Pilar Catalán, Patricia Muñoz, Darío García de Viedma |
| EPI_ISL_13157812 | Laboratorio di Microbiologia e Virologia, Università Vita-Salute San Raffaele, Milano | Laboratorio di Microbiologia e Virologia, Università Vita-Salute San Raffaele, Milano | Benedetta Giuliani, Sofia Sisti, Michela Sampaolo, Elena Criscuolo, Matteo Castelli, Roberto Ferrarese, Roberta Antonia Diotti, Massimo Locatelli, Massimo Clementi, Nicasio Mancini, Nicola Clementi |
| EPI_ISL_13158444 | Laboratorio di Microbiologia e Virologia, Università Vita-Salute San Raffaele, Milano | Laboratorio di Microbiologia e Virologia, Università Vita-Salute San Raffaele, Milano | Sofia Sisti, Michela Sampaolo, Elena Criscuolo, Benedetta Giuliani, Matteo Castelli, Roberto Ferrarese, Martina Libera, Massimo Locatelli, Massimo Clementi, Nicasio Mancini, Nicola Clementi |
| EPI_ISL_13159759 | Laboratorio di Microbiologia e Virologia, Università Vita-Salute San Raffaele, Milano | Laboratorio di Microbiologia e Virologia, Università Vita-Salute San Raffaele, Milano | Roberto Ferrarese, Benedetta Giuliani, Elena Criscuolo, Sofia Sisti, Michela Sampaolo, Matteo Castelli, Roberta Antonia Diotti, Massimo Locatelli, Massimo Clementi, Nicasio Mancini, Nicola Clementi |
| EPI_ISL_13191438 | Instituto de Infectologia Emilio Ribas | Instituto Adolfo Lutz Strategic Laboratory | Claudio Tavares Sacchi, Karoline Rodrigues Campos, Marlon Benedito Nascimento Santos, Alex Domingos Reis, Ariadne Ferreira Amarante, Adriano Abbud, Adriana Bugno, Walkiria Delnoro Almeida Prado, Regiane Cardoso de Paula |
| EPI_ISL_13234112 | Laboratório Central de Saúde Pública do Estado do Rio Grande do Sul | Instituto Adolfo Lutz Strategic Laboratory | Claudio Tavares Sacchi, Karoline Rodrigues Campos, Adriano Abbud, Adriana Bugno |
| EPI_ISL_13242738 | Hospital General Universitario Gregorio Marañón | Hospital General Universitario Gregorio Marañón | Sergio Buenestado Serrano, Rosalía Palomino Cabrera, Daniel Peñas Utrilla, Jorge Rodríguez-Grande, Pedro Sola Campoy, Laura Pérez-Lago, Cristina Rodriguez-Grande, Marta Herranz Martin, Julia Suárez, Pilar Catalán, Patricia Muñoz, Darío García de Viedma |
| EPI_ISL_13244349, EPI_ISL_13244610 | Erasmus Medical Center Department of Virology | Erasmus Medical Center Department of Virology | Bas Oude Munnink, Marjan Boter, Babette Weller, Richard Molenkamp, Janette Rahamat-Langendoen, Reina Sikkema, Marion Koopmans |
| EPI_ISL_13302316 | Laboratory of Clinical Microbiology, Virology and Bioemergencies. ASST-Fatebenefratelli-Sacco, L.Sacco University Hospital | Army Medical and Veterinary Research Center | Silvia Fillo, Riccardo De Sanctis, Giovanni Faggioni, Andrea Ciammaruconi, Anna Anselmo, Vanessa Vera Fain, Simone Di Sabatino, Francesco Giordani, Antonella Fortunato, Rossella Brandi, Giulia Campoli, Marzia Cavalli, Anella Monte, Martina Lipari, Maria Di Spirito, Giorgia Grilli, Silvia Chimienti, Giandomenico Cerreto, Filippo Molinari, Giancarlo Petralito, Davide Mileto, Valeria Micheli, Maria Rita Gismondo, Florigio Lista |
| EPI_ISL_13314740 | Laboratorio de Vigilancia em Saude de Vinhedo | Instituto Adolfo Lutz Strategic Laboratory | Claudio Tavares Sacchi, Karoline Rodrigues Campos, Adriano Abbud, Adriana Bugno |
| EPI_ISL_13331598 | Department for Virology, Molecular Biology and Genome Research, R. G. Lugar Center for Public Health Research, National Center for Disease Control and Public Health (NCDC) of Georgia | Department for Virology, Molecular Biology and Genome Research, R. G. Lugar Center for Public Health Research, National Center for Disease Control and Public Health (NCDC) of Georgia | Giorgi Tomashvili, Salome Javashvili, Meri Pantsulaia, Gvantsa Brachveli, Ana Papkiauri, Giorgi Gogoladze, Gvantsa Chanturia, Adam Kotorashvili, Maia Alkhazashvili, Khatuna Zakhashvili, Paata Imnadze, Amiran Gamkrelidze. |
| EPI_ISL_13338028 | Clinical Virology Unit, Department of Clinical Sciences, Institute of Tropical Medicine of Antwerp | Clinical Virology Unit, Department of Clinical Sciences, Institute of Tropical Medicine of Antwerp | Antonio Mauro Rezende*, Tessa de Block*, Sandra Coppens, Eric Florence, Maartje van Frankenhuijsen, Stefanie Bracke, Isabel Brosius, Laurens Liesenborghs, Patrick Soentjens, Kevin Ariën, Marjan Van Esbroeck, Philippe Selhorst*, Koen Vercauteren* *equal contribution |
| EPI_ISL_13339105 | Microbiology Service, Hospital Universitario Clinico San Cecilio, Granada | Microbiology Service, Hospital Universitario Clinico San Cecilio, Granada | Chueca N, de Salazar A, Viñuela L, Fuentes A, Casimiro-Soriguer CS, Perez-Florido J, Dopazo J, Garcia F |
| EPI_ISL_13342823 | Clinical Virology Unit, Department of Clinical Sciences, Institute of Tropical Medicine of Antwerp | Clinical Virology Unit, Department of Clinical Sciences, Institute of Tropical Medicine of Antwerp | Philippe Selhorst, Antonio Mauro Rezende, Tessa de Block, Sandra Coppens, Eric Florence, Isabel Brosius, Laurens Liesenborghs, Kevin Ariën, Marjan Van Esbroeck, Chris Kenyon, Koen Vercauteren |
| EPI_ISL_13343634 | Instituto de Infectologia Emilio Ribas | Instituto Adolfo Lutz Strategic Laboratory | Claudio Tavares Sacchi, Karoline Rodrigues Campos, Adriano Abbud, Adriana Bugno |
| EPI_ISL_13343697 | Fleury Medicina Dignóstica | Instituto Adolfo Lutz Strategic Laboratory | Claudio Tavares Sacchi, Karoline Rodrigues Campos, Adriano Abbud, Adriana Bugno |
| EPI_ISL_13343718 | Hospital Santa Ignes | Instituto Adolfo Lutz Strategic Laboratory | Claudio Tavares Sacchi, Karoline Rodrigues Campos, Adriano Abbud, Adriana Bugno |
| EPI_ISL_13351002 | B.C. Centre for Disease Control Public Health Laboratory | B.C. Centre for Disease Control Public Health Laboratory | John Tyson, Tracy Lee, Anthea Lam, Josh Quick, Agatha Jassem, Natalie Prystajecky, Linda Hoang, Inna Sekirov, Catherine Hogn, Frankie Tsang, Mel Krajden |
| EPI_ISL_13362760, EPI_ISL_13362764 | Laboratorio di Epidemiologia Molecolare e Sanità Pubblica-Policlinico Bari | Istituto Zooprofilattico Sperimentale della Puglia e della Basilicata | Parisi A, Simone D, Capozzi L, Del Sambro L, Bianco A, Chironna M, Loconsole D, Sallustio F, Galante D, Pace L, Manzulli V, Fasanella A. |
| EPI_ISL_13363142 | Hospital Universitari Vall d'Hebron | Hospital Universitari Vall d'Hebron | Maria Piñana, Cristina Andrés, Alejandra González-Sánchez, Damir Garcia-Cehic, Ariadna Rando, Juliana Esperalba, Maria Gema Codina, Maria Carmen Martin, Carla Castillo, Karen García, Rodrigo Vásquez, Maria Piquer, Tomàs Pumarola, Josep Quer, Andrés Antón |
| EPI_ISL_13374487 | National Public Health Center, National Biosafety Laboratory | National Public Health Center, National Biosafety Laboratory | Judit Henczkó, Dániel Déri, Fruzsina Petrovay, Lili Jármi, Bernadett Pályi, Eszter Balla, Zoltán Kis |
| EPI_ISL_13409177, EPI_ISL_13409178, EPI_ISL_13409179, EPI_ISL_13409180 | Viral Genomics and Bioinformatics, MRC University of Glasgow Centre for Virus Research | Viral Genomics and Bioinformatics, MRC University of Glasgow Centre for Virus Research | Filipe,A., Tong,L., Vattipally,S.B., Maclean,A., Gunson,R., Holden,M.T.G., Barr,D., Ho,A., Palmarini,M., Rambaut,A., Robertson,D.L. and Thomson,E.C. |
| EPI_ISL_13436658 | Coordenadoria de Vigilancia em Saude - Sao Paulo | Instituto Adolfo Lutz Strategic Laboratory | Claudio Tavares Sacchi, Karoline Rodrigues Campos, Ariadne Fereira Amarante, Adriano Abbud, Adriana Bugno |
| EPI_ISL_13436792 | Hospital Santa Ignes | Instituto Adolfo Lutz Strategic Laboratory | Claudio Tavares Sacchi, Karoline Rodrigues Campos, Adriano Abbud, Adriana Bugno |
| EPI_ISL_13437056 | Hosp. Alemao Oswaldo Cruz | Instituto Adolfo Lutz Strategic Laboratory | Claudio Tavares Sacchi, Karoline Rodrigues Campos, Ariadne Ferreira Amarante, Adriano Abbud, Adriana Bugno |
| EPI_ISL_13459346 | CRT-DST-AIDS | Instituto Adolfo Lutz Strategic Laboratory | Claudio Tavares Sacchi, Karoline Rodrigues Campos, Ariadne Ferreira Amarante, Adriano Abbud, Adriana Bugno |
| EPI_ISL_13459347, EPI_ISL_13459482, EPI_ISL_13459483 | Instituto de Infectologia Emilio Ribas | Instituto Adolfo Lutz Strategic Laboratory | Claudio Tavares Sacchi, Karoline Rodrigues Campos, Ariadne Ferreira Amarante, Adriano Abbud, Adriana Bugno |
| EPI_ISL_13484458 | Laboratorio de Enterovirus, Instituto Oswaldo Cruz, Fiocruz | Instituto Oswaldo Cruz FIOCRUZ - Laboratory of Respiratory Viruses and Measles (LVRS) | Paola Resende, Elisa Cavalcante Pereira, Bruna Mendonça da Silva, Jéssica Graça Macedo de Carvalho, Larissa Macedo Pinto, Victor Guimaraes, Marilda Siqueira, Renan da Silva Faustino, Marilia Santini, Edson Elias da Silva on behalf of the Fiocruz Genomic Surveillance Network |
| EPI_ISL_13499566 | Erasmus Medical Center Department of Virology | Erasmus Medical Center Department of Virology | Bas Oude Munnink, Marjan Boter, Babette Weller, Richard Molenkamp, Janette Rahamat-Langendoen, Reina Sikkema, Marion Koopmans |
| EPI_ISL_13502582 | Laboratory of Microbiology and Virology, Ospedale Amedeo di Savoia, ASL "Città di Torino" | Laboratory of Microbiology and Virology, Ospedale Amedeo di Savoia, ASL "Città di Torino" | Francesco Cerutti, Antonella Bottoni, Marisa Cazzadore, Tiziano Allice, Maria Grazia Milia, Gabriella Gregori, Elisa Burdino, Valeria Ghisetti |
| EPI_ISL_13508393 | Hosp. Itacolomy Butanta | Instituto Adolfo Lutz Strategic Laboratory | Claudio Tavares Sacchi, Karoline Rodrigues Campos, Ariadne Ferreira Amarante, Adriano Abbud, Adriana Bugno |
| EPI_ISL_13508471 | Instituto de Infectologia Emilio Ribas | Instituto Adolfo Lutz Strategic Laboratory | Claudio Tavares Sacchi, Karoline Rodrigues Campos, Ariadne Ferreira Amarante, Adriano Abbud, Adriana Bugno |
| EPI_ISL_13511312 | Laboratorio de Salud Pública de Antioquia | Instituto Nacional de Salud- Dirección de Investigación en Salud Pública | Katherine Laiton-Donato, Diego A. Álvarez-Díaz, Carlos Franco-Muñoz, Héctor A. Ruiz-Moreno, Paola Rojas-Estevez, Andres Prada, Alicia Rosales, Marcela Mercado-Reyes |
| EPI_ISL_13537923 | Microbiology, Immunology and Transplantation, KU Leuven, Rega Institute | Microbiology, Immunology and Transplantation, KU Leuven, Rega Institute | Wawina-Bokalanga,T., Vanmechelen,B., Logist,A.-S., Sinnesael,R., Ysebaert,L., Bloemen,M. and Maes,P. |
| EPI_ISL_13537924, EPI_ISL_13537925, EPI_ISL_13537926 | Microbiology, Immunology and Transplantation, KU Leuven, Rega Institute | Microbiology, Immunology and Transplantation, KU Leuven, Rega Institute | Vanmechelen,B., Wawina-Bokalanga,T., Logist,A.-S., Sinnesael,R., Ysebaert,L., Verlinden,J., Van Holm,B., Bloemen,M. and Maes,P. |
| EPI_ISL_13544233 | Public Health Agency of Canada, National Microbiology Laboratory | Public Health Agency of Canada, National Microbiology Laboratory | Duggan,A., Hole,D., Knox,N., Yadav,C., Haidl,E., Chapel,M., Domselaar,G.V., Jolly,G., Audet,J., Fernando,L., Antonation,K., Safronetz,D., Hagan,M., Griffiths,E., Leung,A., Graham,M., Peters,G., Go,A., Laminman,V., Kaplen,B., Eshaghi,A., Gubbay,J.B., Hasso,M., Marchand-Austin,A., Olsha,R. and Patel,S.N. |
| EPI_ISL_13544265, EPI_ISL_13544266, EPI_ISL_13544267 | Public Health Agency of Canada, National Microbiology Laboratory | Public Health Agency of Canada, National Microbiology Laboratory | Duggan,A., Hole,D., Knox,N., Yadav,C., Haidl,E., Chapel,M., Domselaar,G.V., Fernando,L., Graham,M., Antonation,K., Audet,J., Hagan,M., Safronetz,D., Leung,A., Peters,G., Go,A., Laminman,V., Kaplen,B., Jolly,G., Charest,H., Levade,I. and Fafard,J. |
| EPI_ISL_13584854, EPI_ISL_13586184 | Institute for Virology, Philipps-University Marburg | Institute for Virology, Philipps-University Marburg | Eickmann, M., Lier, C., Kowalski, K., Kraft, F., Becker, S. |
| EPI_ISL_13607904 | Servicio de Infectologia, Hospital Universitario Dr. José Eleuterio Gonzalez, Universidad Autonoma de Nuevo Leon | Centro de Investigacion e Innovacion en Virologia Medica, Departamento de Bioquimica y Medicina Molecular, Facultad de Medicina, Universidad Autonoma de Nuevo Leon | Kame A. Galan-Huerta, Manuel Paz Infanzon, Ali F. Ruiz Higareda, Laura Nuzzolo-Shihadeh, Adrian Camacho-Ortiz, Paola Bocanegra-Ibarias, Ana M. Rivas-Estilla, Daniel Zacarias-Villarreal, Luis A. Yamallel-Ortega, Maria D. Guerrero-Putz, Jorge Ocampo-Candiani |
| EPI_ISL_13624509 | Instituto de Diagnóstico y Referencia Epidemiológicos/Jurisdicción Sanitaria Cuauhtémoc/Hospital Ángeles Roma | Instituto de Diagnóstico y Referencia Epidemiológicos/Instituto de Biotecnología UNAM | Adnan Araiza-Rodríguez, Adriana Salvador-Patiño, Alejandro Sánchez-Flores, América del Pilar Mandujano-Martínez, Blanca Taboada, Carlos Eduardo Hernández-Sánchez, Carlos F. Arias, Claudia Elena Wong-Arámbula, Daniel José Regalado-Santiago, David Esaú Fragoso-Fonseca, Elizabeth Andrade-Montiel, Fabiola Garcés-Ayala, Fernando González-Dominguez, Gabriel García-Rodríguez, Gloria Vázquez-Castro, Hugo López Gatell Ramírez, Irma López-Martínez, Jerome Verleyen, Jesús Trujillo, Jorge Ochoa, José Ernesto Ramírez-González, Karel Estrada-Guerra, Lucía Hernández-Rivas, Magaly Guadalupe Landa-Flores, Maribel González-Villa, Mireya Mederos-Michel, Nancy Martínez-Velázquez, Noé Escobar-Escamilla, Oliva López, Ricardo Cortés-Alcalá, Ricardo Grande, Verónica Jiménez-Jacinto |
| EPI_ISL_13632068 | SA Pathology | SA Pathology | Coldbeck-Shackley, R, Selway, C, Adamson, PJ, Lim, CK, Turra, M, Bastian, I, Beazley, R, Flood, L, Leong, LEX |
| EPI_ISL_13632071 | Center of Diagnostics and Vaccine Development, Centers for Disease Control, Taiwan | Center of Diagnostics and Vaccine Development, Centers for Disease Control, Taiwan | Jih-Hui Lin, Shu-Chun Chiu, Hsin-I, Huang, Wei-Lun Huang, Wen-Bin, Fann, Pei-Yu, Hsieh, Jyh-Yuan Yang |

| | | | |
|---|---|---|---|
| EPI_ISL_13651348, EPI_ISL_13651350 | Laboratorio de Referencia Nacional de Virus Respiratorio. Centro Nacional de Salud Publica. Instituto Nacional de Salud. | Laboratorio de Referencia Nacional de Virus Respiratorio. Centro Nacional de Salud Publica. Instituto Nacional de Salud. | Carlos Padilla Rojas, Veronica Hurtado Vela, Iris Silva Molina, Luren Sevilla Castañeda, Victor Jimenez Vasquez, Orson Mestanza Millones, Luis Barcena Flores, Wendy Lizarraga Olivares, Alicia Nuñez Llanos, Steve Acedo Lazo, Francisco Ascue Orosco, Kelly Izarra Rojas, Princesa Medrano Alhuay, Karla Vasquez Cajachahua, Estela Huaman Angeles, Jorge Giraldo Chavez, Lilian Huarca Balbin, Lisbet Roxana Inga Angulo, Maria Sandra Villar Saavedra, Henri Bailon Calderon, Lely Solari Zerpa, Gloria Arotinco Garayar. Equipo de vigilancia genomica del Instituto Nacional de Salud. |
| EPI_ISL_13658019, EPI_ISL_13658021 | Erasmus Medical Center Department of Virology | Erasmus Medical Center Department of Virology | Bas Oude Munnink, Marjan Boter, Babette Weller, Richard Molenkamp, Janette Rahamat-Langendoen, Reina Sikkema, Marion Koopmans |
| EPI_ISL_13660191 | Hospital Center Luxembourg | Laboratoire National de Santé Microbiology | Eric Hugoson, Ines Kozar, Sibel Berger, Anke Wienecke-Baldacchino, Bas Oude Munnink, Michel Kohnen, Jean-Hugues Francois, Tamir Abdelrahman |
| EPI_ISL_13705358 | Hosp. Alemao Oswaldo Cruz | Instituto Adolfo Lutz Strategic Laboratory | Claudio Tavares Sacchi, Karoline Rodrigues Campos, Ariadne Ferreira Amarante, Marlon Benedito Nascimento Santos, Alex Domingos Reis, Adriano Abbud, Adriana Bugno |
| EPI_ISL_13705407 | Hosp. Sirio-Libanes | Instituto Adolfo Lutz Strategic Laboratory | Claudio Tavares Sacchi, Karoline Rodrigues Campos, Ariadne Ferreira Amarante, Marlon Benedito Nascimento Santos, Alex Domingos Reis, Adriano Abbud, Adriana Bugno |
| EPI_ISL_13717674 | Hospital Center Luxembourg | Laboratoire National de Santé Microbiology | Eric Hugoson, Ines Kozar, Sibel Berger, Anke Wienecke-Baldacchino, Bas Oude Munnink, Michel Kohnen, Jean-Hugues Francois, Tamir Abdelrahman |
| EPI_ISL_13728303 | Department of Medical Microbiology & Infection prevention, Amsterdam University Medical Centers location AMC | Department of Medical Microbiology & Infection prevention, Amsterdam University Medical Centers location AMC | Matthijs Welkers, Jelle Koopsen, Robin van Houdt, Marcel Jonges, Sebastien Matamoros, Sjoerd Rebers, Fokla Zorgdrager, Sylvia Bruisten, Judith den Uil, Akke Cornelissen, Janke Schinkel, Menno de Jong, Gini van Rijckevorsel and Mariken van der Lubben on behalf of the Amsterdam Regional Genomic epidemiology and Outbreak Surveillance (ARGOS) consortium |
| EPI_ISL_13732932 | Hosp. Sao Joaquim - Beneficiencia Portuguesa | Instituto Adolfo Lutz Strategic Laboratory | Claudio Tavares Sacchi, Karoline Rodrigues Campos, Ariadne Ferreira Amarante, Marlon Benedito Nascimento Santos, Alex Domingos Reis, Adriano Abbud, Adriana Bugno |
| EPI_ISL_13734269 | Department of Clinical Sciences, Institute of Tropica Medicine | Department of Clinical Sciences, Institute of Tropica Medicine | De Baetselier,I., Van Dijck,C., Kenyon,C., Coppens,J., Smet,H., de Block,T., Coppens,S., Vanroye,F., Bugert,J., Girl,P., Liesenborghs,L., Selhorst,P., Arien,K., Van den Bossche,D., Florence,E., Rezende,A.M., Vercauteren,K. and Van Esbroeck,M. |
| EPI_ISL_13780332 | Laboratorio Central de Saude Publica do Estado de Minas Gerais (LACEN-MG) | Laboratorio Central de Saude Publica do Estado de Minas Gerais (LACEN-MG) | Adriana Aparecida Ribeiro, Felipe Campos de Melo Iani, Ludmila Oliveira Lamounier, Natália Rocha Guimarães,vTalita Emile Ribeiro Adelino, Vagner Fonseca |
| EPI_ISL_13817808 | New York University Langone Health | New York University Langone Health | Adriana Heguy, Dacia Dimartino, Emily Guzman, Christian Marier, Peter Meyn, Sitharam Ramaswami, Gael Westby, Paul Zappile, Yutong Zhang, Guiqing Wang |
| EPI_ISL_13822667, EPI_ISL_13822668, EPI_ISL_13822669, EPI_ISL_13822718 | Erasmus Medical Center Department of Virology | Erasmus Medical Center Department of Virology | Bas Oude Munnink, Marjan Boter, Babette Weller, Richard Molenkamp, Janette Rahamat-Langendoen, Reina Sikkema, Marion Koopmans |
| EPI_ISL_13833194, EPI_ISL_13833195, EPI_ISL_13833196 | Laboratorio de Referencia Nacional de Virus Respiratorio. Centro Nacional de Salud Publica. Instituto Nacional de Salud. | Laboratorio de Referencia Nacional de Virus Respiratorio. Centro Nacional de Salud Publica. Instituto Nacional de Salud. | Carlos Padilla Rojas, Veronica Hurtado Vela, Iris Silva Molina, Luren Sevilla Castañeda, Victor Jimenez Vasquez, Orson Mestanza Millones, Luis Barcena Flores, Wendy Lizarraga Olivares, Alicia Nuñez Llanos, Steve Acedo Lazo, Francisco Ascue Orosco, Kelly Izarra Rojas, Princesa Medrano Alhuay, Karla Vasquez Cajachahua, Estela Huaman Angeles, Jorge Giraldo Chavez, Lilian Huarca Balbin, Lisbet Roxana Inga Angulo, Maria Sandra Villar Saavedra, Henri Bailon Calderon, Lely Solari Zerpa, Gloria Arotinco Garayar. Equipo de vigilancia genomica del Instituto Nacional de Salud. |
| EPI_ISL_13889435, EPI_ISL_13889436, EPI_ISL_13889437, EPI_ISL_13889438, EPI_ISL_13889439, EPI_ISL_13889440, EPI_ISL_13889441, EPI_ISL_13889442, EPI_ISL_13889443, EPI_ISL_13889444, EPI_ISL_13889445, EPI_ISL_13889446, EPI_ISL_13889447, EPI_ISL_13889448, EPI_ISL_13889449, EPI_ISL_13889450, EPI_ISL_13889515, EPI_ISL_13889590, EPI_ISL_13889660, EPI_ISL_13889729, EPI_ISL_13889796, EPI_ISL_13889908, EPI_ISL_13889977, EPI_ISL_13890048, EPI_ISL_13890135, EPI_ISL_13890204, EPI_ISL_13890273, EPI_ISL_13890338, EPI_ISL_13890408, EPI_ISL_13890464, EPI_ISL_13890465, EPI_ISL_13890466, EPI_ISL_13890467, EPI_ISL_13890468, EPI_ISL_13890469, EPI_ISL_13890470, EPI_ISL_13890471, EPI_ISL_13890472, EPI_ISL_13890473, EPI_ISL_13890474, EPI_ISL_13890475, EPI_ISL_13890476, EPI_ISL_13890477, EPI_ISL_13890478, EPI_ISL_13890479, EPI_ISL_13890480, EPI_ISL_13890481, EPI_ISL_13890482 | | | |
| see above | Charité Universitätsmedizin Berlin, Institut für Virologie/Labor Berlin | Charité Universitätsmedizin Berlin, Institut für Virologie | Terry C. Jones, Julia Schneider, Barbara Mühlemann, Talitha Veith, Jörn Beheim-Schwarzbach, Julia Tesch, Marie Luisa Schmidt, Felix Walper, Tobias Bleicker, Caroline Isner, Frieder Pfäfflin, Ricardo Niklas Werner, Victor M. Corman, Christian Drosten |
| EPI_ISL_13891126 | Ministry of Health Turkey | Ministry of Health Turkey | Fatma Bayrakdar, Suleyman Yalcin, Gulay Korukluoglu |
| EPI_ISL_13953610, EPI_ISL_13953611 | Indian Council of Medical Research-National Institute of Virology | Indian Council of Medical Research-National Institute of Virology | Pragya Yadav, Rima Sahay, Anita Aich Shete, Sreelekshmy Mohandas, Priya Abraham |
| EPI_ISL_13983354, EPI_ISL_13983355 | Instituto de Infectologia Emilio Ribas | Instituto Adolfo Lutz Strategic Laboratory | Claudio Tavares Sacchi, Karoline Rodrigues Campos, Ariadne Ferreira Amarante, Marlon Benedito Nascimento Santos, Alex Domingos Reis, Adriano Abbud, Adriana Bugno |
| EPI_ISL_13983356 | INSPI-Centro de Referencia Nacional de Virus Exantemáticos, Gastroentéricos y Transmitido por Vectores. | INSPI-Dirección Técnica de Investigación, Desarrollo e Innovación INSPI-Centro de Referencia Nacional de Genómica, Secuenciación y Bioinformática | Andrés Carrazco-Montalvo, Diana Gutiérrez, Naomi Mora, Silvia Salgado-Cisneros, Johana Parrales-Valdiviezo, Martha Sánchez-Domenech, Diego Morales, Gulnara Borja-Cabrera, Leandro Patiño*. |
| EPI_ISL_13983888 | Bangkok Hospital Phuket | National Institute of Health, Department of Medical Sciences, Ministry of Public Health, Thailand | Pilailuk Okada; Siripaporn Phuygun; Nuttida Thongpramul; Thanutsapa Thanadachakul; Kazuhisa Okada; Archawin Rojanawiwat; Chakkarat Pitayawonganon; Supakit Sirilak |
| EPI_ISL_13993734, EPI_ISL_13993735, EPI_ISL_13993737, EPI_ISL_13993738 | California Department of Public Health | California Department of Public Health | Viral and Rickettsial Disease Laboratory |
| EPI_ISL_14003930 | University of Rochester Medical Center | University of Rochester Medical Center | Andrew Cameron, Mondraya Howard, Sara Connelly, Dwight Hardy, Kelly DeLary |
| EPI_ISL_14021725 | Hosp. Municipal Enf. Antonio Policarpo de Oliveira | Instituto Adolfo Lutz Strategic Laboratory | Claudio Tavares Sacchi, Karoline Rodrigues Campos, Ariadne Ferreira Amarante, Marlon Benedito Nascimento Santos, Alex Domingos Reis, Adriano Abbud, Adriana Bugno |
| EPI_ISL_14070493, EPI_ISL_14070852, EPI_ISL_14070854, EPI_ISL_14070855 | Instituto de Infectologia Emilio Ribas | Instituto Adolfo Lutz Strategic Laboratory | Claudio Tavares Sacchi, Karoline Rodrigues Campos, Ariadne Ferreira Amarante, Marlon Benedito Nascimento Santos, Alex Domingos Reis, Adriano Abbud, Adriana Bugno |
| EPI_ISL_14089382 | Pathogen Genomics Lab, National Institute for Biomedical Research (INRB) | Pathogen Genomics Lab, National Institute for Biomedical Research (INRB) | Placide Mbala-Kingebeni, Eddy Kinganda-Lusamaki, Adrienne Amuri-Aziza, Elisabeth Pukuta, Catherine Pratt, Nicolas Fernandez, Emmanuel Lokilo Lofiko, Gradi Luakanda Ndelemo, Francisca Muyembe Mawete, Jean Claude Makangara Cigolo, Elisabeth Muyamuna, Raphaël Lumembe Numbi, Gabriel Kabamba Lungenyi, Prince Akil Bandali, Pauline Musuamba Kayembe, Rilia Ola Mpumbe, Emile Malembi, Emmanuel Hasivirwe Vakaniaki, Andrew Rambaut, Nick Loman, Kristian Andersen, Michael Wiley, Ahidjo Ayouba, Steve Ahuka-Mundeke, Martine Peeters, Eric Delaporte, Jean-Jacques Muyembe Tamfum |
| EPI_ISL_14153982 | Vajira Hospital | National Institute of Health, Department of Medical Sciences, Ministry of Public Health, Thailand | Pilailuk Okada; Siripaporn Phuygun; Nuttida Thongpramul; Thanutsapa Thanadachakul; Kazuhisa Okada; Archawin Rojanawiwat; Chakkarat Pitayawonganon; Supakit Sirilak |
| EPI_ISL_14167248, EPI_ISL_14167575 | Medical University of Vienna Center for Virology | Medical University of Vienna Center for Virology | Jeremy V. Camp, Monika Redlberger-Fritz, Stephan W. Aberle |
| EPI_ISL_14170200, EPI_ISL_14170201, EPI_ISL_14170202, EPI_ISL_14170203, EPI_ISL_14170204 | Erasmus Medical Center Department of Virology | Erasmus Medical Center Department of Virology | Bas Oude Munnink, Marjan Boter, Babette Weller, Babs Verstrepen, Richard Molenkamp, Janette Rahamat-Langendoen, Reina Sikkema, Marion Koopmans |
| EPI_ISL_14181948, EPI_ISL_14181949, EPI_ISL_14181950, EPI_ISL_14181951, EPI_ISL_14181952, EPI_ISL_14181953, EPI_ISL_14181954, EPI_ISL_14181955, EPI_ISL_14181956, EPI_ISL_14181957, EPI_ISL_14181958, EPI_ISL_14181959, EPI_ISL_14181960 | | | |
| see above | Institute of Health Carlos III, Bioinformatics Unit | Institute of Health Carlos III, Bioinformatics Unit | Cuesta,I. |
| EPI_ISL_14189012 | Los Angeles County Public Health Laboratories | Los Angeles County Public Health Laboratories | P. Hemarajata et al. |
| EPI_ISL_14201640, EPI_ISL_14201641, EPI_ISL_14201642, EPI_ISL_14201643, EPI_ISL_14201644, EPI_ISL_14201645 | Pathogen Genomics Lab, National Institute for Biomedical Research (INRB) | Pathogen Genomics Lab, National Institute for Biomedical Research (INRB) | Placide Mbala-Kingebeni, Eddy Kinganda-Lusamaki, Adrienne Amuri-Aziza, Elisabeth Pukuta, Catherine Pratt, Nicolas Fernandez, Emmanuel Lokilo Lofiko, Gradi Luakanda Ndelemo, Francisca Muyembe Mawete, Jean Claude Makangara Cigolo, Elisabeth Muyamuna, Raphaël Lumembe Numbi, Gabriel Kabamba Lungenyi, Prince Akil Bandali, Pauline Musuamba Kayembe, Rilia Ola Mpumbe, Emile Malembi, Emmanuel Hasivirwe Vakaniaki, Andrew Rambaut, Nick Loman, Kristian Andersen, Michael Wiley, Ahidjo Ayouba, Steve Ahuka-Mundeke, Martine Peeters, Eric Delaporte, Jean-Jacques Muyembe Tamfum |
| EPI_ISL_14207724, EPI_ISL_14207726, EPI_ISL_14207729, EPI_ISL_14207732, EPI_ISL_14207734, EPI_ISL_14207735, EPI_ISL_14207736, EPI_ISL_14207737, EPI_ISL_14207738, EPI_ISL_14207739, EPI_ISL_14207740, EPI_ISL_14207741 | | | |
| see above | Laboratorio de Referencia Nacional de Virus Respiratorio. Centro Nacional de Salud Publica. Instituto Nacional de Salud. | Laboratorio de Referencia Nacional de Virus Respiratorio. Centro Nacional de Salud Publica. Instituto Nacional de Salud. | Carlos Padilla Rojas, Veronica Hurtado Vela, Iris Silva Molina, Luren Sevilla Castañeda, Victor Jimenez Vasquez, Orson Mestanza Millones, Luis Barcena Flores, Wendy Lizarraga Olivares, Alicia Nuñez Llanos, Steve Acedo Lazo, Francisco Ascue Orosco, Kelly Izarra Rojas, Princesa Medrano Alhuay, Karla Vasquez Cajachahua, Estela Huaman Angeles, Jorge Giraldo Chavez, Lilian Huarca Balbin, Lisbet Roxana Inga Angulo, Maria Sandra Villar Saavedra, Henri Bailon Calderon, Lely Solari Zerpa, Gloria Arotinco Garayar. Equipo de vigilancia genomica del Instituto Nacional de Salud. |
| EPI_ISL_14224334 | Genetica Molecular and Subdepartamento de Virologia ISP Chile | Instituto de Salud Publica de Chile | Paulo C. Covarrubias, Andrés E. Castillo, Constanza Campano, Mariela Guajardo, Bárbara Parra, Rodrigo Fasce Pineda, Jorge Fernández |
| EPI_ISL_14251112 | University of Rochester Medical Center | University of Rochester Medical Center | Andrew Cameron, Mondraya Howard, Joel Maki, Sara Connelly, Kelly Delary, Dwight Hardy |
| EPI_ISL_14254435, EPI_ISL_14254436, EPI_ISL_14254437, EPI_ISL_14254438 | Erasmus Medical Center Department of Virology | Erasmus Medical Center Department of Virology | Bas Oude Munnink, Marjan Boter, Babette Weller, Babs Verstrepen, Richard Molenkamp, Janette Rahamat-Langendoen, Reina Sikkema, Marion Koopmans |
| EPI_ISL_14326638, EPI_ISL_14326639, EPI_ISL_14326640, EPI_ISL_14326641, EPI_ISL_14326642, EPI_ISL_14326643 | Environmental, Agricultural, and Occupational Health, University of Nebraska Medical Center, 984388 Nebraska Medical Center | Environmental, Agricultural, and Occupational Health, University of Nebraska Medical Center, 984388 Nebraska Medical Center | Tegomoh,B., Cross,S.T., Chapman,R.C., Bernhard,K., McCutchen,E.L., Fauver,J.R., Pratt,C.B., Warden,D.E., Iwen,P.C., Donahue,M. and Wiley,M.R. |
| EPI_ISL_14326644 | Environmental, Agricultural, and Occupational Health, University of Nebraska Medical Center, 984388 Nebraska Medical Center | Environmental, Agricultural, and Occupational Health, University of Nebraska Medical Center, 984388 Nebraska Medical Center | Tegomoh,B., Cross,S.T., Chapman,R.C., Bernhard,K., McCutchen,E.L., Fauver,J.R., Pratt,C.B., Warden,D.E., Iwen,P.C., Donahue,M. and Wiley,M.R |
| EPI_ISL_14414948 | UMS Parque Industrial Curitiba | Instituto Adolfo Lutz Strategic Laboratory | Claudio Tavares Sacchi, Karoline Rodrigues Campos, Ariadne Ferreira Amarante, Marlon Benedito Nascimento Santos, Alex Domingos Reis, Adriano Abbud, Adriana Bugno |
| EPI_ISL_14415810 | CTA Sao Miguel | Instituto Adolfo Lutz Strategic Laboratory | Claudio Tavares Sacchi, Karoline Rodrigues Campos, Ariadne Ferreira Amarante, Marlon Benedito Nascimento Santos, Alex Domingos Reis, Adriano Abbud, Adriana Bugno |
| EPI_ISL_14438678, EPI_ISL_14438679, EPI_ISL_14438680, EPI_ISL_14438681, EPI_ISL_14438682, EPI_ISL_14438683, EPI_ISL_14438684, EPI_ISL_14438685, EPI_ISL_14438686, EPI_ISL_14438687, EPI_ISL_14438688, EPI_ISL_14438689, EPI_ISL_14438690, EPI_ISL_14438691, EPI_ISL_14438692, EPI_ISL_14438693, EPI_ISL_14438694, EPI_ISL_14438696, EPI_ISL_14438697, EPI_ISL_14438698, EPI_ISL_14441870, EPI_ISL_14441871, EPI_ISL_14441872, EPI_ISL_14441873, EPI_ISL_14441874, EPI_ISL_14441875, EPI_ISL_14441876, EPI_ISL_14441877, EPI_ISL_14441878, EPI_ISL_14441879, EPI_ISL_14441880, EPI_ISL_14441881, EPI_ISL_14441882, EPI_ISL_14441883, EPI_ISL_14441884, EPI_ISL_14441885, EPI_ISL_14441886, EPI_ISL_14441887 | | | |
| see above | Rhode Island State Health Laboratory | Rhode Island State Health Laboratory | Kristin Carpenter-Azevedo, Sean Sierra-Patev, Richard C. Huard |
| EPI_ISL_14445098, EPI_ISL_14445099, EPI_ISL_14445100, EPI_ISL_14445101, EPI_ISL_14445102, EPI_ISL_14445103, EPI_ISL_14445104, EPI_ISL_14445105, EPI_ISL_14445106, EPI_ISL_14445107, EPI_ISL_14445108, EPI_ISL_14445109, EPI_ISL_14445110, EPI_ISL_14445111, EPI_ISL_14445112, EPI_ISL_14445113, EPI_ISL_14445114, EPI_ISL_14445115, EPI_ISL_14445116, EPI_ISL_14445117, EPI_ISL_14445118, EPI_ISL_14445119, EPI_ISL_14445120, EPI_ISL_14445121, EPI_ISL_14445122, EPI_ISL_14445123, EPI_ISL_14445124, EPI_ISL_14445125, EPI_ISL_14445126, EPI_ISL_14445127, EPI_ISL_14445128, EPI_ISL_14445130, EPI_ISL_14445131, EPI_ISL_14445132, EPI_ISL_14445133, EPI_ISL_14445134, EPI_ISL_14445135, EPI_ISL_14445136, EPI_ISL_14445137, EPI_ISL_14445138, EPI_ISL_14445139, EPI_ISL_14445140, EPI_ISL_14445142, EPI_ISL_14445143, EPI_ISL_14445144, EPI_ISL_14445145, EPI_ISL_14445146, EPI_ISL_14445147, EPI_ISL_14445148, EPI_ISL_14445149, EPI_ISL_14445150, EPI_ISL_14445151, EPI_ISL_14445152, EPI_ISL_14445153 | | | |
| see above | Laboratorio de Referencia Nacional de Virus Respiratorio. Centro Nacional de Salud Publica. Instituto Nacional de Salud. | Laboratorio de Referencia Nacional de Virus Respiratorio. Centro Nacional de Salud Publica. Instituto Nacional de Salud. | Carlos Padilla Rojas, Veronica Hurtado Vela, Iris Silva Molina, Luren Sevilla Castañeda, Victor Jimenez Vasquez, Orson Mestanza Millones, Luis Barcena Flores, Wendy Lizarraga Olivares, Alicia Nuñez Llanos, Steve Acedo Lazo, Francisco Ascue Orosco, Kelly Izarra Rojas, Princesa Medrano Alhuay, Karla Vasquez Cajachahua, Estela Huaman Angeles, Jorge Giraldo Chavez, Lilian Huarca Balbin, Lisbet Roxana Inga Angulo, Maria Sandra Villar Saavedra, Henri Bailon Calderon, Lely Solari Zerpa, Gloria Arotinco Garayar. Equipo de vigilancia genomica del Instituto Nacional de Salud. |
| EPI_ISL_14465517 | Centro de Desenvolvimento Científico e Tecnológico (CDCT), Centro Estadual de Vigilância em Saúde (CEVS) da Secretaria Estadual da Saúde (SES-RS) | Centro de Desenvolvimento Científico e Tecnológico (CDCT), Centro Estadual de Vigilância em Saúde (CEVS) da Secretaria Estadual da Saúde (SES-RS) | Richard Steiner Salvato, Regina Bones Barcellos, Fernanda Marques Godinho |
| EPI_ISL_14467428, EPI_ISL_14467429 | Laboratório Central de Saúde Pública do Amazonas - LACEN-AM | Laboratório de Ecologia de Doenças Transmissíveis na Amazônia, Instituto Leônidas e Maria Deane - Fiocruz Amazônia | Victor Souza, Fernanda Nascimento, Matilde Mejía, Dejanane Silva, Luciana Gonçalves, Tatyana Costa Amorim Ramos, Ana Ruth Lima Arcanjo, Valdinete Nascimento, Felipe Naveca on behalf of the Fiocruz COVID-19 Genomic Surveillance Network |

| | | | |
|---|---|---|---|
| EPI_ISL_14486937, EPI_ISL_14487241 | Instituto Nacional de Salud | Instituto Nacional de Salud- Dirección de Investigación en Salud Pública | Katherine Laiton-Donato, Diego A. Álvarez-Díaz, Carlos Franco-Muñoz, Héctor A. Ruiz-Moreno, Paola Rojas-Estevez, Alicia Rosales, Daniel Martinez, Sergio Gómez, Astrid Carolina Flores, Franklin Prieto, Diana Walteros, Marcela Mercado-Reyes |
| EPI_ISL_14515177, EPI_ISL_14515211, EPI_ISL_14515218, EPI_ISL_14515219 | Department of Infectious Diseases, National Institute of Health Doutor Ricardo Jorge, Portugal (INSA) | Department of Infectious Diseases, National Institute of Health Doutor Ricardo Jorge, Portugal (INSA) | Isidro,J., Borges,V., Pinto,M., Sobral,D., Santos,J., Nunes,A., Mixao,V., Ferreira,R., Santos,D., Duarte,S., Vieira,L., Borrego,M.J., Nuncio,S., Lopes de Carvalho,I., Pelerito,A., Cordeiro,R. and Gomes,J.P. |
| EPI_ISL_14526939, EPI_ISL_14526940, EPI_ISL_14526941, EPI_ISL_14526942, EPI_ISL_14526943, EPI_ISL_14526944, EPI_ISL_14526945, EPI_ISL_14526946, EPI_ISL_14526947, EPI_ISL_14526948, EPI_ISL_14526949, EPI_ISL_14526950, EPI_ISL_14526951, EPI_ISL_14526952, EPI_ISL_14526953, EPI_ISL_14526954, EPI_ISL_14526955, EPI_ISL_14526956 | | | |
| see above | Connecticut Department of Public Health | Grubaugh Lab - Yale School of Public Health | Nicholas F. G. Chen, Chrispin Chaguza, Kien Pham, Nathan D. Grubaugh, Christina Nishimura, Claire Pearson, Kutluhan Incekara, Jian Ping Huang, Emily Gagnon, Ethan Reever, Jafar Razeq, Anthony Muyombwe, Chantal B. F. Vogels |
| EPI_ISL_14541645, EPI_ISL_14541654 | Public Health Authority of the Slovak Republic | Laboratory of Genomics and Bioinformatics, Comenius University Science Park | Tomáš Szemes, Edita Staroňová, Elena Tichá, Lucia Ševčíková, Terézia Vrabľová, Tatiana Sedláčková, Miroslav Böhmer, Jaroslav Budiš, Pavol Mišenko |
| EPI_ISL_14553812 | Hospital CIMA San Jose | Inciensa, Instituto de Investigación y Enseñanza en Nutrición y Salud | Francisco Duarte, Ana Isela Ruiz-Gonzalez, Hillary Serrano, Diana Cantillo, Claudio Soto-Garita, Gustavo Vega, Estela Cordero, Adriana Godinez & Melany Calderon |
| EPI_ISL_14561914 | Los Angeles County Public Health Laboratories | Los Angeles County Public Health Laboratories | P. Hemarajata et al. |
| EPI_ISL_14571429 | Hosp. Municipal Dr. Jose de Carvalho Florence | Instituto Adolfo Lutz Strategic Laboratory | Claudio Tavares Sacchi, Karoline Rodrigues Campos, Ariadne Ferreira Amarante, Marlon Benedito Nascimento Santos, Alex Domingos Reis, Adriano Abbud, Adriana Bugno |
| EPI_ISL_14571435 | Secrtetaria Municipal de Saude de Sertaozinho | Instituto Adolfo Lutz Strategic Laboratory | Claudio Tavares Sacchi, Karoline Rodrigues Campos, Ariadne Ferreira Amarante, Marlon Benedito Nascimento Santos, Alex Domingos Reis, Adriano Abbud, Adriana Bugno |
| EPI_ISL_14571439 | Secrtetaria Municipal de Saude de Sata Barbara D Oeste | Instituto Adolfo Lutz Strategic Laboratory | Claudio Tavares Sacchi, Karoline Rodrigues Campos, Ariadne Ferreira Amarante, Marlon Benedito Nascimento Santos, Alex Domingos Reis, Adriano Abbud, Adriana Bugno |
| EPI_ISL_14571441 | Hosp. Municipal Dr. Waldemar Tebaldi | Instituto Adolfo Lutz Strategic Laboratory | Claudio Tavares Sacchi, Karoline Rodrigues Campos, Ariadne Ferreira Amarante, Marlon Benedito Nascimento Santos, Alex Domingos Reis, Adriano Abbud, Adriana Bugno |
| EPI_ISL_14571442 | Instituto de Infectologia Emilio Ribas II Baixada Santista | Instituto Adolfo Lutz Strategic Laboratory | Claudio Tavares Sacchi, Karoline Rodrigues Campos, Ariadne Ferreira Amarante, Marlon Benedito Nascimento Santos, Alex Domingos Reis, Adriano Abbud, Adriana Bugno |
| EPI_ISL_14571444 | UBDS DR. Italo Baruffi Castelo Branco | Instituto Adolfo Lutz Strategic Laboratory | Claudio Tavares Sacchi, Karoline Rodrigues Campos, Ariadne Ferreira Amarante, Marlon Benedito Nascimento Santos, Alex Domingos Reis, Adriano Abbud, Adriana Bugno |
| EPI_ISL_14583298, EPI_ISL_14583299, EPI_ISL_14583300, EPI_ISL_14583301, EPI_ISL_14583302, EPI_ISL_14583303, EPI_ISL_14583304, EPI_ISL_14583305, EPI_ISL_14583306, EPI_ISL_14583307, EPI_ISL_14583308, EPI_ISL_14583309 | | | |
| see above | Rhode Island State Health Laboratory | Rhode Island State Health Laboratory | Kristin Carpenter-Azevedo, Sean Sierra-Patev, Richard C. Huard |
| EPI_ISL_14584274, EPI_ISL_14584275, EPI_ISL_14584276, EPI_ISL_14584277, EPI_ISL_14584278, EPI_ISL_14584279, EPI_ISL_14584280, EPI_ISL_14584281, EPI_ISL_14584282, EPI_ISL_14584283, EPI_ISL_14584284, EPI_ISL_14584285, EPI_ISL_14584286, EPI_ISL_14584287, EPI_ISL_14584288, EPI_ISL_14584289, EPI_ISL_14584290, EPI_ISL_14584291, EPI_ISL_14584292, EPI_ISL_14584293, EPI_ISL_14584294, EPI_ISL_14584295, EPI_ISL_14584296, EPI_ISL_14584297, EPI_ISL_14584298, EPI_ISL_14584299, EPI_ISL_14584300, EPI_ISL_14584301, EPI_ISL_14584302, EPI_ISL_14584303, EPI_ISL_14584304, EPI_ISL_14584305, EPI_ISL_14584306, EPI_ISL_14584307, EPI_ISL_14584308, EPI_ISL_14584309, EPI_ISL_14584310, EPI_ISL_14584311 | | | |
| see above | Laboratorio de Referencia Nacional de Virus Respiratorio. Centro Nacional de Salud Publica. Instituto Nacional de Salud. | Laboratorio de Referencia Nacional de Virus Respiratorio. Centro Nacional de Salud Publica. Instituto Nacional de Salud. | Carlos Padilla Rojas, Veronica Hurtado Vela, Iris Silva Molina, Luren Sevilla Castañeda, Victor Jimenez Vasquez, Orson Mestanza Millones, Luis Barcena Flores, Wendy Lizarraga Olivares, Alicia Nuñez Llanos, Steve Acedo Lazo, Francisco Ascue Orosco, Kelly Izarra Rojas, Princesa Medrano Alhuay, Karla Vasquez Cajachahua, Estela Huaman Angeles, Jorge Giraldo Chavez, Lilian Huarca Balbin, Lisbet Roxana Inga Angulo, Maria Sandra Villar Saavedra, Henri Bailon Calderon, Lely Solari Zerpa, Gloria Arotinco Garayar. Equipo de vigilancia genomica del Instituto Nacional de Salud. |
| EPI_ISL_14615579 | RSUPN dr. Cipto Mangunkusumo | National Institute of Health Research and Development | Hana Apsari Pawestri, Arie Ardiansyah Nugraha, Fajar Nur Sulistiyohadi, Subangkit, Krisna NA Pangesti, Tze Minn Mak, I Gede Made Wirabrata |
| EPI_ISL_14622055 | Instituto de Infectologia Emilio Ribas | Instituto Adolfo Lutz Strategic Laboratory | Claudio Tavares Sacchi, Karoline Rodrigues Campos, Ariadne Ferreira Amarante, Marlon Benedito Nascimento Santos, Alex Domingos Reis, Adriano Abbud, Adriana Bugno |
| EPI_ISL_14622520 | UBS Jovaia | Instituto Adolfo Lutz Strategic Laboratory | Claudio Tavares Sacchi, Karoline Rodrigues Campos, Ariadne Ferreira Amarante, Marlon Benedito Nascimento Santos, Alex Domingos Reis, Adriano Abbud, Adriana Bugno |
| EPI_ISL_14622706 | Centro de Referencia Modulo I SAE II Bauru | Instituto Adolfo Lutz Strategic Laboratory | Claudio Tavares Sacchi, Karoline Rodrigues Campos, Ariadne Ferreira Amarante, Marlon Benedito Nascimento Santos, Alex Domingos Reis, Adriano Abbud, Adriana Bugno |
| EPI_ISL_14622707 | USF Boicucanga I Sao Sebastiao | Instituto Adolfo Lutz Strategic Laboratory | Claudio Tavares Sacchi, Karoline Rodrigues Campos, Ariadne Ferreira Amarante, Marlon Benedito Nascimento Santos, Alex Domingos Reis, Adriano Abbud, Adriana Bugno |
| EPI_ISL_14622913 | Secretaria Municipal de Saude de Caxias do Sul | Instituto Adolfo Lutz Strategic Laboratory | Claudio Tavares Sacchi, Karoline Rodrigues Campos, Ariadne Ferreira Amarante, Marlon Benedito Nascimento Santos, Alex Domingos Reis, Adriano Abbud, Adriana Bugno |
| EPI_ISL_14622953 | Sistema de Vigilancia em Saude Viamao | Instituto Adolfo Lutz Strategic Laboratory | Claudio Tavares Sacchi, Karoline Rodrigues Campos, Ariadne Ferreira Amarante, Marlon Benedito Nascimento Santos, Alex Domingos Reis, Adriano Abbud, Adriana Bugno |
| EPI_ISL_14622960 | Vigilancia Epidemiologica Municipal | Instituto Adolfo Lutz Strategic Laboratory | Claudio Tavares Sacchi, Karoline Rodrigues Campos, Ariadne Ferreira Amarante, Marlon Benedito Nascimento Santos, Alex Domingos Reis, Adriano Abbud, Adriana Bugno |
| EPI_ISL_14623175 | Centro de Referencia em Especialidades Central Rib Preto | Instituto Adolfo Lutz Strategic Laboratory | Claudio Tavares Sacchi, Karoline Rodrigues Campos, Ariadne Ferreira Amarante, Marlon Benedito Nascimento Santos, Alex Domingos Reis, Adriano Abbud, Adriana Bugno |
| EPI_ISL_14623523 | Laboratorio Municipal de Piracicaba | Instituto Adolfo Lutz Strategic Laboratory | Claudio Tavares Sacchi, Karoline Rodrigues Campos, Ariadne Ferreira Amarante, Marlon Benedito Nascimento Santos, Alex Domingos Reis, Adriano Abbud, Adriana Bugno |
| EPI_ISL_14623704 | Unidade Basica de Saude Esplanada | Instituto Adolfo Lutz Strategic Laboratory | Claudio Tavares Sacchi, Karoline Rodrigues Campos, Ariadne Ferreira Amarante, Marlon Benedito Nascimento Santos, Alex Domingos Reis, Adriano Abbud, Adriana Bugno |
| EPI_ISL_14624411 | Hospital Albert Sabin Atibaia | Instituto Adolfo Lutz Strategic Laboratory | Claudio Tavares Sacchi, Karoline Rodrigues Campos, Ariadne Ferreira Amarante, Marlon Benedito Nascimento Santos, Alex Domingos Reis, Adriano Abbud, Adriana Bugno |
| EPI_ISL_14624610 | USAFA Forte | Instituto Adolfo Lutz Strategic Laboratory | Claudio Tavares Sacchi, Karoline Rodrigues Campos, Ariadne Ferreira Amarante, Marlon Benedito Nascimento Santos, Alex Domingos Reis, Adriano Abbud, Adriana Bugno |
| EPI_ISL_14624698 | Centro de Referencia em AIDS SECRAIDS | Instituto Adolfo Lutz Strategic Laboratory | Claudio Tavares Sacchi, Karoline Rodrigues Campos, Ariadne Ferreira Amarante, Marlon Benedito Nascimento Santos, Alex Domingos Reis, Adriano Abbud, Adriana Bugno |
| EPI_ISL_14624832 | Servico de Vigilancia Epidemiologica e de Zoonoses do Guaruja | Instituto Adolfo Lutz Strategic Laboratory | Claudio Tavares Sacchi, Karoline Rodrigues Campos, Ariadne Ferreira Amarante, Marlon Benedito Nascimento Santos, Alex Domingos Reis, Adriano Abbud, Adriana Bugno |
| EPI_ISL_14624915 | SMS Aruja | Instituto Adolfo Lutz Strategic Laboratory | Claudio Tavares Sacchi, Karoline Rodrigues Campos, Ariadne Ferreira Amarante, Marlon Benedito Nascimento Santos, Alex Domingos Reis, Adriano Abbud, Adriana Bugno |
| EPI_ISL_14625156 | Secretaria Municipal de Saude de Suzano | Instituto Adolfo Lutz Strategic Laboratory | Claudio Tavares Sacchi, Karoline Rodrigues Campos, Ariadne Ferreira Amarante, Marlon Benedito Nascimento Santos, Alex Domingos Reis, Adriano Abbud, Adriana Bugno |
| EPI_ISL_14625157 | PSF Vila Nossa Senhora de Fatima Fartura | Instituto Adolfo Lutz Strategic Laboratory | Claudio Tavares Sacchi, Karoline Rodrigues Campos, Ariadne Ferreira Amarante, Marlon Benedito Nascimento Santos, Alex Domingos Reis, Adriano Abbud, Adriana Bugno |
| EPI_ISL_14625230 | UBS Centro Clair Aparecida Pavan | Instituto Adolfo Lutz Strategic Laboratory | Claudio Tavares Sacchi, Karoline Rodrigues Campos, Ariadne Ferreira Amarante, Marlon Benedito Nascimento Santos, Alex Domingos Reis, Adriano Abbud, Adriana Bugno |
| EPI_ISL_14625256 | UMS Campina do Siqueira | Instituto Adolfo Lutz Strategic Laboratory | Claudio Tavares Sacchi, Karoline Rodrigues Campos, Ariadne Ferreira Amarante, Marlon Benedito Nascimento Santos, Alex Domingos Reis, Adriano Abbud, Adriana Bugno |
| EPI_ISL_14625282 | Hospital Edmundo Vasconcelos | Instituto Adolfo Lutz Strategic Laboratory | Claudio Tavares Sacchi, Karoline Rodrigues Campos, Ariadne Ferreira Amarante, Marlon Benedito Nascimento Santos, Alex Domingos Reis, Adriano Abbud, Adriana Bugno |
| EPI_ISL_14664595, EPI_ISL_14665379, EPI_ISL_14665380, EPI_ISL_14665381, EPI_ISL_14665382, EPI_ISL_14665383, EPI_ISL_14665384, EPI_ISL_14665385, EPI_ISL_14665386, EPI_ISL_14665387, EPI_ISL_14665388, EPI_ISL_14665389, EPI_ISL_14665390, EPI_ISL_14665391, EPI_ISL_14665392, EPI_ISL_14665393 | | | |
| see above | CT Department of Public Health | CT Department of Public Health | Claire Pearson, Tu N. Nguyen, Kutluhan Incekara |
| EPI_ISL_14666780 | Public Health Authority of the Slovak Republic | Laboratory of Genomics and Bioinformatics, Comenius University Science Park | Tomáš Szemes, Edita Staroňová, Elena Tichá, Lucia Ševčíková, Terézia Vrabľová, Tatiana Sedláčková, Miroslav Böhmer, Jaroslav Budiš, Pavol Mišenko |
| EPI_ISL_14676265 | Centro de Desenvolvimento Científico e Tecnológico (CDCT)/CEVS/SES-RS | Centro de Desenvolvimento Científico e Tecnológico (CDCT)/CEVS/SES-RS | Richard Steiner Salvato, Regina Bones Barcellos, Fernanda Marques Godinho |
| EPI_ISL_14707250 | Charité Universitätsmedizin Berlin, Institut für Virologie | Charité Universitätsmedizin Berlin, Institut für Virologie | Julia Schneider, Victor M Corman, Terry C Jones, Christian Drosten |
| EPI_ISL_14721255, EPI_ISL_14721256, EPI_ISL_14721257, EPI_ISL_14721258, EPI_ISL_14721259, EPI_ISL_14721260, EPI_ISL_14721261, EPI_ISL_14721262, EPI_ISL_14721263, EPI_ISL_14721264, EPI_ISL_14721265 | | | |
| see above | National Public Health Laboratory, National Centre for Infectious Diseases | National Public Health Laboratory, National Centre for Infectious Diseases | Yichen Ding, Benny Yeo, Daniel Lim, Zhenyang Zhou, Royce Ang, Samuel Loo, Lin Cui, Raymond Tzer Pin Lin |
| EPI_ISL_14736407 | California Department of Public Health | California Department of Public Health | Viral and Rickettsial Disease Laboratory |
| EPI_ISL_14752090, EPI_ISL_14752093, EPI_ISL_14752094, EPI_ISL_14752096 | Environmental, Agricultural, and Occupational Health, University of Nebraska Medical Center | Environmental, Agricultural, and Occupational Health, University of Nebraska Medical Center | Tegomoh,B., Cross,S.T., Chapman,R.C., Bernhard,K., McCutchen,E.L., Fauver,J.R., Pratt,C.B., Warden,D.E., Iwen,P.C., Donahue,M. and Wiley,M.R. |
| EPI_ISL_14752098, EPI_ISL_14752145, EPI_ISL_14752175, EPI_ISL_14752177, EPI_ISL_14752180, EPI_ISL_14752189, EPI_ISL_14752193, EPI_ISL_14752201, EPI_ISL_14752202, EPI_ISL_14752211, EPI_ISL_14752215, EPI_ISL_14752231, EPI_ISL_14752234, EPI_ISL_14752240, EPI_ISL_14752251, EPI_ISL_14752255 | | | |
| see above | Department of Infectious Diseases, National Institute of Health Doutor Ricardo Jorge (INSA) | Department of Infectious Diseases, National Institute of Health Doutor Ricardo Jorge (INSA) | Isidro,J., Borges,V., Pinto,M., Sobral,D., Santos,J., Nunes,A., Mixao,V., Ferreira,R., Santos,D., Duarte,S., Vieira,L., Borrego,M.J., Nuncio,S., Lopes de Carvalho,I., Pelerito,A., Cordeiro,R. and Gomes,J.P. |
| EPI_ISL_14772317 | Policlinica Jacare Wilson Federzoni Cabreuva | Instituto Adolfo Lutz Strategic Laboratory | Claudio Tavares Sacchi, Karoline Rodrigues Campos, Ariadne Ferreira Amarante, Marlon Benedito Nascimento Santos, Alex Domingos Reis, Adriano Abbud, Adriana Bugno |
| EPI_ISL_14772318 | Secretaria Municipal de Saude de Sertaozinho | Instituto Adolfo Lutz Strategic Laboratory | Claudio Tavares Sacchi, Karoline Rodrigues Campos, Ariadne Ferreira Amarante, Marlon Benedito Nascimento Santos, Alex Domingos Reis, Adriano Abbud, Adriana Bugno |
| EPI_ISL_14772912 | USF Jardim Oratorio | Instituto Adolfo Lutz Strategic Laboratory | Claudio Tavares Sacchi, Karoline Rodrigues Campos, Ariadne Ferreira Amarante, Marlon Benedito Nascimento Santos, Alex Domingos Reis, Adriano Abbud, Adriana Bugno |
| EPI_ISL_14772913 | Vigilancia Epidemiologica Jardinopolis - SP | Instituto Adolfo Lutz Strategic Laboratory | Claudio Tavares Sacchi, Karoline Rodrigues Campos, Ariadne Ferreira Amarante, Marlon Benedito Nascimento Santos, Alex Domingos Reis, Adriano Abbud, Adriana Bugno |
| EPI_ISL_14772914 | Pronto Atendimento Infantil e entral de Quimioterapia SJrpreto | Instituto Adolfo Lutz Strategic Laboratory | Claudio Tavares Sacchi, Karoline Rodrigues Campos, Ariadne Ferreira Amarante, Marlon Benedito Nascimento Santos, Alex Domingos Reis, Adriano Abbud, Adriana Bugno |
| EPI_ISL_14773001 | CEDIC CTA | Instituto Adolfo Lutz Strategic Laboratory | Claudio Tavares Sacchi, Karoline Rodrigues Campos, Ariadne Ferreira Amarante, Marlon Benedito Nascimento Santos, Alex Domingos Reis, Adriano Abbud, Adriana Bugno |
| EPI_ISL_14786290 | IRCCS Sacro Cuore Don Calabria Hospital, Department of Infectious, Tropical Diseases & Microbiology | Department of Infectious, Tropical Diseases & Microbiology,IRCCS Sacro Cuore Don Calabria Hospital | Michela Deiana, Antonio Mori, Concetta Castilletti, Chiara Piubelli, Denise Lavezzari, Elena Pomari |
| EPI_ISL_14786346 | IRCCS Sacro Cuore Don Calabria Hospital, Department of Infectious, Tropical Diseases & Microbiology | IRCCS Sacro Cuore Don Calabria Hospital, Department of Infectious, Tropical Diseases & Microbiology | Michela Deiana, Antonio Mori, Concetta Castilletti, Chiara Piubelli, Denise Lavezzari, Elena Pomari |
| EPI_ISL_14793992, EPI_ISL_14795058, EPI_ISL_14795084, EPI_ISL_14795085, EPI_ISL_14795259 | Erasmus Medical Center Department of Virology | Erasmus Medical Center Department of Virology | Bas Oude Munnink, Leonard Schuele, Marjan Boter, Babette Weller, Babs Verstrepen, Richard Molenkamp, Janette Rahamat-Langendoen, Reina Sikkema, Marion Koopmans |
| EPI_ISL_14804638, EPI_ISL_14804639, EPI_ISL_14804640, EPI_ISL_14804641, EPI_ISL_14804642, EPI_ISL_14804643, EPI_ISL_14804644, EPI_ISL_14804645, | Nebraska Public Health Laboratory | University of Nebraska Medical Center, Oklahoma Pathogen Genomics Consortium | Chapman,R.C., Bernhard,K., McCutchen,E.L., Fauver,J.R., O'Dell,J.X., Mannell,M., Wiley,M.R., Cross,S.T. |

| | | | |
|---|---|---|---|
| EPI_ISL_14804646, EPI_ISL_14804647 | | | |
| EPI_ISL_14809096 | AMA Capao Redondo | Instituto Adolfo Lutz Strategic Laboratory | Claudio Tavares Sacchi, Karoline Rodrigues Campos, Ariadne Ferreira Amarante, Marlon Benedito Nascimento Santos, Alex Domingos Reis, Adriano Abbud, Adriana Bugno |
| EPI_ISL_14809097 | Pronto Socorro Municipal de Taubate | Instituto Adolfo Lutz Strategic Laboratory | Claudio Tavares Sacchi, Karoline Rodrigues Campos, Ariadne Ferreira Amarante, Marlon Benedito Nascimento Santos, Alex Domingos Reis, Adriano Abbud, Adriana Bugno |
| EPI_ISL_14809098 | Laboratorio Municipal de Piracicaba | Instituto Adolfo Lutz Strategic Laboratory | Claudio Tavares Sacchi, Karoline Rodrigues Campos, Ariadne Ferreira Amarante, Marlon Benedito Nascimento Santos, Alex Domingos Reis, Adriano Abbud, Adriana Bugno |
| EPI_ISL_14809099 | Centro de Saude Gabriel de Lara | Instituto Adolfo Lutz Strategic Laboratory | Claudio Tavares Sacchi, Karoline Rodrigues Campos, Ariadne Ferreira Amarante, Marlon Benedito Nascimento Santos, Alex Domingos Reis, Adriano Abbud, Adriana Bugno |
| EPI_ISL_14809100 | Secretaria Municipal da Saude de Joanopolis | Instituto Adolfo Lutz Strategic Laboratory | Claudio Tavares Sacchi, Karoline Rodrigues Campos, Ariadne Ferreira Amarante, Marlon Benedito Nascimento Santos, Alex Domingos Reis, Adriano Abbud, Adriana Bugno |
| EPI_ISL_14810370, EPI_ISL_14810404, EPI_ISL_14810405, EPI_ISL_14810406, EPI_ISL_14810407 | Erasmus Medical Center Department of Virology | Erasmus Medical Center Department of Virology | Leonard Schuele, Bas Oude Munnink, Marjan Boter, Babette Weller, Babs Verstrepen, Richard Molenkamp, Janette Rahamat-Langendoen, Reina Sikkema, Marion Koopmans |
| EPI_ISL_14818590, EPI_ISL_14818595, EPI_ISL_14818600, EPI_ISL_14818606 | Los Angeles County Public Health Laboratories | Los Angeles County Public Health Laboratories | P. Hemarajata et al. |
| EPI_ISL_14818783, EPI_ISL_14818784, EPI_ISL_14818785, EPI_ISL_14818786, EPI_ISL_14818787, EPI_ISL_14818788, EPI_ISL_14818789, EPI_ISL_14818790, EPI_ISL_14818791, EPI_ISL_14818792, EPI_ISL_14818793, EPI_ISL_14818794, EPI_ISL_14818795, EPI_ISL_14818796, EPI_ISL_14818797, EPI_ISL_14818798, EPI_ISL_14818799, EPI_ISL_14818800, EPI_ISL_14818801, EPI_ISL_14818802, EPI_ISL_14818803, EPI_ISL_14818804, EPI_ISL_14818805, EPI_ISL_14818806, EPI_ISL_14818807, EPI_ISL_14818808, EPI_ISL_14818809, EPI_ISL_14818810, EPI_ISL_14818811, EPI_ISL_14818812, EPI_ISL_14818813, EPI_ISL_14818814, EPI_ISL_14818815, EPI_ISL_14818816, EPI_ISL_14818817, EPI_ISL_14818818, EPI_ISL_14818819, EPI_ISL_14818820, EPI_ISL_14818821, EPI_ISL_14818822 | | | |
| see above | Laboratorio de Referencia Nacional de Virus Inmunoprevenibles. Centro Nacional de Salud Publica. Instituto Nacional de Salud. | Laboratorio de Referencia Nacional de Virus Inmunoprevenibles. Centro Nacional de Salud Publica. Instituto Nacional de Salud. | Carlos Padilla Rojas, Veronica Hurtado Vela, Iris Silva Molina, Luren Sevilla Castañeda, Victor Jimenez Vasquez, Luis Barcena Flores, Alicia Nuñez Llanos, Kelly Izarra Rojas, Karla Vasquez Cajachahua, Estela Huaman Angeles, Jorge Giraldo Chavez, Lilian Huarca Balbin, Maria Sandra Villar Saavedra, Henri Bailon Calderon, Lely Solari Zerpa, Gloria Arotinco Garayar. Equipo de vigilancia genomica del Instituto Nacional de Salud. |
| EPI_ISL_14838586, EPI_ISL_14838587, EPI_ISL_14838588, EPI_ISL_14838589 | Pathogen Genomics Lab, National Institute for Biomedical Research (INRB) | Pathogen Genomics Lab, National Institute for Biomedical Research (INRB) | Placide Mbala-Kingebeni, Eddy Kinganda-Lusamaki, Adrienne Amuri-Aziza, Elisabeth Pukuta, Catherine Pratt, Nicolas Fernandez, Emmanuel Lokilo Lofiko, Gradi Luakanda Ndelemo, Francisca Muyembe Mawete, Jean Claude Makangara Cigolo, Elisabeth Muyamuna, Raphaël Lumembe Numbi, Gabriel Kabamba Lungenyi, Prince Akil Bandali, Pauline Musuamba Kayembe, Rilia Ola Mpumbe, Emile Malembi, Emmanuel Hasivirwe Vakaniaki, Andrew Rambaut, Nick Loman, Kristian Andersen, Michael Wiley, Ahidjo Ayouba, Steve Ahuka-Mundeke, Martine Peeters, Eric Delaporte, Jean-Jacques Muyembe Tamfum |
| EPI_ISL_14842156, EPI_ISL_14842157 | Division of High Consequence Pathogens and Pathology (DHCPP)-PRB, CDC | Division of High Consequence Pathogens and Pathology (DHCPP)-PRB, CDC | Gigante,C.M., Pavlick,J., Zhao,H., Batra,D., Hetrick,E.E., Howard,D.T., Kovar,L., Seabolt,M.H., Weigand,M.R., Burroughs,M., Lee,J., Wilkins,K., McCollum,A., Hutson,C., Davidson,W., Rao,A., Parrott,T. and Li,Y. |
| EPI_ISL_14863040, EPI_ISL_14863041, EPI_ISL_14863042, EPI_ISL_14863043, EPI_ISL_14863045, EPI_ISL_14863046, EPI_ISL_14863047 | Molecular Epidemiology, Idaho Bureau of Laboratories | Molecular Epidemiology, Idaho Bureau of Laboratories | Ceniseros,A. |
| EPI_ISL_14865785 | UBS J COPA | Instituto Adolfo Lutz Strategic Laboratory | Claudio Tavares Sacchi, Karoline Rodrigues Campos, Ariadne Ferreira Amarante, Marlon Benedito Nascimento Santos, Alex Domingos Reis, Adriano Abbud, Adriana Bugno |
| EPI_ISL_14866481 | PR S da Familia Unidade de Saude Adalberto Rocha | Instituto Adolfo Lutz Strategic Laboratory | Claudio Tavares Sacchi, Karoline Rodrigues Campos, Ariadne Ferreira Amarante, Marlon Benedito Nascimento Santos, Alex Domingos Reis, Adriano Abbud, Adriana Bugno |
| EPI_ISL_14866751 | Pronto Socorro da Vila Dirce | Instituto Adolfo Lutz Strategic Laboratory | Claudio Tavares Sacchi, Karoline Rodrigues Campos, Ariadne Ferreira Amarante, Marlon Benedito Nascimento Santos, Alex Domingos Reis, Adriano Abbud, Adriana Bugno |
| EPI_ISL_14866752 | Secretaria Municipal de Saude Sao Carlos | Instituto Adolfo Lutz Strategic Laboratory | Claudio Tavares Sacchi, Karoline Rodrigues Campos, Ariadne Ferreira Amarante, Marlon Benedito Nascimento Santos, Alex Domingos Reis, Adriano Abbud, Adriana Bugno |
| EPI_ISL_14887952, EPI_ISL_14887953, EPI_ISL_14887954, EPI_ISL_14887955, EPI_ISL_14887956, EPI_ISL_14887957, EPI_ISL_14887958, EPI_ISL_14887959, EPI_ISL_14887960, EPI_ISL_14887961, EPI_ISL_14887962, EPI_ISL_14887963, EPI_ISL_14887964, EPI_ISL_14887965, EPI_ISL_14887966, EPI_ISL_14887967, EPI_ISL_14887968, EPI_ISL_14887969, EPI_ISL_14887970, EPI_ISL_14887971, EPI_ISL_14887972, EPI_ISL_14887973, EPI_ISL_14887974, EPI_ISL_14887975, EPI_ISL_14887976, EPI_ISL_14887977, EPI_ISL_14887978, EPI_ISL_14887979, EPI_ISL_14887980, EPI_ISL_14887981, EPI_ISL_14887982, EPI_ISL_14887983, EPI_ISL_14887984, EPI_ISL_14887985, EPI_ISL_14887986, EPI_ISL_14887987, EPI_ISL_14887988, EPI_ISL_14887989, EPI_ISL_14887990 | | | |
| see above | Viral Genotyping Reference Laboratory, Royal Infirmary of Edinburgh | Viral Genotyping Reference Laboratory, Royal Infirmary of Edinburgh | McHugh,M.P., Maloney,D., Parker,A., Mathers,K., Dewar,R., Kenicer,J., Cotton,S., Wild,J. and Templeton,K.E. |
| EPI_ISL_14917565, EPI_ISL_14917571, EPI_ISL_14917573, EPI_ISL_14917580 | Los Angeles County Public Health Laboratories | Los Angeles County Public Health Laboratories | P. Hemarajata et al. |
| EPI_ISL_14934498, EPI_ISL_14934500, EPI_ISL_14934505, EPI_ISL_14934506, EPI_ISL_14934507, EPI_ISL_14934509, EPI_ISL_14934528, EPI_ISL_14934533, EPI_ISL_14934554, EPI_ISL_14934560, EPI_ISL_14934564, EPI_ISL_14934566, EPI_ISL_14934572, EPI_ISL_14934580, EPI_ISL_14934612 | | | |
| see above | Department of Infectious Diseases, National Institute of Health Doutor Ricardo Jorge, Portugal (INSA) | Department of Infectious Diseases, National Institute of Health Doutor Ricardo Jorge, Portugal (INSA) | Isidro,J., Borges,V., Pinto,M., Sobral,D., Santos,J., Nunes,A., Mixao,V., Ferreira,R., Santos,D., Duarte,S., Vieira,L., Borrego,M.J., Nuncio,S., Lopes de Carvalho,I., Pelerito,A., Cordeiro,R. and Gomes,J.P. |
| EPI_ISL_14944276, EPI_ISL_14944277, EPI_ISL_14944278, EPI_ISL_14944279, EPI_ISL_14944280, EPI_ISL_14944281, EPI_ISL_14944282, EPI_ISL_14944283, EPI_ISL_14944284, EPI_ISL_14944285, EPI_ISL_14944286, EPI_ISL_14944287, EPI_ISL_14944288, EPI_ISL_14944289, EPI_ISL_14944290, EPI_ISL_14944291, EPI_ISL_14944292, EPI_ISL_14944293, EPI_ISL_14944294 | | | |
| see above | Rhode Island State Health Laboratory | Rhode Island State Health Laboratory | Kristin Carpenter-Azevedo, Sean Sierra-Patev, Richard C. Huard |
| EPI_ISL_14945299 | Department of Microbiology, The University of Hong Kong | Department of Microbiology, The University of Hong Kong | Kelvin K.W. To, Kwok-Yung Yuen |
| EPI_ISL_14977306, EPI_ISL_14977307, EPI_ISL_14977308, EPI_ISL_14977309, EPI_ISL_14977310 | Environmental, Agricultural, and Occupational Health, University of Nebraska Medical Center | Environmental, Agricultural, and Occupational Health, University of Nebraska Medical Center | Tegomoh,B., Cross,S.T., Chapman,R.C., Bernhard,K., McCutchen,E.L., Fauver,J.R., Pratt,C.B., Warden,D.E., Iwen,P.C., Donahue,M. and Wiley,M.R. |
| EPI_ISL_14980972, EPI_ISL_14981151 | Kingston Health Sciences Centre | Kingston Health Sciences Centre | Calvin Sjaarda, Henry Wong, Nick Buchner, Drew Roberts, Phung Ta, Jacob Whalen, Sheri Levesque, Prameet Sheth |
| EPI_ISL_14994740 | UBS Vila California Zeilival Bruscagin | Instituto Adolfo Lutz Strategic Laboratory | Claudio Tavares Sacchi, Karoline Rodrigues Campos, Ariadne Ferreira Amarante, Marlon Benedito Nascimento Santos, Alex Domingos Reis, Adriano Abbud, Adriana Bugno |
| EPI_ISL_14995578 | Hosp. Municipal de Ilhabela Gov. Mario Covas Jr. | Instituto Adolfo Lutz Strategic Laboratory | Claudio Tavares Sacchi, Karoline Rodrigues Campos, Ariadne Ferreira Amarante, Marlon Benedito Nascimento Santos, Alex Domingos Reis, Adriano Abbud, Adriana Bugno |
| EPI_ISL_14995579 | Secretaria Municipal de Saude de Feira de Santana | Instituto Adolfo Lutz Strategic Laboratory | Claudio Tavares Sacchi, Karoline Rodrigues Campos, Ariadne Ferreira Amarante, Marlon Benedito Nascimento Santos, Alex Domingos Reis, Adriano Abbud, Adriana Bugno |
| EPI_ISL_14995580 | UBS Alexander Fleming Simioni | Instituto Adolfo Lutz Strategic Laboratory | Claudio Tavares Sacchi, Karoline Rodrigues Campos, Ariadne Ferreira Amarante, Marlon Benedito Nascimento Santos, Alex Domingos Reis, Adriano Abbud, Adriana Bugno |
| EPI_ISL_14995582 | Hosp. Municipla. Dr. Jose de Carvalho Florence | Instituto Adolfo Lutz Strategic Laboratory | Claudio Tavares Sacchi, Karoline Rodrigues Campos, Ariadne Ferreira Amarante, Marlon Benedito Nascimento Santos, Alex Domingos Reis, Adriano Abbud, Adriana Bugno |
| EPI_ISL_14995583 | UBS Agua Rasa | Instituto Adolfo Lutz Strategic Laboratory | Claudio Tavares Sacchi, Karoline Rodrigues Campos, Ariadne Ferreira Amarante, Marlon Benedito Nascimento Santos, Alex Domingos Reis, Adriano Abbud, Adriana Bugno |
| EPI_ISL_14995585 | Pronto Socorro Municipal do Promorar | Instituto Adolfo Lutz Strategic Laboratory | Claudio Tavares Sacchi, Karoline Rodrigues Campos, Ariadne Ferreira Amarante, Marlon Benedito Nascimento Santos, Alex Domingos Reis, Adriano Abbud, Adriana Bugno |
| EPI_ISL_14995586 | UPA Centro | Instituto Adolfo Lutz Strategic Laboratory | Claudio Tavares Sacchi, Karoline Rodrigues Campos, Ariadne Ferreira Amarante, Marlon Benedito Nascimento Santos, Alex Domingos Reis, Adriano Abbud, Adriana Bugno |
| EPI_ISL_14995587 | Centro de Saude 24 horas | Instituto Adolfo Lutz Strategic Laboratory | Claudio Tavares Sacchi, Karoline Rodrigues Campos, Ariadne Ferreira Amarante, Marlon Benedito Nascimento Santos, Alex Domingos Reis, Adriano Abbud, Adriana Bugno |
| EPI_ISL_14995589 | Cresser Centro de Referencia da Saúde Sexual e Reprodutiva | Instituto Adolfo Lutz Strategic Laboratory | Claudio Tavares Sacchi, Karoline Rodrigues Campos, Ariadne Ferreira Amarante, Marlon Benedito Nascimento Santos, Alex Domingos Reis, Adriano Abbud, Adriana Bugno |
| EPI_ISL_14995590, EPI_ISL_14995591 | Instituto de Infectologia Emilio Ribas | Instituto Adolfo Lutz Strategic Laboratory | Claudio Tavares Sacchi, Karoline Rodrigues Campos, Ariadne Ferreira Amarante, Marlon Benedito Nascimento Santos, Alex Domingos Reis, Adriano Abbud, Adriana Bugno |
| EPI_ISL_14995592 | Unidade de Pronto Atendimento Cipo | Instituto Adolfo Lutz Strategic Laboratory | Claudio Tavares Sacchi, Karoline Rodrigues Campos, Ariadne Ferreira Amarante, Marlon Benedito Nascimento Santos, Alex Domingos Reis, Adriano Abbud, Adriana Bugno |
| EPI_ISL_14995593 | SAE DST / Aids Ipiranga Jose Francisco Araujo | Instituto Adolfo Lutz Strategic Laboratory | Claudio Tavares Sacchi, Karoline Rodrigues Campos, Ariadne Ferreira Amarante, Marlon Benedito Nascimento Santos, Alex Domingos Reis, Adriano Abbud, Adriana Bugno |
| EPI_ISL_14995611 | UBS Horto Florestal | Instituto Adolfo Lutz Strategic Laboratory | Claudio Tavares Sacchi, Karoline Rodrigues Campos, Ariadne Ferreira Amarante, Marlon Benedito Nascimento Santos, Alex Domingos Reis, Adriano Abbud, Adriana Bugno |
| EPI_ISL_14995619 | Hosp. Tereza de Lisieux | Instituto Adolfo Lutz Strategic Laboratory | Claudio Tavares Sacchi, Karoline Rodrigues Campos, Ariadne Ferreira Amarante, Marlon Benedito Nascimento Santos, Alex Domingos Reis, Adriano Abbud, Adriana Bugno |
| EPI_ISL_14995622 | UBS Parque Meia Lua | Instituto Adolfo Lutz Strategic Laboratory | Claudio Tavares Sacchi, Karoline Rodrigues Campos, Ariadne Ferreira Amarante, Marlon Benedito Nascimento Santos, Alex Domingos Reis, Adriano Abbud, Adriana Bugno |
| EPI_ISL_14995649 | Instituto de Infectologia Emilio Ribas | Instituto Adolfo Lutz Strategic Laboratory | Claudio Tavares Sacchi, Karoline Rodrigues Campos, Ariadne Ferreira Amarante, Marlon Benedito Nascimento Santos, Alex Domingos Reis, Adriano Abbud, Adriana Bugno |
| EPI_ISL_14995652 | Hosp. Dr. Osiris Florindo Coelho Ferraz de Vasconcelos | Instituto Adolfo Lutz Strategic Laboratory | Claudio Tavares Sacchi, Karoline Rodrigues Campos, Ariadne Ferreira Amarante, Marlon Benedito Nascimento Santos, Alex Domingos Reis, Adriano Abbud, Adriana Bugno |
| EPI_ISL_14995653 | Unidade Basica de Saude Vila Cristina | Instituto Adolfo Lutz Strategic Laboratory | Claudio Tavares Sacchi, Karoline Rodrigues Campos, Ariadne Ferreira Amarante, Marlon Benedito Nascimento Santos, Alex Domingos Reis, Adriano Abbud, Adriana Bugno |
| EPI_ISL_14995723 | Unidade Mista de Atendimento Infantil Carapicuiba | Instituto Adolfo Lutz Strategic Laboratory | Claudio Tavares Sacchi, Karoline Rodrigues Campos, Ariadne Ferreira Amarante, Marlon Benedito Nascimento Santos, Alex Domingos Reis, Adriano Abbud, Adriana Bugno |
| EPI_ISL_14995724 | Hosp. Carlos Chagas | Instituto Adolfo Lutz Strategic Laboratory | Claudio Tavares Sacchi, Karoline Rodrigues Campos, Ariadne Ferreira Amarante, Marlon Benedito Nascimento Santos, Alex Domingos Reis, Adriano Abbud, Adriana Bugno |
| EPI_ISL_15003284, EPI_ISL_15003285, EPI_ISL_15003286, EPI_ISL_15003287, EPI_ISL_15003288, EPI_ISL_15003289, EPI_ISL_15003290, EPI_ISL_15003291, EPI_ISL_15003292, EPI_ISL_15003293, EPI_ISL_15003294, EPI_ISL_15003295, EPI_ISL_15003296, EPI_ISL_15003297 | | | |
| see above | Rhode Island State Health Laboratory | Rhode Island State Health Laboratory | Kristin Carpenter-Azevedo, Sean Sierra-Patev, Richard C. Huard |
| EPI_ISL_15005641 | Chongqing Municipal Center for Disease Control and Prevention | Chongqing Municipal Center for Disease Control and Prevention | Sheng Ye, Yun Tang, Shuang Chen, Mingyue Wang, Zhangping Tan, Zhen Yu |
| EPI_ISL_15008576, EPI_ISL_15008577 | Indian Council of Medical Research-National Institute of Virology | Indian Council of Medical Research-National Institute of Virology | Pragya Yadav, Rima Sahay, Anita Aich Shete, Sreelekshmy Mohandas, Priya Abraham |
| EPI_ISL_15016099, EPI_ISL_15016100, EPI_ISL_15016101, EPI_ISL_15016102, EPI_ISL_15016103 | Florida Bureau of Public Health Laboratories | Florida Bureau of Public Health Laboratories | Sarah Schmedes, Brenna McGruder, George Churchwell, Maria Pedrosa, Phil A. Lee |
| EPI_ISL_15038825 | Johns Hopkins University School of Medicine | Johns Hopkins School of Medicine Pathology | Amary Fall,Katharine Uhteg, C Paul Morris ,Heba H. Mostafa |
| EPI_ISL_15039575, EPI_ISL_15039652 | Johns Hopkins University School of Medicine | Johns Hopkins School of Medicine Pathology | Amary Fall, Katharine Uhteg , C Paul Morris, Heba H. Mostafa |
| EPI_ISL_15055820 | Sicilian Regional Laboratory - AOUP "P. Giaccone" - University of Palermo | Sicilian Regional Laboratory - AOUP "P. Giaccone" - University of Palermo | Fabio Tramuto, Carmelo Massimo Maida, Giulia Randazzo, Valeria Guzzetta, Walter Mazzucco, Giorgio Graziano, Vincenzo Restivo, Claudio Costantino, Francesco Vitale |
| EPI_ISL_15098376, EPI_ISL_15098377, EPI_ISL_15098378, EPI_ISL_15098379, EPI_ISL_15098380, EPI_ISL_15098381, EPI_ISL_15098382, EPI_ISL_15098383, EPI_ISL_15098384, EPI_ISL_15098385, EPI_ISL_15098386, EPI_ISL_15098387, EPI_ISL_15098388, EPI_ISL_15098389, EPI_ISL_15098390, EPI_ISL_15098391, EPI_ISL_15098392, EPI_ISL_15098393, EPI_ISL_15098394, EPI_ISL_15098395 | | | |
| see above | CT Department of Public Health | CT Department of Public Health | Claire Pearson, Tu N. Nguyen, Kutluhan Incekara |
| EPI_ISL_15104903 | Institute for Virology, Philipps-University Marburg | Institute for Virology, Philipps-University Marburg | Eickmann, M., Lier, C., Kowalski, K., Kraft, F., Becker, S. |

| | | | |
|---|---|---|---|
| EPI_ISL_15116301, EPI_ISL_15116302, EPI_ISL_15116303, EPI_ISL_15116304, EPI_ISL_15116305, EPI_ISL_15116306, EPI_ISL_15116307 | Centers for Disease Control & Prevention (CDC), Division of High Consequence Pathogens and Pathology (DHCPP-PRB) | Centers for Disease Control & Prevention (CDC), Division of High Consequence Pathogens and Pathology (DHCPP-PRB) | Gigante,C.M., Plumb,M., Ruprecht,A., Zhao,H., Wicker,V., Wilkins,K., Matheny,A., Khan,T., Davidson,W., Sheth,M., Burgin,A., Burroughs,M., Padilla,J., Lee,J.S., Batra,D., Hetrick,E.E., Howard,D.T., Garfin,J., Tate,L., Hubsmith,S.J., Mendoza,R.M., Stanek,D., Gillani,S., Lee,M., Mangla,A., Blythe,D., SierraPatev,S., Carpenter-Azevedo,K., Huard,R.C., Gallagher,G., Hall,J., Ash,S., Kovar,L., Seabolt,M.H., Weigand,M.R., Damon,I., Satheshkumar,P.S., McCollum,A.M., Hutson,C.L., Wang,X. and Li,Y. |
| EPI_ISL_15120464, EPI_ISL_15120480, EPI_ISL_15120492 | Los Angeles County Public Health Laboratories | Los Angeles County Public Health Laboratories | P. Hemarajata et al. |
| EPI_ISL_15158317, EPI_ISL_15158318, EPI_ISL_15158319, EPI_ISL_15158320, EPI_ISL_15158321, EPI_ISL_15158323, EPI_ISL_15158324, EPI_ISL_15158325, EPI_ISL_15158326, EPI_ISL_15158327, EPI_ISL_15158328, EPI_ISL_15158329, EPI_ISL_15158330, EPI_ISL_15158331, EPI_ISL_15158332, EPI_ISL_15158333, EPI_ISL_15158334, EPI_ISL_15158335, EPI_ISL_15158337, EPI_ISL_15158338, EPI_ISL_15158339, EPI_ISL_15158340, EPI_ISL_15158341, EPI_ISL_15158342, EPI_ISL_15158343, EPI_ISL_15158344, EPI_ISL_15158345, EPI_ISL_15158346, EPI_ISL_15158347, EPI_ISL_15158348, EPI_ISL_15158349, EPI_ISL_15158350, EPI_ISL_15158351, EPI_ISL_15158352, EPI_ISL_15158353, EPI_ISL_15158354, EPI_ISL_15158355, EPI_ISL_15158356, EPI_ISL_15158357, EPI_ISL_15158358, EPI_ISL_15158359, EPI_ISL_15158360, EPI_ISL_15158361, EPI_ISL_15158362, EPI_ISL_15158363, EPI_ISL_15158364, EPI_ISL_15158365, EPI_ISL_15158366, EPI_ISL_15158367, EPI_ISL_15158368, EPI_ISL_15158369, EPI_ISL_15158370, EPI_ISL_15158371, EPI_ISL_15158372, EPI_ISL_15158373, EPI_ISL_15158375, EPI_ISL_15158376, EPI_ISL_15158380, EPI_ISL_15158381, EPI_ISL_15158382, EPI_ISL_15158383, EPI_ISL_15158384, EPI_ISL_15158385, EPI_ISL_15158386, EPI_ISL_15158388, EPI_ISL_15158389, EPI_ISL_15158390, EPI_ISL_15158391, EPI_ISL_15158392, EPI_ISL_15158393, EPI_ISL_15158395, EPI_ISL_15158396, EPI_ISL_15158397, EPI_ISL_15158398 | | | |
| see above | Molecular Biology, Microbiology, and Biochemistry, Southern Illinois University | Molecular Biology, Microbiology, and Biochemistry, Southern Illinois University | Gagnon,K.T. |
| EPI_ISL_15165602, EPI_ISL_15165603, EPI_ISL_15165604, EPI_ISL_15165605, EPI_ISL_15165606, EPI_ISL_15165607, EPI_ISL_15165608, EPI_ISL_15165609, EPI_ISL_15165610, EPI_ISL_15165611, EPI_ISL_15165612, EPI_ISL_15165613, EPI_ISL_15165614, EPI_ISL_15165615, EPI_ISL_15165616, EPI_ISL_15165617, EPI_ISL_15165618 | | | |
| see above | Centro de Desenvolvimento Científico e Tecnológico (CDCT), Centro Estadual de Vigilância em Saúde (CEVS) da Secretaria Estadual da Saúde (SES-RS) | Centro de Desenvolvimento Científico e Tecnológico (CDCT), Centro Estadual de Vigilância em Saúde (CEVS) da Secretaria Estadual da Saúde (SES-RS) | Richard Steiner Salvato, Fernanda Marques Godinho, Regina Bones Barcellos, Patricia Sesterheim, Amanda Pellenz Ruivo, Viviane Horn de Melo, Júlio Augusto Schroder |
| EPI_ISL_15226680 | Los Angeles County Public Health Laboratories | Los Angeles County Public Health Laboratories | P. Hemarajata et al. |
| EPI_ISL_15247221, EPI_ISL_15247222, EPI_ISL_15247223, EPI_ISL_15247224, EPI_ISL_15247225, EPI_ISL_15247226, EPI_ISL_15247227, EPI_ISL_15247228 | Medical University of Vienna Center for Virology | Medical University of Vienna Center for Virology | Jeremy V. Camp, Monika Redlberger-Fritz, Stephan W. Aberle |
| EPI_ISL_15263355 | Sicilian Regional Laboratory - AOUP "P. Giaccone" - University of Palermo | Sicilian Regional Laboratory - AOUP "P. Giaccone" - University of Palermo | Fabio Tramuto, Carmelo Massimo Maida, Giulia Randazzo, Valeria Guzzetta, Walter Mazzucco, Giorgio Graziano, Vincenzo Restivo, Claudio Costantino, Francesco Vitale |
| EPI_ISL_15264003, EPI_ISL_15266514, EPI_ISL_15266516, EPI_ISL_15266518, EPI_ISL_15266615 | Erasmus Medical Center Department of Virology | Erasmus Medical Center Department of Virology | Leonard Schuele, Bas Oude Munnink, Marjan Boter, Babette Weller, Babs Verstrepen, Richard Molenkamp, Janette Rahamat-Langendoen, Reina Sikkema, Marion Koopmans |
| EPI_ISL_15266843 | Erasmus Medical Center Department of Virology | Erasmus Medical Center Department of Virology | Leonard Schuele, Bas Oude Munnink, Marjan Boter, David Nieuwenhuijse, Babette Weller, Babs Verstrepen, Richard Molenkamp, Janette Rahamat-Langendoen, Reina Sikkema, Marion Koopmans |
| EPI_ISL_15266988, EPI_ISL_15267015, EPI_ISL_15267029, EPI_ISL_15267031, EPI_ISL_15267800, EPI_ISL_15268281, EPI_ISL_15269049 | Erasmus Medical Center Department of Virology | Erasmus Medical Center Department of Virology | Leonard Schuele, Bas Oude Munnink, Marjan Boter, Babette Weller, Babs Verstrepen, Richard Molenkamp, Janette Rahamat-Langendoen, Reina Sikkema, Marion Koopmans |
| EPI_ISL_15269237, EPI_ISL_15269238, EPI_ISL_15269375, EPI_ISL_15269384, EPI_ISL_15269386, EPI_ISL_15269387, EPI_ISL_15269388, EPI_ISL_15269544, EPI_ISL_15269595, EPI_ISL_15269598 | Erasmus Medical Center Department of Virology | Erasmus Medical Center Department of Virology | Leonard Schuele, Bas Oude Munnink, Marjan Boter, David Nieuwenhuijse, Babette Weller, Babs Verstrepen, Richard Molenkamp, Janette Rahamat-Langendoen, Reina Sikkema, Marion Koopmans |
| EPI_ISL_15269698, EPI_ISL_15269699, EPI_ISL_15269702, EPI_ISL_15269704, EPI_ISL_15269708 | Erasmus Medical Center Department of Virology | Erasmus Medical Center Department of Virology | Leonard Schuele, Bas Oude Munnink, Marjan Boter, Babette Weller, Babs Verstrepen, Richard Molenkamp, Janette Rahamat-Langendoen, Reina Sikkema, Marion Koopmans |
| EPI_ISL_15292976, EPI_ISL_15292977, EPI_ISL_15292978, EPI_ISL_15292979, EPI_ISL_15292980, EPI_ISL_15292981, EPI_ISL_15292982, EPI_ISL_15292983, EPI_ISL_15292984, EPI_ISL_15292985, EPI_ISL_15292986, EPI_ISL_15292987, EPI_ISL_15292988, EPI_ISL_15292989, EPI_ISL_15292990, EPI_ISL_15292991, EPI_ISL_15292992, EPI_ISL_15292993, EPI_ISL_15292994, EPI_ISL_15292995, EPI_ISL_15292996, EPI_ISL_15292998, EPI_ISL_15292999, EPI_ISL_15293000, EPI_ISL_15293001, EPI_ISL_15293002, EPI_ISL_15293003, EPI_ISL_15293004, EPI_ISL_15293005, EPI_ISL_15293006, EPI_ISL_15293007, EPI_ISL_15293008, EPI_ISL_15293009 | | | |
| see above | Institute of Health Carlos III, Bioinformatics Unit | Institute of Health Carlos III, Bioinformatics Unit | Cuesta,I. |
| EPI_ISL_15293815 | National Institute for Viral Disease Control and Prevention (IVDC), Chinese Center for Disease Control and Prevention , Beijing, China | National Institute for Viral Disease Control and Prevention (IVDC), Chinese Center for Disease Control and Prevention , Beijing, China | Wenjie Tan, Changcheng Wu, Ruhan A, Wenling Wang, Roujian Lu, Li Zhao, Baoying Huang, Fei Ye, Wenbo Xu |
| EPI_ISL_15317149 | Centers for Disease Control & Prevention (CDC), Division of High Consequence Pathogens and Pathology (DHCPP-PRB) | Centers for Disease Control & Prevention (CDC), Division of High Consequence Pathogens and Pathology (DHCPP-PRB) | Gigante,C.M., Xia,D., Zhao,H., Batra,D., Hetrick,E.E., Howard,D.T., Kovar,L., Seabolt,M.H., Weigand,M.R., Burroughs,M., Lee,J., Wilkins,K., McCollum,A., Hutson,C., Davidson,W., Rao,A., Plipat,N. and Li,Y. |
| EPI_ISL_15317153 | Centers for Disease Control & Prevention (CDC), Division of High Consequence Pathogens and Pathology (DHCPP-PRB) | Centers for Disease Control & Prevention (CDC), Division of High Consequence Pathogens and Pathology (DHCPP-PRB) | Gigante,C.M., Culbertson,M., Zhao,H., Batra,D., Hetrick,E.E., Howard,D.T., Kovar,L., Seabolt,M.H., Weigand,M.R., Burroughs,M., Lee,J., Wilkins,K., McCollum,A., Hutson,C., Davidson,W., Rao,A., Pope,B. and Li,Y. |
| EPI_ISL_15317158, EPI_ISL_15317159, EPI_ISL_15317160 | Centers for Disease Control & Prevention (CDC), Division of High Consequence Pathogens and Pathology (DHCPP-PRB) | Centers for Disease Control & Prevention (CDC), Division of High Consequence Pathogens and Pathology (DHCPP-PRB) | Gigante,C.M., Ventura,J., Zhao,H., Batra,D., Hetrick,E.E., Howard,D.T., Kovar,L., Seabolt,M.H., Weigand,M.R., Burroughs,M., Lee,J., Wilkins,K., McCollum,A., Hutson,C., Davidson,W., Rao,A., Nash,J. and Li,Y. |
| EPI_ISL_15321162, EPI_ISL_15321163, EPI_ISL_15321164, EPI_ISL_15321165, EPI_ISL_15321166, EPI_ISL_15321167, EPI_ISL_15321168, EPI_ISL_15321169, EPI_ISL_15321170, EPI_ISL_15321171, EPI_ISL_15321172, EPI_ISL_15321173, EPI_ISL_15321174, EPI_ISL_15321175, EPI_ISL_15321176, EPI_ISL_15321177 | | | |
| see above | Rhode Island State Health Laboratory | Rhode Island State Health Laboratory | Kristin Carpenter-Azevedo, Sean Sierra-Patev, Richard C. Huard |
| EPI_ISL_15325417, EPI_ISL_15325422 | Los Angeles County Public Health Laboratories | Los Angeles County Public Health Laboratories | P. Hemarajata et al. |
| EPI_ISL_15332324 | Centers for Disease Control & Prevention (CDC), Division of High Consequence Pathogens and Pathology (DHCPP-PRB) | Centers for Disease Control & Prevention (CDC), Division of High Consequence Pathogens and Pathology (DHCPP-PRB) | Gigante,C.M., Xia,D., Zhao,H., Batra,D., Hetrick,E.E., Howard,D.T., Kovar,L., Seabolt,M.H., Knipe,K., Burroughs,S., Lee,J., Wilkins,K., McCollum,A., Hutson,C., Davidson,W., Rao,A., Plipat,N. and Li,Y. |
| EPI_ISL_15332333 | Centers for Disease Control & Prevention (CDC), Division of High Consequence Pathogens and Pathology (DHCPP-PRB) | Centers for Disease Control & Prevention (CDC), Division of High Consequence Pathogens and Pathology (DHCPP-PRB) | Gigante,C.M., Lee,P., Zhao,H., Batra,D., Hetrick,E.E., Howard,D.T., Kovar,L., Seabolt,M.H., Knipe,K., Burroughs,S., Lee,J., Wilkins,K., McCollum,A., Hutson,C., Davidson,W., Rao,A., Stanek,D. and Li,Y. |
| EPI_ISL_15370031 | Laboratory for Diagnostics of Zoonoses and WHO Centre, Institute of Microbiology and Immunology, Faculty of Medicine, University of Ljubljana | Laboratory for Diagnostics of Zoonoses and WHO Centre, Institute of Microbiology and Immunology, Faculty of Medicine, University of Ljubljana | Zakotnik,S., Vlaj,D., Suljic,A., Zorec,T.M., Korva,M., Poljak,M. and Avsic Zupanc,T. |
| EPI_ISL_15370073 | Nigeria Centre for Disease Control, National Reference Laboratory | Chemical, Biological and Radiological Sciences, Defence Science and Technology Laboratory | Ndodo,N., Ashcroft,J., Lewandowski,K., Yinka-Ogunleye,A., Chukwu,C., Ahmad,A., King,D., Akinpelu,A., Maluquer de Motes,C., Ribeca,P., Summer,R.P., Rambaut,A., Chester,M., Maishman,T., Babatunde,O., Mba,N., Babatunde,O., Aruna,O., Pullan,S.T., Gannon,B., Brown,C., Ihekweazu,C., Adetifa,I. and Ulaeto,D.O. |
| EPI_ISL_15373792 | Department of immunology and microbiology - Pasteur Institute in Ho Chi Minh city | Department of immunology and microbiology - Pasteur Institute in Ho Chi Minh city | Manh H. Dao, Nhung H. P. Vu, Hang T. T. Pham, Thang M. Cao, Thinh V. Nguyen, Quang D. Pham, Quang C. Luong, Trung V. Nguyen |
| EPI_ISL_15380491 | Research Institute for Tropical Medicine | Molecular Biology Laboratory, Research Institute for Tropical Medicine | Czarina Christelle Alyannah S. Celis, Ezekiel A. Melo, Stephen Paul B.Ortia, Joanna Ina G. Manalo, Amalea Dulcene Nicolasora, Miguel Francisco B. Abulencia, Francisco Gerardo M. Polotan on behalf of the Research Institute for Tropical Medicine |
| EPI_ISL_15380492 | Eastwood Medical City | Molecular Biology Laboratory, Research Institute for Tropical Medicine | Samantha Louise P. Bado, Niquitta B. Galap, Bea C. Mateo, Chelsea Mae M. Reyes, Amalea Dulcene Nicolasora, Miguel Francisco B. Abulencia, Francisco Gerardo M. Polotan on behalf of the Research Institute for Tropical Medicine |
| EPI_ISL_15412318, EPI_ISL_15412320, EPI_ISL_15412322, EPI_ISL_15412324, EPI_ISL_15412326, EPI_ISL_15412328, EPI_ISL_15412330, EPI_ISL_15412331, EPI_ISL_15412333, EPI_ISL_15412335, EPI_ISL_15412336, EPI_ISL_15412338 | | | |
| see above | CT Department of Public Health | CT Department of Public Health | Claire Pearson, Tu N. Nguyen, Kutluhan Incekara, Neranjan V. Perera |
| EPI_ISL_15415308 | Hospital Clínica Bíblica | Inciensa, Instituto de Investigación y Enseñanza en Nutrición y Salud | Francisco Duarte, Ana Isela Ruiz-Gonzalez, Hillary Serrano, Diana Cantillo, Claudio Soto-Garita, Gustavo Vega, Estela Cordero, Adriana Godinez & Melany Calderon |
| EPI_ISL_15418009 | Los Angeles County Public Health Laboratories | Los Angeles County Public Health Laboratories | P. Hemarajata et al. |
| EPI_ISL_15419131 | Instituto de Infectologia Emilio Ribas | Instituto Adolfo Lutz Strategic Laboratory | Claudio Tavares Sacchi, Karoline Rodrigues Campos, Ariadne Ferreira Amarante, Marlon Benedito Nascimento Santos, Adriano Abbud, Adriana Bugno |
| EPI_ISL_15419132 | Centro de Saude de Sao Roque Dr Jose Carvalho Brito | Instituto Adolfo Lutz Strategic Laboratory | Claudio Tavares Sacchi, Karoline Rodrigues Campos, Ariadne Ferreira Amarante, Marlon Benedito Nascimento Santos, Adriano Abbud, Adriana Bugno |
| EPI_ISL_15419133 | UBS II COHAB Presidente Prudente | Instituto Adolfo Lutz Strategic Laboratory | Claudio Tavares Sacchi, Karoline Rodrigues Campos, Ariadne Ferreira Amarante, Marlon Benedito Nascimento Santos, Adriano Abbud, Adriana Bugno |
| EPI_ISL_15419134 | CTA Centro de Testagem e Aconselhamento de Caieiras | Instituto Adolfo Lutz Strategic Laboratory | Claudio Tavares Sacchi, Karoline Rodrigues Campos, Ariadne Ferreira Amarante, Marlon Benedito Nascimento Santos, Adriano Abbud, Adriana Bugno |
| EPI_ISL_15419135 | SAE DST AIDS Cidade Dutra | Instituto Adolfo Lutz Strategic Laboratory | Claudio Tavares Sacchi, Karoline Rodrigues Campos, Ariadne Ferreira Amarante, Marlon Benedito Nascimento Santos, Adriano Abbud, Adriana Bugno |
| EPI_ISL_15419136 | UBS J Nordeste | Instituto Adolfo Lutz Strategic Laboratory | Claudio Tavares Sacchi, Karoline Rodrigues Campos, Ariadne Ferreira Amarante, Marlon Benedito Nascimento Santos, Adriano Abbud, Adriana Bugno |
| EPI_ISL_15419137 | CTA Centro de Testagem e Aconselhamento Favo de Mel | Instituto Adolfo Lutz Strategic Laboratory | Claudio Tavares Sacchi, Karoline Rodrigues Campos, Ariadne Ferreira Amarante, Marlon Benedito Nascimento Santos, Adriano Abbud, Adriana Bugno |
| EPI_ISL_15419138 | Vigilancia Epidemiologica e Controle de Vetores de Pirassununga | Instituto Adolfo Lutz Strategic Laboratory | Claudio Tavares Sacchi, Karoline Rodrigues Campos, Ariadne Ferreira Amarante, Marlon Benedito Nascimento Santos, Adriano Abbud, Adriana Bugno |
| EPI_ISL_15419139 | Policlinica Maria Dirce | Instituto Adolfo Lutz Strategic Laboratory | Claudio Tavares Sacchi, Karoline Rodrigues Campos, Ariadne Ferreira Amarante, Marlon Benedito Nascimento Santos, Adriano Abbud, Adriana Bugno |

| | | | |
|---|---|---|---|
| EPI_ISL_15419140 | Secretaria Municipal de Saude de Batatais SP | Instituto Adolfo Lutz Strategic Laboratory | Claudio Tavares Sacchi, Karoline Rodrigues Campos, Ariadne Ferreira Amarante, Marlon Benedito Nascimento Santos, Adriano Abbud, Adriana Bugno |
| EPI_ISL_15419141 | UBS J Nordeste | Instituto Adolfo Lutz Strategic Laboratory | Claudio Tavares Sacchi, Karoline Rodrigues Campos, Ariadne Ferreira Amarante, Marlon Benedito Nascimento Santos, Adriano Abbud, Adriana Bugno |
| EPI_ISL_15419142 | Instituto de Infectologia Emilio Ribas | Instituto Adolfo Lutz Strategic Laboratory | Claudio Tavares Sacchi, Karoline Rodrigues Campos, Ariadne Ferreira Amarante, Marlon Benedito Nascimento Santos, Adriano Abbud, Adriana Bugno |
| EPI_ISL_15419143 | Santa Casa de Barretos | Instituto Adolfo Lutz Strategic Laboratory | Claudio Tavares Sacchi, Karoline Rodrigues Campos, Ariadne Ferreira Amarante, Marlon Benedito Nascimento Santos, Adriano Abbud, Adriana Bugno |
| EPI_ISL_15419144 | Hospital Vera Cruz | Instituto Adolfo Lutz Strategic Laboratory | Claudio Tavares Sacchi, Karoline Rodrigues Campos, Ariadne Ferreira Amarante, Marlon Benedito Nascimento Santos, Adriano Abbud, Adriana Bugno |
| EPI_ISL_15419145 | NotreDame Intermedica Saude | Instituto Adolfo Lutz Strategic Laboratory | Claudio Tavares Sacchi, Karoline Rodrigues Campos, Ariadne Ferreira Amarante, Marlon Benedito Nascimento Santos, Adriano Abbud, Adriana Bugno |
| EPI_ISL_15419146 | Hospital e Maternidade Santa Maria Cruz Azul | Instituto Adolfo Lutz Strategic Laboratory | Claudio Tavares Sacchi, Karoline Rodrigues Campos, Ariadne Ferreira Amarante, Marlon Benedito Nascimento Santos, Adriano Abbud, Adriana Bugno |
| EPI_ISL_15419147 | Pronto Socorro Central de Diadema | Instituto Adolfo Lutz Strategic Laboratory | Claudio Tavares Sacchi, Karoline Rodrigues Campos, Ariadne Ferreira Amarante, Marlon Benedito Nascimento Santos, Adriano Abbud, Adriana Bugno |
| EPI_ISL_15419148 | NotreDame Intermedica Saude Santo Andre | Instituto Adolfo Lutz Strategic Laboratory | Claudio Tavares Sacchi, Karoline Rodrigues Campos, Ariadne Ferreira Amarante, Marlon Benedito Nascimento Santos, Adriano Abbud, Adriana Bugno |
| EPI_ISL_15419149 | UBS Dom Angelico | Instituto Adolfo Lutz Strategic Laboratory | Claudio Tavares Sacchi, Karoline Rodrigues Campos, Ariadne Ferreira Amarante, Marlon Benedito Nascimento Santos, Adriano Abbud, Adriana Bugno |
| EPI_ISL_15419150 | Secao de Centro de Diagnostico SECEDI | Instituto Adolfo Lutz Strategic Laboratory | Claudio Tavares Sacchi, Karoline Rodrigues Campos, Ariadne Ferreira Amarante, Marlon Benedito Nascimento Santos, Adriano Abbud, Adriana Bugno |
| EPI_ISL_15419151 | UPA 24H Brotas | Instituto Adolfo Lutz Strategic Laboratory | Claudio Tavares Sacchi, Karoline Rodrigues Campos, Ariadne Ferreira Amarante, Marlon Benedito Nascimento Santos, Adriano Abbud, Adriana Bugno |
| EPI_ISL_15419152 | Hospital Nossa Senhora de Lourdes | Instituto Adolfo Lutz Strategic Laboratory | Claudio Tavares Sacchi, Karoline Rodrigues Campos, Ariadne Ferreira Amarante, Marlon Benedito Nascimento Santos, Adriano Abbud, Adriana Bugno |
| EPI_ISL_15419153 | AMA Paraisopolis | Instituto Adolfo Lutz Strategic Laboratory | Claudio Tavares Sacchi, Karoline Rodrigues Campos, Ariadne Ferreira Amarante, Marlon Benedito Nascimento Santos, Adriano Abbud, Adriana Bugno |
| EPI_ISL_15419154 | Pronto Atendimento Infantil e Central de Quimioterapia de Sao Jose do Rio Preto | Instituto Adolfo Lutz Strategic Laboratory | Claudio Tavares Sacchi, Karoline Rodrigues Campos, Ariadne Ferreira Amarante, Marlon Benedito Nascimento Santos, Adriano Abbud, Adriana Bugno |
| EPI_ISL_15419155 | Santa Casa de Atibaia Pro Saude | Instituto Adolfo Lutz Strategic Laboratory | Claudio Tavares Sacchi, Karoline Rodrigues Campos, Ariadne Ferreira Amarante, Marlon Benedito Nascimento Santos, Adriano Abbud, Adriana Bugno |
| EPI_ISL_15419156 | UPA Vila Mariana | Instituto Adolfo Lutz Strategic Laboratory | Claudio Tavares Sacchi, Karoline Rodrigues Campos, Ariadne Ferreira Amarante, Marlon Benedito Nascimento Santos, Adriano Abbud, Adriana Bugno |
| EPI_ISL_15419157 | Secretaria de Saude de Mogi das Cruzes | Instituto Adolfo Lutz Strategic Laboratory | Claudio Tavares Sacchi, Karoline Rodrigues Campos, Ariadne Ferreira Amarante, Marlon Benedito Nascimento Santos, Adriano Abbud, Adriana Bugno |
| EPI_ISL_15419158 | Unidade Mista de saude Mariano Gayoso Castelo Branco | Instituto Adolfo Lutz Strategic Laboratory | Claudio Tavares Sacchi, Karoline Rodrigues Campos, Ariadne Ferreira Amarante, Marlon Benedito Nascimento Santos, Adriano Abbud, Adriana Bugno |
| EPI_ISL_15419159 | Hospital Vivalle | Instituto Adolfo Lutz Strategic Laboratory | Claudio Tavares Sacchi, Karoline Rodrigues Campos, Ariadne Ferreira Amarante, Marlon Benedito Nascimento Santos, Adriano Abbud, Adriana Bugno |
| EPI_ISL_15419160 | UPA Nova Hortolandia Manoel Geogino Lopes | Instituto Adolfo Lutz Strategic Laboratory | Claudio Tavares Sacchi, Karoline Rodrigues Campos, Ariadne Ferreira Amarante, Marlon Benedito Nascimento Santos, Adriano Abbud, Adriana Bugno |
| EPI_ISL_15419161 | Centro de Saude I Albertino Affonso Jaboticabal | Instituto Adolfo Lutz Strategic Laboratory | Claudio Tavares Sacchi, Karoline Rodrigues Campos, Ariadne Ferreira Amarante, Marlon Benedito Nascimento Santos, Adriano Abbud, Adriana Bugno |
| EPI_ISL_15419162 | SAE DST AIDS M Boi Mirim Servico de Atencao Especializada | Instituto Adolfo Lutz Strategic Laboratory | Claudio Tavares Sacchi, Karoline Rodrigues Campos, Ariadne Ferreira Amarante, Marlon Benedito Nascimento Santos, Adriano Abbud, Adriana Bugno |
| EPI_ISL_15419163 | Instituto de Infectologia Emilio Ribas | Instituto Adolfo Lutz Strategic Laboratory | Claudio Tavares Sacchi, Karoline Rodrigues Campos, Ariadne Ferreira Amarante, Marlon Benedito Nascimento Santos, Adriano Abbud, Adriana Bugno |
| EPI_ISL_15458905 | Nebraska Public Health Laboratory (NPHL) | Environmental, Agricultural, and Occupational Health, University of Nebraska Medical Center | Chapman,R.C., Bernhard,K., McCutchen,E.L., Fauver,J.R., O'Dell,J.X., Mannell,M., Wiley,M.R. and Cross,S.T. |
| EPI_ISL_15502328 | Los Angeles County Public Health Laboratories | Los Angeles County Public Health Laboratories | P. Hemarajata et al. |
| EPI_ISL_15593715 | División Diagnóstico Molecular Hospital México | División Diagnóstico Molecular Hospital México | Juan Carlos Villalobos Ugalde, Vanessa Villalobos Alfaro, Carlos Ramirez Chavarria |
| EPI_ISL_15593718 | LESP Mexico City | Instituto de Diagnostico y Referencia Epidemiologicos (INDRE) | Abril Rodríguez-Maldonado; Claudia Wong-Arámbula; Felipe Arguijo-Perez; Helios Cárdenas-Hernández; Carmen Castro-Méndez; Lidia García-Torres; Ruth Madera-Sandoval; América Mandujano-Martínez; Nancy Martínez-Velázquez; Mireya Mederos-Michel; Angélica Pedraza-Meléndez; Joaquín Quiroz-Mercado; Daniel Regalado-Santiago; Silvia Rivero-Arredondo; Erika Sierra-Atanacio; Fernando González-Domínguez; Lucia Hernández-Rivas, Irma López-Martínez; Ernesto Ramírez-González; Maribel González-Villa |
| EPI_ISL_15593719 | LESP Puebla | Instituto de Diagnostico y Referencia Epidemiologicos (INDRE) | Abril Rodríguez-Maldonado; Claudia Wong-Arámbula; Felipe Arguijo-Perez; Helios Cárdenas-Hernández; Carmen Castro-Méndez; Lidia García-Torres; Ruth Madera-Sandoval; América Mandujano-Martínez; Nancy Martínez-Velázquez; Mireya Mederos-Michel; Angélica Pedraza-Meléndez; Joaquín Quiroz-Mercado; Daniel Regalado-Santiago; Silvia Rivero-Arredondo; Erika Sierra-Atanacio; Fernando González-Domínguez; Lucia Hernández-Rivas, Irma López-Martínez; Ernesto Ramírez-González; Maribel González-Villa |
| EPI_ISL_15593720 | LESP Tamaulipas | Instituto de Diagnostico y Referencia Epidemiologicos (INDRE) | Abril Rodríguez-Maldonado; Claudia Wong-Arámbula; Felipe Arguijo-Perez; Helios Cárdenas-Hernández; Carmen Castro-Méndez; Lidia García-Torres; Ruth Madera-Sandoval; América Mandujano-Martínez; Nancy Martínez-Velázquez; Mireya Mederos-Michel; Angélica Pedraza-Meléndez; Joaquín Quiroz-Mercado; Daniel Regalado-Santiago; Silvia Rivero-Arredondo; Erika Sierra-Atanacio; Fernando González-Domínguez; Lucia Hernández-Rivas, Irma López-Martínez; Ernesto Ramírez-González; Maribel González-Villa |
| EPI_ISL_15593721 | LESP Baja California | Instituto de Diagnostico y Referencia Epidemiologicos (INDRE) | Abril Rodríguez-Maldonado; Claudia Wong-Arámbula; Felipe Arguijo-Perez; Helios Cárdenas-Hernández; Carmen Castro-Méndez; Lidia García-Torres; Ruth Madera-Sandoval; América Mandujano-Martínez; Nancy Martínez-Velázquez; Mireya Mederos-Michel; Angélica Pedraza-Meléndez; Joaquín Quiroz-Mercado; Daniel Regalado-Santiago; Silvia Rivero-Arredondo; Erika Sierra-Atanacio; Fernando González-Domínguez; Lucia Hernández-Rivas, Irma López-Martínez; Ernesto Ramírez-González; Maribel González-Villa |
| EPI_ISL_15597042, EPI_ISL_15597046 | UCLA Clinical Micro Lab | Los Angeles County Public Health Laboratories | P. Hemarajata et al. |
| EPI_ISL_15597048, EPI_ISL_15597049 | Quest Diagnostics Nichols Institute | Los Angeles County Public Health Laboratories | P. Hemarajata et al. |
| EPI_ISL_15597058, EPI_ISL_15597059, EPI_ISL_15597063 | Los Angeles County Public Health Laboratories | Los Angeles County Public Health Laboratories | P. Hemarajata et al. |
| EPI_ISL_15608908, EPI_ISL_15608909, EPI_ISL_15608910 | Southern Nevada Public Health Laboratory | Southern Nevada Public Health Laboratory | Michael Picker |
| EPI_ISL_15641541, EPI_ISL_15641542, EPI_ISL_15641543, EPI_ISL_15641544, EPI_ISL_15641545, EPI_ISL_15641546, EPI_ISL_15641547, EPI_ISL_15641548, EPI_ISL_15641549, EPI_ISL_15641550, EPI_ISL_15641551, EPI_ISL_15641552, EPI_ISL_15641553, EPI_ISL_15641554, EPI_ISL_15641555, EPI_ISL_15641556, EPI_ISL_15641557, EPI_ISL_15641558, EPI_ISL_15641559, EPI_ISL_15641560, EPI_ISL_15641561, EPI_ISL_15641562, EPI_ISL_15641563, EPI_ISL_15641564, EPI_ISL_15641565, EPI_ISL_15641566, EPI_ISL_15641567, EPI_ISL_15641568, EPI_ISL_15641569, EPI_ISL_15641570, EPI_ISL_15641571, EPI_ISL_15641572, EPI_ISL_15641573, EPI_ISL_15641574, EPI_ISL_15641575, EPI_ISL_15641576, EPI_ISL_15641577, EPI_ISL_15641578, EPI_ISL_15641579, EPI_ISL_15641580, EPI_ISL_15641581, EPI_ISL_15641582, EPI_ISL_15641583, EPI_ISL_15641584, EPI_ISL_15641585, EPI_ISL_15641586, EPI_ISL_15641587, EPI_ISL_15641588, EPI_ISL_15641589, EPI_ISL_15641590, EPI_ISL_15641591, EPI_ISL_15641592, EPI_ISL_15641593, EPI_ISL_15641594, EPI_ISL_15641595, EPI_ISL_15641596, EPI_ISL_15641597, EPI_ISL_15641598, EPI_ISL_15641599, EPI_ISL_15641600, EPI_ISL_15641601, EPI_ISL_15641602, EPI_ISL_15641603, EPI_ISL_15641604, EPI_ISL_15641605, EPI_ISL_15641606, EPI_ISL_15641607, EPI_ISL_15641608, EPI_ISL_15641609, EPI_ISL_15641610 | | | |
| see above | Public Health Laboratory, Public Health Service Amsterdam, The Netherlands | Department of Medical Microbiology & Infection prevention, Amsterdam University Medical Centers location AMC | Matthijs Welkers, Jelle Koopsen, Robin van Houdt, Marcel Jonges, Sebastien Matamoros, Sjoerd Rebers, Fokla Zorgdrager, Sylvia Bruisten, Akke Cornelissen, Janke Schinkel, Ewout Fanoy, Roisin Bavalia, Menno de Jong and Mariken van der Lubben on behalf of the Amsterdam Regional Genomic epidemiology and Outbreak Surveillance (ARGOS) consortium |
| EPI_ISL_15655944 | Erasmus Medical Center Department of Virology | Erasmus Medical Center Department of Virology | Leonard Schuele, Bas Oude Munnink, Marjan Boter, Babette Weller, Babs Verstrepen, Richard Molenkamp, Janette Rahamat-Langendoen, Reina Sikkema, Marion Koopmans |
| EPI_ISL_15660293 | Johns Hopkins University School of Medicine | Johns Hopkins School of Medicine Pathology | Amary Fall, Heba M Mosta |
| EPI_ISL_15660294, EPI_ISL_15660895, EPI_ISL_15660926 | Johns Hopkins University School of Medicine | Johns Hopkins School of Medicine Pathology | Amary Fall,Katharine Uhteg, C Paul Morris ,Heba H. Mostafa |
| EPI_ISL_15701569 | Institute for Medical Virology, University Hospital, Goethe University Frankfurt, Germany | Institute of Virology, Charité-Universitätsmedizin Berlin, Humboldt-Universität zu Berlin, Germany | Denisa Bojkova, Julia Schneider, Victor M. Corman, Jindrich Cinatl jr. |
| EPI_ISL_15702015, EPI_ISL_15702711, EPI_ISL_15703560 | Erasmus Medical Center Department of Virology | Erasmus Medical Center Department of Virology | Leonard Schuele, Bas Oude Munnink, Marjan Boter, Babette Weller, Babs Verstrepen, Richard Molenkamp, Janette Rahamat-Langendoen, Reina Sikkema, Marion Koopmans |
| EPI_ISL_15704688 | Institute for Medical Virology, University Hospital, Goethe University Frankfurt, Germany | Institute of Virology, Charité-Universitätsmedizin Berlin, Humboldt-Universität zu Berlin, Germany | Denisa Bojkova, Julia Schneider, Victor M. Corman, Jindrich Cinatl jr. |
| EPI_ISL_15704845 | Erasmus Medical Center Department of Virology | Erasmus Medical Center Department of Virology | Leonard Schuele, Bas Oude Munnink, Marjan Boter, Babette Weller, Babs Verstrepen, Richard Molenkamp, Janette Rahamat-Langendoen, Reina Sikkema, Marion Koopmans |
| EPI_ISL_15704861, EPI_ISL_15705166, EPI_ISL_15705315, EPI_ISL_15705498, EPI_ISL_15705640, EPI_ISL_15705771, EPI_ISL_15705945, EPI_ISL_15706105, EPI_ISL_15706238, EPI_ISL_15706380 | Institute for Medical Virology, University Hospital, Goethe University Frankfurt, Germany | Institute of Virology, Charité-Universitätsmedizin Berlin, Humboldt-Universität zu Berlin, Germany | Denisa Bojkova, Julia Schneider, Victor M. Corman, Jindrich Cinatl jr. |
| EPI_ISL_15709387 | División de Microbiología, Hospital Nacional de Niños Dr. Carlos Sáenz Herrera | Inciensa, Instituto de Investigación y Enseñanza en Nutrición y Salud | Francisco Duarte, Ana Isela Ruiz-Gonzalez, Hillary Serrano, Diana Cantillo, Claudio Soto-Garita, Gustavo Vega, Estela Cordero, Adriana Godinez, Melany Calderon & Cristian Pérez-Corrales |
| EPI_ISL_15712853, EPI_ISL_15712860, EPI_ISL_15714234, EPI_ISL_15714244, EPI_ISL_15714245, EPI_ISL_15714286, EPI_ISL_15714287, EPI_ISL_15714288, EPI_ISL_15714299, EPI_ISL_15714539, EPI_ISL_15714920, EPI_ISL_15714989, EPI_ISL_15714990, EPI_ISL_15715289, EPI_ISL_15715292, EPI_ISL_15715305, EPI_ISL_15717862, EPI_ISL_15718514, EPI_ISL_15718920 | | | |
| see above | Erasmus Medical Center Department of Virology | Erasmus Medical Center Department of Virology | Leonard Schuele, Bas Oude Munnink, Marjan Boter, Babette Weller, Babs Verstrepen, Richard Molenkamp, Janette Rahamat-Langendoen, Reina Sikkema, Marion Koopmans |
| EPI_ISL_15747900 | WHO National Influenza Centre Russian Federation | WHO National Influenza Centre Russian Federation | Andrey Komissarov, Artem Fadeev, Nikita Yolshin, Evgeny Venev, Kseniya Komissarova, Daria Danilenko, Dmitry Lioznov |
| EPI_ISL_15763810, EPI_ISL_15763811, EPI_ISL_15763812, EPI_ISL_15763813, EPI_ISL_15763814, EPI_ISL_15763815, EPI_ISL_15763816, EPI_ISL_15763817, EPI_ISL_15763818, EPI_ISL_15763819, EPI_ISL_15763820, EPI_ISL_15763821, EPI_ISL_15763822, EPI_ISL_15763823, EPI_ISL_15763824, EPI_ISL_15763825, EPI_ISL_15763826, EPI_ISL_15763827, EPI_ISL_15763828, EPI_ISL_15763829, EPI_ISL_15763830, EPI_ISL_15763831, EPI_ISL_15763832 | | | |
| see above | National Virus Reference Laboratory | National Virus Reference Laboratory | Gabriel Gonzalez, Michael Carr, Brian Keogan, Jose Maria Urtasun Elizari, Jonathan Dean, Daniel Hare, Cillian F De Gascun |
| EPI_ISL_15831211 | División Diagnóstico Molecular Hospital México | División Diagnóstico Molecular Hospital México | Juan Carlos Villalobos Ugalde, Vanessa Villalobos Alfaro, Carlos Ramirez Chavarría |
| EPI_ISL_15831212 | División Diagnóstico Molecular Hospital México | División Diagnóstico Molecular Hospital México | Juan Carlos Villalobos Ugalde, Vanessa Villalobos Alfaro, Sofía Villalobos Abarca |
| EPI_ISL_15896302 | Los Angeles County Public Health Laboratories | Los Angeles County Public Health Laboratories | P. Hemarajata et al. |
| EPI_ISL_15896309 | Kaiser Permanente Chino Hills Regional Reference Laboratories | Los Angeles County Public Health Laboratories | P. Hemarajata et al. |
| EPI_ISL_15896332, EPI_ISL_15896345 | Los Angeles County Public Health Laboratories | Los Angeles County Public Health Laboratories | P. Hemarajata et al. |

| | | | |
|---|---|---|---|
| EPI_ISL_15896817, EPI_ISL_15896818, EPI_ISL_15897122, EPI_ISL_15897123, EPI_ISL_15897124, EPI_ISL_15897493, EPI_ISL_15897494, EPI_ISL_15897569, EPI_ISL_15897570, EPI_ISL_15897573, EPI_ISL_15897574, EPI_ISL_15897587, EPI_ISL_15897588, EPI_ISL_15897589, EPI_ISL_15897590, EPI_ISL_15897591 | | | |
| see above | Landspitali Department of Clinical Microbiology | Landspitali Department of Clinical Microbiology | Freyja Valsdottir, Zarko Urosevic, Brynja Armannsdottir, Karl Gustaf Kristinsson |
| EPI_ISL_15942296 | Sexually Transmitted Diseases (STDs) outpatient service of Dermatology Unit, Fondazione IRCCS Ca' Granda Ospedale Maggiore Policlinico of Milan | Bioinformatic lab, Scientific Institute IRCCS E. Medea | Diego Forni, Rachele Cagliani, Manuela Sironi, Chiara Moltrasio, Luigia Venegoni, Eleonora Quattri, Angelo Marzano |
| EPI_ISL_15942637, EPI_ISL_15942638, EPI_ISL_15942885, EPI_ISL_15942886 | Sexually Transmitted Diseases (STDs) outpatient service of Dermatology Unit, Fondazione IRCCS Ca' Granda Ospedale Maggiore Policlinico of Milan | Bioinformatic Lab, Scientific Institute IRCCS E. Medea | Diego Forni, Rachele Cagliani, Manuela Sironi, Chiara Moltrasio, Luigia Venegoni, Eleonora Quattri, Angelo Marzano |
| EPI_ISL_15950021, EPI_ISL_15950022, EPI_ISL_15950023, EPI_ISL_15950024, EPI_ISL_15950025, EPI_ISL_15950026, EPI_ISL_15950027, EPI_ISL_15950028, EPI_ISL_15950029, EPI_ISL_15950030, EPI_ISL_15950031, EPI_ISL_15950032 | | | |
| see above | Rhode Island State Health Laboratory | Rhode Island State Health Laboratory | Kristin Carpenter-Azevedo, Sean Sierra-Patev, Richard C. Huard |
| EPI_ISL_15955334, EPI_ISL_15955345, EPI_ISL_15955346, EPI_ISL_15955347, EPI_ISL_15955354 | Kaiser Permanente Chino Hills Regional Reference Laboratories | Los Angeles County Public Health Laboratories | P. Hemarajata et al. |
| EPI_ISL_15965179 | Institute of Microbiology, Universidad San Francisco de Quito | Institute of Microbiology, Universidad San Francisco de Quito | Belén Prado-Vivar, Mateo Carvajal, Sully Márquez, Erika B. Muñoz, Rommel Guevara, Maritza Paez, Estefanía Rivadeneira, Evelyn Sánchez Espinoza, Josefina Coloma, Verónica Barragán, Patricio Rojas-Silva, Gabriel Trueba, Michelle Grunauer, Paúl Cárdenas |
| EPI_ISL_15972402, EPI_ISL_15972403, EPI_ISL_15972404, EPI_ISL_15972405, EPI_ISL_15972406, EPI_ISL_15972407, EPI_ISL_15972408, EPI_ISL_15972409 | Laboratorio Central, Ministerio de Salud Córdoba | Laboratorio Central, Ministerio de Salud Córdoba | Castro, G.; Sicilia, P.; Poklepovich, T.; Campos, J.; Barbas, G. |
| EPI_ISL_15992095 | LESP State of Mexico | Instituto de Diagnostico y Referencia Epidemiologicos (INDRE) | Abril Rodríguez-Maldonado; Claudia Wong-Arámbula; Felipe Arguijo-Perez; Helios Cárdenas-Hernández; Carmen Castro-Méndez; Lidia García-Torres; Ruth Madera-Sandoval; América Mandujano-Martínez; Nancy Martínez-Velázquez; Mireya Mederos-Michel; Angélica Pedraza-Meléndez; Joaquín Quiroz-Mercado; Daniel Regalado-Santiago; Silvia Rivero-Arredondo; Erika Sierra-Atanacio; Fernando González-Domínguez; Lucia Hernández-Rivas, Irma López-Martínez; Ernesto Ramírez-González; Maribel González-Villa |
| EPI_ISL_15992096 | LESP Jalisco | Instituto de Diagnostico y Referencia Epidemiologicos (INDRE) | Abril Rodríguez-Maldonado; Claudia Wong-Arámbula; Felipe Arguijo-Perez; Helios Cárdenas-Hernández; Carmen Castro-Méndez; Lidia García-Torres; Ruth Madera-Sandoval; América Mandujano-Martínez; Nancy Martínez-Velázquez; Mireya Mederos-Michel; Angélica Pedraza-Meléndez; Joaquín Quiroz-Mercado; Daniel Regalado-Santiago; Silvia Rivero-Arredondo; Erika Sierra-Atanacio; Fernando González-Domínguez; Lucia Hernández-Rivas, Irma López-Martínez; Ernesto Ramírez-González; Maribel González-Villa |
| EPI_ISL_15992097 | LESP Morelos | Instituto de Diagnostico y Referencia Epidemiologicos (INDRE) | Abril Rodríguez-Maldonado; Claudia Wong-Arámbula; Felipe Arguijo-Perez; Helios Cárdenas-Hernández; Carmen Castro-Méndez; Lidia García-Torres; Ruth Madera-Sandoval; América Mandujano-Martínez; Nancy Martínez-Velázquez; Mireya Mederos-Michel; Angélica Pedraza-Meléndez; Joaquín Quiroz-Mercado; Daniel Regalado-Santiago; Silvia Rivero-Arredondo; Erika Sierra-Atanacio; Fernando González-Domínguez; Lucia Hernández-Rivas, Irma López-Martínez; Ernesto Ramírez-González; Maribel González-Villa |
| EPI_ISL_15992098 | LESP Nuevo Leon | Instituto de Diagnostico y Referencia Epidemiologicos (INDRE) | Abril Rodríguez-Maldonado; Claudia Wong-Arámbula; Felipe Arguijo-Perez; Helios Cárdenas-Hernández; Carmen Castro-Méndez; Lidia García-Torres; Ruth Madera-Sandoval; América Mandujano-Martínez; Nancy Martínez-Velázquez; Mireya Mederos-Michel; Angélica Pedraza-Meléndez; Joaquín Quiroz-Mercado; Daniel Regalado-Santiago; Silvia Rivero-Arredondo; Erika Sierra-Atanacio; Fernando González-Domínguez; Lucia Hernández-Rivas, Irma López-Martínez; Ernesto Ramírez-González; Maribel González-Villa |
| EPI_ISL_15992099 | LESP Hidalgo | Instituto de Diagnostico y Referencia Epidemiologicos (INDRE) | Abril Rodríguez-Maldonado; Claudia Wong-Arámbula; Felipe Arguijo-Perez; Helios Cárdenas-Hernández; Carmen Castro-Méndez; Lidia García-Torres; Ruth Madera-Sandoval; América Mandujano-Martínez; Nancy Martínez-Velázquez; Mireya Mederos-Michel; Angélica Pedraza-Meléndez; Joaquín Quiroz-Mercado; Daniel Regalado-Santiago; Silvia Rivero-Arredondo; Erika Sierra-Atanacio; Fernando González-Domínguez; Lucia Hernández-Rivas, Irma López-Martínez; Ernesto Ramírez-González; Maribel González-Villa |
| EPI_ISL_15992100 | LESP Campeche | Instituto de Diagnostico y Referencia Epidemiologicos (INDRE) | Abril Rodríguez-Maldonado; Claudia Wong-Arámbula; Felipe Arguijo-Perez; Helios Cárdenas-Hernández; Carmen Castro-Méndez; Lidia García-Torres; Ruth Madera-Sandoval; América Mandujano-Martínez; Nancy Martínez-Velázquez; Mireya Mederos-Michel; Angélica Pedraza-Meléndez; Joaquín Quiroz-Mercado; Daniel Regalado-Santiago; Silvia Rivero-Arredondo; Erika Sierra-Atanacio; Fernando González-Domínguez; Lucia Hernández-Rivas, Irma López-Martínez; Ernesto Ramírez-González; Maribel González-Villa |
| EPI_ISL_15992101 | LESP Tlaxcala | Instituto de Diagnostico y Referencia Epidemiologicos (INDRE) | Abril Rodríguez-Maldonado; Claudia Wong-Arámbula; Felipe Arguijo-Perez; Helios Cárdenas-Hernández; Carmen Castro-Méndez; Lidia García-Torres; Ruth Madera-Sandoval; América Mandujano-Martínez; Nancy Martínez-Velázquez; Mireya Mederos-Michel; Angélica Pedraza-Meléndez; Joaquín Quiroz-Mercado; Daniel Regalado-Santiago; Silvia Rivero-Arredondo; Erika Sierra-Atanacio; Fernando González-Domínguez; Lucia Hernández-Rivas, Irma López-Martínez; Ernesto Ramírez-González; Maribel González-Villa |
| EPI_ISL_15992102 | LESP Aguascalientes | Instituto de Diagnostico y Referencia Epidemiologicos (INDRE) | Abril Rodríguez-Maldonado; Claudia Wong-Arámbula; Felipe Arguijo-Perez; Helios Cárdenas-Hernández; Carmen Castro-Méndez; Lidia García-Torres; Ruth Madera-Sandoval; América Mandujano-Martínez; Nancy Martínez-Velázquez; Mireya Mederos-Michel; Angélica Pedraza-Meléndez; Joaquín Quiroz-Mercado; Daniel Regalado-Santiago; Silvia Rivero-Arredondo; Erika Sierra-Atanacio; Fernando González-Domínguez; Lucia Hernández-Rivas, Irma López-Martínez; Ernesto Ramírez-González; Maribel González-Villa |
| EPI_ISL_15992103, EPI_ISL_15992104 | LESP San Luis Potosi | Instituto de Diagnostico y Referencia Epidemiologicos (INDRE) | Abril Rodríguez-Maldonado; Claudia Wong-Arámbula; Felipe Arguijo-Perez; Helios Cárdenas-Hernández; Carmen Castro-Méndez; Lidia García-Torres; Ruth Madera-Sandoval; América Mandujano-Martínez; Nancy Martínez-Velázquez; Mireya Mederos-Michel; Angélica Pedraza-Meléndez; Joaquín Quiroz-Mercado; Daniel Regalado-Santiago; Silvia Rivero-Arredondo; Erika Sierra-Atanacio; Fernando González-Domínguez; Lucia Hernández-Rivas, Irma López-Martínez; Ernesto Ramírez-González; Maribel González-Villa |
| EPI_ISL_16006560 | Sequencing/Bioinformatics, Delaware Public Health Lab | Sequencing/Bioinformatics, Delaware Public Health Lab | Bajwa,M.I. and Miller,H. |
| EPI_ISL_16012468, EPI_ISL_16012469, EPI_ISL_16012470, EPI_ISL_16012471, EPI_ISL_16012472, EPI_ISL_16012473, EPI_ISL_16012474, EPI_ISL_16012475, EPI_ISL_16012476, EPI_ISL_16012477, EPI_ISL_16012478, EPI_ISL_16012479, EPI_ISL_16012480, EPI_ISL_16012481, EPI_ISL_16012482, EPI_ISL_16012483, EPI_ISL_16012484, EPI_ISL_16012485, EPI_ISL_16012486, EPI_ISL_16012487, EPI_ISL_16012488, EPI_ISL_16012489, EPI_ISL_16012490, EPI_ISL_16012491, EPI_ISL_16012492, EPI_ISL_16012493, EPI_ISL_16012494, EPI_ISL_16012495, EPI_ISL_16012496, EPI_ISL_16012497, EPI_ISL_16012498, EPI_ISL_16012499, EPI_ISL_16012500, EPI_ISL_16012501, EPI_ISL_16012502, EPI_ISL_16012503, EPI_ISL_16012504, EPI_ISL_16012505, EPI_ISL_16012506, EPI_ISL_16012507, EPI_ISL_16012508, EPI_ISL_16012509, EPI_ISL_16012510, EPI_ISL_16012511, EPI_ISL_16012512, EPI_ISL_16012513, EPI_ISL_16012514, EPI_ISL_16012515, EPI_ISL_16012516, EPI_ISL_16012517, EPI_ISL_16012518, EPI_ISL_16012519, EPI_ISL_16012520, EPI_ISL_16012521, EPI_ISL_16012522, EPI_ISL_16012523, EPI_ISL_16012524, EPI_ISL_16012525, EPI_ISL_16012526, EPI_ISL_16012527, EPI_ISL_16012528, EPI_ISL_16012529, EPI_ISL_16012530, EPI_ISL_16012531, EPI_ISL_16012532, EPI_ISL_16012533, EPI_ISL_16012534, EPI_ISL_16012535, EPI_ISL_16012536 | | | |
| see above | National Virus Reference Laboratory | National Virus Reference Laboratory | Gabriel Gonzalez, Michael Carr, Brian Keogan, Jose Maria Urtasun Elizari, Jonathan Dean, Daniel Hare, Cillian F De Gascun |
| EPI_ISL_16074741 | Virology Lab, Institute of Tropical Medicine of Sao Paulo, School of Medicine, Universidade de Sao Paulo | Virology Lab | Antonio Charlys da Costa, Maria Cassia Mendes-Correa |
| EPI_ISL_16074742 | Virology Lab, Institute of Tropical Medicine of Sao Paulo, School of Medicine, Universidade de Sao Paulo | Virology Lab, Institute of Tropical Medicine of Sao Paulo, School of Medicine, Universidade de Sao Paulo | Antonio Charlys da Costa, Maria Cassia Mendes-Correa |
| EPI_ISL_16080585, EPI_ISL_16080586, EPI_ISL_16080587, EPI_ISL_16080588, EPI_ISL_16080589, EPI_ISL_16080590 | Institute of Virology, Faculty of Medicine and University Hospital Cologne, University of Cologne, Cologne, Germany | Institute of Virology, Faculty of Medicine and University Hospital Cologne, University of Cologne, Cologne, Germany | Eva Heger, Michael Böhm, Ulrike Wieland, Alexander Kreuter |
| EPI_ISL_16104842, EPI_ISL_16104843, EPI_ISL_16104844, EPI_ISL_16104845, EPI_ISL_16104846, EPI_ISL_16104847, EPI_ISL_16104848, EPI_ISL_16104849, EPI_ISL_16104850, EPI_ISL_16104851, EPI_ISL_16104852, EPI_ISL_16104853, EPI_ISL_16104854, EPI_ISL_16104855, EPI_ISL_16104856, EPI_ISL_16104857, EPI_ISL_16104858, EPI_ISL_16104859, EPI_ISL_16104860, EPI_ISL_16104861, EPI_ISL_16104862, EPI_ISL_16104863, EPI_ISL_16104864, EPI_ISL_16104865, EPI_ISL_16104866, EPI_ISL_16104867, EPI_ISL_16104868, EPI_ISL_16104869, EPI_ISL_16104870, EPI_ISL_16104871, EPI_ISL_16104872, EPI_ISL_16104873, EPI_ISL_16104874, EPI_ISL_16104875, EPI_ISL_16104876, EPI_ISL_16104877, EPI_ISL_16104878, EPI_ISL_16104879, EPI_ISL_16104880, EPI_ISL_16104881, EPI_ISL_16104882, EPI_ISL_16104883, EPI_ISL_16104884, EPI_ISL_16104885, EPI_ISL_16104886, EPI_ISL_16104887, EPI_ISL_16104888, EPI_ISL_16104889, EPI_ISL_16104890, EPI_ISL_16104891, EPI_ISL_16104892, EPI_ISL_16104893, EPI_ISL_16104894, EPI_ISL_16104895, EPI_ISL_16104896, EPI_ISL_16104897, EPI_ISL_16104898, EPI_ISL_16104899, EPI_ISL_16104900, EPI_ISL_16104901, EPI_ISL_16104902, EPI_ISL_16104903, EPI_ISL_16104904, EPI_ISL_16104905, EPI_ISL_16104906, EPI_ISL_16104907, EPI_ISL_16104908, EPI_ISL_16104909, EPI_ISL_16104910, EPI_ISL_16104911, EPI_ISL_16104912, EPI_ISL_16104913, EPI_ISL_16104914, EPI_ISL_16104915, EPI_ISL_16104916, EPI_ISL_16104917, EPI_ISL_16104918, EPI_ISL_16104919, EPI_ISL_16104920, EPI_ISL_16104921, EPI_ISL_16104922, EPI_ISL_16116727, EPI_ISL_16116728, EPI_ISL_16116729, EPI_ISL_16116730, EPI_ISL_16116733, EPI_ISL_16116734, EPI_ISL_16116735, EPI_ISL_16116736, EPI_ISL_16116737, EPI_ISL_16116738, EPI_ISL_16116739, EPI_ISL_16116740, EPI_ISL_16116741, EPI_ISL_16116742, EPI_ISL_16116743, EPI_ISL_16116744, EPI_ISL_16116745, EPI_ISL_16116746, EPI_ISL_16116747, EPI_ISL_16116748, EPI_ISL_16116749, EPI_ISL_16116750, EPI_ISL_16116751, EPI_ISL_16116752, EPI_ISL_16116753, EPI_ISL_16116754, EPI_ISL_16116755, EPI_ISL_16116756, EPI_ISL_16116757, EPI_ISL_16116758, EPI_ISL_16116759, EPI_ISL_16116760, EPI_ISL_16116761, EPI_ISL_16116762, EPI_ISL_16116763, EPI_ISL_16116764, EPI_ISL_16116765 | | | |
| see above | Laboratorio de Referencia Nacional de Viruas Inmunoprevenibles. Centro Nacional de Salud Publica. Instiuto Nacional de Salud | Laboratorio de Referencia Nacional de Viruas Inmunoprevenibles. Centro Nacional de Salud Publica. Instiuto Nacional de Salud | Carlos Patricio Padilla Rojas, Carmen Verónica Hurtado Vela, Juana Iris Silva Molina, Luis Bárcena Flores, Víctor Jiménez Vásquez, Alicia Elizabeth Núñez Llanos, Wendy Lizarraga Olivares, Luren Nieves Sevilla Catañeda, Kelly Vanessa Izarra Rojas, Karla Vasquez Cajachahua, Steve Vladimir Acedo Lazo, Omar Alberto Cáceres Rey, Henri Bailón Calderón, Priscila Nayu Lope Pari, Nancy Rojas Serrano, Gloria Arotinco Garayar. Equipo de vigilancia genomica del Instituto Nacional de Salud. |
| EPI_ISL_16138916, EPI_ISL_16138917, EPI_ISL_16138918, EPI_ISL_16138919, EPI_ISL_16138920, EPI_ISL_16138921, EPI_ISL_16138922, EPI_ISL_16138923, EPI_ISL_16138924, EPI_ISL_16138925, EPI_ISL_16138926, EPI_ISL_16138927, EPI_ISL_16138928, EPI_ISL_16138929, EPI_ISL_16138930, EPI_ISL_16138931, EPI_ISL_16138933, EPI_ISL_16138934, EPI_ISL_16138935, EPI_ISL_16138936, EPI_ISL_16138937, EPI_ISL_16138938, EPI_ISL_16138939, EPI_ISL_16138940, EPI_ISL_16138941, EPI_ISL_16138944, EPI_ISL_16138945 | | | |
| see above | California Department of Public Health | California Department of Public Health | Kath, C., Haw, M., Espinosa, A., and Hacker, J. |
| EPI_ISL_16233781 | Complejo Hospitalario Universitario de Pontevedra | Microbiology Department. Complexo Hospitalario Universitario de Vigo | Daviña C, Pizcueta J, Trigo M, Perez-Castro S |
| EPI_ISL_16233782, EPI_ISL_16233783, EPI_ISL_16233784, EPI_ISL_16233785, EPI_ISL_16233786, EPI_ISL_16233787, EPI_ISL_16233788 | Microbiology Department. Complexo Hospitalario Universitario de Vigo | Microbiology Department. Complexo Hospitalario Universitario de Vigo | Daviña C, Pizcueta J, Perez-Castro S |
| EPI_ISL_16467111 | IRCCS Sacro Cuore Don Calabria Hospital, Department of Infectious, Tropical Diseases & Microbiology | IRCCS Sacro Cuore Don Calabria Hospital, Department of Infectious, Tropical Diseases & Microbiology | Michela Deiana, Denise Lavezzari, Silvia Accordini, Concetta Castilletti, Antonio Mori, Elena Pomari, Chiara Piubelli |
| EPI_ISL_16505425 | Hopital Saint Louis | Hopital Saint Louis | Zeggagh,J., Ferraris,O., Salmona,M., Tarantola,A., Molina,J.M. and Delaugerre,C. |
| EPI_ISL_16510131, EPI_ISL_16510132, EPI_ISL_16510134, EPI_ISL_16510136, EPI_ISL_16510138, EPI_ISL_16510140, EPI_ISL_16510141, EPI_ISL_16510143, EPI_ISL_16510145, EPI_ISL_16510147, EPI_ISL_16510148, EPI_ISL_16510150, EPI_ISL_16510151, EPI_ISL_16510153, EPI_ISL_16510154, EPI_ISL_16510155, EPI_ISL_16510156, EPI_ISL_16510157, EPI_ISL_16510158, EPI_ISL_16510159, EPI_ISL_16510160, EPI_ISL_16510161, EPI_ISL_16510162, EPI_ISL_16510163, EPI_ISL_16510164, EPI_ISL_16510165, EPI_ISL_16510166, EPI_ISL_16510167, EPI_ISL_16510168, EPI_ISL_16510169, EPI_ISL_16510170, EPI_ISL_16510171, EPI_ISL_16510172, EPI_ISL_16510173, EPI_ISL_16510174, EPI_ISL_16510175, EPI_ISL_16510176, EPI_ISL_16510177, EPI_ISL_16510178, EPI_ISL_16510179, EPI_ISL_16510180, EPI_ISL_16510181, EPI_ISL_16510182, EPI_ISL_16510183, EPI_ISL_16510184, EPI_ISL_16510185 | | | |
| see above | National Virus Reference Laboratory | National Virus Reference Laboratory | Gabriel Gonzalez, Michael Carr, Brian Keogan, Jose Maria Urtasun Elizari, Jonathan Dean, Daniel Hare, Cillian F De Gascun |
| EPI_ISL_16588110, EPI_ISL_16588825, EPI_ISL_16589442 | Erasmus Medical Center Department of Virology | Erasmus Medical Center Department of Virology | Leonard Schuele, Bas Oude Munnink, Marjan Boter, Babette Weller, Babs Verstrepen, Richard Molenkamp, Janette Rahamat-Langendoen, Reina Sikkema, Marion Koopmans |
| EPI_ISL_16645206 | Division of High-risk Pathogens, Korea Disease Control and Prevention Agency | Division of High-risk Pathogens, Korea Disease Control and Prevention Agency | Rhie,G.-e. |
| EPI_ISL_16650224, EPI_ISL_16650225, EPI_ISL_16650226, EPI_ISL_16650227, EPI_ISL_16650228, EPI_ISL_16650229, EPI_ISL_16650230, EPI_ISL_16650231, EPI_ISL_16650232, EPI_ISL_16650234, EPI_ISL_16650237, EPI_ISL_16650238, EPI_ISL_16650240, EPI_ISL_16650241, EPI_ISL_16650243, EPI_ISL_16650244, EPI_ISL_16650246, EPI_ISL_16650247, EPI_ISL_16650248, EPI_ISL_16650249, EPI_ISL_16650251, EPI_ISL_16650253, EPI_ISL_16650255, EPI_ISL_16650258, EPI_ISL_16650260, EPI_ISL_16650262, EPI_ISL_16650265 | | | |
| see above | Laboratorio Central de Saude Publica do Estado de Minas Gerais (Lacen-MG) | Laboratorio Central de Saude Publica do Estado de Minas Gerais (Lacen-MG) | Felipe Campos de Melo Iani, Ludmila Oliveira Lamounier, Luiz Marcelo Ribeiro Tomé, Natália Rocha Guimarães,Talita Emile Ribeiro Adelino. |

| | | | |
|---|---|---|---|
| EPI_ISL_16679206 | Los Angeles County Public Health Laboratories | Los Angeles County Public Health Laboratories | P. Hemarajata et al. |
| EPI_ISL_16679210, EPI_ISL_16679223, EPI_ISL_16679225, EPI_ISL_16679235 | Kaiser Permanente Chino Hills Regional Reference Laboratories | Los Angeles County Public Health Laboratories | P. Hemarajata et al. |
| EPI_ISL_16679242, EPI_ISL_16679243, EPI_ISL_16679244, EPI_ISL_16679245, EPI_ISL_16679246, EPI_ISL_16679247, EPI_ISL_16679248, EPI_ISL_16679249, EPI_ISL_16679250, EPI_ISL_16679251, EPI_ISL_16679252, EPI_ISL_16679253, EPI_ISL_16679254, EPI_ISL_16679255, EPI_ISL_16679256, EPI_ISL_16679257, EPI_ISL_16679258, EPI_ISL_16679259, EPI_ISL_16679260, EPI_ISL_16679261, EPI_ISL_16679262, EPI_ISL_16679263, EPI_ISL_16679264, EPI_ISL_16679265, EPI_ISL_16679266, EPI_ISL_16679267, EPI_ISL_16679268, EPI_ISL_16679269, EPI_ISL_16679270, EPI_ISL_16679271, EPI_ISL_16679272, EPI_ISL_16679273, EPI_ISL_16679274, EPI_ISL_16679275, EPI_ISL_16679276, EPI_ISL_16679277, EPI_ISL_16679278, EPI_ISL_16679279, EPI_ISL_16679280, EPI_ISL_16679281, EPI_ISL_16679282, EPI_ISL_16679283, EPI_ISL_16679284, EPI_ISL_16679285, EPI_ISL_16679286, EPI_ISL_16679287, EPI_ISL_16679288, EPI_ISL_16679289, EPI_ISL_16679290, EPI_ISL_16679291, EPI_ISL_16679292, EPI_ISL_16679293, EPI_ISL_16679294, EPI_ISL_16679295, EPI_ISL_16679296, EPI_ISL_16679297, EPI_ISL_16679298, EPI_ISL_16679299, EPI_ISL_16679300, EPI_ISL_16679301, EPI_ISL_16679302, EPI_ISL_16679303, EPI_ISL_16679304, EPI_ISL_16679305 | | | |
| see above | California Department of Public Health | California Department of Public Health | Probert,W., Espinosa,A., Kath,C., Haw,M., O'Neil,R., Bell,J. and Hacker,J. |
| EPI_ISL_16727186, EPI_ISL_16727549 | Erasmus Medical Center, Department of Virology | Erasmus Medical Center Department of Virology | Leonard Schuele, Bas Oude Munnink, Marjan Boter, Babette Weller, Babs Verstrepen, Richard Molenkamp, Janette Rahamat-Langendoen, Reina Sikkema, Marion Koopmans |
| EPI_ISL_16751085, EPI_ISL_16751092 | Centre for Biological Threats, Highly Pathogenic Viruses, Robert Koch Institute | Centre for Biological Threats, Highly Pathogenic Viruses, Robert Koch Institute | Brinkmann,A., Kohl,C., Pape,K., Schrick,L., Michel,J., Schaade,L. and Nitsche,A. |
| EPI_ISL_16751099, EPI_ISL_16751100 | Centers for Disease Control & Prevention (CDC), Division of High Consequence Pathogens and Pathology (DHCPP-PRB) | Centers for Disease Control & Prevention (CDC), Division of High Consequence Pathogens and Pathology (DHCPP-PRB) | Gigante,C., Murray,J., Zhao,H., Batra,D., Hetrick,E., Howard,D., Kovar,L., Seabolt,M., Morrison,S., Desch,M., Knipe,K., Weigand,M., Sheth,M., Burroughs,A.B., Lee,J., Wilkins,K., McCollum,A., Hutson,C., Davidson,W., Rao,A., Atkinson,A. and Li,Y. |
| EPI_ISL_16751110, EPI_ISL_16751111 | Centers for Disease Control & Prevention (CDC), Division of High Consequence Pathogens and Pathology (DHCPP-PRB) | Centers for Disease Control & Prevention (CDC), Division of High Consequence Pathogens and Pathology (DHCPP-PRB) | Gigante,C., Bradley,A., Zhao,H., Batra,D., Hetrick,E., Howard,D., Kovar,L., Seabolt,M., Morrison,S., Desch,M., Knipe,K., Sheth,M.R., Burgin,A., Burroughs,M., Lee,J., Wilkins,K., McCollum,A., Hutson,C., Davidson,W., Rao,A., Anderson,J. and Li,Y. |
| EPI_ISL_16751112 | Centers for Disease Control & Prevention (CDC), Division of High Consequence Pathogens and Pathology (DHCPP-PRB) | Centers for Disease Control & Prevention (CDC), Division of High Consequence Pathogens and Pathology (DHCPP-PRB) | Gigante,C., Johnson,S., Zhao,H., Batra,D., Hetrick,E., Howard,D., Kovar,L., Seabolt,M., Morrison,S., Weigand,M., Knipe,K., Sheth,M., Burgin,A., Burroughs,M., Lee,J., Wilkins,K., McCollum,A., Hutson,C., Davidson,W., Rao,A., Riner,D. and Li,Y. |
| EPI_ISL_16751113 | Centers for Disease Control & Prevention (CDC), Division of High Consequence Pathogens and Pathology (DHCPP-PRB) | Centers for Disease Control & Prevention (CDC), Division of High Consequence Pathogens and Pathology (DHCPP-PRB) | Gigante,C., Cleavinger,K., Zhao,H., Batra,D., Hetrick,E., Howard,D., Kovar,L., Seabolt,M., Morrison,S., Desch,M., Knipe,K., Burroughs,M.R., Lee,J., Wilkins,K., McCollum,A., Hutson,C., Davidson,W., Rao,A., Sinn,M. and Li,Y. |
| EPI_ISL_16751116 | Centers for Disease Control & Prevention (CDC), Division of High Consequence Pathogens and Pathology (DHCPP-PRB) | Centers for Disease Control & Prevention (CDC), Division of High Consequence Pathogens and Pathology (DHCPP-PRB) | Gigante,C., Buttery,E., Zhao,H., Batra,D., Hetrick,E., Howard,D., Kovar,L., Seabolt,M., Morrison,S., Desch,M., Knipe,K., Weigand,M., Sheth,M., Burroughs,A.B., Lee,J., Wilkins,K., McCollum,A., Hutson,C., Davidson,W., Rao,A., Raman,D. and Li,Y. |
| EPI_ISL_16751118, EPI_ISL_16751122 | Centers for Disease Control & Prevention (CDC), Division of High Consequence Pathogens and Pathology (DHCPP-PRB) | Centers for Disease Control & Prevention (CDC), Division of High Consequence Pathogens and Pathology (DHCPP-PRB) | Gigante,C., Ruiz,V., Zhao,H., Batra,D., Hetrick,E., Howard,D., Kovar,L., Seabolt,M., Morrison,S., Desch,M., Knipe,K., Weigand,M., Sheth,M., Burgin,A., Burroughs,M., Lee,J., Wilkins,K., McCollum,A., Hutson,C., Davidson,W., Rao,A., Wang,J. and Li,Y. |
| EPI_ISL_16751124 | Centers for Disease Control & Prevention (CDC), Division of High Consequence Pathogens and Pathology (DHCPP-PRB) | Centers for Disease Control & Prevention (CDC), Division of High Consequence Pathogens and Pathology (DHCPP-PRB) | Gigante,C., Kubin,G., Zhao,H., Batra,D., Hetrick,E., Howard,D., Kovar,L., Seabolt,M., Morrison,S., Desch,M., Knipe,K., Sheth,M.R., Burgin,A., Burroughs,M., Lee,J., Wilkins,K., McCollum,A., Hutson,C., Davidson,W., Rao,A., White,S. and Li,Y. |
| EPI_ISL_16751134 | Centers for Disease Control & Prevention (CDC), Division of High Consequence Pathogens and Pathology (DHCPP-PRB) | Centers for Disease Control & Prevention (CDC), Division of High Consequence Pathogens and Pathology (DHCPP-PRB) | Gigante,C., Hauser,J., Zhao,H., Batra,D., Hetrick,E., Howard,D., Kovar,L., Seabolt,M., Knipe,K., Burroughs,M.S., Lee,J., Wilkins,K., McCollum,A., Hutson,C., Davidson,W., Rao,A., Mangla,A. and Li,Y. |
| EPI_ISL_16751135 | Centers for Disease Control & Prevention (CDC), Division of High Consequence Pathogens and Pathology (DHCPP-PRB) | Centers for Disease Control & Prevention (CDC), Division of High Consequence Pathogens and Pathology (DHCPP-PRB) | Gigante,C., Lee,P., Zhao,H., Batra,D., Hetrick,E., Howard,D., Kovar,L., Seabolt,M., Morrison,S., Desch,M., Knipe,K., Weigand,M., Burroughs,M.S., Lee,J., Wilkins,K., McCollum,A., Hutson,C., Davidson,W., Rao,A., Stanek,D. and Li,Y. |
| EPI_ISL_16847486, EPI_ISL_16847487 | Institute for Medical Virology, University Hospital, Goethe University | Institute for Medical Virology, University Hospital, Goethe University | Denisa Bojkova, Julia Schneider, Victor M. Corman, Martin Michaelis, Jindrich Cinatl jr. |
| EPI_ISL_16871158, EPI_ISL_16871159, EPI_ISL_16871160, EPI_ISL_16871161, EPI_ISL_16871162, EPI_ISL_16871163 | Laboratorio de Enterovirus, Instituto Oswaldo Cruz, Fiocruz | Instituto Oswaldo Cruz FIOCRUZ - Laboratory of Respiratory Viruses and Measles (LVRS) | Paola Resende, Elisa Cavalcante Pereira, Bruna Mendonça da Silva, Jéssica Graça Macedo de Carvalho, Larissa Macedo Pinto, Victor Guimaraes, Marilda Siqueira, Renan da Silva Faustino, Marilia Santini, Edson Elias da Silva on behalf of the Fiocruz Genomic Surveillance Network |
| EPI_ISL_16930151, EPI_ISL_16930157, EPI_ISL_16930168 | California Department of Public Health | California Department of Public Health | Probert,W., Espinosa,A., Kath,C., Haw,M., O'Neil,R., Bell,J. and Hacker,J. |
| EPI_ISL_16946400 | Division de Microbiologia, Hospital Nacional de Niños Carlos Saenz Herrera | Instituto Costarricense de Investigacion y Enseñanza en Nutricion y Salud, Inciensa | Diana Cantillo, Hillary Serrano, Ana Isela Ruiz, Gustavo Vega, Claudio Soto-Garita, Adriana Godinez, Estela Cordero, Melany Calderon, Francisco Duarte |
| EPI_ISL_16955153, EPI_ISL_16955154, EPI_ISL_16955155, EPI_ISL_16955156, EPI_ISL_16955157, EPI_ISL_16955158, EPI_ISL_16955159, EPI_ISL_16955160 | CT Department of Public Health | CT Department of Public Health | Claire Pearson, Tu N. Nguyen, Kutluhan Incekara, Neranjan V. Perera |
| EPI_ISL_16955204, EPI_ISL_16955205, EPI_ISL_16955206, EPI_ISL_16955207, EPI_ISL_16955208, EPI_ISL_16955209, EPI_ISL_16955210, EPI_ISL_16955211, EPI_ISL_16955212, EPI_ISL_16955213, EPI_ISL_16955214, EPI_ISL_16955215, EPI_ISL_16955216, EPI_ISL_16955217, EPI_ISL_16955218, EPI_ISL_16955219, EPI_ISL_16955220, EPI_ISL_16955221, EPI_ISL_16955222, EPI_ISL_16955227, EPI_ISL_16955228, EPI_ISL_16955229, EPI_ISL_16955230, EPI_ISL_16955239, EPI_ISL_16955240, EPI_ISL_16955241, EPI_ISL_16955242, EPI_ISL_16955243, EPI_ISL_16955244, EPI_ISL_16955245, EPI_ISL_16955246, EPI_ISL_16955247, EPI_ISL_16955248, EPI_ISL_16955249, EPI_ISL_16955250, EPI_ISL_16955252, EPI_ISL_16955253, EPI_ISL_16955254, EPI_ISL_16955255, EPI_ISL_16955256, EPI_ISL_16955257, EPI_ISL_16955258, EPI_ISL_16955259, EPI_ISL_16955260, EPI_ISL_16955261, EPI_ISL_16955262, EPI_ISL_16955263, EPI_ISL_16955264, EPI_ISL_16955265, EPI_ISL_16955266, EPI_ISL_16955267, EPI_ISL_16955268, EPI_ISL_16955270, EPI_ISL_16955271, EPI_ISL_16955272, EPI_ISL_16955273, EPI_ISL_16955274, EPI_ISL_16955275, EPI_ISL_16955276, EPI_ISL_16955277, EPI_ISL_16955278, EPI_ISL_16955279, EPI_ISL_16955280, EPI_ISL_16955281, EPI_ISL_16955282, EPI_ISL_16955283, EPI_ISL_16955284, EPI_ISL_16955285, EPI_ISL_16955286, EPI_ISL_16955287, EPI_ISL_16955288, EPI_ISL_16955289, EPI_ISL_16955290, EPI_ISL_16955291, EPI_ISL_16955292, EPI_ISL_16955294, EPI_ISL_16955295, EPI_ISL_16955296, EPI_ISL_16955297, EPI_ISL_16955298, EPI_ISL_16955299, EPI_ISL_16955300, EPI_ISL_16955301, EPI_ISL_16955302, EPI_ISL_16955303, EPI_ISL_16955304, EPI_ISL_16955305, EPI_ISL_16955306, EPI_ISL_16955307, EPI_ISL_16955308, EPI_ISL_16955309, EPI_ISL_16955310, EPI_ISL_16955311, EPI_ISL_16955312, EPI_ISL_16955313, EPI_ISL_16955314, EPI_ISL_16955315, EPI_ISL_16955316 | | | |
| see above | Public Health Laboratory, NYC Department of Health and Mental Hygiene | Public Health Laboratory, NYC Department of Health and Mental Hygiene | Wang,J.C., Amin,H.S., Clabby,T.T., Taki,F., Su,M., Rahat,A., De La Cruz,N., Olsen,A., Thi,C., Silver,S., Akther,S., Chowdhury,M., Omoregie,E. and Hughes,S. |
| EPI_ISL_16985950, EPI_ISL_16985951, EPI_ISL_16985952, EPI_ISL_16985953, EPI_ISL_16985954, EPI_ISL_16985955, EPI_ISL_16985956, EPI_ISL_16985957, EPI_ISL_16985958, EPI_ISL_16985959, EPI_ISL_16985960, EPI_ISL_16985961, EPI_ISL_16985962, EPI_ISL_16985963, EPI_ISL_16985964, EPI_ISL_16985965, EPI_ISL_16985966, EPI_ISL_16985967, EPI_ISL_16985968, EPI_ISL_16985969, EPI_ISL_16985970, EPI_ISL_16985971, EPI_ISL_16985972 | | | |
| see above | National Virus Reference Laboratory | National Virus Reference Laboratory | Gabriel Gonzalez, Michael Carr, Emer O'Byrne, Weronika Banka, Brian Keogan, Jose Maria Urtasun Elizari, Jonathan Dean, Daniel Hare, Cillian F De Gascun |
| EPI_ISL_16997394 | California Department of Public Health | California Department of Public Health | Probert,W., Espinosa,A., Kath,C., Haw,M., O'Neil,R., Bell,J. and Hacker,J. |
| EPI_ISL_16997415, EPI_ISL_16997417, EPI_ISL_16997418, EPI_ISL_16997421, EPI_ISL_16997422, EPI_ISL_16997423, EPI_ISL_16997424, EPI_ISL_16997429, EPI_ISL_16997432, EPI_ISL_16997433, EPI_ISL_16997434, EPI_ISL_16997435, EPI_ISL_16997436, EPI_ISL_16997437, EPI_ISL_16997438, EPI_ISL_16997439, EPI_ISL_16997440, EPI_ISL_16997444, EPI_ISL_16997445, EPI_ISL_16997446 | | | |
| see above | Kaiser Permanente Chino Hills Regional Reference Laboratories | Los Angeles County Public Health Laboratories | P. Hemarajata et al. |
| EPI_ISL_16997447 | Laboratory Corporation of America | Los Angeles County Public Health Laboratories | P. Hemarajata et al. |
| EPI_ISL_16997455 | Los Angeles County Public Health Laboratories | Los Angeles County Public Health Laboratories | P. Hemarajata et al. |
| EPI_ISL_16997460 | Quest Diagnostics Nichols Institute | Los Angeles County Public Health Laboratories | P. Hemarajata et al. |
| EPI_ISL_16997470 | UCLA Clinical Micro Lab | Los Angeles County Public Health Laboratories | P. Hemarajata et al. |
| EPI_ISL_16999059, EPI_ISL_16999060, EPI_ISL_16999061, EPI_ISL_16999062, EPI_ISL_16999063, EPI_ISL_16999064, EPI_ISL_16999065, EPI_ISL_16999066, EPI_ISL_16999067, EPI_ISL_16999068, EPI_ISL_16999069, EPI_ISL_16999070, EPI_ISL_16999071, EPI_ISL_16999072, EPI_ISL_16999073, EPI_ISL_16999074, EPI_ISL_16999075, EPI_ISL_16999076, EPI_ISL_16999077, EPI_ISL_16999078, EPI_ISL_16999079, EPI_ISL_16999080, EPI_ISL_16999081, EPI_ISL_16999082, EPI_ISL_16999083, EPI_ISL_16999084, EPI_ISL_16999085, EPI_ISL_16999086, EPI_ISL_16999087, EPI_ISL_16999088, EPI_ISL_16999089, EPI_ISL_16999090, EPI_ISL_16999091, EPI_ISL_16999092, EPI_ISL_16999093, EPI_ISL_16999094, EPI_ISL_16999095, EPI_ISL_16999096, EPI_ISL_16999097, EPI_ISL_16999098, EPI_ISL_16999099, EPI_ISL_16999100, EPI_ISL_16999101, EPI_ISL_16999102, EPI_ISL_16999103, EPI_ISL_16999104, EPI_ISL_16999105, EPI_ISL_16999106, EPI_ISL_16999107, EPI_ISL_16999108, EPI_ISL_16999109, EPI_ISL_16999110, EPI_ISL_16999111, EPI_ISL_16999112, EPI_ISL_16999113, EPI_ISL_16999114, EPI_ISL_16999115, EPI_ISL_16999116, EPI_ISL_16999117, EPI_ISL_16999118, EPI_ISL_16999119, EPI_ISL_16999120, EPI_ISL_16999121, EPI_ISL_16999122, EPI_ISL_16999123, EPI_ISL_16999124, EPI_ISL_16999125, EPI_ISL_16999126, EPI_ISL_16999127, EPI_ISL_16999128, EPI_ISL_16999129, EPI_ISL_16999130, EPI_ISL_16999131, EPI_ISL_16999132, EPI_ISL_16999133, EPI_ISL_16999134, EPI_ISL_16999135, EPI_ISL_16999136, EPI_ISL_16999137, EPI_ISL_16999138, EPI_ISL_16999139 | | | |
| see above | Laboratorio de Referencia Nacional de Virus Inmunoprevenibles. Centro Nacional de Salud Publica. Instiuto Nacional de Salud | Equipo de Vigilancia Genomica. Area de Innovación y Desarrollo. Centro Nacional de Salud Publica. Instiuto Nacional de Salud | Carlos Patricio Padilla Rojas, Carmen Verónica Hurtado Vela, Juana Iris Silva Molina, Luis Bárcena Flores, Víctor Jiménez Vásquez, Alicia Elizabeth Núñez Llanos, Wendy Lizarraga Olivares, Luren Nieves Sevilla Catañeda, Kelly Vanessa Izarra Rojas, Karla Vasquez Cajachahua, Steve Vladimir Acedo Lazo, Omar Alberto Cáceres Rey, Henri Bailón Calderón, Priscila Nayu Lope Pari, Nancy Rojas Serrano, Gloria Arotinco Garayar. Equipo de Vigilancia Genomica del Instituto Nacional de Salud. |
| EPI_ISL_17012019, EPI_ISL_17012020, EPI_ISL_17012023, EPI_ISL_17012024, EPI_ISL_17012025, EPI_ISL_17012026, EPI_ISL_17012027, EPI_ISL_17012028, EPI_ISL_17012029, EPI_ISL_17012030, EPI_ISL_17012031, EPI_ISL_17012032, EPI_ISL_17012033, EPI_ISL_17012034, EPI_ISL_17012035, EPI_ISL_17012036, EPI_ISL_17012037, EPI_ISL_17012038, EPI_ISL_17012039, EPI_ISL_17012040, EPI_ISL_17012041, EPI_ISL_17012042, EPI_ISL_17012043, EPI_ISL_17012044, EPI_ISL_17012046, EPI_ISL_17012047, EPI_ISL_17012048, EPI_ISL_17012049, EPI_ISL_17012050, EPI_ISL_17012051, EPI_ISL_17012052, EPI_ISL_17012053, EPI_ISL_17012054, EPI_ISL_17012055, EPI_ISL_17012056, EPI_ISL_17012057, EPI_ISL_17012058, EPI_ISL_17012059, EPI_ISL_17012060, EPI_ISL_17012061, EPI_ISL_17012062, EPI_ISL_17012063, EPI_ISL_17012064, EPI_ISL_17012066, EPI_ISL_17012067, EPI_ISL_17012068, EPI_ISL_17012069, EPI_ISL_17012071 | | | |
| see above | Laboratorio de Virus Exantematicos, Gastroentéricos y Otros Transmitidos por Vectores | Centro de Referencia Nacional de Genomica, Secuenciacion y Bioinformatica GENSBIO, INSPI-CZ9 | Andrés Carrazco, Silvia Salgado, Diana Gutiérrez, Damaris Alarcón, Andrés Herrera, Andrés Tinizaray, Martha Sánchez, Johanna Parrales, Diego Morales, Jorge Bejarano, Leandro Patiño. |
| EPI_ISL_17012072 | Laboratorio de Virus Exantematicos, Gastroentéricos y Otros Transmitidos por Vectores | Centro de Referencia Nacional de Genomica, Secuenciacion y Bioinformatica GENSBIO, INSPI-CZ11 | Andrés Carrazco, Silvia Salgado, Diana Gutiérrez, Damaris Alarcón, Andrés Herrera, Andrés Tinizaray, Martha Sánchez, Johanna Parrales, Diego Morales, Jorge Bejarano, Leandro Patiño. |
| EPI_ISL_17012073 | Laboratorio de Virus Exantematicos, Gastroentéricos y Otros Transmitidos por Vectores | Centro de Referencia Nacional de Genomica, Secuenciacion y Bioinformatica GENSBIO, INSPI-CZ12 | Andrés Carrazco, Silvia Salgado, Diana Gutiérrez, Damaris Alarcón, Andrés Herrera, Andrés Tinizaray, Martha Sánchez, Johanna Parrales, Diego Morales, Jorge Bejarano, Leandro Patiño. |
| EPI_ISL_17012074 | Laboratorio de Virus Exantematicos, Gastroentéricos y Otros Transmitidos por Vectores | Centro de Referencia Nacional de Genomica, Secuenciacion y Bioinformatica GENSBIO, INSPI-CZ14 | Andrés Carrazco, Silvia Salgado, Diana Gutiérrez, Damaris Alarcón, Andrés Herrera, Andrés Tinizaray, Martha Sánchez, Johanna Parrales, Diego Morales, Jorge Bejarano, Leandro Patiño. |
| EPI_ISL_17012075 | Laboratorio de Virus Exantematicos, Gastroentéricos y Otros Transmitidos por Vectores | Centro de Referencia Nacional de Genomica, Secuenciacion y Bioinformatica GENSBIO, INSPI-CZ15 | Andrés Carrazco, Silvia Salgado, Diana Gutiérrez, Damaris Alarcón, Andrés Herrera, Andrés Tinizaray, Martha Sánchez, Johanna Parrales, Diego Morales, Jorge Bejarano, Leandro Patiño. |
| EPI_ISL_17012076 | Laboratorio de Virus Exantematicos, Gastroentéricos y Otros Transmitidos por Vectores | Centro de Referencia Nacional de Genomica, Secuenciacion y Bioinformatica GENSBIO, INSPI-CZ16 | Andrés Carrazco, Silvia Salgado, Diana Gutiérrez, Damaris Alarcón, Andrés Herrera, Andrés Tinizaray, Martha Sánchez, Johanna Parrales, Diego Morales, Jorge Bejarano, Leandro Patiño. |
| EPI_ISL_17012077 | Laboratorio de Virus Exantematicos, Gastroentéricos y | Centro de Referencia Nacional de Genomica, | Andrés Carrazco, Silvia Salgado, Diana Gutiérrez, Damaris Alarcón, Andrés Herrera, Andrés Tinizaray, Martha Sánchez, Johanna Parrales, Diego Morales, Jorge Bejarano, Leandro Patiño. |

| | | | |
|---|---|---|---|
| | Otros Transmitidos por Vectores | Secuenciacion y Bioinformatica GENSBIO, INSPI-CZ18 | |
| EPI_ISL_17012078, EPI_ISL_17012079, EPI_ISL_17012080, EPI_ISL_17012081, EPI_ISL_17012082, EPI_ISL_17012083, EPI_ISL_17012084, EPI_ISL_17012085, EPI_ISL_17012086, EPI_ISL_17012087, EPI_ISL_17012088, EPI_ISL_17012089, EPI_ISL_17012090, EPI_ISL_17012091, EPI_ISL_17012092, EPI_ISL_17012094, EPI_ISL_17012095, EPI_ISL_17012096, EPI_ISL_17012097, EPI_ISL_17012100, EPI_ISL_17012101, EPI_ISL_17012102, EPI_ISL_17012103, EPI_ISL_17012104, EPI_ISL_17012105, EPI_ISL_17012106, EPI_ISL_17012107, EPI_ISL_17012108, EPI_ISL_17012109, EPI_ISL_17012110, EPI_ISL_17012111, EPI_ISL_17012113, EPI_ISL_17012115 | | | |
| see above | Laboratorio de Virus Exantematicos, Gastroentéricos y Otros Transmitidos por Vectores | Centro de Referencia Nacional de Genomica, Secuenciacion y Bioinformatica GENSBIO, INSPI-CZ9 | Andrés Carrazco-Motalvo, Silvia Salgado, Diana Gutiérrez, Damaris Alarcón, Andrés Herrera, Andrés Tinizaray, Ruth Gómez, Martha Sánchez, Johanna Parrales, Diego Morales, Jorge Bejarano, Leandro Patiño. |
| EPI_ISL_17018429, EPI_ISL_17018431, EPI_ISL_17018433, EPI_ISL_17018434, EPI_ISL_17018436, EPI_ISL_17018438, EPI_ISL_17018439 | National Institute for Infectious Diseases "Matei Bals" | National Institute for Infectious Diseases "Matei Bals" | Robert Hohan, Marius Surleac, Leontina Banica, Andreea Tudor, Simona Paraschiv |
| EPI_ISL_17019459, EPI_ISL_17019461, EPI_ISL_17019462, EPI_ISL_17019464, EPI_ISL_17019467 | Parkland Health and Hospital System | Dallas County Health & Human Services Public Health Laboratory | Kabir, Farruk; Plaisance, Erin; Stringer, Joey; Short, Luke. |
| EPI_ISL_17019470 | MD Progressive Care | Dallas County Health & Human Services Public Health Laboratory | Kabir, Farruk; Plaisance, Erin; Stringer, Joey; Short, Luke. |
| EPI_ISL_17019472 | White Rock Medical Center | Dallas County Health & Human Services Public Health Laboratory | Kabir, Farruk; Plaisance, Erin; Stringer, Joey; Short, Luke. |
| EPI_ISL_17019473 | Children's Health Dallas Texas | Dallas County Health & Human Services Public Health Laboratory | Kabir, Farruk; Plaisance, Erin; Stringer, Joey; Short, Luke. |
| EPI_ISL_17019476 | Dallas County Jail | Dallas County Health & Human Services Public Health Laboratory | Kabir, Farruk; Plaisance, Erin; Stringer, Joey; Short, Luke. |
| EPI_ISL_17048204, EPI_ISL_17048205, EPI_ISL_17048206, EPI_ISL_17048207 | Laboratorio Central de Saude Publica do Estado da Bahia (LACEN/BA) | Laboratory of Respiratory Viruses and Measles, Oswaldo Cruz Institute, FIOCRUZ | Paola Resende, Fernando Motta, Elisa Cavalcante Pereira, Bruna Mendonça da Silva, Jéssica Graça Macedo de Carvalho, Larissa Macedo Pinto, Victor Guimaraes, Felicidade Pereira, Marilda Siqueira, Renan da Silva Faustino, Marilia Santini, Edson Elias da Silva on behalf of the Fiocruz COVID-19 Genomic Surveillance Network |
| EPI_ISL_17048208 | Laboratorio de Enterovirus, Instituto Oswaldo Cruz, Fiocruz | Laboratory of Respiratory Viruses and Measles, Oswaldo Cruz Institute, FIOCRUZ | Paola Resende, Elisa Cavalcante Pereira, Bruna Mendonça da Silva, Jéssica Graça Macedo de Carvalho, Larissa Macedo Pinto, Victor Guimaraes, Marilda Siqueira, Renan da Silva Faustino, Marilia Santini, Edson Elias da Silva on behalf of the Fiocruz Genomic Surveillance Network |
| EPI_ISL_17085647, EPI_ISL_17085649, EPI_ISL_17085650, EPI_ISL_17085652, EPI_ISL_17085653, EPI_ISL_17085655, EPI_ISL_17085656, EPI_ISL_17085658, EPI_ISL_17085659, EPI_ISL_17085661, EPI_ISL_17085662, EPI_ISL_17085664, EPI_ISL_17085665, EPI_ISL_17085667, EPI_ISL_17085668, EPI_ISL_17085670, EPI_ISL_17085671, EPI_ISL_17085673, EPI_ISL_17085674, EPI_ISL_17085676, EPI_ISL_17085678, EPI_ISL_17085679, EPI_ISL_17085681, EPI_ISL_17085682, EPI_ISL_17085684, EPI_ISL_17085685, EPI_ISL_17085687, EPI_ISL_17085688, EPI_ISL_17085690, EPI_ISL_17085692, EPI_ISL_17085693, EPI_ISL_17085695, EPI_ISL_17085696, EPI_ISL_17085698, EPI_ISL_17085700, EPI_ISL_17085701, EPI_ISL_17085703, EPI_ISL_17085704, EPI_ISL_17085706, EPI_ISL_17085708, EPI_ISL_17085709, EPI_ISL_17085711, EPI_ISL_17085712 | | | |
| see above | Public Health Laboratory, NYC Department of Health and Mental Hygiene | Public Health Laboratory, NYC Department of Health and Mental Hygiene | Wang,J.C., Amin,H.S., Clabby,T.T., Taki,F., Su,M., Rahat,A., De La Cruz,N., Olsen,A., Thi,C., Silver,S., Akther,S., Chowdhury,M., Omoregie,E. and Hughes,S. |
| EPI_ISL_17085714, EPI_ISL_17085715, EPI_ISL_17085717, EPI_ISL_17085719, EPI_ISL_17085720, EPI_ISL_17085722, EPI_ISL_17085723, EPI_ISL_17085725, EPI_ISL_17085726, EPI_ISL_17085728, EPI_ISL_17085730, EPI_ISL_17085731, EPI_ISL_17085733, EPI_ISL_17085734, EPI_ISL_17085736, EPI_ISL_17085737, EPI_ISL_17085739, EPI_ISL_17085740, EPI_ISL_17085742, EPI_ISL_17085743, EPI_ISL_17085745, EPI_ISL_17085746, EPI_ISL_17085748, EPI_ISL_17085750, EPI_ISL_17085752, EPI_ISL_17085753, EPI_ISL_17085755, EPI_ISL_17085756, EPI_ISL_17085758, EPI_ISL_17085760, EPI_ISL_17085761, EPI_ISL_17085762, EPI_ISL_17085764, EPI_ISL_17085765, EPI_ISL_17085767, EPI_ISL_17085769, EPI_ISL_17085770, EPI_ISL_17085771, EPI_ISL_17085773, EPI_ISL_17085774, EPI_ISL_17085776, EPI_ISL_17085777, EPI_ISL_17085779, EPI_ISL_17085781, EPI_ISL_17085782, EPI_ISL_17085784, EPI_ISL_17085785, EPI_ISL_17085787, EPI_ISL_17085788, EPI_ISL_17085790, EPI_ISL_17085792, EPI_ISL_17085793, EPI_ISL_17085794, EPI_ISL_17085795, EPI_ISL_17085796, EPI_ISL_17085797, EPI_ISL_17085798, EPI_ISL_17085920, EPI_ISL_17085921, EPI_ISL_17085922, EPI_ISL_17085923, EPI_ISL_17085924, EPI_ISL_17085925, EPI_ISL_17085926, EPI_ISL_17085927, EPI_ISL_17085928, EPI_ISL_17085929, EPI_ISL_17085930, EPI_ISL_17085931, EPI_ISL_17085932, EPI_ISL_17085933, EPI_ISL_17085934, EPI_ISL_17085936, EPI_ISL_17085937, EPI_ISL_17085938, EPI_ISL_17085939, EPI_ISL_17085940, EPI_ISL_17085941, EPI_ISL_17085942, EPI_ISL_17085943, EPI_ISL_17085944, EPI_ISL_17085945, EPI_ISL_17085946, EPI_ISL_17085947, EPI_ISL_17085948, EPI_ISL_17085949, EPI_ISL_17085950, EPI_ISL_17085951, EPI_ISL_17085952, EPI_ISL_17085953, EPI_ISL_17085954, EPI_ISL_17085955, EPI_ISL_17085956, EPI_ISL_17085957, EPI_ISL_17085958, EPI_ISL_17085959, EPI_ISL_17085960, EPI_ISL_17085961 | | | |
| see above | Public Health Laboratory, NYC Department of Health and Mental Hygiene | Public Health Laboratory, NYC Department of Health and Mental Hygiene | Clabby,T.T., Amin,H.S., Wang,J.C., Taki,F., Su,M., Rahat,A., De La Cruz,N., Olsen,A., Thi,C., Silver,S., Akther,S., Chowdhury,M., Omoregie,E. and Hughes,S. |
| EPI_ISL_17085962, EPI_ISL_17085963, EPI_ISL_17085964, EPI_ISL_17085965, EPI_ISL_17085966, EPI_ISL_17085967, EPI_ISL_17085968, EPI_ISL_17085969, EPI_ISL_17085970, EPI_ISL_17085971, EPI_ISL_17085972, EPI_ISL_17085973, EPI_ISL_17085974, EPI_ISL_17085975, EPI_ISL_17085976, EPI_ISL_17085977, EPI_ISL_17085978, EPI_ISL_17085979, EPI_ISL_17085980, EPI_ISL_17085981, EPI_ISL_17085982, EPI_ISL_17085983, EPI_ISL_17085984, EPI_ISL_17085985, EPI_ISL_17085986, EPI_ISL_17085987, EPI_ISL_17085988, EPI_ISL_17085989, EPI_ISL_17085990, EPI_ISL_17085991, EPI_ISL_17085992, EPI_ISL_17085993, EPI_ISL_17085994, EPI_ISL_17085995, EPI_ISL_17085996, EPI_ISL_17085997, EPI_ISL_17085998, EPI_ISL_17085999, EPI_ISL_17086000, EPI_ISL_17086001, EPI_ISL_17086002, EPI_ISL_17086003, EPI_ISL_17086004, EPI_ISL_17086005, EPI_ISL_17086006, EPI_ISL_17086007, EPI_ISL_17086008, EPI_ISL_17086009, EPI_ISL_17086010, EPI_ISL_17086011, EPI_ISL_17086012, EPI_ISL_17086013, EPI_ISL_17086014, EPI_ISL_17086015, EPI_ISL_17086016, EPI_ISL_17086017, EPI_ISL_17086018, EPI_ISL_17086019, EPI_ISL_17086020, EPI_ISL_17086021, EPI_ISL_17086022, EPI_ISL_17086023, EPI_ISL_17086024, EPI_ISL_17086025, EPI_ISL_17086026, EPI_ISL_17086027, EPI_ISL_17086028, EPI_ISL_17086029, EPI_ISL_17086030, EPI_ISL_17086031, EPI_ISL_17086032, EPI_ISL_17086033, EPI_ISL_17086035, EPI_ISL_17086036, EPI_ISL_17086037, EPI_ISL_17086038, EPI_ISL_17086039, EPI_ISL_17086040, EPI_ISL_17086041, EPI_ISL_17086042, EPI_ISL_17086043, EPI_ISL_17086044, EPI_ISL_17086045, EPI_ISL_17086046, EPI_ISL_17086047, EPI_ISL_17086048, EPI_ISL_17086049, EPI_ISL_17086050, EPI_ISL_17086051, EPI_ISL_17086052, EPI_ISL_17086053, EPI_ISL_17086054, EPI_ISL_17086055, EPI_ISL_17086056, EPI_ISL_17086057, EPI_ISL_17086058, EPI_ISL_17086059, EPI_ISL_17086060, EPI_ISL_17086061 | | | |
| see above | Public Health Laboratory, NYC Department of Health and Mental Hygiene | Public Health Laboratory, NYC Department of Health and Mental Hygiene | Amin,H.S., Clabby,T.T., Wang,J.C., Taki,F., Su,M., Rahat,A., De La Cruz,N., Olsen,A., Thi,C., Silver,S., Akther,S., Chowdhury,M., Omoregie,E. and Hughes,S. |
| EPI_ISL_17086062, EPI_ISL_17086063, EPI_ISL_17086064, EPI_ISL_17086065, EPI_ISL_17086066, EPI_ISL_17086067, EPI_ISL_17086068, EPI_ISL_17086069, EPI_ISL_17086070, EPI_ISL_17086071, EPI_ISL_17086072, EPI_ISL_17086073, EPI_ISL_17086074, EPI_ISL_17086075, EPI_ISL_17086076, EPI_ISL_17086077, EPI_ISL_17086078, EPI_ISL_17086079, EPI_ISL_17086080, EPI_ISL_17086081, EPI_ISL_17086082, EPI_ISL_17086083, EPI_ISL_17086084, EPI_ISL_17086085, EPI_ISL_17086086, EPI_ISL_17086087, EPI_ISL_17086088, EPI_ISL_17086089, EPI_ISL_17086090, EPI_ISL_17086091, EPI_ISL_17086092, EPI_ISL_17086093, EPI_ISL_17086095, EPI_ISL_17086096, EPI_ISL_17086097, EPI_ISL_17086098, EPI_ISL_17086099, EPI_ISL_17086100, EPI_ISL_17086101, EPI_ISL_17086102, EPI_ISL_17086103, EPI_ISL_17086104, EPI_ISL_17086105, EPI_ISL_17086106, EPI_ISL_17086107, EPI_ISL_17086108, EPI_ISL_17086109, EPI_ISL_17086110, EPI_ISL_17086111, EPI_ISL_17086112, EPI_ISL_17086113, EPI_ISL_17086114, EPI_ISL_17086115, EPI_ISL_17086116, EPI_ISL_17086117, EPI_ISL_17086118, EPI_ISL_17086119, EPI_ISL_17086120, EPI_ISL_17086121, EPI_ISL_17086122, EPI_ISL_17086123, EPI_ISL_17086124, EPI_ISL_17086125, EPI_ISL_17086126, EPI_ISL_17086127, EPI_ISL_17086128, EPI_ISL_17086129, EPI_ISL_17086130, EPI_ISL_17086131, EPI_ISL_17086132, EPI_ISL_17086133, EPI_ISL_17086135, EPI_ISL_17086136, EPI_ISL_17086137, EPI_ISL_17086138, EPI_ISL_17086139, EPI_ISL_17086140, EPI_ISL_17086141, EPI_ISL_17086142, EPI_ISL_17086143, EPI_ISL_17086144, EPI_ISL_17086145, EPI_ISL_17086146, EPI_ISL_17086147, EPI_ISL_17086148, EPI_ISL_17086149, EPI_ISL_17086150, EPI_ISL_17086151, EPI_ISL_17086152, EPI_ISL_17086153, EPI_ISL_17086154, EPI_ISL_17086155, EPI_ISL_17086156, EPI_ISL_17086157, EPI_ISL_17086158, EPI_ISL_17086159, EPI_ISL_17086160, EPI_ISL_17086161, EPI_ISL_17086162, EPI_ISL_17086163, EPI_ISL_17086164, EPI_ISL_17086165, EPI_ISL_17086166, EPI_ISL_17086167, EPI_ISL_17086168, EPI_ISL_17086169, EPI_ISL_17086170, EPI_ISL_17086171, EPI_ISL_17086172, EPI_ISL_17086173, EPI_ISL_17086175, EPI_ISL_17086176, EPI_ISL_17086177, EPI_ISL_17086178, EPI_ISL_17086179, EPI_ISL_17086180, EPI_ISL_17086181, EPI_ISL_17086182, EPI_ISL_17086183, EPI_ISL_17086184, EPI_ISL_17086185, EPI_ISL_17086186, EPI_ISL_17086187, EPI_ISL_17086188, EPI_ISL_17086189, EPI_ISL_17086190, EPI_ISL_17086191, EPI_ISL_17086192, EPI_ISL_17086193, EPI_ISL_17086194, EPI_ISL_17086195, EPI_ISL_17086196, EPI_ISL_17086197, EPI_ISL_17086198, EPI_ISL_17086199, EPI_ISL_17086200, EPI_ISL_17086201, EPI_ISL_17086202, EPI_ISL_17086203, EPI_ISL_17086204, EPI_ISL_17086205, EPI_ISL_17086206, EPI_ISL_17086207, EPI_ISL_17086208, EPI_ISL_17086209, EPI_ISL_17086210, EPI_ISL_17086211, EPI_ISL_17086212, EPI_ISL_17086213, EPI_ISL_17086214, EPI_ISL_17086215, EPI_ISL_17086216, EPI_ISL_17086217, EPI_ISL_17086218, EPI_ISL_17086219, EPI_ISL_17086232, EPI_ISL_17086233, EPI_ISL_17086234, EPI_ISL_17086235, EPI_ISL_17086236, EPI_ISL_17086237, EPI_ISL_17086238, EPI_ISL_17086239, EPI_ISL_17086240, EPI_ISL_17086241, EPI_ISL_17086242, EPI_ISL_17086243, EPI_ISL_17086244, EPI_ISL_17086245, EPI_ISL_17086246, EPI_ISL_17086247, EPI_ISL_17086248, EPI_ISL_17086249, EPI_ISL_17086250, EPI_ISL_17086251, EPI_ISL_17086252, EPI_ISL_17086253, EPI_ISL_17086254, EPI_ISL_17086255, EPI_ISL_17086256, EPI_ISL_17086257, EPI_ISL_17086258, EPI_ISL_17086259, EPI_ISL_17086260, EPI_ISL_17086261, EPI_ISL_17086262, EPI_ISL_17086263, EPI_ISL_17086264, EPI_ISL_17086265, EPI_ISL_17086266, EPI_ISL_17086267, EPI_ISL_17086268, EPI_ISL_17086269, EPI_ISL_17086270, EPI_ISL_17086271, EPI_ISL_17086272, EPI_ISL_17086273 | | | |
| see above | Public Health Laboratory, NYC Department of Health and Mental Hygiene | Public Health Laboratory, NYC Department of Health and Mental Hygiene | Wang,J.C., Amin,H.S., Clabby,T.T., Taki,F., Su,M., Rahat,A., De La Cruz,N., Olsen,A., Thi,C., Silver,S., Akther,S., Chowdhury,M., Omoregie,E. and Hughes,S. |
| EPI_ISL_17086274, EPI_ISL_17086275, EPI_ISL_17086276, EPI_ISL_17086277, EPI_ISL_17086278, EPI_ISL_17086279, EPI_ISL_17086280, EPI_ISL_17086281, EPI_ISL_17086282, EPI_ISL_17086283, EPI_ISL_17086284, EPI_ISL_17086285, EPI_ISL_17086286, EPI_ISL_17086287, EPI_ISL_17086288, EPI_ISL_17086289, EPI_ISL_17086290, EPI_ISL_17086291, EPI_ISL_17086292, EPI_ISL_17086293, EPI_ISL_17086294, EPI_ISL_17086295, EPI_ISL_17086296, EPI_ISL_17086297, EPI_ISL_17086298, EPI_ISL_17086299, EPI_ISL_17086300, EPI_ISL_17086301, EPI_ISL_17086302, EPI_ISL_17086304, EPI_ISL_17086305, EPI_ISL_17086306, EPI_ISL_17086307, EPI_ISL_17086308, EPI_ISL_17086309, EPI_ISL_17086310, EPI_ISL_17086311, EPI_ISL_17086312, EPI_ISL_17086313, EPI_ISL_17086314, EPI_ISL_17086315, EPI_ISL_17086316, EPI_ISL_17086317, EPI_ISL_17086318, EPI_ISL_17086319, EPI_ISL_17086320, EPI_ISL_17086321, EPI_ISL_17086322, EPI_ISL_17086323, EPI_ISL_17086324, EPI_ISL_17086325, EPI_ISL_17086326, EPI_ISL_17086327, EPI_ISL_17086328, EPI_ISL_17086329, EPI_ISL_17086330, EPI_ISL_17086331, EPI_ISL_17086332, EPI_ISL_17104180, EPI_ISL_17104181, EPI_ISL_17104182, EPI_ISL_17104183, EPI_ISL_17104184, EPI_ISL_17104185, EPI_ISL_17104186, EPI_ISL_17104188, EPI_ISL_17104189, EPI_ISL_17104190, EPI_ISL_17104191, EPI_ISL_17104192, EPI_ISL_17104193, EPI_ISL_17104194, EPI_ISL_17104195, EPI_ISL_17104196, EPI_ISL_17104197, EPI_ISL_17104198, EPI_ISL_17104200, EPI_ISL_17104201, EPI_ISL_17104202, EPI_ISL_17104203, EPI_ISL_17104204, EPI_ISL_17104205, EPI_ISL_17104206, EPI_ISL_17104207, EPI_ISL_17104208, EPI_ISL_17104209, EPI_ISL_17104210, EPI_ISL_17104212, EPI_ISL_17104213, EPI_ISL_17104214, EPI_ISL_17104215, EPI_ISL_17104216, EPI_ISL_17104217, EPI_ISL_17104218, EPI_ISL_17104219, EPI_ISL_17104220, EPI_ISL_17104221, EPI_ISL_17104222, EPI_ISL_17104224, EPI_ISL_17104225, EPI_ISL_17104226, EPI_ISL_17104227, EPI_ISL_17104228, EPI_ISL_17104229, EPI_ISL_17104230, EPI_ISL_17104231, EPI_ISL_17104232, EPI_ISL_17104233, EPI_ISL_17104235, EPI_ISL_17104236, EPI_ISL_17104237, EPI_ISL_17104238, EPI_ISL_17104239, EPI_ISL_17104240, EPI_ISL_17104241, EPI_ISL_17104242, EPI_ISL_17104243, EPI_ISL_17104244, EPI_ISL_17104245, EPI_ISL_17104247, EPI_ISL_17104248, EPI_ISL_17104249, EPI_ISL_17104250, EPI_ISL_17104251, EPI_ISL_17104252, EPI_ISL_17104253, EPI_ISL_17104254, EPI_ISL_17104255, EPI_ISL_17104256, EPI_ISL_17104257, EPI_ISL_17104259, EPI_ISL_17104260, EPI_ISL_17104261, EPI_ISL_17104262, EPI_ISL_17104263, EPI_ISL_17104264, EPI_ISL_17104265, EPI_ISL_17104266, EPI_ISL_17104267, EPI_ISL_17104268, EPI_ISL_17104269, EPI_ISL_17104271, EPI_ISL_17104272, EPI_ISL_17104273, EPI_ISL_17104274, EPI_ISL_17104275, EPI_ISL_17104276, EPI_ISL_17104277, EPI_ISL_17104278, EPI_ISL_17104279, EPI_ISL_17104280, EPI_ISL_17104281, EPI_ISL_17104282, EPI_ISL_17104284, EPI_ISL_17104285, EPI_ISL_17104286, EPI_ISL_17104287, EPI_ISL_17104288, EPI_ISL_17104350, EPI_ISL_17104352, EPI_ISL_17104353, EPI_ISL_17104354, EPI_ISL_17104355, EPI_ISL_17104356, EPI_ISL_17104357, EPI_ISL_17104358, EPI_ISL_17104359, EPI_ISL_17104360, EPI_ISL_17104361, EPI_ISL_17104362, EPI_ISL_17104364, EPI_ISL_17104365, EPI_ISL_17104366, EPI_ISL_17104367, EPI_ISL_17104368, EPI_ISL_17104369, EPI_ISL_17104370, EPI_ISL_17104371, EPI_ISL_17104372, EPI_ISL_17104373, EPI_ISL_17104374, EPI_ISL_17104375, EPI_ISL_17104377, EPI_ISL_17104378, EPI_ISL_17104379, EPI_ISL_17104380, EPI_ISL_17104381, EPI_ISL_17104382, EPI_ISL_17104383, EPI_ISL_17104384, EPI_ISL_17104385, EPI_ISL_17104386, EPI_ISL_17104387, EPI_ISL_17104389, EPI_ISL_17104390, EPI_ISL_17104391, EPI_ISL_17104392, EPI_ISL_17104393, EPI_ISL_17104394, EPI_ISL_17104395, EPI_ISL_17104396, EPI_ISL_17104397, EPI_ISL_17104398, EPI_ISL_17104399, EPI_ISL_17104401, EPI_ISL_17104402, EPI_ISL_17104403, EPI_ISL_17104404, EPI_ISL_17104405, EPI_ISL_17104406, EPI_ISL_17104407, EPI_ISL_17104408, EPI_ISL_17104409, EPI_ISL_17104410, EPI_ISL_17104412, EPI_ISL_17104413, EPI_ISL_17104414, EPI_ISL_17104415, EPI_ISL_17104416, EPI_ISL_17104417, EPI_ISL_17104418, EPI_ISL_17104419, EPI_ISL_17104420, EPI_ISL_17104421, EPI_ISL_17104422, EPI_ISL_17104423, EPI_ISL_17104425, EPI_ISL_17104426, EPI_ISL_17104427, EPI_ISL_17104428, EPI_ISL_17104429, EPI_ISL_17104430, EPI_ISL_17104431, EPI_ISL_17104432, EPI_ISL_17104433, EPI_ISL_17104434, EPI_ISL_17104435, EPI_ISL_17104437, EPI_ISL_17104438, EPI_ISL_17104439, EPI_ISL_17104440, EPI_ISL_17104441, EPI_ISL_17104442, EPI_ISL_17104443, EPI_ISL_17104444, EPI_ISL_17104445, EPI_ISL_17104446, EPI_ISL_17104447, EPI_ISL_17104448, EPI_ISL_17104450, EPI_ISL_17104451, EPI_ISL_17104452, EPI_ISL_17104453, EPI_ISL_17104454, EPI_ISL_17104455, EPI_ISL_17104456, EPI_ISL_17104457, EPI_ISL_17104458, EPI_ISL_17104490, EPI_ISL_17104491, EPI_ISL_17104492, EPI_ISL_17104493, EPI_ISL_17104494, EPI_ISL_17104496, EPI_ISL_17104497, EPI_ISL_17104498, EPI_ISL_17104499, EPI_ISL_17104500, EPI_ISL_17104501, EPI_ISL_17104502, EPI_ISL_17104503, EPI_ISL_17104504, EPI_ISL_17104506, EPI_ISL_17104507, EPI_ISL_17104508, EPI_ISL_17104509, EPI_ISL_17104510, EPI_ISL_17104511, EPI_ISL_17104512, EPI_ISL_17104513, EPI_ISL_17104514, EPI_ISL_17104515, EPI_ISL_17104516, EPI_ISL_17104517, EPI_ISL_17104518, EPI_ISL_17104520, EPI_ISL_17104521, EPI_ISL_17104522, EPI_ISL_17104523, EPI_ISL_17104524, EPI_ISL_17104525, EPI_ISL_17104526, EPI_ISL_17104527, EPI_ISL_17104528, EPI_ISL_17104529, EPI_ISL_17104530, EPI_ISL_17104531, EPI_ISL_17104534, EPI_ISL_17104535, EPI_ISL_17104536, EPI_ISL_17104537, EPI_ISL_17104538, EPI_ISL_17104539, EPI_ISL_17104540, EPI_ISL_17104541, EPI_ISL_17104542, EPI_ISL_17104543, EPI_ISL_17104545, EPI_ISL_17104546, EPI_ISL_17104547, EPI_ISL_17104548, EPI_ISL_17104549, EPI_ISL_17104551, EPI_ISL_17104552, EPI_ISL_17104553, EPI_ISL_17104554, EPI_ISL_17104555, EPI_ISL_17104556, EPI_ISL_17104557, EPI_ISL_17104559, EPI_ISL_17104560, EPI_ISL_17104561, EPI_ISL_17104562, EPI_ISL_17104563, EPI_ISL_17104564, EPI_ISL_17104565, EPI_ISL_17104566, EPI_ISL_17104567, EPI_ISL_17104568, EPI_ISL_17104569, EPI_ISL_17104570, EPI_ISL_17104572, EPI_ISL_17104573, EPI_ISL_17104574, EPI_ISL_17104575, EPI_ISL_17104576, EPI_ISL_17104577, EPI_ISL_17104578, EPI_ISL_17104579, EPI_ISL_17104580, EPI_ISL_17104581, EPI_ISL_17104582, EPI_ISL_17104583, EPI_ISL_17104585, EPI_ISL_17104586, EPI_ISL_17104587, EPI_ISL_17104588, EPI_ISL_17104589, EPI_ISL_17104590, EPI_ISL_17104591, EPI_ISL_17104592, EPI_ISL_17104593, EPI_ISL_17104594, EPI_ISL_17104595, EPI_ISL_17104597, EPI_ISL_17104598, EPI_ISL_17104599, EPI_ISL_17104613, EPI_ISL_17104614, EPI_ISL_17104615, EPI_ISL_17104616, EPI_ISL_17104617, EPI_ISL_17104618, EPI_ISL_17104619, EPI_ISL_17104620, EPI_ISL_17104621, EPI_ISL_17104622, EPI_ISL_17104623, EPI_ISL_17104625, EPI_ISL_17104626, EPI_ISL_17104627, EPI_ISL_17104628, EPI_ISL_17104629, EPI_ISL_17104630, EPI_ISL_17104631, EPI_ISL_17104632, EPI_ISL_17104634, EPI_ISL_17104635, EPI_ISL_17104637, EPI_ISL_17104638, EPI_ISL_17104639, EPI_ISL_17104640, EPI_ISL_17104641, EPI_ISL_17104642, EPI_ISL_17104643, EPI_ISL_17104644, EPI_ISL_17104645, EPI_ISL_17104646, EPI_ISL_17104647, EPI_ISL_17104648, EPI_ISL_17104649, EPI_ISL_17104650, EPI_ISL_17104651, EPI_ISL_17104652, EPI_ISL_17104653, EPI_ISL_17104655, EPI_ISL_17104656, EPI_ISL_17104657 | | | |
| see above | Public Health Laboratory, NYC Department of Health and Mental Hygiene | Public Health Laboratory, NYC Department of Health and Mental Hygiene | Clabby,T.T., Amin,H.S., Wang,J.C., Taki,F., Su,M., Rahat,A., De La Cruz,N., Olsen,A., Thi,C., Silver,S., Akther,S., Chowdhury,M., Omoregie,E. and Hughes,S. |
| EPI_ISL_17118598, EPI_ISL_17118599, EPI_ISL_17118600, EPI_ISL_17118601, EPI_ISL_17118602, EPI_ISL_17118603, EPI_ISL_17118604, EPI_ISL_17118605, EPI_ISL_17118606, EPI_ISL_17118607, EPI_ISL_17118608, EPI_ISL_17118609, EPI_ISL_17118610, EPI_ISL_17118611, EPI_ISL_17118612, EPI_ISL_17118613, EPI_ISL_17118614, EPI_ISL_17118615, EPI_ISL_17118616, EPI_ISL_17118617, EPI_ISL_17118618, EPI_ISL_17118619, EPI_ISL_17118620, EPI_ISL_17118621, EPI_ISL_17118622, EPI_ISL_17118623, EPI_ISL_17118624, EPI_ISL_17118625, EPI_ISL_17118626, EPI_ISL_17118627, EPI_ISL_17118628, EPI_ISL_17118629, EPI_ISL_17118630, EPI_ISL_17118631, EPI_ISL_17118632, EPI_ISL_17118633, EPI_ISL_17118634, EPI_ISL_17118635, EPI_ISL_17118636, EPI_ISL_17118637, EPI_ISL_17118638, EPI_ISL_17118639, EPI_ISL_17118640, EPI_ISL_17118641, EPI_ISL_17118642, EPI_ISL_17118643, EPI_ISL_17118644, EPI_ISL_17118645, EPI_ISL_17118646, EPI_ISL_17118647, EPI_ISL_17118648, EPI_ISL_17118649, EPI_ISL_17118650, EPI_ISL_17118651, EPI_ISL_17118652, EPI_ISL_17118653, EPI_ISL_17118654, EPI_ISL_17118655, EPI_ISL_17118656, EPI_ISL_17118657, EPI_ISL_17118658, EPI_ISL_17118659, EPI_ISL_17118660, EPI_ISL_17118661, EPI_ISL_17118662, EPI_ISL_17118663, EPI_ISL_17118664, EPI_ISL_17118665, EPI_ISL_17118666, EPI_ISL_17118667, EPI_ISL_17118668, EPI_ISL_17118669, EPI_ISL_17118670, EPI_ISL_17118671, EPI_ISL_17118672, EPI_ISL_17118673, EPI_ISL_17118674, EPI_ISL_17118675, EPI_ISL_17118676, EPI_ISL_17118677, EPI_ISL_17118678, EPI_ISL_17118679, EPI_ISL_17118680, EPI_ISL_17118681, EPI_ISL_17118682, EPI_ISL_17118683, EPI_ISL_17118684, EPI_ISL_17118685, EPI_ISL_17118686, EPI_ISL_17118687, EPI_ISL_17118688, EPI_ISL_17118689, EPI_ISL_17118690, EPI_ISL_17118691, EPI_ISL_17118692, EPI_ISL_17118693, EPI_ISL_17118694, EPI_ISL_17118695, EPI_ISL_17118696, EPI_ISL_17118697 | | | |
| see above | Public Health Laboratory, NYC Department of Health and Mental Hygiene | Public Health Laboratory, NYC Department of Health and Mental Hygiene | Wang,J.C., Amin,H.S., Clabby,T.T., Taki,F., Su,M., Rahat,A., De La Cruz,N., Olsen,A., Thi,C., Silver,S., Akther,S., Chowdhury,M., Omoregie,E. and Hughes,S. |
| EPI_ISL_17165679, EPI_ISL_17165680, EPI_ISL_17165681, EPI_ISL_17165682, EPI_ISL_17165683, EPI_ISL_17165684, EPI_ISL_17165685, EPI_ISL_17165686, EPI_ISL_17165687, EPI_ISL_17165688, EPI_ISL_17165689, EPI_ISL_17165690, EPI_ISL_17165691, EPI_ISL_17165692, EPI_ISL_17165693, EPI_ISL_17165694, EPI_ISL_17165695, EPI_ISL_17165696, EPI_ISL_17165697, EPI_ISL_17165698, EPI_ISL_17165699, EPI_ISL_17165700, EPI_ISL_17165701, EPI_ISL_17165702, EPI_ISL_17165703, EPI_ISL_17165704, EPI_ISL_17165705, EPI_ISL_17165706, EPI_ISL_17165707, EPI_ISL_17165708, EPI_ISL_17165709, EPI_ISL_17165710, EPI_ISL_17165711, EPI_ISL_17165712, EPI_ISL_17165713, EPI_ISL_17165714, EPI_ISL_17165715, EPI_ISL_17165716, EPI_ISL_17165717, EPI_ISL_17165718, EPI_ISL_17165719, EPI_ISL_17165720, EPI_ISL_17165721, EPI_ISL_17165722, EPI_ISL_17165723, EPI_ISL_17165724, EPI_ISL_17165725, EPI_ISL_17165726, EPI_ISL_17165727, EPI_ISL_17165728, EPI_ISL_17165729, EPI_ISL_17165730, EPI_ISL_17165731, EPI_ISL_17165732, EPI_ISL_17165733, EPI_ISL_17165734, EPI_ISL_17165735, EPI_ISL_17165736, EPI_ISL_17165737, EPI_ISL_17165738, EPI_ISL_17165739, EPI_ISL_17165740, EPI_ISL_17165741, EPI_ISL_17165742, EPI_ISL_17165743, EPI_ISL_17165744, EPI_ISL_17165745, EPI_ISL_17165746, EPI_ISL_17165747, EPI_ISL_17165748, EPI_ISL_17165749, EPI_ISL_17165750, EPI_ISL_17165751, EPI_ISL_17165752, EPI_ISL_17165753, EPI_ISL_17165754, EPI_ISL_17165755, EPI_ISL_17165756, EPI_ISL_17165757, EPI_ISL_17165758, EPI_ISL_17165759, EPI_ISL_17165760, EPI_ISL_17165761, EPI_ISL_17165762, EPI_ISL_17165763, EPI_ISL_17165764, EPI_ISL_17165765, EPI_ISL_17165766, EPI_ISL_17165767, EPI_ISL_17165768, EPI_ISL_17165769, EPI_ISL_17165770, EPI_ISL_17165771, EPI_ISL_17165772, EPI_ISL_17165773, EPI_ISL_17165774, EPI_ISL_17165775, EPI_ISL_17165776, EPI_ISL_17165777, EPI_ISL_17165778 | | | |
| see above | Public Health Laboratory, NYC Department of Health and Mental Hygiene (DOHMH) | Public Health Laboratory, NYC Department of Health and Mental Hygiene (DOHMH) | Clabby,T.T., Amin,H.S., Wang,J.C., Taki,F., Su,M., Rahat,A., De La Cruz,N., Olsen,A., Thi,C., Silver,S., Akther,S., Chowdhury,M., Omoregie,E. and Hughes,S. |
| EPI_ISL_17170656, EPI_ISL_17170657, EPI_ISL_17170658, EPI_ISL_17170659, EPI_ISL_17170660, EPI_ISL_17170661, EPI_ISL_17170662, EPI_ISL_17170663, EPI_ISL_17170664, EPI_ISL_17170665, EPI_ISL_17170666, EPI_ISL_17170667, EPI_ISL_17170668, EPI_ISL_17170669, EPI_ISL_17170670, EPI_ISL_17170671 | | | |
| see above | California Department of Public Health | California Department of Public Health | Haw, M. , Kath, C., Espinosa, A., O'Neil, R., and Hacker, J. |
| EPI_ISL_17179627, EPI_ISL_17179628 | St Jame's Hospital, Virology Department | National Virus Reference Laboratory | Patrice Keane, Yvonne Lynagh, Brendan Crowley, Gabriel Gonzalez, Michael Carr, Emer O'Byrne, Weronika Banka, Brian Keogan, Jose Maria Urtasun Elizari, Jonathan Dean, Daniel Hare, Cillian F De Gascun |
| EPI_ISL_17179629, EPI_ISL_17179630, EPI_ISL_17179631, EPI_ISL_17179632 | National Virus Reference Laboratory | National Virus Reference Laboratory | Gabriel Gonzalez, Michael Carr, Emer O'Byrne, Weronika Banka, Brian Keogan, Jose Maria Urtasun Elizari, Jonathan Dean, Daniel Hare, Cillian F De Gascun |
| EPI_ISL_17179633 | St Jame's Hospital, Virology Department | National Virus Reference Laboratory | Patrice Keane, Yvonne Lynagh, Brendan Crowley, Gabriel Gonzalez, Michael Carr, Emer O'Byrne, Weronika Banka, Brian Keogan, Jose Maria Urtasun Elizari, Jonathan Dean, Daniel Hare, Cillian F De Gascun |

| | | | |
|---|---|---|---|
| EPI_ISL_17179634, EPI_ISL_17179635, EPI_ISL_17179636, EPI_ISL_17179637 | National Virus Reference Laboratory | National Virus Reference Laboratory | Gabriel Gonzalez, Michael Carr, Emer O'Byrne, Weronika Banka, Brian Keogan, Jose Maria Urtasun Elizari, Jonathan Dean, Daniel Hare, Cillian F De Gascun |
| EPI_ISL_17179638, EPI_ISL_17179639, EPI_ISL_17179640, EPI_ISL_17179641, EPI_ISL_17179642, EPI_ISL_17179643 | St Jame's Hospital, Virology Department | National Virus Reference Laboratory | Patrice Keane, Yvonne Lynagh, Brendan Crowley, Gabriel Gonzalez, Michael Carr, Emer O'Byrne, Weronika Banka, Brian Keogan, Jose Maria Urtasun Elizari, Jonathan Dean, Daniel Hare, Cillian F De Gascun |
| EPI_ISL_17187497, EPI_ISL_17187498 | Vajira Hospital | Thai Red Cross Emerging Infectious Diseases Clinical Center and Faculty of Medicine, Chulalongkorn University | Suppasit srisaeng, Praepoly Ruekmuang, Kusuma Swangpun, Arriya Panchaiyaphum, Pakita Salaeh, Natpusda Kongmaung, Pornsiri Limwattanawong, Noree Pholprasert, Montriya Unteamsom, Kanjana Jeknok, Withak Withaksabut, Sunisa Nilda, Artorn Niakul, Sopon Iamsirithaworn, Thitipong Yingyong, Rossaporn Kittiyaowamarn, Rome Buathong, Ratanaporn Tangwangvivat, Supaporn Wacharapluesadee, Sininat Petcharat, Ananporn Supataragul, Stefan Fernandez, Achawin Rojanaviwat, Chonticha Klungthong, Pilailuk Okada, Khajohn Joonlasak, Chakkarat Pitayawonganon, Opass Putcharoen |
| EPI_ISL_17187499 | Department of Disease Control, Ministry of Public Health | Thai Red Cross Emerging Infectious Diseases Clinical Center and Faculty of Medicine, Chulalongkorn University | Suppasit srisaeng, Praepoly Ruekmuang, Kusuma Swangpun, Arriya Panchaiyaphum, Pakita Salaeh, Natpusda Kongmaung, Pornsiri Limwattanawong, Noree Pholprasert, Montriya Unteamsom, Kanjana Jeknok, Withak Withaksabut, Sunisa Nilda, Artorn Niakul, Sopon Iamsirithaworn, Thitipong Yingyong, Rossaporn Kittiyaowamarn, Rome Buathong, Ratanaporn Tangwangvivat, Supaporn Wacharapluesadee, Sininat Petcharat, Ananporn Supataragul, Stefan Fernandez, Achawin Rojanaviwat, Chonticha Klungthong, Pilailuk Okada, Khajohn Joonlasak, Chakkarat Pitayawonganon, Opass Putcharoen |
| EPI_ISL_17187500 | Bangkok Hospital Phuket | Thai Red Cross Emerging Infectious Diseases Clinical Center and Faculty of Medicine, Chulalongkorn University | Nungrathai Srisong, Praepoly Ruekmuang, Kusuma Swangpun, Arriya Panchaiyaphum, Pakita Salaeh, Natpusda Kongmaung, Pornsiri Limwattanawong, Noree Pholprasert, Montriya Unteamsom, Kanjana Jeknok, Withak Withaksabut, Sunisa Nilda, Artorn Niakul, Sopon Iamsirithaworn, Thitipong Yingyong, Rossaporn Kittiyaowamarn, Rome Buathong, Ratanaporn Tangwangvivat, Supaporn Wacharapluesadee, Sininat Petcharat, Ananporn Supataragul, Stefan Fernandez, Achawin Rojanaviwat, Chonticha Klungthong, Pilailuk Okada, Khajohn Joonlasak, Chakkarat Pitayawonganon, Opass Putcharoen |
| EPI_ISL_17187502 | Department of Disease Control, Ministry of Public Health | Thai Red Cross Emerging Infectious Diseases Clinical Center and Faculty of Medicine, Chulalongkorn University | Supanut Chotichavalrattanakul, , Praepoly Ruekmuang, Kusuma Swangpun, Arriya Panchaiyaphum, Pakita Salaeh, Natpusda Kongmaung, Pornsiri Limwattanawong, Noree Pholprasert, Montriya Unteamsom, Kanjana Jeknok, Withak Withaksabut, Sunisa Nilda, Artorn Niakul, Sopon Iamsirithaworn, Thitipong Yingyong, Rossaporn Kittiyaowamarn, Rome Buathong, Ratanaporn Tangwangvivat, Supaporn Wacharapluesadee, Sininat Petcharat, Ananporn Supataragul, Stefan Fernandez, Achawin Rojanaviwat, Chonticha Klungthong, Pilailuk Okada, Khajohn Joonlasak, Chakkarat Pitayawonganon, Opass Putcharoen |
| EPI_ISL_17187504 | Suvarnabhumi Airport | Thai Red Cross Emerging Infectious Diseases Clinical Center and Faculty of Medicine, Chulalongkorn University | Phawinee Montri, Praepoly Ruekmuang, Kusuma Swangpun, Arriya Panchaiyaphum, Pakita Salaeh, Natpusda Kongmaung, Pornsiri Limwattanawong, Noree Pholprasert, Montriya Unteamsom, Kanjana Jeknok, Withak Withaksabut, Sunisa Nilda, Artorn Niakul, Sopon Iamsirithaworn, Thitipong Yingyong, Rossaporn Kittiyaowamarn, Rome Buathong, Ratanaporn Tangwangvivat, Supaporn Wacharapluesadee, Sininat Petcharat, Ananporn Supataragul, Stefan Fernandez, Achawin Rojanaviwat, Chonticha Klungthong, Pilailuk Okada, Khajohn Joonlasak, Chakkarat Pitayawonganon, Opass Putcharoen |
| EPI_ISL_17206607, EPI_ISL_17206608, EPI_ISL_17206609, EPI_ISL_17206610, EPI_ISL_17206611, EPI_ISL_17206612, EPI_ISL_17206613, EPI_ISL_17206614, EPI_ISL_17206615, EPI_ISL_17206616, EPI_ISL_17206617, EPI_ISL_17206618, EPI_ISL_17206619, EPI_ISL_17206620 | | | |
| see above | California Department of Public Health | California Department of Public Health | Haw,M., Kath,C., Espinosa,A., O'Neil,R., and Hacker,J. |
| EPI_ISL_17206621, EPI_ISL_17206622 | California Department of Public Health | California Department of Public Health | Kath, C., Haw, M., Espinosa, A., and Hacker, J. |
| EPI_ISL_17211324 | Kaiser Permanente Chino Hills Regional Reference Laboratories | Los Angeles County Public Health Laboratories | P. Hemarajata et al. |
| EPI_ISL_17211331 | Los Angeles County Public Health Laboratories | Los Angeles County Public Health Laboratories | P. Hemarajata et al. |
| EPI_ISL_17211335 | Laboratory Corporation of America | Los Angeles County Public Health Laboratories | P. Hemarajata et al. |
| EPI_ISL_17271956, EPI_ISL_17271957 | Rhode Island State Health Laboratory | Rhode Island State Health Laboratory | Kristin Carpenter-Azevedo, Sean Sierra-Patev, Richard C. Huard |
| EPI_ISL_17390796, EPI_ISL_17390797, EPI_ISL_17390798, EPI_ISL_17390799, EPI_ISL_17390800, EPI_ISL_17390801, EPI_ISL_17390802, EPI_ISL_17390803, EPI_ISL_17390804, EPI_ISL_17390807 | Antioquia, Laboratorio Departamental de Salud Publica de Antioquia | Antioquia, Laboratorio Departamental de Salud Publica de Antioquia | Betancur,I.I.B., Velarde Hoyos,C.A.C.V., Gomez,R.R.G. and Mercado-Reyes,M.M.R. |
| EPI_ISL_17406093, EPI_ISL_17406094, EPI_ISL_17406095, EPI_ISL_17406096, EPI_ISL_17406097, EPI_ISL_17406098, EPI_ISL_17406099, EPI_ISL_17406100, EPI_ISL_17406101, EPI_ISL_17406102, EPI_ISL_17406103, EPI_ISL_17406104, EPI_ISL_17406105, EPI_ISL_17406106, EPI_ISL_17406107, EPI_ISL_17406108, EPI_ISL_17406109, EPI_ISL_17406110, EPI_ISL_17406111, EPI_ISL_17406112, EPI_ISL_17406113, EPI_ISL_17406114, EPI_ISL_17406115, EPI_ISL_17406116, EPI_ISL_17406117, EPI_ISL_17406118, EPI_ISL_17406119, EPI_ISL_17406120, EPI_ISL_17406121, EPI_ISL_17406122, EPI_ISL_17406123, EPI_ISL_17406124 | | | |
| see above | CDCT/CEVS/SES-RS | CDCT/CEVS/SES-RS | Richard Steiner Salvato, Fernanda Marques Godinho, Regina Bones Barcellos, Patricia Sesterheim, Amanda Pellenz Ruivo, Viviane Horn de Melo, Júlio Augusto Schroder |
| EPI_ISL_17424656, EPI_ISL_17424657, EPI_ISL_17424658, EPI_ISL_17424659, EPI_ISL_17424660, EPI_ISL_17424661, EPI_ISL_17424662, EPI_ISL_17424663, EPI_ISL_17424664, EPI_ISL_17424665, EPI_ISL_17424666, EPI_ISL_17424667, EPI_ISL_17424668, EPI_ISL_17424669, EPI_ISL_17424670, EPI_ISL_17424671, EPI_ISL_17424672, EPI_ISL_17424673, EPI_ISL_17424674, EPI_ISL_17424675, EPI_ISL_17424676, EPI_ISL_17424677, EPI_ISL_17424678 | | | |
| see above | Molecular Microbiology Laboratory, Department of Pathology, Molecular and Cell-Based Medicine, Icahn School of Medicine at Mount Sinai, | Molecular Microbiology Laboratory, Department of Pathology, Molecular and Cell-Based Medicine, Icahn School of Medicine at Mount Sinai, | Luz H. Patiño, Susana Guerra, Marina Muñoz, Nicolas Luna , Keith Farrugia, Adriana van de Guchte, Zain Khalil , Ana Silvia Gonzalez-Reiche, Matthew M. Hernandez ,Radhika Banu, Paras Shrestha, Bernadette Liggayu, Adolfo Firpo Betancourt, David Reich, Carlos Cordon-Cardo, Randy Albrechtf, Rebecca Pearlf, Viviana Simona, Aria Rookera, Emilia Mia Sordillo, Harm van Bakeld, Adolfo García-Sastre, Dusan Bogunovic, Gustavo Palacios, Alberto Paniz Mondolfi, Juan David Ramírez |
| EPI_ISL_17445514, EPI_ISL_17445515, EPI_ISL_17445516, EPI_ISL_17445517, EPI_ISL_17445518, EPI_ISL_17445519 | Tokyo Metropolitan Institute of Public Health | Tokyo Metropolitan Institute of Public Health | Fumi Kasuya, Wakaba Okada, Ryota Kumagai, Sachiko Harada, Arisa Amano, Michiya Hasegawa, Mami Nagashima, Kenji Sadamasu |
| EPI_ISL_17471100, EPI_ISL_17471101, EPI_ISL_17471102, EPI_ISL_17471103, EPI_ISL_17471104, EPI_ISL_17471105, EPI_ISL_17471106, EPI_ISL_17471107, EPI_ISL_17471108, EPI_ISL_17471109, EPI_ISL_17471110 | | | |
| see above | Laboratorio de Enterovirus, Instituto Oswaldo Cruz, Fiocruz | Laboratory of Respiratory Viruses and Measles, Oswaldo Cruz Institute, FIOCRUZ | Paola Resende, Elisa Cavalcante Pereira, Bruna Mendonça da Silva, Jéssica Graça Macedo de Carvalho, Larissa Macedo Pinto, Victor Guimaraes, Marilda Siqueira, Renan da Silva Faustino, Marilia Santini, Beatriz Grinsztejn, Mayara Secco Torres da Silva, Edson Elias da Silva on behalf of the Fiocruz Genomic Surveillance Network |
| EPI_ISL_17485343 | Laboratorio de Enterovirus, Instituto Oswaldo Cruz, Fiocruz | Instituto Oswaldo Cruz FIOCRUZ - Laboratory of Respiratory Viruses and Measles (LVRS) | Paola Resende, Elisa Cavalcante Pereira, Bruna Mendonça da Silva, Jéssica Graça Macedo de Carvalho, Larissa Macedo Pinto, Victor Guimaraes, Marilda Siqueira, Renan da Silva Faustino, Marilia Santini, Beatriz Grinsztejn, Mayara Secco Torres da Silva, Edson Elias da Silva on behalf of the Fiocruz Genomic Surveillance Network |
| EPI_ISL_17502583 | Public Health Laboratory, Public Health Service Amsterdam, The Netherlands | Department of Medical Microbiology & Infection prevention, Amsterdam University Medical Centers location AMC | Matthijs Welkers, Jelle Koopsen, Robin van Houdt, Marcel Jonges, Sebastien Matamoros, Sjoerd Rebers, Fokla Zorgdrager, Sylvia Bruisten, Akke Cornelissen, Janke Schinkel, Ewout Fanoy, Roisin Bavalia, Menno de Jong and Mariken van der Lubben on behalf of the Amsterdam Regional Genomic epidemiology and Outbreak Surveillance (ARGOS) consortium |
| EPI_ISL_17518107 | Virology Section, Division of Microbiology,Osaka Institute of Public Health | Virology Section, Division of Microbiology,Osaka Institute of Public Health | Daiki Kanbayashi, Takako Kurata, Takuya Kawahata, Fumiya Bannno, Minami Hama, Kazushi Motomuta |
| EPI_ISL_17525484 | Division de Microbiologia, Hospital Nacional de Niños Carlos Saenz Herrera | Inciensa, Investigación y Enseñanza en Nutrición y Salud Centro Nacional de Referencia de Virología | Cristian Perez Corrales, Christopher Mairena Acuña, Diana Cantillo, Hillary Serrano, Ana Isela Ruiz, Gustavo Vega, Claudio Soto-Garita, Adriana Godinez, Estela Cordero, Melany Calderon, Francisco Duarte |
| EPI_ISL_17529367, EPI_ISL_17529368 | Laboratorio de Virus Exantematicos, Gastroentéricos y Otros Transmitidos por Vectores | Centro de Referencia Nacional de Genomica, Secuenciacion y Bioinformatica GENSBIO, INSPI-CZ9 | Andrés Carrazco*, SIlvia Salgado, Diana Gutiérrez, Damaris Alarcón, Andrés Tinizaray, Ruth Gómez, Martha Sánchez, Johanna Parrales, Eva Nicola, Jorge Bejarano, Leandro Patiño. |
| EPI_ISL_17536780 | Department of Virology, National Institute of Health, Islamabad, Pakistan | Department of Virology, National Institute of Health, Islamabad, Pakistan | Massab Umair, Muhammad Ammar, Syed Adnan Haider, Rabia Hakim, Qasim Malik, Muhammad Salman, Ghazala Parveen, and Naseem Akhtar |
| EPI_ISL_17536782, EPI_ISL_17536783, EPI_ISL_17536784, EPI_ISL_17536785 | Laboratorio de Enterovirus, Instituto Oswaldo Cruz, Fiocruz | Instituto Oswaldo Cruz FIOCRUZ - Laboratory of Respiratory Viruses and Measles (LVRS) | Paola Resende, Elisa Cavalcante Pereira, Bruna Mendonça da Silva, Jéssica Graça Macedo de Carvalho, Larissa Macedo Pinto, Victor Guimaraes, Marilda Siqueira, Renan da Silva Faustino, Marilia Santini, Edson Elias da Silva on behalf of the Fiocruz Genomic Surveillance Network |
| EPI_ISL_17582853 | Quest Diagnostics Nichols Institute | Los Angeles County Public Health Laboratories | P. Hemarajata et al. |
| EPI_ISL_17584292 | Centro Medico ABC | Instituto Nacional de Medicina Genomica | Cedro Tanda Alberto, Roxana Trejo González, Laura Gomez-Romero, Alfredo Mendoza-Vargas, Dora Garnica-Lopez, Alfredo Hidalgo-Miranda, Luis A Herrera. |
| EPI_ISL_17592665, EPI_ISL_17592666, EPI_ISL_17592667, EPI_ISL_17592668, EPI_ISL_17592669, EPI_ISL_17592670 | Tokyo Metropolitan Institute of Public Health | Tokyo Metropolitan Institute of Public Health | Fumi Kasuya, Wakaba Okada, Ryota Kumagai, Sachiko Harada, Arisa Amano, Michiya Hasegawa, Mami Nagashima, Kenji Sadamasu |
| EPI_ISL_17595305 | Kaiser Permanente Chino Hills Regional Reference Laboratories | Los Angeles County Public Health Laboratories | P. Hemarajata et al. |
| EPI_ISL_17614017, EPI_ISL_17614018, EPI_ISL_17614019, EPI_ISL_17614020, EPI_ISL_17614021, EPI_ISL_17614022, EPI_ISL_17614023, EPI_ISL_17614024, EPI_ISL_17614025, EPI_ISL_17614026, EPI_ISL_17614027, EPI_ISL_17614028, EPI_ISL_17614029, EPI_ISL_17614030, EPI_ISL_17614031, EPI_ISL_17614032, EPI_ISL_17614033, EPI_ISL_17614034, EPI_ISL_17614035, EPI_ISL_17614036, EPI_ISL_17614037, EPI_ISL_17614038, EPI_ISL_17614039, EPI_ISL_17614040, EPI_ISL_17614041, EPI_ISL_17614042, EPI_ISL_17614043, EPI_ISL_17614044, EPI_ISL_17614045, EPI_ISL_17614046, EPI_ISL_17614047, EPI_ISL_17614048, EPI_ISL_17614049 | | | |
| see above | Laboratorio de Enterovirus, Instituto Oswaldo Cruz, Fiocruz | Instituto Oswaldo Cruz FIOCRUZ - Laboratory of Respiratory Viruses and Measles (LVRS) | Paola Resende, Elisa Cavalcante Pereira, Bruna Mendonça da Silva, Jéssica Graça Macedo de Carvalho, Larissa Macedo Pinto, Victor Guimaraes, Marilda Siqueira, Renan da Silva Faustino, Marilia Santini, Edson Elias da Silva on behalf of the Fiocruz Genomic Surveillance Network |
| EPI_ISL_17665624, EPI_ISL_17665625, EPI_ISL_17665626, EPI_ISL_17665627 | Tokyo Metropolitan Institute of Public Health | Tokyo Metropolitan Institute of Public Health | Fumi Kasuya, Wakaba Okada, Ryota Kumagai, Sachiko Harada, Arisa Amano, Michiya Hasegawa, Mami Nagashima, Kenji Sadamasu |
| EPI_ISL_17672206 | LESP State of Mexico | Instituto de Diagnostico y Referencia Epidemiologicos (INDRE) | Abril Rodríguez-Maldonado; Claudia Wong-Arámbula; Silvia Rivero-Arredondo; Ruth Madera-Sandoval; Joaquín Quiroz-Mercado; Fernando González-Domínguez; Lucia Hernández-Rivas, Irma López-Martínez; Ernesto Ramírez-González; Maribel González-Villa |
| EPI_ISL_17672208 | LESP Queretaro | Instituto de Diagnostico y Referencia Epidemiologicos (INDRE) | Abril Rodríguez-Maldonado; Claudia Wong-Arámbula; Silvia Rivero-Arredondo; Ruth Madera-Sandoval; Joaquín Quiroz-Mercado; Fernando González-Domínguez; Lucia Hernández-Rivas, Irma López-Martínez; Ernesto Ramírez-González; Maribel González-Villa |
| EPI_ISL_17672209 | LESP Yucatan | Instituto de Diagnostico y Referencia Epidemiologicos (INDRE) | Abril Rodríguez-Maldonado; Claudia Wong-Arámbula; Silvia Rivero-Arredondo; Ruth Madera-Sandoval; Joaquín Quiroz-Mercado; Fernando González-Domínguez; Lucia Hernández-Rivas, Irma López-Martínez; Ernesto Ramírez-González; Maribel González-Villa |
| EPI_ISL_17672210 | LESP Quintana Roo | Instituto de Diagnostico y Referencia Epidemiologicos (INDRE) | Abril Rodríguez-Maldonado; Claudia Wong-Arámbula; Silvia Rivero-Arredondo; Ruth Madera-Sandoval; Joaquín Quiroz-Mercado; Fernando González-Domínguez; Lucia Hernández-Rivas, Irma López-Martínez; Ernesto Ramírez-González; Maribel González-Villa |
| EPI_ISL_17672211 | LESP Mexico City | Instituto de Diagnostico y Referencia Epidemiologicos (INDRE) | Abril Rodríguez-Maldonado; Claudia Wong-Arámbula; Silvia Rivero-Arredondo; Ruth Madera-Sandoval; Joaquín Quiroz-Mercado; Fernando González-Domínguez; Lucia Hernández-Rivas, Irma López-Martínez; Ernesto Ramírez-González; Maribel González-Villa |
| EPI_ISL_17672212 | LESP Tamaulipas | Instituto de Diagnostico y Referencia Epidemiologicos (INDRE) | Abril Rodríguez-Maldonado; Claudia Wong-Arámbula; Silvia Rivero-Arredondo; Ruth Madera-Sandoval; Joaquín Quiroz-Mercado; Fernando González-Domínguez; Lucia Hernández-Rivas, Irma López-Martínez; Ernesto Ramírez-González; Maribel González-Villa |
| EPI_ISL_17672213 | LESP Puebla | Instituto de Diagnostico y Referencia Epidemiologicos (INDRE) | Abril Rodríguez-Maldonado; Claudia Wong-Arámbula; Silvia Rivero-Arredondo; Ruth Madera-Sandoval; Joaquín Quiroz-Mercado; Fernando González-Domínguez; Lucia Hernández-Rivas, Irma López-Martínez; Ernesto Ramírez-González; Maribel González-Villa |
| EPI_ISL_17672214 | LESP Guerrero | Instituto de Diagnostico y Referencia Epidemiologicos (INDRE) | Abril Rodríguez-Maldonado; Claudia Wong-Arámbula; Silvia Rivero-Arredondo; Ruth Madera-Sandoval; Joaquín Quiroz-Mercado; Fernando González-Domínguez; Lucia Hernández-Rivas, Irma López-Martínez; Ernesto Ramírez-González; Maribel González-Villa |

| | | | |
|---|---|---|---|
| EPI_ISL_17672217 | LESP Zacatecas | Instituto de Diagnostico y Referencia Epidemiologicos (INDRE) | Abril Rodríguez-Maldonado; Claudia Wong-Arámbula; Silvia Rivero-Arredondo; Ruth Madera-Sandoval; Joaquín Quiroz-Mercado; Fernando González-Domínguez; Lucia Hernández-Rivas, Irma López-Martínez; Ernesto Ramírez-González; Maribel González-Villa |
| EPI_ISL_17672218 | LESP Tabasco | Instituto de Diagnostico y Referencia Epidemiologicos (INDRE) | Abril Rodríguez-Maldonado; Claudia Wong-Arámbula; Silvia Rivero-Arredondo; Ruth Madera-Sandoval; Joaquín Quiroz-Mercado; Fernando González-Domínguez; Lucia Hernández-Rivas, Irma López-Martínez; Ernesto Ramírez-González; Maribel González-Villa |
| EPI_ISL_17672221 | LESP Chihuahua | Instituto de Diagnostico y Referencia Epidemiologicos (INDRE) | Abril Rodríguez-Maldonado; Claudia Wong-Arámbula; Silvia Rivero-Arredondo; Ruth Madera-Sandoval; Joaquín Quiroz-Mercado; Fernando González-Domínguez; Lucia Hernández-Rivas, Irma López-Martínez; Ernesto Ramírez-González; Maribel González-Villa |
| EPI_ISL_17672222 | LESP Veracruz | Instituto de Diagnostico y Referencia Epidemiologicos (INDRE) | Abril Rodríguez-Maldonado; Claudia Wong-Arámbula; Silvia Rivero-Arredondo; Ruth Madera-Sandoval; Joaquín Quiroz-Mercado; Fernando González-Domínguez; Lucia Hernández-Rivas, Irma López-Martínez; Ernesto Ramírez-González; Maribel González-Villa |
| EPI_ISL_17672223 | LESP Baja California | Instituto de Diagnostico y Referencia Epidemiologicos (INDRE) | Abril Rodríguez-Maldonado; Claudia Wong-Arámbula; Silvia Rivero-Arredondo; Ruth Madera-Sandoval; Joaquín Quiroz-Mercado; Fernando González-Domínguez; Lucia Hernández-Rivas, Irma López-Martínez; Ernesto Ramírez-González; Maribel González-Villa |
| EPI_ISL_17672224 | LESP Tamaulipas | Instituto de Diagnostico y Referencia Epidemiologicos (INDRE) | Abril Rodríguez-Maldonado; Claudia Wong-Arámbula; Silvia Rivero-Arredondo; Ruth Madera-Sandoval; Joaquín Quiroz-Mercado; Fernando González-Domínguez; Lucia Hernández-Rivas, Irma López-Martínez; Ernesto Ramírez-González; Maribel González-Villa |
| EPI_ISL_17672225 | LESP Campeche | Instituto de Diagnostico y Referencia Epidemiologicos (INDRE) | Abril Rodríguez-Maldonado; Claudia Wong-Arámbula; Silvia Rivero-Arredondo; Ruth Madera-Sandoval; Joaquín Quiroz-Mercado; Fernando González-Domínguez; Lucia Hernández-Rivas, Irma López-Martínez; Ernesto Ramírez-González; Maribel González-Villa |
| EPI_ISL_17672226 | LESP Hidalgo | Instituto de Diagnostico y Referencia Epidemiologicos (INDRE) | Abril Rodríguez-Maldonado; Claudia Wong-Arámbula; Silvia Rivero-Arredondo; Ruth Madera-Sandoval; Joaquín Quiroz-Mercado; Fernando González-Domínguez; Lucia Hernández-Rivas, Irma López-Martínez; Ernesto Ramírez-González; Maribel González-Villa |
| EPI_ISL_17672227 | LESP Mexico City | Instituto de Diagnostico y Referencia Epidemiologicos (INDRE) | Abril Rodríguez-Maldonado; Claudia Wong-Arámbula; Silvia Rivero-Arredondo; Ruth Madera-Sandoval; Joaquín Quiroz-Mercado; Fernando González-Domínguez; Lucia Hernández-Rivas, Irma López-Martínez; Ernesto Ramírez-González; Maribel González-Villa |
| EPI_ISL_17672228 | LESP Guerrero | Instituto de Diagnostico y Referencia Epidemiologicos (INDRE) | Abril Rodríguez-Maldonado; Claudia Wong-Arámbula; Silvia Rivero-Arredondo; Ruth Madera-Sandoval; Joaquín Quiroz-Mercado; Fernando González-Domínguez; Lucia Hernández-Rivas, Irma López-Martínez; Ernesto Ramírez-González; Maribel González-Villa |
| EPI_ISL_17672229 | LESP Coahuila | Instituto de Diagnostico y Referencia Epidemiologicos (INDRE) | Abril Rodríguez-Maldonado; Claudia Wong-Arámbula; Silvia Rivero-Arredondo; Ruth Madera-Sandoval; Joaquín Quiroz-Mercado; Fernando González-Domínguez; Lucia Hernández-Rivas, Irma López-Martínez; Ernesto Ramírez-González; Maribel González-Villa |
| EPI_ISL_17672230 | LESP Tabasco | Instituto de Diagnostico y Referencia Epidemiologicos (INDRE) | Abril Rodríguez-Maldonado; Claudia Wong-Arámbula; Silvia Rivero-Arredondo; Ruth Madera-Sandoval; Joaquín Quiroz-Mercado; Fernando González-Domínguez; Lucia Hernández-Rivas, Irma López-Martínez; Ernesto Ramírez-González; Maribel González-Villa |
| EPI_ISL_17672231 | LESP Chiapas | Instituto de Diagnostico y Referencia Epidemiologicos (INDRE) | Abril Rodríguez-Maldonado; Claudia Wong-Arámbula; Silvia Rivero-Arredondo; Ruth Madera-Sandoval; Joaquín Quiroz-Mercado; Fernando González-Domínguez; Lucia Hernández-Rivas, Irma López-Martínez; Ernesto Ramírez-González; Maribel González-Villa |
| EPI_ISL_17672232 | LESP Yucatan | Instituto de Diagnostico y Referencia Epidemiologicos (INDRE) | Abril Rodríguez-Maldonado; Claudia Wong-Arámbula; Silvia Rivero-Arredondo; Ruth Madera-Sandoval; Joaquín Quiroz-Mercado; Fernando González-Domínguez; Lucia Hernández-Rivas, Irma López-Martínez; Ernesto Ramírez-González; Maribel González-Villa |
| EPI_ISL_17672233 | LESP Quintana Roo | Instituto de Diagnostico y Referencia Epidemiologicos (INDRE) | Abril Rodríguez-Maldonado; Claudia Wong-Arámbula; Silvia Rivero-Arredondo; Ruth Madera-Sandoval; Joaquín Quiroz-Mercado; Fernando González-Domínguez; Lucia Hernández-Rivas, Irma López-Martínez; Ernesto Ramírez-González; Maribel González-Villa |
| EPI_ISL_17672234 | LESP Jalisco | Instituto de Diagnostico y Referencia Epidemiologicos (INDRE) | Abril Rodríguez-Maldonado; Claudia Wong-Arámbula; Silvia Rivero-Arredondo; Ruth Madera-Sandoval; Joaquín Quiroz-Mercado; Fernando González-Domínguez; Lucia Hernández-Rivas, Irma López-Martínez; Ernesto Ramírez-González; Maribel González-Villa |
| EPI_ISL_17672235 | LESP State of Mexico | Instituto de Diagnostico y Referencia Epidemiologicos (INDRE) | Abril Rodríguez-Maldonado; Claudia Wong-Arámbula; Silvia Rivero-Arredondo; Ruth Madera-Sandoval; Joaquín Quiroz-Mercado; Fernando González-Domínguez; Lucia Hernández-Rivas, Irma López-Martínez; Ernesto Ramírez-González; Maribel González-Villa |
| EPI_ISL_17672236 | Laboratorio Estatal de Salud Pública Puebla | Instituto de Diagnostico y Referencia Epidemiologicos (INDRE) | Abril Rodríguez-Maldonado; Claudia Wong-Arámbula; Silvia Rivero-Arredondo; Ruth Madera-Sandoval; Joaquín Quiroz-Mercado; Fernando González-Domínguez; Lucia Hernández-Rivas, Irma López-Martínez; Ernesto Ramírez-González; Maribel González-Villa |
| EPI_ISL_17672237 | LESP Campeche | Instituto de Diagnostico y Referencia Epidemiologicos (INDRE) | Abril Rodríguez-Maldonado; Claudia Wong-Arámbula; Silvia Rivero-Arredondo; Ruth Madera-Sandoval; Joaquín Quiroz-Mercado; Fernando González-Domínguez; Lucia Hernández-Rivas, Irma López-Martínez; Ernesto Ramírez-González; Maribel González-Villa |
| EPI_ISL_17672238 | LESP State of Mexico | Instituto de Diagnostico y Referencia Epidemiologicos (INDRE) | Abril Rodríguez-Maldonado; Claudia Wong-Arámbula; Silvia Rivero-Arredondo; Ruth Madera-Sandoval; Joaquín Quiroz-Mercado; Fernando González-Domínguez; Lucia Hernández-Rivas, Irma López-Martínez; Ernesto Ramírez-González; Maribel González-Villa |
| EPI_ISL_17672239 | LESP Chiapas | Instituto de Diagnostico y Referencia Epidemiologicos (INDRE) | Abril Rodríguez-Maldonado; Claudia Wong-Arámbula; Silvia Rivero-Arredondo; Ruth Madera-Sandoval; Joaquín Quiroz-Mercado; Fernando González-Domínguez; Lucia Hernández-Rivas, Irma López-Martínez; Ernesto Ramírez-González; Maribel González-Villa |
| EPI_ISL_17672241 | LESP Jalisco | Instituto de Diagnostico y Referencia Epidemiologicos (INDRE) | Abril Rodríguez-Maldonado; Claudia Wong-Arámbula; Silvia Rivero-Arredondo; Ruth Madera-Sandoval; Joaquín Quiroz-Mercado; Fernando González-Domínguez; Lucia Hernández-Rivas, Irma López-Martínez; Ernesto Ramírez-González; Maribel González-Villa |
| EPI_ISL_17672242 | LESP Nuevo Leon | Instituto de Diagnostico y Referencia Epidemiologicos (INDRE) | Abril Rodríguez-Maldonado; Claudia Wong-Arámbula; Silvia Rivero-Arredondo; Ruth Madera-Sandoval; Joaquín Quiroz-Mercado; Fernando González-Domínguez; Lucia Hernández-Rivas, Irma López-Martínez; Ernesto Ramírez-González; Maribel González-Villa |
| EPI_ISL_17672244 | LESP Hidalgo | Instituto de Diagnostico y Referencia Epidemiologicos (INDRE) | Abril Rodríguez-Maldonado; Claudia Wong-Arámbula; Silvia Rivero-Arredondo; Ruth Madera-Sandoval; Joaquín Quiroz-Mercado; Fernando González-Domínguez; Lucia Hernández-Rivas, Irma López-Martínez; Ernesto Ramírez-González; Maribel González-Villa |
| EPI_ISL_17672245 | LESP Tamaulipas | Instituto de Diagnostico y Referencia Epidemiologicos (INDRE) | Abril Rodríguez-Maldonado; Claudia Wong-Arámbula; Silvia Rivero-Arredondo; Ruth Madera-Sandoval; Joaquín Quiroz-Mercado; Fernando González-Domínguez; Lucia Hernández-Rivas, Irma López-Martínez; Ernesto Ramírez-González; Maribel González-Villa |
| EPI_ISL_17672246 | LESP Veracruz | Instituto de Diagnostico y Referencia Epidemiologicos (INDRE) | Abril Rodríguez-Maldonado; Claudia Wong-Arámbula; Silvia Rivero-Arredondo; Ruth Madera-Sandoval; Joaquín Quiroz-Mercado; Fernando González-Domínguez; Lucia Hernández-Rivas, Irma López-Martínez; Ernesto Ramírez-González; Maribel González-Villa |
| EPI_ISL_17672247 | LESP Quintana Roo | Instituto de Diagnostico y Referencia Epidemiologicos (INDRE) | Abril Rodríguez-Maldonado; Claudia Wong-Arámbula; Silvia Rivero-Arredondo; Ruth Madera-Sandoval; Joaquín Quiroz-Mercado; Fernando González-Domínguez; Lucia Hernández-Rivas, Irma López-Martínez; Ernesto Ramírez-González; Maribel González-Villa |
| EPI_ISL_17672248 | LESP Queretaro | Instituto de Diagnostico y Referencia Epidemiologicos (INDRE) | Abril Rodríguez-Maldonado; Claudia Wong-Arámbula; Silvia Rivero-Arredondo; Ruth Madera-Sandoval; Joaquín Quiroz-Mercado; Fernando González-Domínguez; Lucia Hernández-Rivas, Irma López-Martínez; Ernesto Ramírez-González; Maribel González-Villa |
| EPI_ISL_17672249 | LESP Oaxaca | Instituto de Diagnostico y Referencia Epidemiologicos (INDRE) | Abril Rodríguez-Maldonado; Claudia Wong-Arámbula; Silvia Rivero-Arredondo; Ruth Madera-Sandoval; Joaquín Quiroz-Mercado; Fernando González-Domínguez; Lucia Hernández-Rivas, Irma López-Martínez; Ernesto Ramírez-González; Maribel González-Villa |
| EPI_ISL_17672250 | LESP Yucatan | Instituto de Diagnostico y Referencia Epidemiologicos (INDRE) | Abril Rodríguez-Maldonado; Claudia Wong-Arámbula; Silvia Rivero-Arredondo; Ruth Madera-Sandoval; Joaquín Quiroz-Mercado; Fernando González-Domínguez; Lucia Hernández-Rivas, Irma López-Martínez; Ernesto Ramírez-González; Maribel González-Villa |
| EPI_ISL_17672251 | LESP Guanajuato | Instituto de Diagnostico y Referencia Epidemiologicos (INDRE) | Abril Rodríguez-Maldonado; Claudia Wong-Arámbula; Silvia Rivero-Arredondo; Ruth Madera-Sandoval; Joaquín Quiroz-Mercado; Fernando González-Domínguez; Lucia Hernández-Rivas, Irma López-Martínez; Ernesto Ramírez-González; Maribel González-Villa |
| EPI_ISL_17672252 | LESP Veracruz | Instituto de Diagnostico y Referencia Epidemiologicos (INDRE) | Abril Rodríguez-Maldonado; Claudia Wong-Arámbula; Silvia Rivero-Arredondo; Ruth Madera-Sandoval; Joaquín Quiroz-Mercado; Fernando González-Domínguez; Lucia Hernández-Rivas, Irma López-Martínez; Ernesto Ramírez-González; Maribel González-Villa |
| EPI_ISL_17672253 | LESP Mexico City | Instituto de Diagnostico y Referencia Epidemiologicos (INDRE) | Abril Rodríguez-Maldonado; Claudia Wong-Arámbula; Silvia Rivero-Arredondo; Ruth Madera-Sandoval; Joaquín Quiroz-Mercado; Fernando González-Domínguez; Lucia Hernández-Rivas, Irma López-Martínez; Ernesto Ramírez-González; Maribel González-Villa |
| EPI_ISL_17672254 | LESP Durango | Instituto de Diagnostico y Referencia Epidemiologicos (INDRE) | Abril Rodríguez-Maldonado; Claudia Wong-Arámbula; Silvia Rivero-Arredondo; Ruth Madera-Sandoval; Joaquín Quiroz-Mercado; Fernando González-Domínguez; Lucia Hernández-Rivas, Irma López-Martínez; Ernesto Ramírez-González; Maribel González-Villa |
| EPI_ISL_17672255 | LESP San Luis Potosi | Instituto de Diagnostico y Referencia Epidemiologicos (INDRE) | Abril Rodríguez-Maldonado; Claudia Wong-Arámbula; Silvia Rivero-Arredondo; Ruth Madera-Sandoval; Joaquín Quiroz-Mercado; Fernando González-Domínguez; Lucia Hernández-Rivas, Irma López-Martínez; Ernesto Ramírez-González; Maribel González-Villa |
| EPI_ISL_17672256 | LESP Nuevo Leon | Instituto de Diagnostico y Referencia Epidemiologicos (INDRE) | Abril Rodríguez-Maldonado; Claudia Wong-Arámbula; Silvia Rivero-Arredondo; Ruth Madera-Sandoval; Joaquín Quiroz-Mercado; Fernando González-Domínguez; Lucia Hernández-Rivas, Irma López-Martínez; Ernesto Ramírez-González; Maribel González-Villa |
| EPI_ISL_17672257 | LESP Quintana Roo | Instituto de Diagnostico y Referencia Epidemiologicos (INDRE) | Abril Rodríguez-Maldonado; Claudia Wong-Arámbula; Silvia Rivero-Arredondo; Ruth Madera-Sandoval; Joaquín Quiroz-Mercado; Fernando González-Domínguez; Lucia Hernández-Rivas, Irma López-Martínez; Ernesto Ramírez-González; Maribel González-Villa |
| EPI_ISL_17672259 | LESP Mexico City | Instituto de Diagnostico y Referencia Epidemiologicos (INDRE) | Abril Rodríguez-Maldonado; Claudia Wong-Arámbula; Silvia Rivero-Arredondo; Ruth Madera-Sandoval; Joaquín Quiroz-Mercado; Fernando González-Domínguez; Lucia Hernández-Rivas, Irma López-Martínez; Ernesto Ramírez-González; Maribel González-Villa |
| EPI_ISL_17672260 | LESP Veracruz | Instituto de Diagnostico y Referencia Epidemiologicos (INDRE) | Abril Rodríguez-Maldonado; Claudia Wong-Arámbula; Silvia Rivero-Arredondo; Ruth Madera-Sandoval; Joaquín Quiroz-Mercado; Fernando González-Domínguez; Lucia Hernández-Rivas, Irma López-Martínez; Ernesto Ramírez-González; Maribel González-Villa |
| EPI_ISL_17692268, EPI_ISL_17692269 | Tokyo Metropolitan Institute of Public Health | Tokyo Metropolitan Institute of Public Health | Fumi Kasuya, Wakaba Okada, Ryota Kumagai, Sachiko Harada, Arisa Amano, Michiya Hasegawa, Mami Nagashima, Kenji Sadamasu |
| EPI_ISL_17703564 | LESP State of Mexico | Instituto de Diagnostico y Referencia Epidemiologicos (INDRE) | Abril Rodríguez-Maldonado; Claudia Wong-Arámbula; Felipe Arguijo-Perez; Helios Cárdenas-Hernández; Carmen Castro-Méndez; Lidia García-Torres; Ruth Madera-Sandoval; América Mandujano-Martínez; Nancy Martínez-Velázquez; Mireya Mederos-Michel; Angélica Pedraza-Meléndez; Joaquín Quiroz-Mercado; Daniel Regalado-Santiago; Silvia Rivero-Arredondo; Erika Sierra-Atanacio; Fernando González-Domínguez; Lucia Hernández-Rivas, Irma López-Martínez; Ernesto Ramírez-González; Maribel González-Villa |
| EPI_ISL_17703566 | LESP Jalisco | Instituto de Diagnostico y Referencia Epidemiologicos (INDRE) | Abril Rodríguez-Maldonado; Claudia Wong-Arámbula; Felipe Arguijo-Perez; Helios Cárdenas-Hernández; Carmen Castro-Méndez; Lidia García-Torres; Ruth Madera-Sandoval; América Mandujano-Martínez; Nancy Martínez-Velázquez; Mireya Mederos-Michel; Angélica Pedraza-Meléndez; Joaquín Quiroz-Mercado; Daniel Regalado-Santiago; Silvia Rivero-Arredondo; Erika Sierra-Atanacio; Fernando González-Domínguez; Lucia Hernández-Rivas, Irma López-Martínez; Ernesto Ramírez-González; Maribel González-Villa |
| EPI_ISL_17703570 | LESP Mexico City | Instituto de Diagnostico y Referencia Epidemiologicos (INDRE) | Abril Rodríguez-Maldonado; Claudia Wong-Arámbula; Felipe Arguijo-Perez; Helios Cárdenas-Hernández; Carmen Castro-Méndez; Lidia García-Torres; Ruth Madera-Sandoval; América Mandujano-Martínez; Nancy Martínez-Velázquez; Mireya Mederos-Michel; Angélica Pedraza-Meléndez; Joaquín Quiroz-Mercado; Daniel Regalado-Santiago; Silvia Rivero-Arredondo; Erika Sierra-Atanacio; Fernando González-Domínguez; Lucia Hernández-Rivas, Irma López-Martínez; Ernesto Ramírez-González; Maribel González-Villa |
| EPI_ISL_17703571 | LESP Sinaloa | Instituto de Diagnostico y Referencia Epidemiologicos (INDRE) | Abril Rodríguez-Maldonado; Claudia Wong-Arámbula; Felipe Arguijo-Perez; Helios Cárdenas-Hernández; Carmen Castro-Méndez; Lidia García-Torres; Ruth Madera-Sandoval; América Mandujano-Martínez; Nancy Martínez-Velázquez; Mireya Mederos-Michel; Angélica Pedraza-Meléndez; Joaquín Quiroz-Mercado; Daniel Regalado-Santiago; Silvia Rivero-Arredondo; Erika Sierra-Atanacio; Fernando González-Domínguez; Lucia Hernández-Rivas, Irma López-Martínez; Ernesto Ramírez-González; Maribel González-Villa |

| | | | |
|---|---|---|---|
| EPI_ISL_17703572 | LESP State of Mexico | Instituto de Diagnostico y Referencia Epidemiologicos (INDRE) | Abril Rodríguez-Maldonado; Claudia Wong-Arámbula; Felipe Arguijo-Perez; Helios Cárdenas-Hernández; Carmen Castro-Méndez; Lidia García-Torres; Ruth Madera-Sandoval; América Mandujano-Martínez; Nancy Martínez-Velázquez; Mireya Mederos-Michel; Angélica Pedraza-Meléndez; Joaquín Quiroz-Mercado; Daniel Regalado-Santiago; Silvia Rivero-Arredondo; Erika Sierra-Atanacio; Fernando González-Domínguez; Lucia Hernández-Rivas, Irma López-Martínez; Ernesto Ramírez-González; Maribel González-Villa |
| EPI_ISL_17703573 | LESP Mexico City | Instituto de Diagnostico y Referencia Epidemiologicos (INDRE) | Abril Rodríguez-Maldonado; Claudia Wong-Arámbula; Felipe Arguijo-Perez; Helios Cárdenas-Hernández; Carmen Castro-Méndez; Lidia García-Torres; Ruth Madera-Sandoval; América Mandujano-Martínez; Nancy Martínez-Velázquez; Mireya Mederos-Michel; Angélica Pedraza-Meléndez; Joaquín Quiroz-Mercado; Daniel Regalado-Santiago; Silvia Rivero-Arredondo; Erika Sierra-Atanacio; Fernando González-Domínguez; Lucia Hernández-Rivas, Irma López-Martínez; Ernesto Ramírez-González; Maribel González-Villa |
| EPI_ISL_17703574 | LESP State of Mexico | Instituto de Diagnostico y Referencia Epidemiologicos (INDRE) | Abril Rodríguez-Maldonado; Claudia Wong-Arámbula; Felipe Arguijo-Perez; Helios Cárdenas-Hernández; Carmen Castro-Méndez; Lidia García-Torres; Ruth Madera-Sandoval; América Mandujano-Martínez; Nancy Martínez-Velázquez; Mireya Mederos-Michel; Angélica Pedraza-Meléndez; Joaquín Quiroz-Mercado; Daniel Regalado-Santiago; Silvia Rivero-Arredondo; Erika Sierra-Atanacio; Fernando González-Domínguez; Lucia Hernández-Rivas, Irma López-Martínez; Ernesto Ramírez-González; Maribel González-Villa |
| EPI_ISL_17703575 | LESP Mexico City | Instituto de Diagnostico y Referencia Epidemiologicos (INDRE) | Abril Rodríguez-Maldonado; Claudia Wong-Arámbula; Felipe Arguijo-Perez; Helios Cárdenas-Hernández; Carmen Castro-Méndez; Lidia García-Torres; Ruth Madera-Sandoval; América Mandujano-Martínez; Nancy Martínez-Velázquez; Mireya Mederos-Michel; Angélica Pedraza-Meléndez; Joaquín Quiroz-Mercado; Daniel Regalado-Santiago; Silvia Rivero-Arredondo; Erika Sierra-Atanacio; Fernando González-Domínguez; Lucia Hernández-Rivas, Irma López-Martínez; Ernesto Ramírez-González; Maribel González-Villa |
| EPI_ISL_17703576 | LESP Quintana Roo | Instituto de Diagnostico y Referencia Epidemiologicos (INDRE) | Abril Rodríguez-Maldonado; Claudia Wong-Arámbula; Felipe Arguijo-Perez; Helios Cárdenas-Hernández; Carmen Castro-Méndez; Lidia García-Torres; Ruth Madera-Sandoval; América Mandujano-Martínez; Nancy Martínez-Velázquez; Mireya Mederos-Michel; Angélica Pedraza-Meléndez; Joaquín Quiroz-Mercado; Daniel Regalado-Santiago; Silvia Rivero-Arredondo; Erika Sierra-Atanacio; Fernando González-Domínguez; Lucia Hernández-Rivas, Irma López-Martínez; Ernesto Ramírez-González; Maribel González-Villa |
| EPI_ISL_17703577 | LESP Mexico City | Instituto de Diagnostico y Referencia Epidemiologicos (INDRE) | Abril Rodríguez-Maldonado; Claudia Wong-Arámbula; Felipe Arguijo-Perez; Helios Cárdenas-Hernández; Carmen Castro-Méndez; Lidia García-Torres; Ruth Madera-Sandoval; América Mandujano-Martínez; Nancy Martínez-Velázquez; Mireya Mederos-Michel; Angélica Pedraza-Meléndez; Joaquín Quiroz-Mercado; Daniel Regalado-Santiago; Silvia Rivero-Arredondo; Erika Sierra-Atanacio; Fernando González-Domínguez; Lucia Hernández-Rivas, Irma López-Martínez; Ernesto Ramírez-González; Maribel González-Villa |
| EPI_ISL_17703578 | LESP Yucatan | Instituto de Diagnostico y Referencia Epidemiologicos (INDRE) | Abril Rodríguez-Maldonado; Claudia Wong-Arámbula; Felipe Arguijo-Perez; Helios Cárdenas-Hernández; Carmen Castro-Méndez; Lidia García-Torres; Ruth Madera-Sandoval; América Mandujano-Martínez; Nancy Martínez-Velázquez; Mireya Mederos-Michel; Angélica Pedraza-Meléndez; Joaquín Quiroz-Mercado; Daniel Regalado-Santiago; Silvia Rivero-Arredondo; Erika Sierra-Atanacio; Fernando González-Domínguez; Lucia Hernández-Rivas, Irma López-Martínez; Ernesto Ramírez-González; Maribel González-Villa |
| EPI_ISL_17703579 | LESP Quintana Roo | Instituto de Diagnostico y Referencia Epidemiologicos (INDRE) | Abril Rodríguez-Maldonado; Claudia Wong-Arámbula; Felipe Arguijo-Perez; Helios Cárdenas-Hernández; Carmen Castro-Méndez; Lidia García-Torres; Ruth Madera-Sandoval; América Mandujano-Martínez; Nancy Martínez-Velázquez; Mireya Mederos-Michel; Angélica Pedraza-Meléndez; Joaquín Quiroz-Mercado; Daniel Regalado-Santiago; Silvia Rivero-Arredondo; Erika Sierra-Atanacio; Fernando González-Domínguez; Lucia Hernández-Rivas, Irma López-Martínez; Ernesto Ramírez-González; Maribel González-Villa |
| EPI_ISL_17703580 | LESP Hidalgo | Instituto de Diagnostico y Referencia Epidemiologicos (INDRE) | Abril Rodríguez-Maldonado; Claudia Wong-Arámbula; Felipe Arguijo-Perez; Helios Cárdenas-Hernández; Carmen Castro-Méndez; Lidia García-Torres; Ruth Madera-Sandoval; América Mandujano-Martínez; Nancy Martínez-Velázquez; Mireya Mederos-Michel; Angélica Pedraza-Meléndez; Joaquín Quiroz-Mercado; Daniel Regalado-Santiago; Silvia Rivero-Arredondo; Erika Sierra-Atanacio; Fernando González-Domínguez; Lucia Hernández-Rivas, Irma López-Martínez; Ernesto Ramírez-González; Maribel González-Villa |
| EPI_ISL_17703582 | LESP Nuevo Leon | Instituto de Diagnostico y Referencia Epidemiologicos (INDRE) | Abril Rodríguez-Maldonado; Claudia Wong-Arámbula; Felipe Arguijo-Perez; Helios Cárdenas-Hernández; Carmen Castro-Méndez; Lidia García-Torres; Ruth Madera-Sandoval; América Mandujano-Martínez; Nancy Martínez-Velázquez; Mireya Mederos-Michel; Angélica Pedraza-Meléndez; Joaquín Quiroz-Mercado; Daniel Regalado-Santiago; Silvia Rivero-Arredondo; Erika Sierra-Atanacio; Fernando González-Domínguez; Lucia Hernández-Rivas, Irma López-Martínez; Ernesto Ramírez-González; Maribel González-Villa |
| EPI_ISL_17703583 | LESP Quintana Roo | Instituto de Diagnostico y Referencia Epidemiologicos (INDRE) | Abril Rodríguez-Maldonado; Claudia Wong-Arámbula; Felipe Arguijo-Perez; Helios Cárdenas-Hernández; Carmen Castro-Méndez; Lidia García-Torres; Ruth Madera-Sandoval; América Mandujano-Martínez; Nancy Martínez-Velázquez; Mireya Mederos-Michel; Angélica Pedraza-Meléndez; Joaquín Quiroz-Mercado; Daniel Regalado-Santiago; Silvia Rivero-Arredondo; Erika Sierra-Atanacio; Fernando González-Domínguez; Lucia Hernández-Rivas, Irma López-Martínez; Ernesto Ramírez-González; Maribel González-Villa |
| EPI_ISL_17703584 | LESP Chihuahua | Instituto de Diagnostico y Referencia Epidemiologicos (INDRE) | Abril Rodríguez-Maldonado; Claudia Wong-Arámbula; Felipe Arguijo-Perez; Helios Cárdenas-Hernández; Carmen Castro-Méndez; Lidia García-Torres; Ruth Madera-Sandoval; América Mandujano-Martínez; Nancy Martínez-Velázquez; Mireya Mederos-Michel; Angélica Pedraza-Meléndez; Joaquín Quiroz-Mercado; Daniel Regalado-Santiago; Silvia Rivero-Arredondo; Erika Sierra-Atanacio; Fernando González-Domínguez; Lucia Hernández-Rivas, Irma López-Martínez; Ernesto Ramírez-González; Maribel González-Villa |
| EPI_ISL_17703585 | LESP Yucatan | Instituto de Diagnostico y Referencia Epidemiologicos (INDRE) | Abril Rodríguez-Maldonado; Claudia Wong-Arámbula; Felipe Arguijo-Perez; Helios Cárdenas-Hernández; Carmen Castro-Méndez; Lidia García-Torres; Ruth Madera-Sandoval; América Mandujano-Martínez; Nancy Martínez-Velázquez; Mireya Mederos-Michel; Angélica Pedraza-Meléndez; Joaquín Quiroz-Mercado; Daniel Regalado-Santiago; Silvia Rivero-Arredondo; Erika Sierra-Atanacio; Fernando González-Domínguez; Lucia Hernández-Rivas, Irma López-Martínez; Ernesto Ramírez-González; Maribel González-Villa |
| EPI_ISL_17703586, EPI_ISL_17703587 | LESP Nuevo Leon | Instituto de Diagnostico y Referencia Epidemiologicos (INDRE) | Abril Rodríguez-Maldonado; Claudia Wong-Arámbula; Felipe Arguijo-Perez; Helios Cárdenas-Hernández; Carmen Castro-Méndez; Lidia García-Torres; Ruth Madera-Sandoval; América Mandujano-Martínez; Nancy Martínez-Velázquez; Mireya Mederos-Michel; Angélica Pedraza-Meléndez; Joaquín Quiroz-Mercado; Daniel Regalado-Santiago; Silvia Rivero-Arredondo; Erika Sierra-Atanacio; Fernando González-Domínguez; Lucia Hernández-Rivas, Irma López-Martínez; Ernesto Ramírez-González; Maribel González-Villa |
| EPI_ISL_17703588, EPI_ISL_17703589 | LESP Quintana Roo | Instituto de Diagnostico y Referencia Epidemiologicos (INDRE) | Abril Rodríguez-Maldonado; Claudia Wong-Arámbula; Felipe Arguijo-Perez; Helios Cárdenas-Hernández; Carmen Castro-Méndez; Lidia García-Torres; Ruth Madera-Sandoval; América Mandujano-Martínez; Nancy Martínez-Velázquez; Mireya Mederos-Michel; Angélica Pedraza-Meléndez; Joaquín Quiroz-Mercado; Daniel Regalado-Santiago; Silvia Rivero-Arredondo; Erika Sierra-Atanacio; Fernando González-Domínguez; Lucia Hernández-Rivas, Irma López-Martínez; Ernesto Ramírez-González; Maribel González-Villa |
| EPI_ISL_17703590 | LESP State of Mexico | Instituto de Diagnostico y Referencia Epidemiologicos (INDRE) | Abril Rodríguez-Maldonado; Claudia Wong-Arámbula; Felipe Arguijo-Perez; Helios Cárdenas-Hernández; Carmen Castro-Méndez; Lidia García-Torres; Ruth Madera-Sandoval; América Mandujano-Martínez; Nancy Martínez-Velázquez; Mireya Mederos-Michel; Angélica Pedraza-Meléndez; Joaquín Quiroz-Mercado; Daniel Regalado-Santiago; Silvia Rivero-Arredondo; Erika Sierra-Atanacio; Fernando González-Domínguez; Lucia Hernández-Rivas, Irma López-Martínez; Ernesto Ramírez-González; Maribel González-Villa |
| EPI_ISL_17703591 | LESP Campeche | Instituto de Diagnostico y Referencia Epidemiologicos (INDRE) | Abril Rodríguez-Maldonado; Claudia Wong-Arámbula; Felipe Arguijo-Perez; Helios Cárdenas-Hernández; Carmen Castro-Méndez; Lidia García-Torres; Ruth Madera-Sandoval; América Mandujano-Martínez; Nancy Martínez-Velázquez; Mireya Mederos-Michel; Angélica Pedraza-Meléndez; Joaquín Quiroz-Mercado; Daniel Regalado-Santiago; Silvia Rivero-Arredondo; Erika Sierra-Atanacio; Fernando González-Domínguez; Lucia Hernández-Rivas, Irma López-Martínez; Ernesto Ramírez-González; Maribel González-Villa |
| EPI_ISL_17703592 | LESP Nuevo Leon | Instituto de Diagnostico y Referencia Epidemiologicos (INDRE) | Abril Rodríguez-Maldonado; Claudia Wong-Arámbula; Felipe Arguijo-Perez; Helios Cárdenas-Hernández; Carmen Castro-Méndez; Lidia García-Torres; Ruth Madera-Sandoval; América Mandujano-Martínez; Nancy Martínez-Velázquez; Mireya Mederos-Michel; Angélica Pedraza-Meléndez; Joaquín Quiroz-Mercado; Daniel Regalado-Santiago; Silvia Rivero-Arredondo; Erika Sierra-Atanacio; Fernando González-Domínguez; Lucia Hernández-Rivas, Irma López-Martínez; Ernesto Ramírez-González; Maribel González-Villa |
| EPI_ISL_17703593, EPI_ISL_17703594 | LESP Jalisco | Instituto de Diagnostico y Referencia Epidemiologicos (INDRE) | Abril Rodríguez-Maldonado; Claudia Wong-Arámbula; Felipe Arguijo-Perez; Helios Cárdenas-Hernández; Carmen Castro-Méndez; Lidia García-Torres; Ruth Madera-Sandoval; América Mandujano-Martínez; Nancy Martínez-Velázquez; Mireya Mederos-Michel; Angélica Pedraza-Meléndez; Joaquín Quiroz-Mercado; Daniel Regalado-Santiago; Silvia Rivero-Arredondo; Erika Sierra-Atanacio; Fernando González-Domínguez; Lucia Hernández-Rivas, Irma López-Martínez; Ernesto Ramírez-González; Maribel González-Villa |
| EPI_ISL_17703595, EPI_ISL_17703596 | LESP Nuevo Leon | Instituto de Diagnostico y Referencia Epidemiologicos (INDRE) | Abril Rodríguez-Maldonado; Claudia Wong-Arámbula; Felipe Arguijo-Perez; Helios Cárdenas-Hernández; Carmen Castro-Méndez; Lidia García-Torres; Ruth Madera-Sandoval; América Mandujano-Martínez; Nancy Martínez-Velázquez; Mireya Mederos-Michel; Angélica Pedraza-Meléndez; Joaquín Quiroz-Mercado; Daniel Regalado-Santiago; Silvia Rivero-Arredondo; Erika Sierra-Atanacio; Fernando González-Domínguez; Lucia Hernández-Rivas, Irma López-Martínez; Ernesto Ramírez-González; Maribel González-Villa |
| EPI_ISL_17703598 | LESP Quintana Roo | Instituto de Diagnostico y Referencia Epidemiologicos (INDRE) | Abril Rodríguez-Maldonado; Claudia Wong-Arámbula; Felipe Arguijo-Perez; Helios Cárdenas-Hernández; Carmen Castro-Méndez; Lidia García-Torres; Ruth Madera-Sandoval; América Mandujano-Martínez; Nancy Martínez-Velázquez; Mireya Mederos-Michel; Angélica Pedraza-Meléndez; Joaquín Quiroz-Mercado; Daniel Regalado-Santiago; Silvia Rivero-Arredondo; Erika Sierra-Atanacio; Fernando González-Domínguez; Lucia Hernández-Rivas, Irma López-Martínez; Ernesto Ramírez-González; Maribel González-Villa |
| EPI_ISL_17703599, EPI_ISL_17703600, EPI_ISL_17703601, EPI_ISL_17703602, EPI_ISL_17703603 | LESP Nuevo Leon | Instituto de Diagnostico y Referencia Epidemiologicos (INDRE) | Abril Rodríguez-Maldonado; Claudia Wong-Arámbula; Felipe Arguijo-Perez; Helios Cárdenas-Hernández; Carmen Castro-Méndez; Lidia García-Torres; Ruth Madera-Sandoval; América Mandujano-Martínez; Nancy Martínez-Velázquez; Mireya Mederos-Michel; Angélica Pedraza-Meléndez; Joaquín Quiroz-Mercado; Daniel Regalado-Santiago; Silvia Rivero-Arredondo; Erika Sierra-Atanacio; Fernando González-Domínguez; Lucia Hernández-Rivas, Irma López-Martínez; Ernesto Ramírez-González; Maribel González-Villa |
| EPI_ISL_17703605 | LESP Chihuahua | Instituto de Diagnostico y Referencia Epidemiologicos (INDRE) | Abril Rodríguez-Maldonado; Claudia Wong-Arámbula; Felipe Arguijo-Perez; Helios Cárdenas-Hernández; Carmen Castro-Méndez; Lidia García-Torres; Ruth Madera-Sandoval; América Mandujano-Martínez; Nancy Martínez-Velázquez; Mireya Mederos-Michel; Angélica Pedraza-Meléndez; Joaquín Quiroz-Mercado; Daniel Regalado-Santiago; Silvia Rivero-Arredondo; Erika Sierra-Atanacio; Fernando González-Domínguez; Lucia Hernández-Rivas, Irma López-Martínez; Ernesto Ramírez-González; Maribel González-Villa |
| EPI_ISL_17703606 | LESP Nuevo Leon | Instituto de Diagnostico y Referencia Epidemiologicos (INDRE) | Abril Rodríguez-Maldonado; Claudia Wong-Arámbula; Felipe Arguijo-Perez; Helios Cárdenas-Hernández; Carmen Castro-Méndez; Lidia García-Torres; Ruth Madera-Sandoval; América Mandujano-Martínez; Nancy Martínez-Velázquez; Mireya Mederos-Michel; Angélica Pedraza-Meléndez; Joaquín Quiroz-Mercado; Daniel Regalado-Santiago; Silvia Rivero-Arredondo; Erika Sierra-Atanacio; Fernando González-Domínguez; Lucia Hernández-Rivas, Irma López-Martínez; Ernesto Ramírez-González; Maribel González-Villa |
| EPI_ISL_17703607, EPI_ISL_17703608, EPI_ISL_17703609, EPI_ISL_17703610 | LESP Mexico City | Instituto de Diagnostico y Referencia Epidemiologicos (INDRE) | Abril Rodríguez-Maldonado; Claudia Wong-Arámbula; Felipe Arguijo-Perez; Helios Cárdenas-Hernández; Carmen Castro-Méndez; Lidia García-Torres; Ruth Madera-Sandoval; América Mandujano-Martínez; Nancy Martínez-Velázquez; Mireya Mederos-Michel; Angélica Pedraza-Meléndez; Joaquín Quiroz-Mercado; Daniel Regalado-Santiago; Silvia Rivero-Arredondo; Erika Sierra-Atanacio; Fernando González-Domínguez; Lucia Hernández-Rivas, Irma López-Martínez; Ernesto Ramírez-González; Maribel González-Villa |
| EPI_ISL_17703611 | LESP Tlaxcala | Instituto de Diagnostico y Referencia Epidemiologicos (INDRE) | Abril Rodríguez-Maldonado; Claudia Wong-Arámbula; Felipe Arguijo-Perez; Helios Cárdenas-Hernández; Carmen Castro-Méndez; Lidia García-Torres; Ruth Madera-Sandoval; América Mandujano-Martínez; Nancy Martínez-Velázquez; Mireya Mederos-Michel; Angélica Pedraza-Meléndez; Joaquín Quiroz-Mercado; Daniel Regalado-Santiago; Silvia Rivero-Arredondo; Erika Sierra-Atanacio; Fernando González-Domínguez; Lucia Hernández-Rivas, Irma López-Martínez; Ernesto Ramírez-González; Maribel González-Villa |
| EPI_ISL_17703612 | LESP Nuevo Leon | Instituto de Diagnostico y Referencia Epidemiologicos (INDRE) | Abril Rodríguez-Maldonado; Claudia Wong-Arámbula; Felipe Arguijo-Perez; Helios Cárdenas-Hernández; Carmen Castro-Méndez; Lidia García-Torres; Ruth Madera-Sandoval; América Mandujano-Martínez; Nancy Martínez-Velázquez; Mireya Mederos-Michel; Angélica Pedraza-Meléndez; Joaquín Quiroz-Mercado; Daniel Regalado-Santiago; Silvia Rivero-Arredondo; Erika Sierra-Atanacio; Fernando González-Domínguez; Lucia Hernández-Rivas, Irma López-Martínez; Ernesto Ramírez-González; Maribel González-Villa |
| EPI_ISL_17703743, EPI_ISL_17703744 | Parasitology Laboratory, Institute of Tropical Medicine of Sao Paulo, School of Medicine, University of Sao Paulo | Parasitology Laboratory | Raissa Heloisa de Araujo Eliodoro, Ingra Claro Morales, Ester Cerdeira Sabino |
| EPI_ISL_17703745 | Parasitology Laboratory, Institute of Tropical Medicine of Sao Paulo, School of Medicine, University of Sao Paulo | Parasitology Laboratory | Raissa Heloisa de Araujo Eliodoro, Ingra Morales Claro, Ester Cerdeira Sabino |
| EPI_ISL_17703746 | Parasitology Laboratory, Institute of Tropical Medicine of Sao Paulo, School of Medicine, University of Sao Paulo | Parasitology Laboratory | Raissa Heloisa de Araujo Eliodoro, Ingra Claro Morales, Ester Cerdeira Sabino |
| EPI_ISL_17718458 | Department of Virology, National Institute of Health, Islamabad, Pakistan | Department of Virology, National Institute of Health, Islamabad, Pakistan | Massab Umair, Muhammad Ammar, Syed Adnan Haider, Rabia Hakim, Qasim Malik, Muhammad Salman, Ghazala Parveen, and Naseem Akhtar |
| EPI_ISL_17722468 | Tokyo Metropolitan Institute of Public Health | Tokyo Metropolitan Institute of Public Health | Fumi Kasuya, Wakaba Okada, Ryota Kumagai, Sachiko Harada, Arisa Amano, Michiya Hasegawa, Mami Nagashima, Kenji Sadamasu |
| EPI_ISL_17736865, EPI_ISL_17736866, EPI_ISL_17736867, EPI_ISL_17736868, EPI_ISL_17736869, EPI_ISL_17736870, EPI_ISL_17736871, EPI_ISL_17736872, EPI_ISL_17736873, EPI_ISL_17736874, EPI_ISL_17736875, EPI_ISL_17736876, EPI_ISL_17736877, EPI_ISL_17736878, EPI_ISL_17736879, EPI_ISL_17736880, EPI_ISL_17736881, EPI_ISL_17736882, EPI_ISL_17736883, EPI_ISL_17736884, EPI_ISL_17736885, EPI_ISL_17736886, EPI_ISL_17736887, EPI_ISL_17736888, EPI_ISL_17736889, EPI_ISL_17736890, EPI_ISL_17736891 | | | |

| | | | |
|---|---|---|---|
| see above | Charité Universitätsmedizin Berlin, Institute for Virology/Laboratory Berlin | Charité Universitätsmedizin Berlin, Institute for Virology | Terry C. Jones, Julia Melchert, Barbara Mühlemann, Talitha Veith, Jörn Beheim-Schwarzbach, Julia Tesch, Marie Luisa Schmidt, Felix Walper, Tobias Bleicker, Caroline Isner, Frieder Pfäfflin, Ricardo Niklas Werner, Victor M. Corman, Christian Drosten |
| EPI_ISL_17737476, EPI_ISL_17737493, EPI_ISL_17737495, EPI_ISL_17737500, EPI_ISL_17737507, EPI_ISL_17737527, EPI_ISL_17737538 | Public Health Ontario | Public Health Ontario | Isabel S, Eshaghi A, Duvvuri VR, Gubbay JB, Cronin K, Li A, Hasso M, Clark ST, Hopkins JP, Patel SN, Braukmann TWA |
| EPI_ISL_17762484, EPI_ISL_17762485 | Tokyo Metropolitan Institute of Public Health | Tokyo Metropolitan Institute of Public Health | Fumi Kasuya, Wakaba Okada, Ryota Kumagai, Sachiko Harada, Arisa Amano, Michiya Hasegawa, Mami Nagashima, Kenji Sadamasu |
| EPI_ISL_17809521 | Hangzhou Center for Disease Control and Prevention | Hangzhou Center for Disease Control and Prevention | Lijiao Ao , Jun Li , Yue Yu |
| EPI_ISL_17817239, EPI_ISL_17817240, EPI_ISL_17817241 | Tokyo Metropolitan Institute of Public Health | Tokyo Metropolitan Institute of Public Health | Fumi Kasuya, Wakaba Okada, Ryota Kumagai, Sachiko Harada, Arisa Amano, Michiya Hasegawa, Mami Nagashima, Kenji Sadamasu |
| EPI_ISL_17821080, EPI_ISL_17821081, EPI_ISL_17821082, EPI_ISL_17821084, EPI_ISL_17821085, EPI_ISL_17821086, EPI_ISL_17821087, EPI_ISL_17821088 | National Institute for Infectious Diseases "Matei Bals" | National Institute for Infectious Diseases "Matei Bals" | Robert Hohan, Ovidiu Vlaicu, Marius Surleac, Leontina Banica, Andreea Tudor, Simona Paraschiv |
| EPI_ISL_17837266, EPI_ISL_17837267, EPI_ISL_17837268, EPI_ISL_17959214, EPI_ISL_17959215, EPI_ISL_17959216 | Tokyo Metropolitan Institute of Public Health | Tokyo Metropolitan Institute of Public Health | Fumi Kasuya, Wakaba Okada, Ryota Kumagai, Sachiko Harada, Arisa Amano, Michiya Hasegawa, Mami Nagashima, Kenji Sadamasu |
| EPI_ISL_17960863, EPI_ISL_17960864, EPI_ISL_17960865, EPI_ISL_17960866 | Laboratorio Nacional de Salud Pública Dr. Defilló | Laboratorio Nacional de Salud Pública Dr. Defilló | Isaac Miguel Sánchez, Carlos Vergara Castillo, Edwin Féliz, Anny Peña, Pedro Martinez, Yeny E. Lara Perez, Robinson Agramonte |
| EPI_ISL_17977751 | Centro Medico ABC | Instituto Nacional de Medicina Genomica | Cedro Tanda Alberto, Roxana Trejo Gonzalez, Laura Gomez-Romero, Alfredo Mendoza-Vargas, Dora Garnica-Lopez, Alfredo Hidalgo-Miranda. |
| EPI_ISL_17988349, EPI_ISL_17988350, EPI_ISL_17988351, EPI_ISL_17988352, EPI_ISL_17988353, EPI_ISL_17988354, EPI_ISL_17988355, EPI_ISL_17988356, EPI_ISL_17988357, EPI_ISL_17988358, EPI_ISL_17988359, EPI_ISL_17988360, EPI_ISL_17988361, EPI_ISL_17988362, EPI_ISL_17988363, EPI_ISL_17988364, EPI_ISL_17988365, EPI_ISL_17988366, EPI_ISL_17988367, EPI_ISL_17988368, EPI_ISL_17988369, EPI_ISL_17988370, EPI_ISL_17988371, EPI_ISL_17988372, EPI_ISL_17988373, EPI_ISL_17988374, EPI_ISL_17988375, EPI_ISL_17988376, EPI_ISL_17988377, EPI_ISL_17988378 | | | |
| see above | Laboratorio Central de Salud Publica | Laboratorio Central de Salud Publica | Cynthia Vazquez, Vagner Fonseca, Andrea Gomez de la Fuente, Sandra Gonzalez, Fatima Fleitas, Mauricio Lima, Natalia R. Guimaraes, Felipe C. M. Iani, Analia Rojas, Tania Alfonso, Cesar Cantero, Julio Barrios, Shirley Villalba, Maria Jose Ortega, Juan Torales, Maria Liz Gamarra, Carolina Aquino, Jairo Mendez Rico, Luiz Carlos Junior Alcantara, Marta Giovanetti |
| EPI_ISL_18055899, EPI_ISL_18055900 | Tokyo Metropolitan Institute of Public Health | Tokyo Metropolitan Institute of Public Health | Fumi Kasuya, Wakaba Okada, Ryota Kumagai, Sachiko Harada, Arisa Amano, Michiya Hasegawa, Mami Nagashima, Kenji Sadamasu |
| EPI_ISL_18059182, EPI_ISL_18059183, EPI_ISL_18059184 | Department of Acute Infectious Diseases Control and Prevention, Yunnan Center for Disease Control and Prevention | Department of Acute Infectious Diseases Control and Prevention, Yunnan Center for Disease Control and Prevention | Meiling Zhang, Ruize Ni, Xiaoqing Fu |
| EPI_ISL_18064640, EPI_ISL_18064641, EPI_ISL_18064642, EPI_ISL_18064643, EPI_ISL_18064644, EPI_ISL_18064645, EPI_ISL_18064648 | Laboratory of Microbiology and Virology, Ospedale Amedeo di Savoia, ASL "Città di Torino" | Laboratory of Microbiology and Virology, Ospedale Amedeo di Savoia, ASL "Città di Torino" | Francesco Cerutti, Tiziano Allice, Maria Grazia Milia, Gabriella Gregori, Elisa Burdino, Sara Monteleone, Marisa Cazzadore, Valeria Ghisetti |
| EPI_ISL_18075506, EPI_ISL_18075507, EPI_ISL_18075508, EPI_ISL_18097375 | Tokyo Metropolitan Institute of Public Health | Tokyo Metropolitan Institute of Public Health | Fumi Kasuya, Wakaba Okada, Ryota Kumagai, Sachiko Harada, Arisa Amano, Michiya Hasegawa, Mami Nagashima, Kenji Sadamasu |
| EPI_ISL_18137807, EPI_ISL_18137808 | Northwestern Medicine | RIPHL at Rush University Medical Center | Stefan Green, Kevin Kunstman, Hannah Barbian, Sofiya Bobrovska, Felix Araujo Perez, Edith Perez, Cecilia Chau, Giancarlo Balangue, Lok Yiu Ashley Wu, Trisha Jeon, Marisol Dominguez, Latifah Boyd, Lacy Simons |
| EPI_ISL_18137812 | Quest Diagnostics | RIPHL at Rush University Medical Center | Stefan Green, Kevin Kunstman, Hannah Barbian, Sofiya Bobrovska, Felix Araujo Perez, Edith Perez, Cecilia Chau, Giancarlo Balangue, Lok Yiu Ashley Wu, Trisha Jeon, Marisol Dominguez, Latifah Boyd |
| EPI_ISL_18161269, EPI_ISL_18161271, EPI_ISL_18161272, EPI_ISL_18161275 | Quest Diagnostics Nichols Institute | Los Angeles County Public Health Laboratories | J. Garrigues et al. |
| EPI_ISL_18161280 | UCLA Clinical Micro Lab | Los Angeles County Public Health Laboratories | J. Garrigues et al. |
| EPI_ISL_18161284 | Quest Diagnostics Nichols Institute | Los Angeles County Public Health Laboratories | J. Garrigues et al. |
| EPI_ISL_18161288 | Los Angeles County Public Health Laboratories | Los Angeles County Public Health Laboratories | J. Garrigues et al. |
| EPI_ISL_18161293, EPI_ISL_18161294 | Laboratory Corporation of America | Los Angeles County Public Health Laboratories | J. Garrigues et al. |
| EPI_ISL_18161299, EPI_ISL_18161301, EPI_ISL_18161302, EPI_ISL_18161305, EPI_ISL_18161311, EPI_ISL_18161314 | Quest Diagnostics Nichols Institute | Los Angeles County Public Health Laboratories | J. Garrigues et al. |
| EPI_ISL_18161321, EPI_ISL_18161323 | Laboratory Corporation of America | Los Angeles County Public Health Laboratories | J. Garrigues et al. |
| EPI_ISL_18161325 | Los Angeles County Public Health Laboratories | Los Angeles County Public Health Laboratories | J. Garrigues et al. |
| EPI_ISL_18213374, EPI_ISL_18213375 | Institute for Hepatology,Shenzhen Third People's Hospital | Institute for Hepatology,Shenzhen Third People's Hospital | Lin Cheng,Zheng Zhang |
| EPI_ISL_18238305, EPI_ISL_18238308, EPI_ISL_18238309, EPI_ISL_18238318, EPI_ISL_18238324, EPI_ISL_18238325 | DPH, Massachusetts State Public Health Laboratory | DPH, Massachusetts State Public Health Laboratory | Doucette,M., Gagne,L. and Smole,S.C. |
| EPI_ISL_18241786, EPI_ISL_18241787, EPI_ISL_18241788, EPI_ISL_18241789, EPI_ISL_18241790, EPI_ISL_18241791, EPI_ISL_18241792 | Unidade de Genômica - UFRJ | Unidade de Genômica - UFRJ | Carolina Moreira Voloch, Filipe Romero Rebello Moreira, Diana Mariani, Rafael Mello Galliez, Debora Souza Faffe, Terezinha Marta Pereira Pinto Castiñeiras, Clarissa Damaso, Amílcar Tanuri. |
| EPI_ISL_18257122 | Haidian CDC | Haidian District Center for Disease Control and Prevention Microbiological Laboratory | Fangyao Liu, Lifei Shi,Feng Liu, Heng Zhang |
| EPI_ISL_18285960, EPI_ISL_18285966, EPI_ISL_18285967, EPI_ISL_18285968, EPI_ISL_18285970 | Quest Diagnostics Nichols Institute | Los Angeles County Public Health Laboratories | J. Garrigues et al. |
| EPI_ISL_18285978 | Quest Diagnostics | RIPHL at Rush University Medical Center | Stefan Green, Kevin Kunstman, Hannah Barbian, Sofiya Bobrovska, Felix Araujo Perez, Edith Perez, Cecilia Chau, Giancarlo Balangue, Lok Yiu Ashley Wu, Trisha Jeon, Marisol Dominguez, Latifah Boyd |
| EPI_ISL_18308395, EPI_ISL_18308396, EPI_ISL_18308397, EPI_ISL_18308398, EPI_ISL_18308399 | National Virus Reference Laboratory | National Virus Reference Laboratory | Gabriel Gonzalez, Michael Carr, Emer O'Byrne, Weronika Banka, Brian Keogan, Jonathan Dean, Daniel Hare, Cillian F De Gascun |
| EPI_ISL_18323779, EPI_ISL_18323780, EPI_ISL_18323781, EPI_ISL_18323782, EPI_ISL_18323783, EPI_ISL_18323784, EPI_ISL_18323785, EPI_ISL_18323786, EPI_ISL_18323787, EPI_ISL_18323788, EPI_ISL_18323789, EPI_ISL_18323790, EPI_ISL_18323791, EPI_ISL_18323792, EPI_ISL_18323793, EPI_ISL_18323794, EPI_ISL_18324980, EPI_ISL_18324981, EPI_ISL_18324982, EPI_ISL_18324983, EPI_ISL_18324984, EPI_ISL_18324985, EPI_ISL_18324986, EPI_ISL_18324987, EPI_ISL_18324988, EPI_ISL_18324989, EPI_ISL_18324990, EPI_ISL_18324991, EPI_ISL_18324992, EPI_ISL_18324993, EPI_ISL_18324994, EPI_ISL_18324995, EPI_ISL_18324996, EPI_ISL_18324997, EPI_ISL_18324998, EPI_ISL_18324999, EPI_ISL_18325000, EPI_ISL_18325001, EPI_ISL_18325002, EPI_ISL_18325003, EPI_ISL_18325004, EPI_ISL_18325005, EPI_ISL_18325006, EPI_ISL_18325007, EPI_ISL_18325008, EPI_ISL_18325009, EPI_ISL_18325010, EPI_ISL_18325011 | | | |
| see above | California Department of Public Health | California Department of Public Health | Kath, C., Haw, M., Espinosa, A., and Hacker, J. |
| EPI_ISL_18352302, EPI_ISL_18352303, EPI_ISL_18352304, EPI_ISL_18352305, EPI_ISL_18352306 | Tokyo Metropolitan Institute of Public Health | Tokyo Metropolitan Institute of Public Health | Fumi Kasuya, Wakaba Okada, Ryota Kumagai, Sachiko Harada, Arisa Amano, Michiya Hasegawa, Mami Nagashima, Kenji Sadamasu |
| EPI_ISL_18360394 | Haidian District Center for Disease Control and Prevention Microbiological Laboratory | Haidian District Center for Disease Control and Prevention Microbiological Laboratory | Fangyao Liu, Lifei Shi,Feng Liu, Heng Zhang |
| EPI_ISL_18387016 | Institut Pasteur de Dakar, Virology Unit | Institut Pasteur de Dakar, Virology Unit | Martin,F., Anges,Y., Benjamin,H., Amadou,S.A. and Ousmane,F. |
| EPI_ISL_18399134, EPI_ISL_18399135, EPI_ISL_18399136, EPI_ISL_18399137, EPI_ISL_18399138, EPI_ISL_18399139, EPI_ISL_18399140, EPI_ISL_18399141, EPI_ISL_18399142, EPI_ISL_18399143, EPI_ISL_18399144, EPI_ISL_18399145, EPI_ISL_18399146, EPI_ISL_18399147 | | | |
| see above | California Department of Public Health | California Department of Public Health | Kath, C., Haw, M., Espinosa, A., and Hacker, J. |
| EPI_ISL_18427687, EPI_ISL_18427688 | Quest Diagnostics | RIPHL at Rush University Medical Center | Stefan Green, Kevin Kunstman, Hannah Barbian, Sofiya Bobrovska, Felix Araujo Perez, Edith Perez, Cecilia Chau, Giancarlo Balangue, Lok Yiu Ashley Wu, Trisha Jeon, Marisol Dominguez, Latifah Boyd |
| EPI_ISL_18436041 | PKC Jatinegara | National Institute of Health Research and Development | Fajar Nur Sulistyahadi, Arie Ardiansyah Nugraha, Hana Apsari Pawestri, Kartika Dewi Puspa, Herna, Subangkit, IGM Wirabrata |
| EPI_ISL_18443030, EPI_ISL_18443031, EPI_ISL_18443032, EPI_ISL_18443033, EPI_ISL_18443034, EPI_ISL_18443035, EPI_ISL_18443036, EPI_ISL_18443037, EPI_ISL_18443038, EPI_ISL_18443039, EPI_ISL_18443040, EPI_ISL_18443041, EPI_ISL_18443042, EPI_ISL_18443043, EPI_ISL_18443044, EPI_ISL_18443045, EPI_ISL_18452332, EPI_ISL_18452333, EPI_ISL_18452334, EPI_ISL_18452335, EPI_ISL_18452336, EPI_ISL_18452337, EPI_ISL_18452338, EPI_ISL_18452339, EPI_ISL_18452340, EPI_ISL_18452341, EPI_ISL_18452342, EPI_ISL_18452343, EPI_ISL_18452344, EPI_ISL_18452345, EPI_ISL_18452346, EPI_ISL_18452347, EPI_ISL_18458948, EPI_ISL_18458949, EPI_ISL_18458950, EPI_ISL_18458951, EPI_ISL_18458952, EPI_ISL_18458953, EPI_ISL_18458954, EPI_ISL_18458955, EPI_ISL_18458956, EPI_ISL_18458957, EPI_ISL_18458958, EPI_ISL_18458959, EPI_ISL_18458960, EPI_ISL_18458961, EPI_ISL_18458962, EPI_ISL_18458963, EPI_ISL_18458964, EPI_ISL_18458965, EPI_ISL_18458966, EPI_ISL_18460494, EPI_ISL_18460495, EPI_ISL_18460496, EPI_ISL_18460497, EPI_ISL_18460498, EPI_ISL_18460499, EPI_ISL_18460500, EPI_ISL_18460501, EPI_ISL_18460502, EPI_ISL_18460503, EPI_ISL_18460504, EPI_ISL_18460505 | | | |
| see above | California Department of Public Health | California Department of Public Health | Kath, C., Haw, M., Espinosa, A., and Hacker, J. |
| EPI_ISL_18463158 | PKM Kembangan | National Institute of Health Research and Development | Hana Apsari Pawestri, Arie Ardiansyah Nugraha, Fajar Nur Sulistyahadi, Hartanti Dian Ikawati, Kartika Dewi Puspa, Markus Evan Anggia, Subangkit, Nelis Imaningsih, IGM Wirabrata |
| EPI_ISL_18463159 | PKC Setiabudi | National Institute of Health Research and Development | Hana Apsari Pawestri, Arie Ardiansyah Nugraha, Fajar Nur Sulistyahadi, Hartanti Dian Ikawati, Kartika Dewi Puspa, Markus Evan Anggia, Subangkit, Nelis Imaningsih, IGM Wirabrata |
| EPI_ISL_18463160 | RSUPN Dr Cipto Mangunkusumo | National Institute of Health Research and | Hana Apsari Pawestri, Arie Ardiansyah Nugraha, Fajar Nur Sulistyahadi, Hartanti Dian Ikawati, Kartika Dewi Puspa, Markus Evan Anggia, Subangkit, Nelis Imaningsih, IGM Wirabrata |

| | | | |
|---|---|---|---|
| | | Development | |
| EPI_ISL_18463161 | RSUD Kembangan | National Institute of Health Research and Development | Hana Apsari Pawestri, Arie Ardiansyah Nugraha, Fajar Nur Sulistyahadi, Hartanti Dian Ikawati, Kartika Dewi Puspa, Markus Evan Anggia, Subangkit, Nelis Imaningsih, IGM Wirabrata |
| EPI_ISL_18467794 | Eka Hospital BSD | National Institute of Health Research and Development | Fajar Nur Sulistyahadi, Hana Apsari Pawestri, Arie Ardiansyah Nugraha, Hartanti Dian Ikawati, Kartika Dewi Puspa, Subangkit, IGM Wirabrata |
| EPI_ISL_18467795, EPI_ISL_18467796 | PKM Kembangan | National Institute of Health Research and Development | Hana Apsari Pawestri, Arie Ardiansyah Nugraha, Fajar Nur Sulistyahadi, Hartanti Dian Ikawati, Kartika Dewi Puspa, Subangkit, IGM Wirabrata |
| EPI_ISL_18467797 | PKC Cengkareng | National Institute of Health Research and Development | Arie Ardiansyah Nugraha, Fajar Nur Sulistyahadi, Hartanti Dian Ikawati, Kartika Dewi Puspa, Hana Apsari Pawestri, Subangkit, IGM Wirabrata |
| EPI_ISL_18467798 | PKC Grogol Petamburan | National Institute of Health Research and Development | Fajar Nur Sulistyahadi, Hana Apsari Pawestri, Arie Ardiansyah Nugraha, Hartanti Dian Ikawati, Kartika Dewi Puspa, Subangkit, IGM Wirabrata |
| EPI_ISL_18467799 | PKC Setiabudi | National Institute of Health Research and Development | Hana Apsari Pawestri, Arie Ardiansyah Nugraha, Fajar Nur Sulistyahadi, Hartanti Dian Ikawati, Kartika Dewi Puspa, Subangkit, IGM Wirabrata |
| EPI_ISL_18467800 | PKC Pancoran | National Institute of Health Research and Development | Arie Ardiansyah Nugraha, Fajar Nur Sulistyahadi, Hartanti Dian Ikawati, Kartika Dewi Puspa, Hana Apsari Pawestri, Subangkit, IGM Wirabrata |
| EPI_ISL_18467801 | PKM Mampang Prapatan | National Institute of Health Research and Development | Arie Ardiansyah Nugraha, Fajar Nur Sulistyahadi, Hartanti Dian Ikawati, Kartika Dewi Puspa, Hana Apsari Pawestri, Subangkit, IGM Wirabrata |
| EPI_ISL_18467802 | PKC Cilandak | National Institute of Health Research and Development | Fajar Nur Sulistyahadi, Hana Apsari Pawestri, Arie Ardiansyah Nugraha, Hartanti Dian Ikawati, Kartika Dewi Puspa, Subangkit, IGM Wirabrata |
| EPI_ISL_18467803 | RS Brawijaya Saharjo | National Institute of Health Research and Development | Fajar Nur Sulistyahadi, Hana Apsari Pawestri, Arie Ardiansyah Nugraha, Hartanti Dian Ikawati, Kartika Dewi Puspa, Subangkit, IGM Wirabrata |
| EPI_ISL_18467804 | PKC Pulogadung | National Institute of Health Research and Development | Hana Apsari Pawestri, Arie Ardiansyah Nugraha, Fajar Nur Sulistyahadi, Hartanti Dian Ikawati, Kartika Dewi Puspa, Subangkit, IGM Wirabrata |
| EPI_ISL_18467805 | PKC Kramat Jati | National Institute of Health Research and Development | Arie Ardiansyah Nugraha, Fajar Nur Sulistyahadi, Hartanti Dian Ikawati, Kartika Dewi Puspa, Hana Apsari Pawestri, Subangkit, IGM Wirabrata |
| EPI_ISL_18467806 | PKM Tanjung Priuk | National Institute of Health Research and Development | Fajar Nur Sulistyahadi, Hana Apsari Pawestri, Arie Ardiansyah Nugraha, Hartanti Dian Ikawati, Kartika Dewi Puspa, Subangkit, IGM Wirabrata |
| EPI_ISL_18467807 | PKC Kelapa Gading | National Institute of Health Research and Development | Fajar Nur Sulistyahadi, Hana Apsari Pawestri, Arie Ardiansyah Nugraha, Hartanti Dian Ikawati, Kartika Dewi Puspa, Subangkit, IGM Wirabrata |
| EPI_ISL_18467808 | RSUP Dr Hasan Sadikin | National Institute of Health Research and Development | Arie Ardiansyah Nugraha, Fajar Nur Sulistyahadi, Hartanti Dian Ikawati, Kartika Dewi Puspa, Hana Apsari Pawestri, Subangkit, IGM Wirabrata |
| EPI_ISL_18486349, EPI_ISL_18486350, EPI_ISL_18486351 | U.O. Microbiologia Laboratorio Unico Centro Servizi - Azienda Unità Sanitarie Locali della Romagna | U.O. Microbiologia Laboratorio Unico Centro Servizi - Azienda Unità Sanitarie Locali della Romagna - DIMEC - Università di Bologna | Alessandra Scagliarini, Vittorio Sambri, Maria Elena Turba, Fabio Gentilini, Francesca Taddei, Giorgio Dirani, Silvia Zannoli, Giulia Gatti, Martina Brandolini, Alessandra Mistral De Pascali, Monica Cricca |
| EPI_ISL_18557816, EPI_ISL_18557817, EPI_ISL_18557818 | Tokyo Metropolitan Institute of Public Health | Tokyo Metropolitan Institute of Public Health | Fumi Kasuya, Wakaba Okada, Ryota Kumagai, Sachiko Harada, Arisa Amano, Michiya Hasegawa, Mami Nagashima, Kenji Sadamasu |
| EPI_ISL_18567806, EPI_ISL_18567807 | Southern Nevada Public Health Laboratory | Southern Nevada Public Health Laboratory | Horng-Yuan Kan |
| EPI_ISL_18634755, EPI_ISL_18634756 | National Virus Reference Laboratory | National Virus Reference Laboratory | Gabriel Gonzalez, Michael Carr, Emer O'Byrne, Weronika Banka, Brian Keogan, Jose Maria Urtasun Elizari, Jonathan Dean, Daniel Hare, Cillian F De Gascun |
| EPI_ISL_18642356 | PKC Senen | National Institute of Health Research and Development | Hana Apsari Pawestri, Arie Ardiansyah Nugraha, Fajar Nur Sulistyahadi, Markus Evan Anggia, Subangkit, Herna, IGM Wirabrata |
| EPI_ISL_18642357 | PKC Cakung | National Institute of Health Research and Development | Hana Apsari Pawestri, Arie Ardiansyah Nugraha, Fajar Nur Sulistyahadi, Markus Evan Anggia, Subangkit, Herna, IGM Wirabrata |
| EPI_ISL_18642358 | RS Mitra Keluarga Gading | National Institute of Health Research and Development | Hana Apsari Pawestri, Arie Ardiansyah Nugraha, Fajar Nur Sulistyahadi, Markus Evan Anggia, Subangkit, Herna, IGM Wirabrata |
| EPI_ISL_18642359 | RSUP Persahabatan | National Institute of Health Research and Development | Hana Apsari Pawestri, Arie Ardiansyah Nugraha, Fajar Nur Sulistyahadi, Markus Evan Anggia, Subangkit, Herna, IGM Wirabrata |
| EPI_ISL_18642360 | PKC Cilandak | National Institute of Health Research and Development | Hana Apsari Pawestri, Arie Ardiansyah Nugraha, Fajar Nur Sulistyahadi, Markus Evan Anggia, Subangkit, Herna, IGM Wirabrata |
| EPI_ISL_18642361 | PKC Mampang Prapatan | National Institute of Health Research and Development | Hana Apsari Pawestri, Arie Ardiansyah Nugraha, Fajar Nur Sulistyahadi, Markus Evan Anggia, Subangkit, Herna, IGM Wirabrata |
| EPI_ISL_18642362 | Dinkes Kabupaten Cirebon | National Institute of Health Research and Development | Hana Apsari Pawestri, Arie Ardiansyah Nugraha, Fajar Nur Sulistyahadi, Markus Evan Anggia, Subangkit, Herna, IGM Wirabrata |
| EPI_ISL_18642363 | PKC Setiabudi | National Institute of Health Research and Development | Hana Apsari Pawestri, Arie Ardiansyah Nugraha, Fajar Nur Sulistyahadi, Markus Evan Anggia, Subangkit, Herna, IGM Wirabrata |
| EPI_ISL_18642364 | PKM Pancoran | National Institute of Health Research and Development | Hana Apsari Pawestri, Arie Ardiansyah Nugraha, Fajar Nur Sulistyahadi, Markus Evan Anggia, Subangkit, Herna, IGM Wirabrata |
| EPI_ISL_18642365 | Puskesmas Bambu Apus | National Institute of Health Research and Development | Hana Apsari Pawestri, Arie Ardiansyah Nugraha, Fajar Nur Sulistyahadi, Markus Evan Anggia, Subangkit, Herna, IGM Wirabrata |
| EPI_ISL_18659828 | Erasmus Medical Center Department of Virology | Erasmus Medical Center Department of Virology | Leonard Schuele, Bas Oude Munnink, Marjan Boter, Babette Weller, Babs Verstrepen, Richard Molenkamp, Reina Sikkema, Marion Koopmans |
| EPI_ISL_18659829, EPI_ISL_18659846 | Erasmus Medical Center Department of Virology | Erasmus Medical Center Department of Virology | Leonard Schuele, Marjan Boter, Hayley Cassidy, Babette Weller, Babs Verstrepen, Richard Molenkamp, Marion Koopmans, Bas Oude Munnink |
| EPI_ISL_18697752 | National Institute of Public Health | Institut Pasteur du Cambodge, Virology Unit | Janin Nouhin, Leakhena Pum, Jurre Y Siegers, Chin Savuth, Chau Darapheak, Veasna Duong, Erik A Karlsson |
| EPI_ISL_18719998, EPI_ISL_18719999 | CT Department of Public Health | CT Department of Public Health | Claire Pearson, Tu N. Nguyen, Kutluhan Incekara, Neranjan V. Perera |
| EPI_ISL_18722047 | Dallas Regional Medical Center | Dallas County Health and Human Services Public Health Laboratory | Kabir, Farruk; Plaisance, Erin; Stringer, Joey; Short, Luke. |
| EPI_ISL_18722048 | DCHHS Sexual Health Clinic | Dallas County Health and Human Services Public Health Laboratory | Kabir, Farruk; Plaisance, Erin; Stringer, Joey; Short, Luke. |
| EPI_ISL_18722049, EPI_ISL_18722050, EPI_ISL_18722051, EPI_ISL_18722052, EPI_ISL_18722053, EPI_ISL_18722054 | Parkland Health and Hospital System | Dallas County Health and Human Services Public Health Laboratory | Kabir, Farruk; Plaisance, Erin; Stringer, Joey; Short, Luke. |
| EPI_ISL_18723946 | Dallas Regional Medical Center | Dallas County Health & Human Services Public Health Laboratory | Kabir, Farruk; Plaisance, Erin; Stringer, Joey; Short, Luke. |
| EPI_ISL_18723947, EPI_ISL_18723948 | DCHHS Sexual Health Clinic | Dallas County Health & Human Services Public Health Laboratory | Kabir, Farruk; Plaisance, Erin; Stringer, Joey; Short, Luke. |
| EPI_ISL_18723949 | Texas Health Presbyterian at Dallas | Dallas County Health & Human Services Public Health Laboratory | Kabir, Farruk; Plaisance, Erin; Stringer, Joey; Short, Luke. |
| EPI_ISL_18723950 | DCHHS Sexual Health Clinic | Dallas County Health & Human Services Public Health Laboratory | Kabir, Farruk; Plaisance, Erin; Stringer, Joey; Short, Luke. |
| EPI_ISL_18723951 | Baylor Scott and White Sunnyvale | Dallas County Health & Human Services Public Health Laboratory | Kabir, Farruk; Plaisance, Erin; Stringer, Joey; Short, Luke. |
| EPI_ISL_18723952, EPI_ISL_18723953 | Parkland Health and Hospital System | Dallas County Health & Human Services Public Health Laboratory | Kabir, Farruk; Plaisance, Erin; Stringer, Joey; Short, Luke. |
| EPI_ISL_18723954, EPI_ISL_18723955 | DCHHS Sexual Health Clinic | Dallas County Health & Human Services Public Health Laboratory | Kabir, Farruk; Plaisance, Erin; Stringer, Joey; Short, Luke. |
| EPI_ISL_18723956 | MD Progressive Care | Dallas County Health & Human Services Public Health Laboratory | Kabir, Farruk; Plaisance, Erin; Stringer, Joey; Short, Luke. |
| EPI_ISL_18723957 | Collin County Health Care Services | Dallas County Health & Human Services Public Health Laboratory | Kabir, Farruk; Plaisance, Erin; Stringer, Joey; Short, Luke. |
| EPI_ISL_18723958 | Baylor University Medical Center Dallas | Dallas County Health & Human Services Public Health Laboratory | Kabir, Farruk; Plaisance, Erin; Stringer, Joey; Short, Luke. |

| | | | |
|---|---|---|---|
| EPI_ISL_18723959, EPI_ISL_18723960, EPI_ISL_18723961, EPI_ISL_18723962 | Parkland Health and Hospital System | Dallas County Health & Human Services Public Health Laboratory | Kabir, Farruk; Plaisance, Erin; Stringer, Joey; Short, Luke. |
| EPI_ISL_18723963 | DCHHS Sexual Health Clinic | Dallas County Health & Human Services Public Health Laboratory | Kabir, Farruk; Plaisance, Erin; Stringer, Joey; Short, Luke. |
| EPI_ISL_18723964, EPI_ISL_18723965, EPI_ISL_18723966, EPI_ISL_18723967, EPI_ISL_18723968 | Parkland Health and Hospital System | Dallas County Health & Human Services Public Health Laboratory | Kabir, Farruk; Plaisance, Erin; Stringer, Joey; Short, Luke. |
| EPI_ISL_18723969 | DCHHS Sexual Health Clinic | Dallas County Health & Human Services Public Health Laboratory | Kabir, Farruk; Plaisance, Erin; Stringer, Joey; Short, Luke. |
| EPI_ISL_18723970 | Texas Health Presbyterian at Dallas | Dallas County Health & Human Services Public Health Laboratory | Kabir, Farruk; Plaisance, Erin; Stringer, Joey; Short, Luke. |
| EPI_ISL_18723971 | White Rock Medical Center | Dallas County Health & Human Services Public Health Laboratory | Kabir, Farruk; Plaisance, Erin; Stringer, Joey; Short, Luke. |
| EPI_ISL_18723972 | Dallas Regional Medical Center | Dallas County Health & Human Services Public Health Laboratory | Kabir, Farruk; Plaisance, Erin; Stringer, Joey; Short, Luke. |
| EPI_ISL_18723973 | MD Progressive Care | Dallas County Health & Human Services Public Health Laboratory | Kabir, Farruk; Plaisance, Erin; Stringer, Joey; Short, Luke. |
| EPI_ISL_18723974, EPI_ISL_18723975, EPI_ISL_18723976, EPI_ISL_18723977, EPI_ISL_18723978, EPI_ISL_18723979, EPI_ISL_18723980 | Parkland Health and Hospital System | Dallas County Health & Human Services Public Health Laboratory | Kabir, Farruk; Plaisance, Erin; Stringer, Joey; Short, Luke. |
| EPI_ISL_18723981 | MD Progressive Care | Dallas County Health & Human Services Public Health Laboratory | Kabir, Farruk; Plaisance, Erin; Stringer, Joey; Short, Luke. |
| EPI_ISL_18723982 | Dallas Regional Medical Center | Dallas County Health & Human Services Public Health Laboratory | Kabir, Farruk; Plaisance, Erin; Stringer, Joey; Short, Luke. |
| EPI_ISL_18723983 | Parkland Health and Hospital System | Dallas County Health & Human Services Public Health Laboratory | Kabir, Farruk; Plaisance, Erin; Stringer, Joey; Short, Luke. |
| EPI_ISL_18723984 | Dallas County Health and Human Services Sexual Health Clinic | Dallas County Health & Human Services Public Health Laboratory | Kabir, Farruk; Plaisance, Erin; Stringer, Joey; Short, Luke. |
| EPI_ISL_18723985 | Dallas Regional Medical Center | Dallas County Health & Human Services Public Health Laboratory | Kabir, Farruk; Plaisance, Erin; Stringer, Joey; Short, Luke. |
| EPI_ISL_18723986, EPI_ISL_18723987 | Parkland Health and Hospital System | Dallas County Health & Human Services Public Health Laboratory | Kabir, Farruk; Plaisance, Erin; Stringer, Joey; Short, Luke. |
| EPI_ISL_18723988 | Children's Health Dallas Texas | Dallas County Health & Human Services Public Health Laboratory | Kabir, Farruk; Plaisance, Erin; Stringer, Joey; Short, Luke. |
| EPI_ISL_18723989 | Dallas County Health and Human Services Sexual Health Clinic | Dallas County Health & Human Services Public Health Laboratory | Kabir, Farruk; Plaisance, Erin; Stringer, Joey; Short, Luke. |
| EPI_ISL_18723990, EPI_ISL_18737442, EPI_ISL_18737443 | Parkland Health and Hospital System | Dallas County Health & Human Services Public Health Laboratory | Kabir, Farruk; Plaisance, Erin; Stringer, Joey; Short, Luke. |
| EPI_ISL_18737444 | Dallas Veteran Affairs Hospital | Dallas County Health & Human Services Public Health Laboratory | Kabir, Farruk; Plaisance, Erin; Stringer, Joey; Short, Luke. |
| EPI_ISL_18737445, EPI_ISL_18737446, EPI_ISL_18737447, EPI_ISL_18737448, EPI_ISL_18737449, EPI_ISL_18737450, EPI_ISL_18737451, EPI_ISL_18737520 | Parkland Health and Hospital System | Dallas County Health & Human Services Public Health Laboratory | Kabir, Farruk; Plaisance, Erin; Stringer, Joey; Short, Luke. |
| EPI_ISL_18737522 | Dallas Veteran Affairs Hospital | Dallas County Health & Human Services Public Health Laboratory | Kabir, Farruk; Plaisance, Erin; Stringer, Joey; Short, Luke. |
| EPI_ISL_18737523, EPI_ISL_18737524, EPI_ISL_18737525, EPI_ISL_18737526, EPI_ISL_18737527, EPI_ISL_18737528, EPI_ISL_18737529, EPI_ISL_18737530, EPI_ISL_18737544, EPI_ISL_18737545, EPI_ISL_18737546, EPI_ISL_18737547, EPI_ISL_18737548 | | | |
| see above | Parkland Health and Hospital System | Dallas County Health & Human Services Public Health Laboratory | Kabir, Farruk; Plaisance, Erin; Stringer, Joey; Short, Luke. |
| EPI_ISL_18737549, EPI_ISL_18737550 | MD Progressive Care | Dallas County Health & Human Services Public Health Laboratory | Kabir, Farruk; Plaisance, Erin; Stringer, Joey; Short, Luke. |
| EPI_ISL_18737551, EPI_ISL_18737552, EPI_ISL_18737553, EPI_ISL_18737554, EPI_ISL_18737555 | Parkland Health and Hospital System | Dallas County Health & Human Services Public Health Laboratory | Kabir, Farruk; Plaisance, Erin; Stringer, Joey; Short, Luke. |
| EPI_ISL_18737556 | Dallas County Health & Human Services Sexual Health Clinic | Dallas County Health & Human Services Public Health Laboratory | Kabir, Farruk; Plaisance, Erin; Stringer, Joey; Short, Luke. |
| EPI_ISL_18737557 | Parkland Health and Hospital System | Dallas County Health & Human Services Public Health Laboratory | Kabir, Farruk; Plaisance, Erin; Stringer, Joey; Short, Luke. |
| EPI_ISL_18739594, EPI_ISL_18739595 | Tokyo Metropolitan Institute of Public Health | Tokyo Metropolitan Institute of Public Health | Fumi Kasuya, Wakaba Okada, Ryota Kumagai, Sachiko Harada, Arisa Amano, Michiya Hasegawa, Mami Nagashima, Kenji Sadamasu |
| EPI_ISL_18744048, EPI_ISL_18744050 | National Medical Center | National Medical Center | Jun-sun Park, Hongsoon Yim, Jihye Um, Hyang Su Kim, BumSik Chin, Jaehyun Jeon, Yeonjae Kim, Min-Kyung Kim |
| EPI_ISL_18746882, EPI_ISL_18746884, EPI_ISL_18746885 | Erasmus Medical Center Department of Virology | Erasmus Medical Center Department of Virology | Leonard Schuele, Marjan Boter, Babs Verstrepen, Richard Molenkamp, Marion Koopmans, Bas Oude Munnink |
| EPI_ISL_18746956, EPI_ISL_18746957, EPI_ISL_18746958, EPI_ISL_18746959, EPI_ISL_18746960, EPI_ISL_18746961, EPI_ISL_18746962, EPI_ISL_18746963, EPI_ISL_18746964, EPI_ISL_18746965, EPI_ISL_18746966, EPI_ISL_18746967 | | | |
| see above | Parkland Health and Hospital System | Dallas County Health & Human Services Public Health Laboratory | Kabir, Farruk; Plaisance, Erin; Stringer, Joey; Short, Luke. |
| EPI_ISL_18746968 | DCHHS Sexual Health Clinic | Dallas County Health & Human Services Public Health Laboratory | Kabir, Farruk; Plaisance, Erin; Stringer, Joey; Short, Luke. |
| EPI_ISL_18746969 | Parkland Health and Hospital System | Dallas County Health & Human Services Public Health Laboratory | Kabir, Farruk; Plaisance, Erin; Stringer, Joey; Short, Luke. |
| EPI_ISL_18746970 | DCHHS Sexual Health Clinic | Dallas County Health & Human Services Public Health Laboratory | Kabir, Farruk; Plaisance, Erin; Stringer, Joey; Short, Luke. |
| EPI_ISL_18746971, EPI_ISL_18746972, EPI_ISL_18746973, EPI_ISL_18746974, EPI_ISL_18746975, EPI_ISL_18746976, EPI_ISL_18746977, EPI_ISL_18746978, EPI_ISL_18746979, EPI_ISL_18746980, EPI_ISL_18746981, EPI_ISL_18746982, EPI_ISL_18746983, EPI_ISL_18746984, EPI_ISL_18746985, EPI_ISL_18746986, EPI_ISL_18746987, EPI_ISL_18746988, EPI_ISL_18746989, EPI_ISL_18746990, EPI_ISL_18746991, EPI_ISL_18746992, EPI_ISL_18746993, EPI_ISL_18746994, EPI_ISL_18746995, EPI_ISL_18746996, EPI_ISL_18746997 | | | |
| see above | Parkland Health and Hospital System | Dallas County Health & Human Services Public Health Laboratory | Kabir, Farruk; Plaisance, Erin; Stringer, Joey; Short, Luke. |
| EPI_ISL_18746998 | White Rock Medical Center | Dallas County Health & Human Services Public Health Laboratory | Kabir, Farruk; Plaisance, Erin; Stringer, Joey; Short, Luke. |
| EPI_ISL_18746999 | DCHHS Sexual Health Clinic | Dallas County Health & Human Services Public Health Laboratory | Kabir, Farruk; Plaisance, Erin; Stringer, Joey; Short, Luke. |
| EPI_ISL_18747000, EPI_ISL_18747001, EPI_ISL_18747002, EPI_ISL_18747003, EPI_ISL_18747004, EPI_ISL_18747005, EPI_ISL_18747006, EPI_ISL_18747007, EPI_ISL_18747008, EPI_ISL_18747009, EPI_ISL_18747010, EPI_ISL_18747011 | | | |
| see above | Parkland Health and Hospital System | Dallas County Health & Human Services Public Health Laboratory | Kabir, Farruk; Plaisance, Erin; Stringer, Joey; Short, Luke. |
| EPI_ISL_18747012 | DCHHS Sexual Health Clinic | Dallas County Health & Human Services Public Health Laboratory | Kabir, Farruk; Plaisance, Erin; Stringer, Joey; Short, Luke. |
| EPI_ISL_18747013, EPI_ISL_18747014, EPI_ISL_18747015, EPI_ISL_18747016, EPI_ISL_18747017, EPI_ISL_18747018, EPI_ISL_18747019, EPI_ISL_18747020, EPI_ISL_18747021, EPI_ISL_18747022, EPI_ISL_18747023, EPI_ISL_18747024, EPI_ISL_18747025, EPI_ISL_18747026, EPI_ISL_18747027, EPI_ISL_18747028 | | | |
| see above | Parkland Health and Hospital System | Dallas County Health & Human Services Public Health Laboratory | Kabir, Farruk; Plaisance, Erin; Stringer, Joey; Short, Luke. |
| EPI_ISL_18747029 | DCHHS Sexual Health Clinic | Dallas County Health & Human Services Public Health Laboratory | Kabir, Farruk; Plaisance, Erin; Stringer, Joey; Short, Luke. |

| | | | |
|---|---|---|---|
| EPI_ISL_18755970, EPI_ISL_18755971, EPI_ISL_18755972, EPI_ISL_18755973, EPI_ISL_18755974, EPI_ISL_18755975, EPI_ISL_18755976, EPI_ISL_18755977, EPI_ISL_18755978, EPI_ISL_18755979 | Erasmus Medical Center Department of Virology | Erasmus Medical Center Department of Virology | Leonard Schuele, Marjan Boter, Babs Verstrepen, Richard Molenkamp, Marion Koopmans, Bas Oude Munnink |
| EPI_ISL_18773109, EPI_ISL_18773110, EPI_ISL_18773111, EPI_ISL_18773112, EPI_ISL_18773114, EPI_ISL_18773116, EPI_ISL_18773118, EPI_ISL_18773119, EPI_ISL_18773121, EPI_ISL_18773122, EPI_ISL_18773123, EPI_ISL_18773124, EPI_ISL_18773126, EPI_ISL_18773127, EPI_ISL_18773128, EPI_ISL_18773129, EPI_ISL_18773130, EPI_ISL_18773131, EPI_ISL_18773132, EPI_ISL_18773133, EPI_ISL_18773134, EPI_ISL_18773135, EPI_ISL_18773139, EPI_ISL_18773140, EPI_ISL_18773141, EPI_ISL_18773144, EPI_ISL_18773147, EPI_ISL_18773148, EPI_ISL_18773149, EPI_ISL_18773150 | | | |
| see above | Animal Health - Istituto Zooprofilattico Sperimentale del Mezzogiorno | Animal Health - Istituto Zooprofilattico Sperimentale del Mezzogiorno | Viscardi,M., Cozzolino,L., Rinaldi,A., De Martinis,C., Cardillo,L., Tiberio,C., Falco,R., Guarino,V., D'Auria,G., Nappo,F., Atripaldi,L., Coppola,M.G. and Fusco,G. |
| EPI_ISL_18781624, EPI_ISL_18781625, EPI_ISL_18781626, EPI_ISL_18781627, EPI_ISL_18781628, EPI_ISL_18781629, EPI_ISL_18781630, EPI_ISL_18781631, EPI_ISL_18781632, EPI_ISL_18781633, EPI_ISL_18781634, EPI_ISL_18781635, EPI_ISL_18781636, EPI_ISL_18781637, EPI_ISL_18781785, EPI_ISL_18781786, EPI_ISL_18781787, EPI_ISL_18781788, EPI_ISL_18781790, EPI_ISL_18781791, EPI_ISL_18781792, EPI_ISL_18781793, EPI_ISL_18781794, EPI_ISL_18781795, EPI_ISL_18781796, EPI_ISL_18781797, EPI_ISL_18781798, EPI_ISL_18781799, EPI_ISL_18781806, EPI_ISL_18781807, EPI_ISL_18781808, EPI_ISL_18781809, EPI_ISL_18781810, EPI_ISL_18781811, EPI_ISL_18781812, EPI_ISL_18781813, EPI_ISL_18781815, EPI_ISL_18781816, EPI_ISL_18781817, EPI_ISL_18781818, EPI_ISL_18781819, EPI_ISL_18781820, EPI_ISL_18781833, EPI_ISL_18781834, EPI_ISL_18781835, EPI_ISL_18781836, EPI_ISL_18781837, EPI_ISL_18781839, EPI_ISL_18781840, EPI_ISL_18781841, EPI_ISL_18781842, EPI_ISL_18781843, EPI_ISL_18781844, EPI_ISL_18781845, EPI_ISL_18781846, EPI_ISL_18781847, EPI_ISL_18781848, EPI_ISL_18781849, EPI_ISL_18781912, EPI_ISL_18781913, EPI_ISL_18781915, EPI_ISL_18781916, EPI_ISL_18781917, EPI_ISL_18781918, EPI_ISL_18781919, EPI_ISL_18781920, EPI_ISL_18781921, EPI_ISL_18781922, EPI_ISL_18781923, EPI_ISL_18781924, EPI_ISL_18781925, EPI_ISL_18781926, EPI_ISL_18781928, EPI_ISL_18781929, EPI_ISL_18786340, EPI_ISL_18786341, EPI_ISL_18786342, EPI_ISL_18786343, EPI_ISL_18786346, EPI_ISL_18786347, EPI_ISL_18786348, EPI_ISL_18786349, EPI_ISL_18786350, EPI_ISL_18786351, EPI_ISL_18786352, EPI_ISL_18786354, EPI_ISL_18786356, EPI_ISL_18786357, EPI_ISL_18786358, EPI_ISL_18786359 | | | |
| see above | California Department of Public Health | California Department of Public Health | Kath, C., Haw, M., Espinosa, A., and Hacker, J. |
| EPI_ISL_18798834 | PKM Mampang Prapatan | National Institute of Health Research and Development | Hana Apsari Pawestri, Arie Ardiansyah Nugraha, Fajar Nur Sulistyahadi, Markus Evan Anggia, Subangkit |
| EPI_ISL_18798835 | PKC Kebayoran Lama | National Institute of Health Research and Development | Hana Apsari Pawestri, Arie Ardiansyah Nugraha, Fajar Nur Sulistyahadi, Markus Evan Anggia, Subangkit |
| EPI_ISL_18798836 | RS Grha Kedoya Jakarta | National Institute of Health Research and Development | Hana Apsari Pawestri, Arie Ardiansyah Nugraha, Fajar Nur Sulistyahadi, Markus Evan Anggia, Subangkit |
| EPI_ISL_18798837 | PKC Kebayoran Baru | National Institute of Health Research and Development | Hana Apsari Pawestri, Arie Ardiansyah Nugraha, Fajar Nur Sulistyahadi, Markus Evan Anggia, Subangkit |
| EPI_ISL_18798838 | PKC Tanah Abang | National Institute of Health Research and Development | Hana Apsari Pawestri, Arie Ardiansyah Nugraha, Fajar Nur Sulistyahadi, Markus Evan Anggia, Subangkit |
| EPI_ISL_18798839 | PKM Bogor Timur | National Institute of Health Research and Development | Hana Apsari Pawestri, Arie Ardiansyah Nugraha, Fajar Nur Sulistyahadi, Markus Evan Anggia, Subangkit |
| EPI_ISL_18798840 | PKM Warung Jambu | National Institute of Health Research and Development | Hana Apsari Pawestri, Arie Ardiansyah Nugraha, Fajar Nur Sulistyahadi, Markus Evan Anggia, Subangkit |
| EPI_ISL_18798841 | PKC Pademangan | National Institute of Health Research and Development | Hana Apsari Pawestri, Arie Ardiansyah Nugraha, Fajar Nur Sulistyahadi, Markus Evan Anggia, Subangkit |
| EPI_ISL_18798842 | PKC Kebayoran Baru | National Institute of Health Research and Development | Hana Apsari Pawestri, Arie Ardiansyah Nugraha, Fajar Nur Sulistyahadi, Markus Evan Anggia, Subangkit |
| EPI_ISL_18809375, EPI_ISL_18809376 | California Department of Public Health | California Department of Public Health | Kath, C., Haw, M., Espinosa, A., and Hacker, J. |
| EPI_ISL_18822108, EPI_ISL_18822109 | Quest Diagnostics Nichols Institute | Los Angeles County Public Health Laboratories | S. McCann et al |
| EPI_ISL_18879931 | Central Public Health Laboratories, Ministry of Health Egypt | Center of Scientific Excellence for Influenza Viruses, National Research Centre (NRC), Egypt. | Wael H. Roshdy, Rabeh El-Shesheny, Yassmin Moatasim, Mina N. Kamel, Shaymaa Shawky, Mokhtar Gomaa, Galal Mahmoud, Amer Sayed, Amel Naguib, Nancy El Guindy, Ahmed Kandeil, Mohamed A. Ali, Amr Kandeel |
| EPI_ISL_18886301 | Reseau lab Bukavu-Kamituga DPS/ Sud -Kivu | Reseau lab Bukavu-Kamituga DPS/ Sud-Kivu | Leandre Murhula Masirika, Jean Claude Udahemuka, Leonard Schuele, Pacifique Ndishimye, Saria Otani ,Justin Bengehya Mbiribindi, Jean M. Marekani, Léandre Mutimbwa Mambo, Marjan Boter, David F. Nieuwenhuijse,Ernest Balyahamwabo Kalalizi,Trudie Lang, Jean Pierre Musabyimana, Frank M. Aarestrup, Marion Koopmans, Bas B. Oude Munnink, Freddy Belesi Siangoli |
| EPI_ISL_18886467 | Reseau Lab Bukavu -Kamituga DPS/Sud-Kivu | Reseau Lab Bukavu-Kamituga DPS /Sud-Kivu | Leandre Murhula Masirika, Jean Claude Udahemuka, Leonard Schuele, Pacifique Ndishimye, Saria Otani ,Justin Bengehya Mbiribindi, Jean M. Marekani, Léandre Mutimbwa Mambo, Marjan Boter, David F. Nieuwenhuijse,Ernest Balyahamwabo Kalalizi,Trudie Lang, Jean Pierre Musabyimana, Frank M. Aarestrup, Marion Koopmans, Bas B. Oude Munnink, Freddy Belesi Siangoli |
| EPI_ISL_18886588 | Reseau lab Bukavu-Kamituga DPS/Sud-Kivu | Reseau lab Bukavu -Kamituga DPS /Sud -Kivu | Leandre Murhula Masirika, Jean Claude Udahemuka, Leonard Schuele, Pacifique Ndishimye, Saria Otani ,Justin Bengehya Mbiribindi, Jean M. Marekani, Léandre Mutimbwa Mambo, Marjan Boter, David F. Nieuwenhuijse,Ernest Balyahamwabo Kalalizi,Trudie Lang, Jean Pierre Musabyimana, Frank M. Aarestrup, Marion Koopmans, Bas B. Oude Munnink, Freddy Belesi Siangoli |
| EPI_ISL_18886634 | Reseau Lab Bukavu-Kamituga DPS/Sud-Kivu | Reseau Lab Bukavu-Kamituga DPS/Sud Kivu | Leandre Murhula Masirika, Jean Claude Udahemuka, Leonard Schuele, Pacifique Ndishimye, Saria Otani ,Justin Bengehya Mbiribindi, Jean M. Marekani, Léandre Mutimbwa Mambo, Marjan Boter, David F. Nieuwenhuijse,Ernest Balyahamwabo Kalalizi,Trudie Lang, Jean Pierre Musabyimana, Frank M. Aarestrup, Marion Koopmans, Bas B. Oude Munnink, Freddy Belesi Siangoli |
| EPI_ISL_18886635 | Reseau Lab Bukavu-Kamituga DPS/Sud Kivu | Reseau Lab Bukavu-Kamituga DPS/Sud Kivu | Leandre Murhula Masirika, Jean Claude Udahemuka, Leonard Schuele, Pacifique Ndishimye, Saria Otani ,Justin Bengehya Mbiribindi, Jean M. Marekani, Léandre Mutimbwa Mambo, Marjan Boter, David F. Nieuwenhuijse,Ernest Balyahamwabo Kalalizi,Trudie Lang, Jean Pierre Musabyimana, Frank M. Aarestrup, Marion Koopmans, Bas B. Oude Munnink, Freddy Belesi Siangoli |
| EPI_ISL_18886639 | Reseau Lab Bukavu-Kamituga DPS/ SUD-Kivu | Reseau Lab Bukavu-Kamituga DPS/ Sud-Kivu | Leandre Murhula Masirika, Jean Claude Udahemuka, Leonard Schuele, Pacifique Ndishimye, Saria Otani ,Justin Bengehya Mbiribindi, Jean M. Marekani, Léandre Mutimbwa Mambo, Marjan Boter, David F. Nieuwenhuijse,Ernest Balyahamwabo Kalalizi,Trudie Lang, Jean Pierre Musabyimana, Frank M. Aarestrup, Marion Koopmans, Bas B. Oude Munnink, Freddy Belesi Siangoli |
| EPI_ISL_18899228 | QUEST DIAGNOSTICS NICHOLS INSTITUTE | Los Angeles County Public Health Laboratories | S. McCann et. al. |
| EPI_ISL_18899230 | QUEST DIAGNOSTICS WEST HILLS | Los Angeles County Public Health Laboratories | S. McCann et. al. |
| EPI_ISL_18899231 | LOS ANGELES COUNTY PUBLIC HEALTH LABORATORY | Los Angeles County Public Health Laboratories | S. McCann et. al. |
| EPI_ISL_18899233 | QUEST DIAGNOSTICS NICHOLS INSTITUTE | Los Angeles County Public Health Laboratories | S. McCann et. al. |
| EPI_ISL_18899234, EPI_ISL_18899235 | LABCORP | Los Angeles County Public Health Laboratories | S. McCann et. al. |
| EPI_ISL_18899237, EPI_ISL_18899239 | LOS ANGELES COUNTY PUBLIC HEALTH LABORATORY | Los Angeles County Public Health Laboratories | S. McCann et. al. |
| EPI_ISL_18899240, EPI_ISL_18899241, EPI_ISL_18899242 | QUEST DIAGNOSTICS WEST HILLS | Los Angeles County Public Health Laboratories | S. McCann et. al. |
| EPI_ISL_18899243, EPI_ISL_18899244 | ARUP LABORATORIES | Los Angeles County Public Health Laboratories | S. McCann et. al. |
